# Diagnostic tests for Schistosomiasis for low prevalence settings: a systematic review and Meta-Analysis

**DOI:** 10.1101/2021.05.05.21256678

**Authors:** Michel T Vaillant, Fred Philippy, Jessica Barré, Dmitry Bulaev, Amadou T Garba

## Abstract

**Background:** Tests for diagnosing schistosomiasis in areas where prevalence is low due to control programme of the disease should be suffiently sensitive to detect the residual disease. If they had sufficient diagnostic accuracy they could replace conventional microscopy as they provide a quicker answer and are easier to use.

**Objectives:** To compare sensitivity and specificity of new tests, especially rapid diagnostic tests (RDTs), with regard to a certain reference test.

**Methods:** We searched the electronic databases Pubmed, EMBASE, the Cochrane Library and LILACS up to February 2021. Furthermore we searched results from the previous meta-analyses.

We included studies that used microscopy as the reference standard: for S. haematobium, microscopy of urine prepared by filtration, centrifugation, or sedimentation methods; and for S. mansoni, microscopy of stool by Kato-Katz thick smear.

Two review authors independently extracted data, assessed quality of the data using QUADAS-2, and performed meta-analysis where appropriate. Grading of evidence was done with the GRADE methodology by using GradePro. Using the variability of test thresholds, we used a bayesian bivariate random-effects summary receiver operating characteristic model for all eligible tests. We investigated heterogeneity, and carried out sensitivity analyses where possible. Results for sensitivity and specificity are presented as percentages with 95% confidence intervals (CI).

**Results:** The review gathered 203 articles stating a diagnostic test for the diagnosis of S. haematobium and S. mansoni out of which 114 entered the analyses. Microscopy of Urine filtration or Kato-Katz smears were used as the reference standard.

Compared with Kato-Katz smears, AWE-SEA ELISA (Se=94%; Sp=64%) is comparable to CCA1 (Se=87%; Sp=60%). IgG ELISA (Se=93%; Sp=68%) has also a very good ability to detect true positive as well as CAA cassette (Se=73%; Sp=68%). For S. haematobium, proteinuria (Se=59%; Sp=83%) and haematuria (Se=74%; Sp=87%) reagent strips showed reasonably high specificities with a considerably better sensitivity for the haematuria test.

There are interesting promising new diagnostic tests that were tested in field studies. However prevalences of the locations where these studies took place are variable and there are no specific study with a high number of patients in areas with low level of schistosomiasis infection.

## Background

Schistosomiasis is an acute and chronic parasitic disease caused by blood flukes (trematode worms) of the genus Schistosoma. Transmission occurs when people suffering from schistosomiasis contaminate freshwater sources with their excreta containing parasite eggs. Preventive chemotherapy for schistosomiasis, where people and communities are targeted for large-scale treatment, is required in 52 endemic countries with moderate-to-high transmission. Estimates show that at least 218 million people required preventive treatment worldwide in 2015. The neglected tropical diseases (NTD) road map launched in 2012 followed by the London Declaration of NTD created a new emphasis for the control of schistosomiasis with commitments of various partners to support the fight against schistosomiasis. The NTD road map set as target to reach the 75-100% coverage of school aged children in 2020, and to eliminate the disease in some regions.

Currently there is no guidance available for the evaluation of the interruption of the transmission of schistosomiasis. The current implementation guidelines based mainly on expert opinion need to be revised according to the available scientific evidence.

The goal of the WHO guidelines is to provide evidence based recommendations to countries in their effort to move from control to interruption of transmission. It will help countries on the implementation of preventive chemotherapy for schistosomiasis and on how to verify if the transmission of the disease is interrupted in the country.

Currently, the guideline in use for the morbidity control of schistosomiasis is based on the recommendations by the expert committee in 2002, updated in 2006 to take into account additional strategies, the treatment in low prevalence areas and of special groups at risk and in 2017 for the use of the Circulating Cathodic Antigen (CCA) diagnostic tests.

New updated guidelines are needed, in particular, because new sensitive diagnostic tools have been developed and to guide their utilisation in low transmission areas. The target condition being diagnosed are infections to Schistosoma mansoni and Schistosoma Heamatobium.

For S. mansoni, the reference standard test is the index microscopy test namely the two Kato-Katz smear performed on 2 sample of feces (quadruple KK). However double KK is also found as well as 6KK. It is Urine Microscopy for S. heamatobium. The comparators are based on the Hemastix dipstick, CCA, CAA, PCR, serology. All diagnostic tests found in the literature search are in Table 1.

**Table 1.**
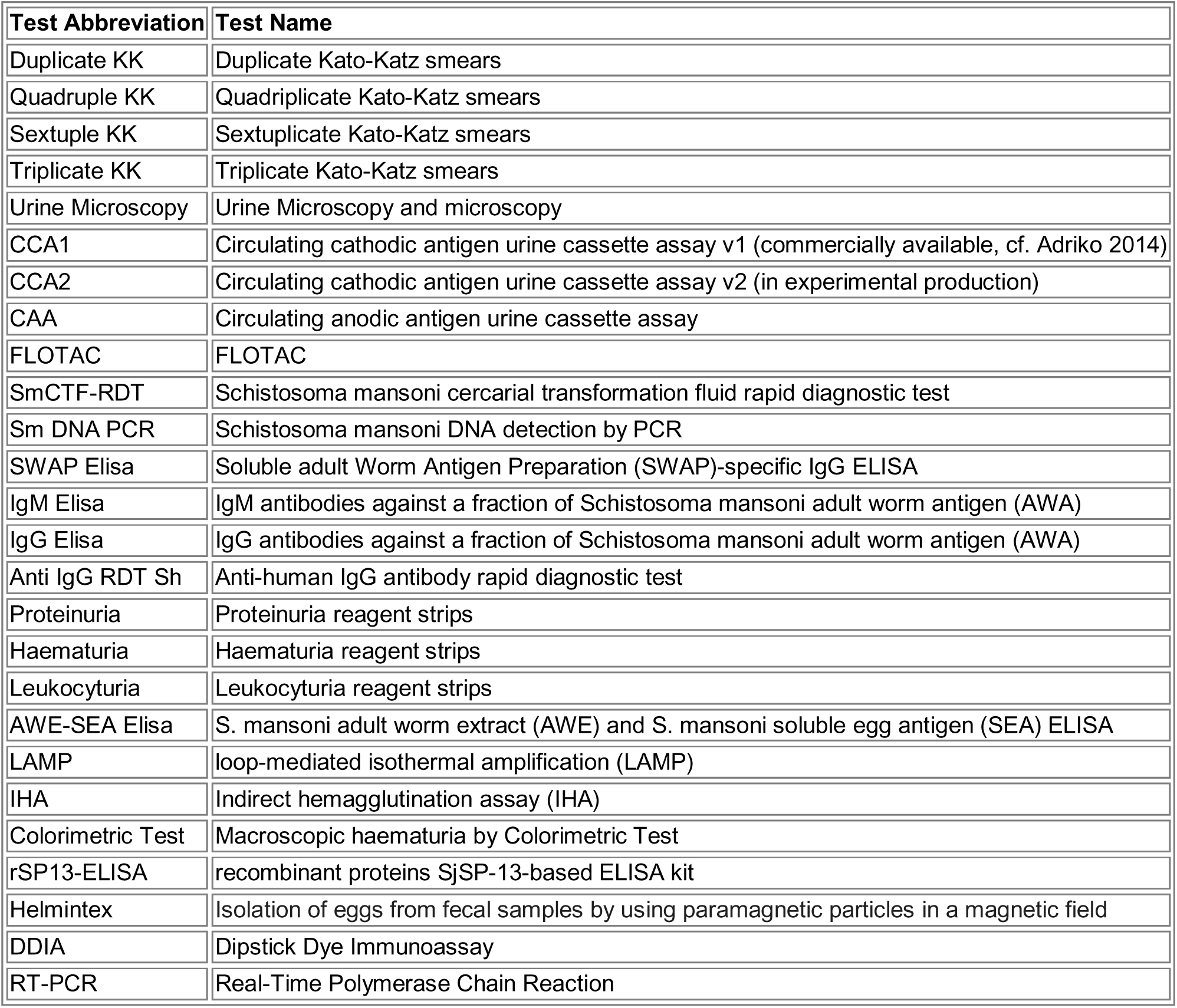
Diagnostic tests

| Test Abbreviation | Test Name |
| --- | --- |
| Duplicate KK | Duplicate Kato-Katz smears |
| Quadruple KK | Quadruplicate Kato-Katz smears |
| Sextuple KK | Sextuplicate Kato-Katz smears |
| Triplicate KK | Triplicate Kato-Katz smears |
| Urine Microscopy | Urine Microscopy and microscopy |
| CCA1 | Circulating cathodic antigen urine cassette assay v1 (commercially available, cf. Adriko 2014) |
| CCA2 | Circulating cathodic antigen urine cassette assay v2 (in experimental production) |
| CAA | Circulating anodic antigen urine cassette assay |
| FLOTAC | FLOTAC |
| SmCTF-RDT | Schistosoma mansoni cercarial transformation fluid rapid diagnostic test |
| Sm DNA PCR | Schistosoma mansoni DNA detection by PCR |
| SWAP Elisa | Soluble adult Worm Antigen Preparation (SWAP)-specific IgG ELISA |
| IgM Elisa | IgM antibodies against a fraction of Schistosoma mansoni adult worm antigen (AWA) |
| IgG Elisa | IgG antibodies against a fraction of Schistosoma mansoni adult worm antigen (AWA) |
| Anti IgG RDT Sh | Anti-human IgG antibody rapid diagnostic test |
| Proteinuria | Proteinuria reagent strips |
| Haematuria | Haematuria reagent strips |
| Leukocyturia | Leukocyturia reagent strips |
| AWE-SEA Elisa | S. mansoni adult worm extract (AWE) and S. mansoni soluble egg antigen (SEA) ELISA |
| LAMP | loop-mediated isothermal amplification (LAMP) |
| IHA | Indirect hemagglutination assay (IHA) |
| Colorimetric Test | Macroscopic haematuria by Colorimetric Test |
| rSP13-ELISA | recombinant proteins SjSP-13-based ELISA kit |
| Helmintex | Isolation of eggs from fecal samples by using paramagnetic particles in a magnetic field |
| DDIA | Dipstick Dye Immunoassay |
| RT-PCR | Real-Time Polymerase Chain Reaction |

Theoretical sensitivity of some of the diagnostic tests described in Table 1 were investigated (Table 2).

**Table 2.** Theoretical sensitivity of diagnostic tests

| Theoretical analytic sensitivity | [Insert text] | [Insert text] |
| --- | --- | --- |
| Kato-Katz | 24 eggs per gram of stool (EPG) | Enk MJ 2008, de Vlas SJ 1992 |
| FLOTAC basic technique | 1 EPG | Cringoli G 2010 |
| FLOTAC dual technique | 2 EPG | Cringoli G 2010 |
| FLOTAC double technique | 2 EPG | Cringoli G 2010 |
| FLOTAC pellet | 2 EPG | Cringoli G 2010 |

The objectives of the systematic review and meta-analysis were to compare sensitivity and specificity of new tests with regard to the Kato-Katz test.

## Materials and Methods

### Criteria for considering studies for this review

#### Types of studies

Use of diagnostic test, validation study, availability of tests performances and/or quantity of patients/data in crossed categories of at least 2 tests

#### Participants

Adults and Children (SAC and pre SAC) living in endemic areas that have received elimination interventions

#### Target conditions

Schistosomiasis based on infection by *Schistosoma mansoni* or *Schistosoma haematobium*

### Search methods for identification of studies

#### Electronic searches

To identify articles relevant to the question, a search in Pubmed, EMBASE, the Cochrane Library and LILACS up to February 2021 will be undergone using analogous search terms. A combination of medical subject headings (MeSH) and title and abstract keywords such as “Schistosomiasis”, “Schistosoma”, “”, or “”, focusing on terms to describe the relevant population: ((schistosomiasis[Title] OR schistosoma[Title]) OR (schistosomiasis[Other Term] OR schistosoma[Other Term])) AND ((diagnostic*[Title]) OR (diagnostic*[Other Term])).

More precisely, the full electronic database search result can be broken down as follows:

- Search in EMBASE on 10/12/2020

○ MeSH: (’schistosomiasis’:ti OR ’schistosoma’:ti OR ’schistosomiasis’:kw OR ’schistosoma’:kw) AND (’diagnostic*’:ti OR ’diagnostic*’:kw)
○ 345 results
- Search in PubMed on 10/12/2020

○ MeSH: (((“schistosomiasis”[Title] OR “schistosoma”[Title]) OR (“schistosomiasis”[Other Term] OR “schistosoma”[Other Term])) AND (“diagnostic*”[Title] OR “diagnostic*”[Other Term])) OR ((“schistosomiasis”[Title] OR “schistosoma”[Title] OR (“schistosomiasis”[Other Term] OR “schistosoma”[Other Term])) AND (“specificity”[Title] OR “sensitivity”[Title] OR (“specificity”[Other Term] OR “sensitivity”[Other Term]))))
○ 416 results
- Search in EMBASE on 03/02/2021 for the years 2018-2021

○ MeSH: (’schistosomiasis’:ti OR ’schistosoma’:ti OR ’schistosomiasis’:kw OR ’schistosoma’:kw) AND (’diagnostic*’:ti OR ’diagnostic*’:kw) AND (2018:py OR 2019:py OR 2020:py OR 2021:py)
○ 80 results
- Search in PubMed on 03/02/2021 for the years 2018-2021

○ MeSH: (((“schistosomiasis”[Title] OR “schistosoma”[Title]) OR (“schistosomiasis”[Other Term] OR “schistosoma”[Other Term])) AND (“diagnostic*”[Title] OR “diagnostic*”[Other Term])) OR ((“schistosomiasis”[Title] OR “schistosoma”[Title] OR (“schistosomiasis”[Other Term] OR “schistosoma”[Other Term])) AND (“specificity”[Title] OR “sensitivity”[Title] OR (“specificity”[Other Term] OR “sensitivity”[Other Term])))) Filters: from 2018 – 2021
○ 95 results
- Search in LILACS on 03/02/2021

○ MeSH: ti:(schistosomiasis OR schistomosa) AND (ti:diagnostic* OR mh:diagnostic*) AND (db:(“LILACS”) AND la:(“en”))
○ 38 results
- Search in Cochrane on 03/02/2021

○ MeSH: (schistosomiasis OR schistosoma) AND diagnostic*
○ 20 results
- 76 additional references could be found by screening bibliographies of other meta-analyses or by executing specific searches
- 370 database search results from previous study

All in all, the search yielded 1440 results, of which 756 were identified as duplicates.

### Data collection and analysis

For each analysis paired forest plots of sensitivity and specificity were created and summary ROC plots were produced:

- Plots of the summary ROC curve,
- Average operating points including 95% confidence intervals and 95% prediction regions
- Bayesian bivariate random effects model used to estimae Sentivity, specificity.

#### Selection of studies

We looked for the use of diagnostic tests, validation studies, availability of test performance measurements and/or quantity of patients/data in crossed categories of at least 2 tests.

All search results have been uploaded in the web app Rayyan QRCI (https://rayyan.qcri.org/welcome) to perform sorting and selection of articles based on an extended list of keywords by two review authors. After submitting all their inclusion and exclusion decisions on the 674 uploaded references, the results were compared. 203 references were declared eligible for full-text assessment.

Subsequently, data could be extracted from 114 references. The quality of the data has been assessed using QUADAS-2, and meta-analysis was performed where appropriate.

Reasons for the exclusion of studies can be found in Appendix B.

#### Data extraction and management

We included studies that provide participant data. Only studies in which true-positives (TPs), true-negatives (TNs), false-positives (FPs), and false-negatives (FNs) were reported or could be extracted from the data were included.

Other data were extracted such as:

- Study authors, publication year, and journal
- Study design.
- Study participants’ age and sex.
- Prevalence of schistosomiasis.
- Treatment status of participants with praziquantel before study
- Reference standard (microscopy), including number of samples per individual
- Index tests

#### Assessment of methodological quality

This review is based on the QUADAS-2 guidelines for the review of diagnostic accuracy. It collects estimates from the scientific literature and aggregate them to provide meta-analytic estimates

<u>Assessment of methodological quality table:</u>

The assessment of the recommended 11 QUADAS items was performed.

Associated charcacteristics of the studies, as well as the 11 QUADAS items for every included study can be found in Appendix B.

#### Grading of the evidence

The GRADE methodology was used to rate the evidence concerning each diagnostic tool. The GradePro GDT tool (https://gradepro.org/) helped in gathering information and rating. Summary of Findings (SoF) tables will be created for every test comparison to evaluate quality of evidence (or certainty) of evidence and strength of recommendations. These tables can be found in Appendix C.

#### Statistical analysis and data synthesis

Intervention:

Diagnostic tests: Urine Microscopy, Hemastix dipstick, CCA, CAA, PCR, serology tests, reagent strips

Comparison:

Comparison of various tests found in the litterature

Outcomes:

- Performances of the tests (sensitivity, specificity, PPV, NPV)
- Disease prevalence with the reference test and with the index test

Using the variability of test thresholds, we used a bayesian bivariate random-effects summary receiver operating characteristic model for all eligible tests.

In order to improve the summary measures considering the group reference test, the Kato-Katz thick smears (single, duplicate, triplicate, etc.) were lumped together based on known differences in sensitivity. This work was carried out for CCA1, CCA2. For FLOTAC we also considered lumping FLOTAC fresh, FLOTAC (10 days) and FLOTAC (30 days). This has been added in the respective tools paragraphs.

#### Investigations of heterogeneity

We investigated heterogeneity by examining the forest plots, and carried out sensitivity analyses where possible.

#### Assessment of reporting bias

We did not assess reporting bias.

### PICO question

In people living in schistosomiasis endemic settings (s. mansoni and s. haematobium), do newer diagnostics when compared to conventional standard (KK for s. mansoni and urine microscopy for s. haem) have better sensitivity and specificity?

## Results

### Results of the search

Together with the records yielded during the first attempt of this study, our search yielded 1364 records. Out of the 1364 records, 608 records were suitable for the review after duplicates were removed. Abstracts were reviewed and 127 articles were eligible for-full text assessment. 76 additionnal records were identified through reading of the references tables of these articles, which results in a total of 203 records eligible for full-text assessment.

Out of these 203 records, 89 were excluded for the following reasons:

- Article in Chinese (n=5)
- Article in Portuguese (n=1)
- Use of a combined gold standard (n=17)
- Empirical data not available (n=5)
- Ongoing study (n=1)
- No indication regarding the reference test (n=2)
- Meta-analysis (n=3)
- Wrong type of study (n=40)
- Statistical analysis (n=5)
- Paper retracted (n=1)
- Article was not retrieved (n=9)

The PRISMA flow diagram can be found in Appendix A (Figure 1).

**Figure 1.**
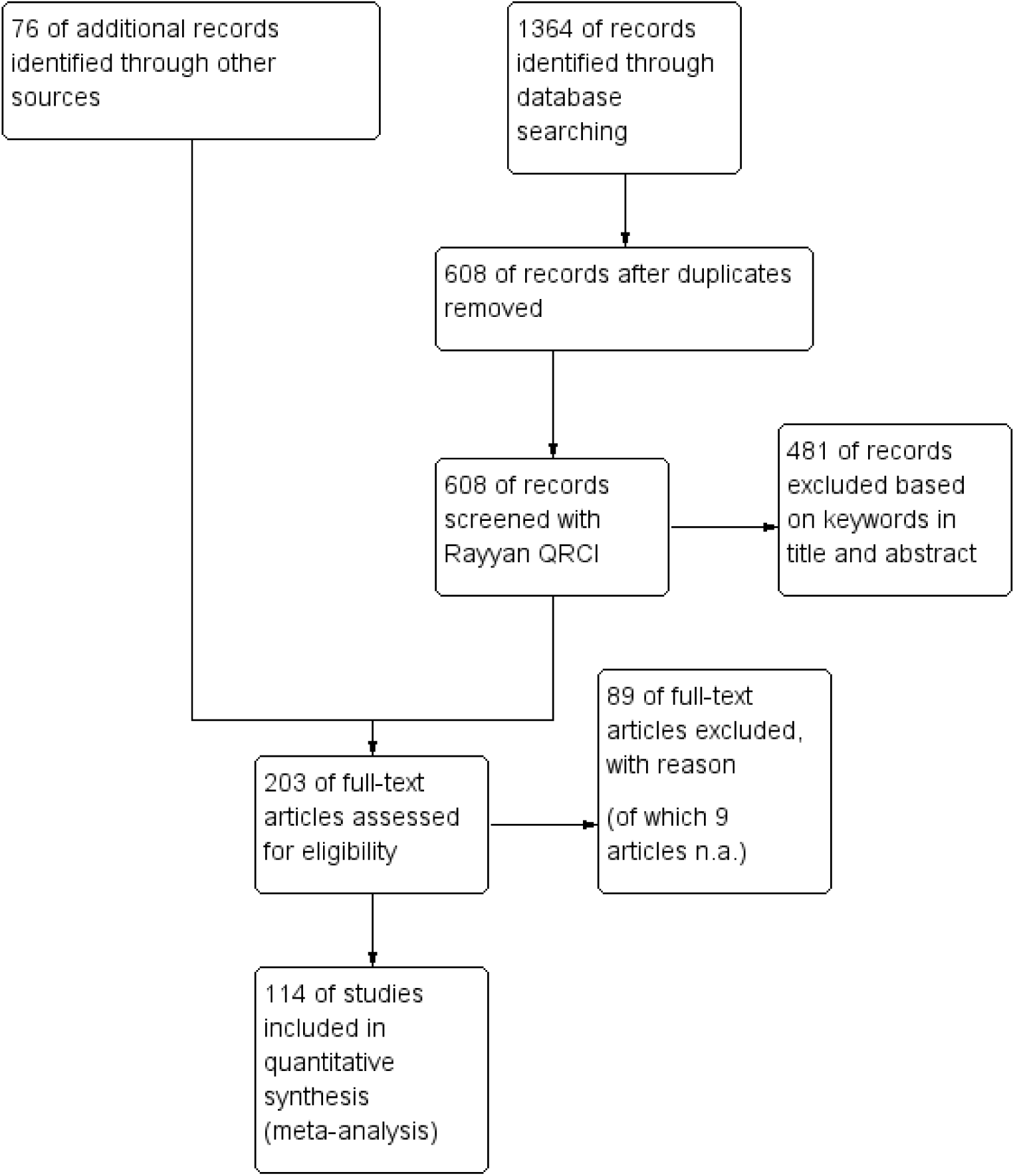
PRISMA Flow Diagram

### Methodological quality of included studies

Using the Quadas-2 tool we evaluated the risk of bias in the quality appraisal of the included studies. We evaluated an unclear risk of bias of about 12.30% in patient selection and 6.56% in the use of the index test. Concerning the reference standard the risk of bias was 29.50%. In terms of applicability concerns, only patient selection would show a 2.46% unclear evaluation.

Figures 2 and 3 provide a visual illustration of the evaluation of risk of bias and applicability concerns.

**Figure 2.**
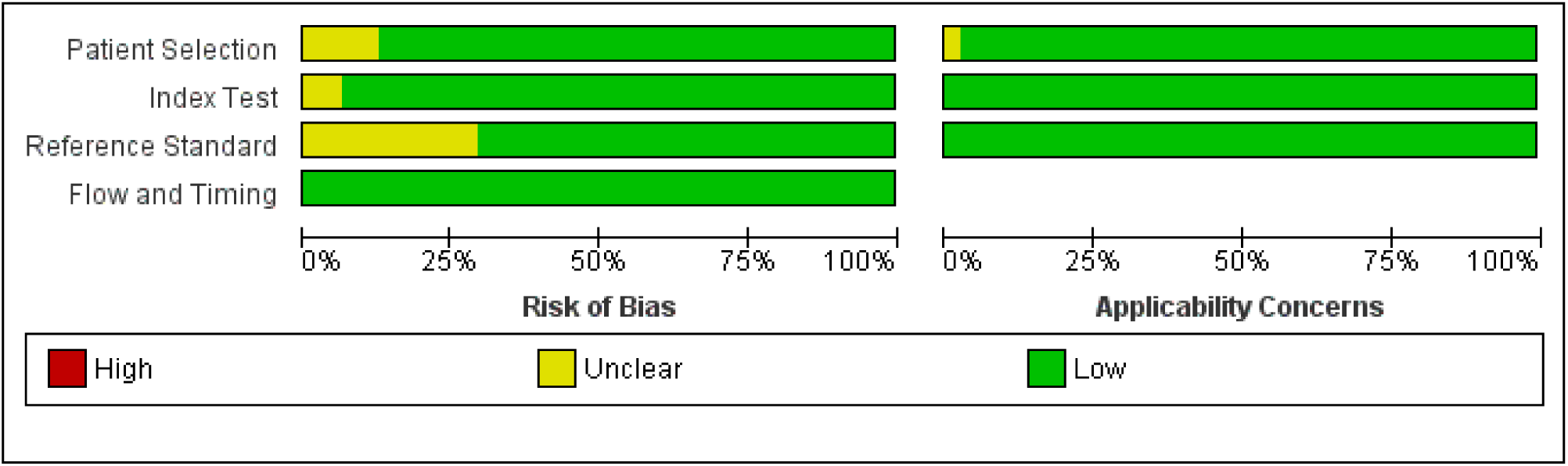
Risk of bias and applicability concerns graph: review authors’ judgements about each domain presented as percentages across included studies

**Figure 3.**
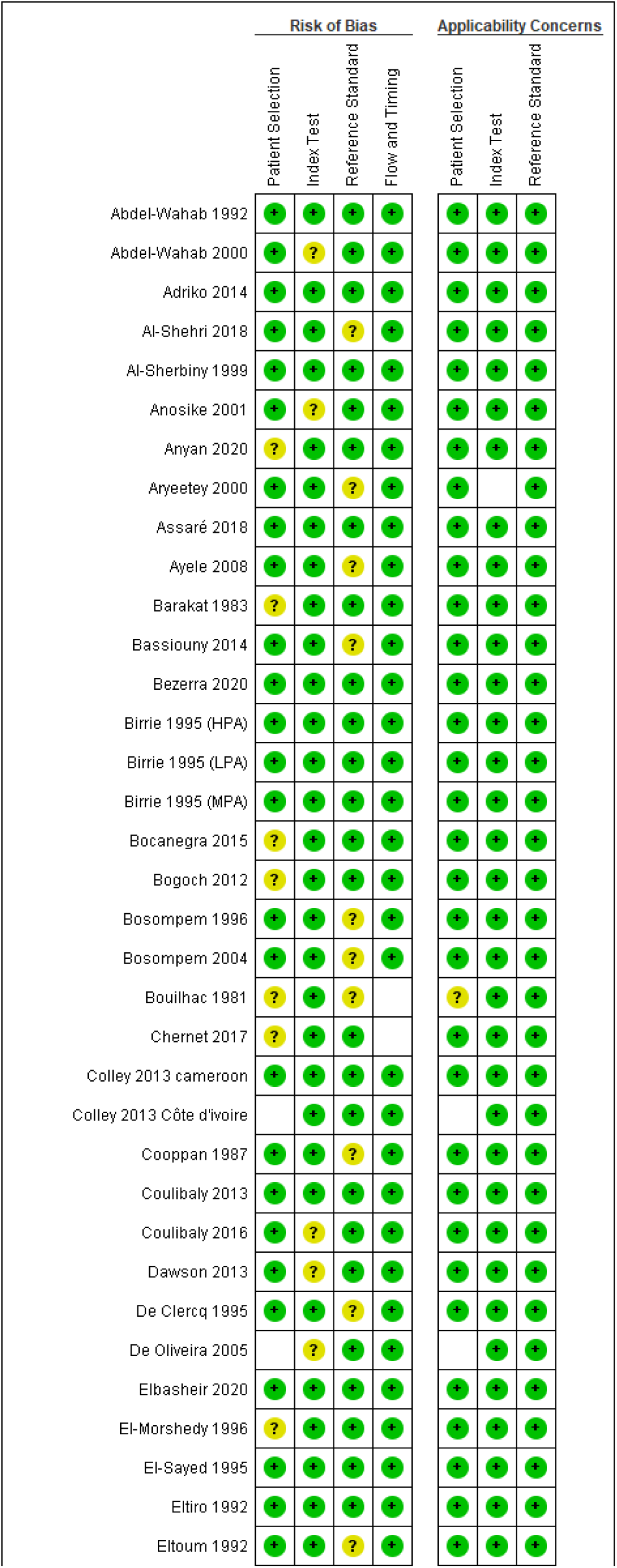
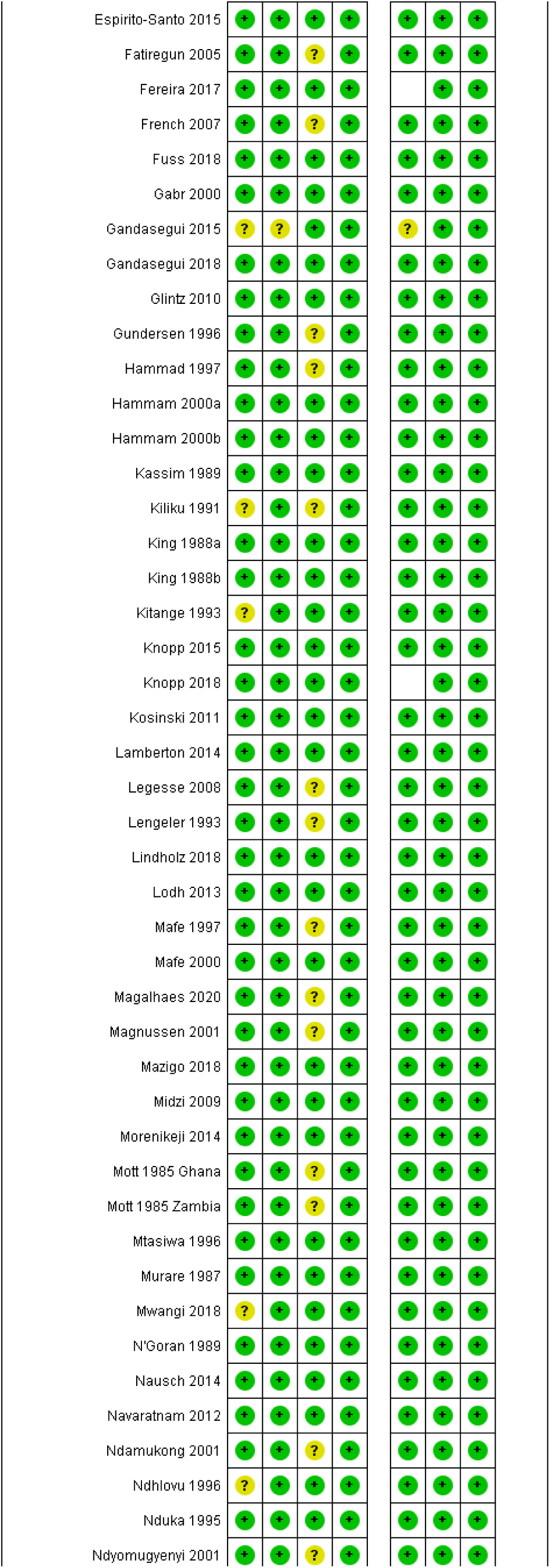
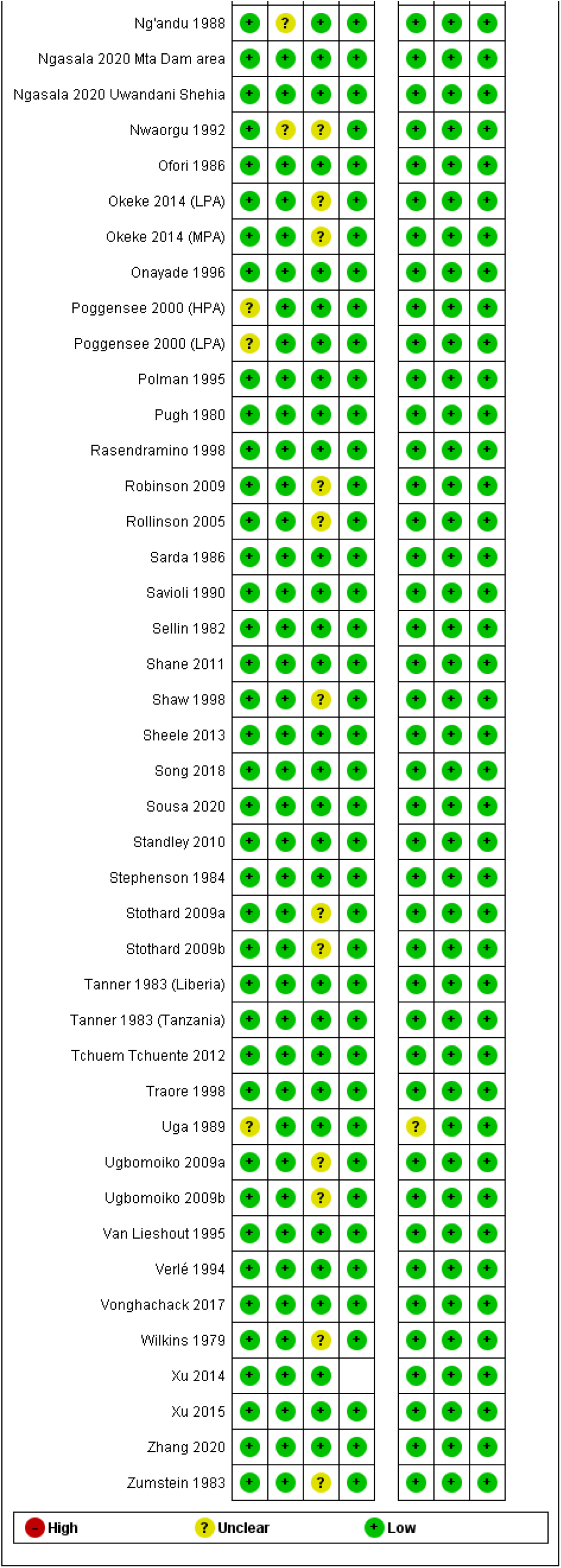
Risk of bias and applicability concerns summary : review authors’judgements about each domain for each included study

### Findings

#### Prevalence of schistosomiasis

The raw prevalences detected in the different studies that were included in the review are shown in Table 3.

**Table 3.** Schistosomiasis infection prevalences

| Study | country | Population | Prevalence<br>(ref. test) | [95% CI] | Prevalence<br>(Index test) | (95% CI |
| --- | --- | --- | --- | --- | --- | --- |
| Abdel-Wahab 1992 | Egypt |  | 0.336 |  | Haem.: 0.4313<br>Protein.: 0.1493<br>Leuko.: 0,1564 |  |
| Abdel-Wahab 2000 | Egypt |  | 0.137 |  | Haem.: 0.3<br>Protein.: 0.035 |  |
| Adriko 2014 | Uganda |  | 0.250 |  | 0.626 |  |
| Al-Shehri 2018 | Uganda |  | 0.567 | [0.508;0.63] | KK: 0.441<br>SEA-EL: 0.751<br>PCR: 0.674 | KK: [0.38;0.502]<br>SEA-EL:<br>[0.698;0.804]<br>PCR: [0.616;0.731] |
| Al-Sherbiny 1999 | Nigeria |  | 0.158 |  |  |  |
| Anosike 2001 | Nigeria |  | 0.499 |  | 0.295 |  |
| Anyan 2020 | Ghana |  |  |  | S. haem. :<br>0.485<br>S. mans: 0.466 | [0.41;0.56]<br>[0.39;0.54] |
| Aryeetey 2000 | Ghana |  |  |  | Haem.: |  |
| Assaré 2018 | Côte d'Ivoire |  | 0.066 | [0.047;0.084] | 0.33 | [0.295;0.366] |
| Ayele 2008 |  |  | 0.48 |  | 0.432 |  |
| Barakat 1983 | Egypt |  | 0,3787 |  | 0,3984 |  |
| Bassiouny 2014 | Yemen |  | 0.181 |  | 0.161 |  |
| Bezerra 2020 | Brazil |  | 0.4882 |  | 0.4488 |  |
| Birrie 1995 (HPA) | Ethiopia |  | 0.305 |  | 0.36 |  |
| Birrie 1995 (LPA) | Ethiopia |  | 0.032 |  | 0.045 |  |
| Birrie 1995 (MPA) | Ethiopia |  | 0.21 |  | 0.308 |  |
| Bocanegra 2015 | Central Angola |  | 0.6118 |  | 0.657 |  |
| Bogoch 2012 | Northern Ghana |  | 0.068 |  |  |  |
| Bosompem 1996 | Ghana |  | 0.476 |  | Haem.: 0.397<br>Protein.: 0.236 |  |
| Bosompem 2004 | Ghana |  | 0.603 |  | Haem.: 0.27<br>Protein.: 0.305 |  |
| Bouilhac 1981 |  |  | 0.3506 |  | 0.3052 |  |
| Chernet 2017 | Las Palmas de Gran Canaria, Spain | sub saharan immigrants | 0.4148 |  |  |  |
| Colley 2013 cameroon | Cameroon |  | 0.384 |  | 0.622 |  |
| Colley 2013 Côte | Côte d'Ivoire |  | 0.479 |  | 0.455 |  |
| d'ivoire |  |  |  |  |  |  |
| Cooppan 1987 | South Africa |  | 0.809 |  | Haem.: 0.697<br>Protein.: 0,774 |  |
| Coulibaly 2013 | South Côte d'Ivoire |  | 0.212 | [0.144;0.299] | 0.669 | [0.576;0.752] |
| Coulibaly 2016 | Côte d'Ivoire |  | 0.933 | [0.892;0.973] | 0.953 | [0.919;0.987] |
| Dawson 2013 | Uganda |  | <3y: 0.3<br>>3y: 0.6 | [0.18;0.47]<br>[0.45;0.65] | (CCA) 0.5<br>0.78 | [0.32;0.62]<br>[0.6;0.9] |
| De Clercq 1995 | Mali |  | 0,576 |  | 0,6372 |  |
| De Oliveira 2005 | Brazil |  | 0.3649 |  |  |  |
| Elbasheir 2020 | Sudan |  | 0.358 |  | 0.438 |  |
| El-Morshedy 1996 | Egypt |  | 0.57 |  | 0.455 |  |
| El-Sayed 1995 | Egypt |  | 0.033 |  | 0.29 |  |
| El tiro 1992 | Ethiopia |  | 0.8350 |  | 0.9300 |  |
| Eltoum 1992 | Sudan |  | 0.42 |  | 0.62 |  |
| Espirito-Santo 2015 | Brazil |  | 0.008 |  | ELISA-IgG:<br>0.116<br>ELISA-IgM:<br>0.214<br>COPT: 0.54<br>ITF-IgM: 0.158<br>qPCR-feces:<br>0.98<br>qPCR-serum:<br>0.15 |  |
| Fatiregun 2005 | Nigeria |  | 0.122 |  | 0.166 |  |
| Ferreira 2017 | Brazil |  | 0.08 |  | 0.273 |  |
| French 2007 | Tanzania |  | 0.1314 |  |  |  |
| Fuss 2018 | Tanzania |  | 0.852 | [0.812;0.892] | PCR: 0.929<br>CCA: 0.949 | PCR: [0.9;0.958]<br>CCA: [0.924;0.974] |
| Gabr 2000 | Egypt |  | 0.089 | [0.081;0.097] | Haem.: 0.219<br>Protein.: 0.0455 |  |
| Gandasegui 2015 | Spain |  |  |  |  |  |
| Gandasegui 2018 | Angola |  | 0.506 |  | microhaem.:<br>0.483<br>LAMP: 0.738 |  |
| Glantz 2010 | South Côte d'Ivoire |  | 0.643 |  | 0.536 |  |
| Gundersen 1996 | Malawi |  | 0.1962 |  | Haem.: 0.8<br>Protein.: 0.7923<br>Leuk.: 0.7577 |  |
| Hammad 1997 | Egypt |  |  |  |  |  |
| Hammam 2000a | Egypt |  | 0.048 | [0.041;0.055] | Haem.: 0.238<br>Protein.: 0.119 |  |
| Hammam 2000b | Egypt |  | 0.052 | [0.047;0.057] | Haem.: 0.179<br>Protein.: 0.008 |  |
| Kassim 1989 | Nigeria |  | 0,1302 |  | Haem.: 0,1193<br>Protein.: 0,2104 |  |
| Kiliku 1991 | Kenya |  | 0.5939 |  | Haem.: 0.4061<br>Protein.: 0.6197 |  |
| King 1988a | Kenya |  | 0.69 |  | Haem.: 0.54<br>Protein.: 0.56 |  |
| King 1988b | Kenya |  | 0.65 |  | 0,3709 |  |
| Kitange 1993 | Tanzania |  | 0.3281 |  | Haem.: 0.3834<br>Protein.: 0.1225 |  |
| Knopp 2015 | Tanzania |  | 0,0475 |  | 0.0408 |  |
| Knopp 2018 | Zanzibar |  | 0.0451 |  | 0.0786 |  |
| Kosinski 2011 | Ghana |  | Girls=0.204<br>Boys=0.316 |  |  |  |
| Lamberton 2014 | Uganda |  | 0.9474 | [0.872;0.979] | 0.8816 |  |
| Legesse 2008 | Ethiopia |  | 0.364 |  | 0.652 |  |
| Lengeler 1993 | Tanzania |  | 0.223 |  | 0.331 |  |
| Lindholz 2018 | Brazil |  | 0.119 |  | HTX: 0.406<br>CCA: 0.716 |  |
| Lodh 2013 | Zambia |  | 0.51 |  | 0.6 |  |
| Mafe 1997 | Nigeria |  | 0.574 |  | 0.48 |  |
| Mafe 2000 | Tanzania |  | 0.325 |  | 0.369 |  |
| Magalhaes 2020 | Brazil |  | 0.55 |  | 2KK: 0.199<br>6KK: 0.37<br>Helm.: 0.40<br>CCA: 0.48 |  |
| Magnussen 2001 | Tanzania |  |  |  | 0.30 |  |
| Mazigo 2018 | Tanzania |  | 0.578 | [0.529;0.624] | 0.874 | [0.835;0.9] |
| Midzi 2009 | Zimbabwe |  | 0.40 |  | 0.65 |  |
| Morenikeji 2014 | Nigeria |  | 0.571 |  | Macroheam.:<br>0.164<br>Microhaem.: 0.5<br>Proteinuria:<br>0.655 |  |
| Mott 1985 Ghana | Ghana |  | 0,6904 |  | Haem.: 0,5107<br>Protein.: 0,8359 |  |
| Mott 1985 Zambia | Zambia |  | 0,6951 |  | Haem.: 0,596<br>Protein.: 0,6959 |  |
| Mtasiwa 1996 | Tanzania |  | 0.6757 |  | 0.6708 |  |
| Murare 1987 | Zimbabwe |  | 0,6983 |  | Haem.: 0,5948<br>Protein.: 0,7112 |  |
| Mwangi 2018 | Kenya |  | 0,4595 |  | 0,4465 |  |
| N'Goran 1989 | Ivory Coast |  | 0.3278 |  | 0.4963 |  |
| Nausch 2014 | Zimbabwe |  | Sh>50%,<br>Sm<20%<br>Sh=0.528,<br>Sm=0.11 | Sh=[0.420;0.633]<br>Sm=[0.054;0.193] | 0.8351 |  |
| Navaratnam 2012 | Uganda |  | 0.322 | [0.283;0.362] | 0.4 | [0.358;0.44] |
| Ndamukong 2001 | Cameroon |  | 0.536 |  | Haem.: 0.498<br>Protein.: 0.495 |  |
| Ndhlovu 1996 | Zimbabwe |  | 0.5363 |  | 0.8715 |  |
| Nduka 1995 | Nigeria |  | 0.2103 |  | 0.0352 |  |
| Ndyomugenyi 2001 | Tanzania |  | 0.476 |  | 0.522 |  |
| Ng'andu 1988 | Zambia |  | 0,4102 |  | Haem.: 0,4199<br>Protein.: 0,3592 |  |
| Ngasala 2020 Mta Dam area | Tanzania |  | 0.068 | [0.046;0.099] | 0.515 |  |
| Ngasala 2020 Uwandani Shehia | Tanzania |  | 0.387 | [0.312;0.468] | 0.452 |  |
| Nwaorgu 1992 | Nigeria |  | 0.57 |  |  |  |
| Ofori 1986 | Ghana |  | 0,5424 |  | Haem.: 0,3814<br>Protein.: 0,4661 |  |
| Okeke 2014 (LPA) | Nigeria |  | 0.051 | [0.0259;0.0761] | Micro-haem.: 0.064<br>Macro-haem.: 0.002<br>Proteinuria: 0.243 | Micro-haem.: [0.0361;0.0919]<br>Macro-haem.: [0.0041;0.0359]<br>Proteinuria: [0.1941;0.2919] |
| Okeke 2014 (MPA) | Nigeria |  | 0.266 | [0.2022;0.3298] | Micro-haem.: 0.207<br>Macro-haem.: 0.043<br>Proteinuria: 0.179 | Micro-haem.: [0.1485;0.2655]<br>Macro-haem.: [0.0137;0.0723]<br>Proteinuria: [0.1485;0.2655] |
| Onayade 1996 | Nigeria |  | 0.8857 |  | 0.5413 |  |
| Poggensee 2000 (HPA) | Tanzania |  | 0.53 |  | Haem.: 0.547<br>Protein.: 0.109<br>Leuk.: 0.547 |  |
| Poggensee 2000 (LPA) | Tanzania |  | 0.04 |  | Haem.: 0.0297<br>Protein.: 0.086<br>Leuk.: 0.55 |  |
| Polman 1995 | Senegal |  | 0.91 |  | s-CAA: 0.94<br>u-CCA: 0.95 |  |
|  |  |  |  |  | s-CCA: 0.83 |  |
| Pugh 1980 | Nigeria |  | 0,1733 |  | Haem.: 0,1601<br>Protein.: 0,2599 |  |
| Rasendramino 1998 | Madagascar |  | 0.7678 |  | Haem.: 0.7020<br>Protein.: 0.6143<br>Leuko.: 0.4899 |  |
| Robinson 2009 | Southern Sudan |  | 0.03 | [0.025;0.036] | 0.075 |  |
| Rollinson 2005 | Tanzania |  | 0,5393 |  | 0,5036 |  |
| Sarda 1986 | Tanzania |  | 0.2523 |  | Haem.: 0.2531<br>Protein.: 0.3131 |  |
| Savioli 1990 | Tanzania |  | 0.34 |  | 0.29 |  |
| Sellin 1982 | Burkina Faso |  | 0,463 |  | Haem.: 0,7048<br>Protein.: 0,5189 |  |
| Shane 2011 | Kenya |  | 0.388 | [0.0.344;0.433] | 0.624 | [0.576;0.670] |
| Shaw 1998 | Senegal |  | 0,3932 |  | 0,3746 |  |
| Sheele 2013 | Kenya |  | 0.4875 |  | 0.675 |  |
| Song 2018 | Sudan |  | 0.3893 |  | 0.7987 |  |
| Sousa 2020 | Brazil |  | 1KK: 0.055<br>2KK: 0.078<br>16KK: 0.143<br>HTX: 0.244 |  | 0.29 |  |
| Standley 2010 | Kenya & Tanzania |  | 0.686 | [0.607;0.757] | tr(-): 0.713<br>tr(+): 0.942 | tr(-): [0.639;0.788]<br>tr(+): [0.895;0.972] |
| Stephenson 1984 | Kenya |  | 0.476 |  | Haem.: 0.435<br>Protein.: 0.35 |  |
| Stothard 2009a | Tanzania |  | 0.307 | [0.234;0.387] | CCA: 0.04<br>SEA-EL: 0.48 | CCA: 0.015;0.085]<br>SEA-EL:<br>0.398;0.563] |
| Stothard 2009b | Tanzania |  | 0.652 | [0.524;0.765] | 0.773 | [0.653;0.867] |
| Tanner 1983<br>(Liberia) | Liberia |  | 0.7079 |  | Haem.: 0.5206<br>Protein.: 0.4419 |  |
| Tanner 1983<br>(Tanzania) | Tanzania |  | 0.2956 |  | Haem.: 0.3303<br>Protein.: 0.3723 |  |
| Tchuem Tchuente 2012 | Ethiopia |  | 0.430 |  | 0.660 |  |
| Traore 1998 | Nigeria |  | 0.552 |  | Haem.: 0.476<br>Protein.: 0.408 |  |
| Uga 1989 | Kenya |  | 0.78 |  | 0.76 |  |
| Ugbomoiko 2009a | Nigeria |  | 1992: 0.629<br>2006: 0.514 | [0.588;0.670]<br>[0.488;0.540] | 0.583<br>0.465 | [0.542;0.624]<br>[0.429;0.491] |
| Ugbomoiko 2009b | Nigeria |  | 0.508 | [0.46;0.555] | Microhaem.:<br>0.683 |  |
|  |  |  |  |  | Protein.: 0.533 |  |
| Van Lieshout 1995 | Surinam |  | 0.29 | [0.23;0.34] | s-CAA: 0.23<br>u-CAA: 0.03<br>s-CCA: 0.17<br>u-CCA: 0.28 | s-CAA: [0.19;0.28]<br>u-CAA: [0.01;0.04]<br>s-CCA: [0.13;0.21]<br>u-CCA: [0.23;0.33] |
| Verlé 1994 | Senegal |  | 0,8693 |  | Haem.: 0,625<br>Protein.: 0,5369 |  |
| Vonghachack 2017 | Lao People's<br>Democratic<br>Republic<br>Cambodia |  | 0.127<br>0.005 |  | 0.238<br>0.184 |  |
| Wilkins 1979 | Gambia |  | 0,5545 |  | Haem.: 0,3498<br>Protein.: 0,4897 |  |
| Xu 2014 | China |  | 0.05 |  | 0.339 |  |
| Xu 2015 | China |  |  |  | ELISA: 0.845<br>IHA: 0.918<br>LAMP: 0. 945 |  |
| Zhang 2020 | Zambia |  | 0.61 | [0.53;0.69] | DDIA: 0.51<br>IHA: 0.56 | DDIA: [0.43;0.59]<br>IHA: [0.48;0.64] |
| Zumstein 1983 | Tanzania |  | 0.46 |  | 0.81 |  |

#### Number of studies and participants

The number of studies and the number of participants for each study are given in Table 6.

**Table 4.**
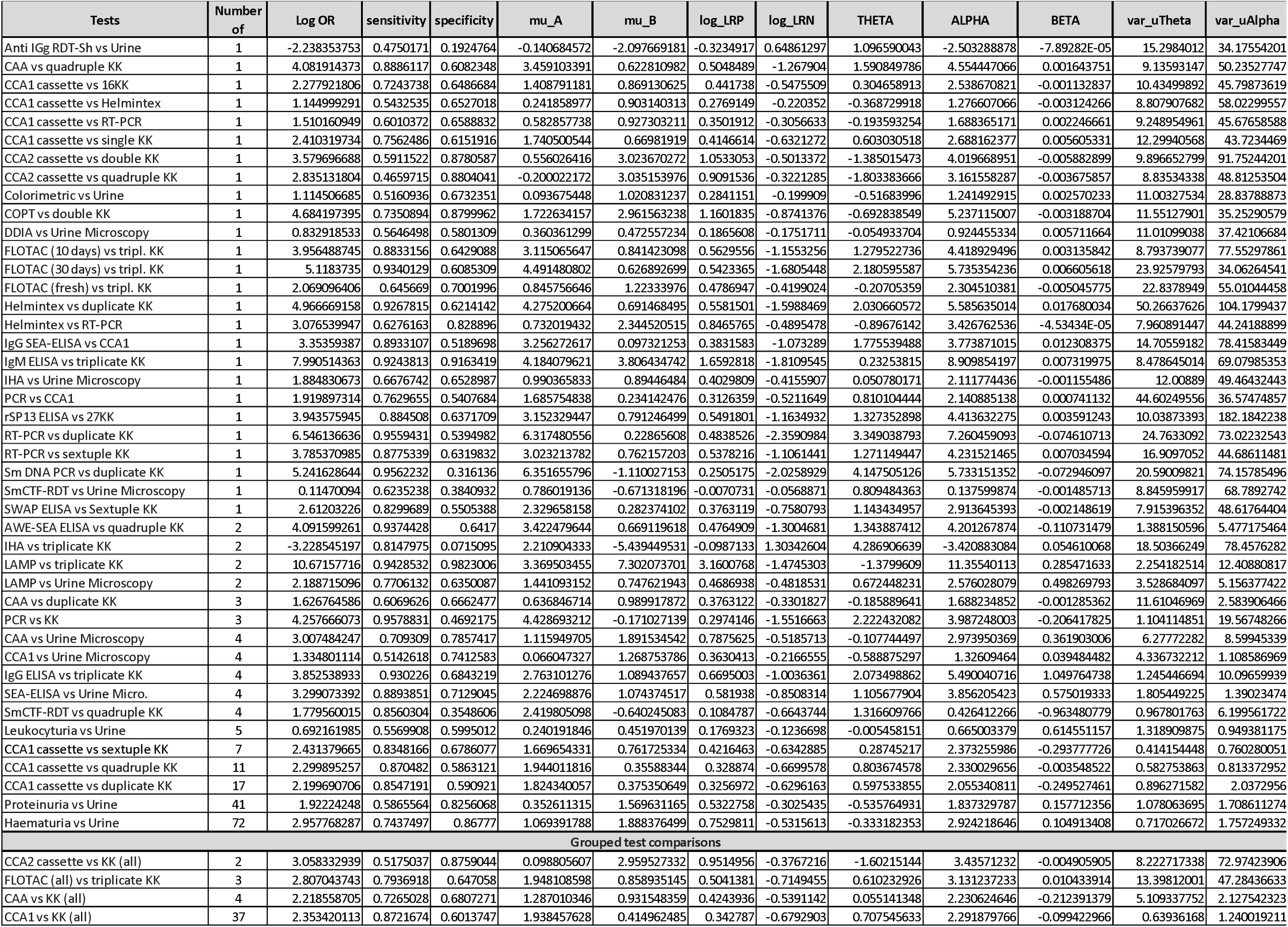
Output parameters of the bayesian bivariate random effects model for each diagnostic test

| Tests | Number of | Log OR | sensitivity | specificity | mu_A | mu_B | log_LRP | log_LRN | THETA | ALPHA | BETA | var_uTheta | var_uAlpha |
| --- | --- | --- | --- | --- | --- | --- | --- | --- | --- | --- | --- | --- | --- |
| Anti IgG RDT-Sh vs Urine | 1 | -2.238353753 | 0.4750171 | 0.1924764 | -0.140684572 | -2.097669181 | -0.3234917 | 0.64861297 | 1.096590043 | -2.503288878 | -7.89282E-05 | 15.2984012 | 34.17554201 |
| CAA vs quadruple KK | 1 | 4.081914373 | 0.8886117 | 0.6082348 | 3.459103391 | 0.622810982 | 0.5048489 | -1.267904 | 1.590849786 | 4.554447066 | 0.001643751 | 9.13593147 | 50.23527747 |
| CCA1 cassette vs 16KK | 1 | 2.277921806 | 0.7243738 | 0.6486684 | 1.408791181 | 0.869130625 | 0.441738 | -0.5475509 | 0.304658913 | 2.538670821 | -0.001132837 | 10.43499892 | 45.79873619 |
| CCA1 cassette vs Helmintex | 1 | 1.144999291 | 0.5432535 | 0.6527018 | 0.241858977 | 0.903140313 | 0.2769149 | -0.220352 | -0.368729918 | 1.276607066 | -0.003124266 | 8.807907682 | 58.02299557 |
| CCA1 cassette vs RT-PCR | 1 | 1.510160949 | 0.6010372 | 0.6588832 | 0.582857738 | 0.927303211 | 0.3501912 | -0.3056633 | -0.193593254 | 1.688365171 | 0.002246661 | 9.248954961 | 45.67658588 |
| CCA1 cassette vs single KK | 1 | 2.410319734 | 0.7562486 | 0.6151916 | 1.740500544 | 0.66981919 | 0.4146614 | -0.6321272 | 0.603030518 | 2.688162377 | 0.005605331 | 12.29940568 | 43.7234469 |
| CCA2 cassette vs double KK | 1 | 3.579696688 | 0.5911522 | 0.8780587 | 0.556026416 | 3.023670272 | 1.0533053 | -0.5013372 | -1.385015473 | 4.019668951 | -0.005882899 | 9.896652799 | 91.75244201 |
| CCA2 cassette vs quadruple KK | 1 | 2.835131804 | 0.4659715 | 0.8804041 | -0.200022172 | 3.035153976 | 0.9091536 | -0.3221285 | -1.803383666 | 3.161558287 | -0.003675857 | 8.83534338 | 48.81253504 |
| Colorimetric vs Urine | 1 | 1.114506685 | 0.5160936 | 0.6732351 | 0.093675448 | 1.020831237 | 0.2841151 | -0.199909 | -0.51683996 | 1.241492915 | 0.002570233 | 11.00327534 | 28.83788873 |
| COPT vs double KK | 1 | 4.684197395 | 0.7350894 | 0.8799962 | 1.722634157 | 2.961563238 | 1.1601835 | -0.8741376 | -0.692838549 | 5.237115007 | -0.003188704 | 11.55127901 | 35.25290579 |
| DDiA vs Urine Microscopy | 1 | 0.832918533 | 0.5646498 | 0.5801309 | 0.360361299 | 0.472557234 | 0.1865608 | -0.1751711 | -0.054933704 | 0.924455334 | 0.005711664 | 11.01099038 | 37.42106684 |
| FLOTAC (10 days) vs tripl. KK | 1 | 3.956488745 | 0.8833156 | 0.6429088 | 3.115065647 | 0.841423098 | 0.5629556 | -1.1553256 | 1.279522736 | 4.418929496 | 0.003135842 | 8.793739077 | 77.55297861 |
| FLOTAC (30 days) vs tripl. KK | 1 | 5.1183735 | 0.9340129 | 0.6085309 | 4.491480802 | 0.626892699 | 0.5423365 | -1.6805448 | 2.180595587 | 5.735354236 | 0.006605618 | 23.92579793 | 34.06264541 |
| FLOTAC (fresh) vs tripl. KK | 1 | 2.069096406 | 0.645669 | 0.7001996 | 0.845756646 | 1.22333976 | 0.4786947 | -0.4199024 | -0.20705359 | 2.304510381 | -0.005045775 | 22.8378949 | 55.01044458 |
| Helmintex vs duplicate KK | 1 | 4.966669158 | 0.9267815 | 0.6214142 | 4.275200664 | 0.691468495 | 0.5581501 | -1.5988469 | 2.030660572 | 5.585635014 | 0.017680034 | 50.26637626 | 104.1799437 |
| Helmintex vs RT-PCR | 1 | 3.076539947 | 0.6276163 | 0.828896 | 0.732019432 | 2.344520515 | 0.8465765 | -0.4895478 | -0.89676142 | 3.426762536 | -4.53434E-05 | 7.960891447 | 44.24188899 |
| IgG SEA-ELISA vs CCA1 | 1 | 3.35359387 | 0.8933107 | 0.5189698 | 3.256272617 | 0.097321253 | 0.3831583 | -1.073289 | 1.775539488 | 3.773871015 | 0.012308375 | 14.70559182 | 78.41583449 |
| IgM ELISA vs triplicate KK | 1 | 7.990514363 | 0.9243813 | 0.9163419 | 4.184079621 | 3.806434742 | 1.6592818 | -1.8109545 | 0.23253815 | 8.909854197 | 0.007319975 | 8.478645014 | 69.07985353 |
| IHA vs Urine Microscopy | 1 | 1.884830673 | 0.6676742 | 0.6528987 | 0.990365833 | 0.89446484 | 0.4029809 | -0.4155907 | 0.050780171 | 2.111774436 | -0.001155486 | 12.00889 | 49.46432443 |
| PCR vs CCA1 | 1 | 1.919897314 | 0.7629655 | 0.5407684 | 1.685754838 | 0.234142476 | 0.3126359 | -0.5211649 | 0.810104444 | 2.140885138 | 0.000741132 | 44.60249556 | 36.57474857 |
| rSP 13 ELISA vs 27KK | 1 | 3.943575945 | 0.884508 | 0.6371709 | 3.152329447 | 0.791246499 | 0.5491801 | -1.1634932 | 1.327352898 | 4.413632275 | 0.003591243 | 10.03873393 | 182.1842238 |
| RT-PCR vs duplicate KK | 1 | 6.546136636 | 0.9559431 | 0.5394982 | 6.317480556 | 0.22865608 | 0.4838526 | -2.3590984 | 3.349038793 | 7.260459093 | -0.074610713 | 24.7633092 | 73.02232543 |
| RT-PCR vs sextuple KK | 1 | 3.785370985 | 0.8775339 | 0.6319832 | 3.023213782 | 0.762157203 | 0.5378216 | -1.1061441 | 1.271149447 | 4.231521465 | 0.007034594 | 16.9097052 | 44.68611481 |
| Sm DNA PCR vs duplicate KK | 1 | 5.241628644 | 0.9562232 | 0.316136 | 6.351655796 | -1.110027153 | 0.2505175 | -2.0258929 | 4.147505126 | 5.733151352 | -0.072946097 | 20.59009821 | 74.15785496 |
| SmCTF-RDT vs Urine Microscopy | 1 | 0.11470094 | 0.6235238 | 0.3840932 | 0.786019136 | -0.671318196 | -0.0070731 | -0.0568871 | 0.809484363 | 0.137599874 | -0.001485713 | 8.845959917 | 68.7892742 |
| SWAP ELISA vs Sextuple KK | 1 | 2.61203226 | 0.8299689 | 0.5505388 | 2.329658158 | 0.282374102 | 0.3763119 | -0.7580793 | 1.143434957 | 2.913645393 | -0.002148619 | 7.915396352 | 48.61764404 |
| AWE-SEA ELISA vs quadruple KK | 2 | 4.091599261 | 0.9374428 | 0.6417 | 3.422479644 | 0.669119618 | 0.4764909 | -1.3004681 | 1.343887412 | 4.201267874 | -0.110731479 | 1.388150596 | 5.477175464 |
| IHA vs triplicate KK | 2 | -3.228545197 | 0.8147975 | 0.0715095 | 2.210904333 | -5.439449531 | -0.0987133 | 1.30342604 | 4.286906639 | -3.420883084 | 0.054610068 | 18.50366249 | 78.4576282 |
| LAMP vs triplicate KK | 2 | 10.67157716 | 0.9428532 | 0.9823006 | 3.369503455 | 7.302073701 | 3.1600768 | -1.4745303 | -1.3799609 | 11.35540113 | 0.285471633 | 2.254182514 | 12.40880817 |
| LAMP vs Urine Microscopy | 2 | 2.188715096 | 0.7706132 | 0.6350087 | 1.441093152 | 0.747621943 | 0.4686938 | -0.4818531 | 0.672448231 | 2.576028079 | 0.498269793 | 3.528684097 | 5.156377422 |
| CAA vs duplicate KK | 3 | 1.626764586 | 0.6069626 | 0.6662477 | 0.636846714 | 0.989917872 | 0.3763122 | -0.3301827 | -0.185889641 | 1.688234852 | -0.001285362 | 11.61046969 | 2.583906466 |
| PCR vs KK | 3 | 4.257666073 | 0.9578831 | 0.4692175 | 4.428693212 | -0.171027139 | 0.2974146 | -1.5516663 | 2.222432082 | 3.987248003 | -0.206417825 | 1.104114851 | 19.56748266 |
| CAA vs Urine Microscopy | 4 | 3.007484247 | 0.709309 | 0.7857417 | 1.115949705 | 1.891534542 | 0.7875625 | -0.5185713 | -0.107744497 | 2.973950369 | 0.361903006 | 6.27772282 | 8.59945339 |
| CCA1 vs Urine Microscopy | 4 | 1.334801114 | 0.5142618 | 0.7412583 | 0.066047327 | 1.268753786 | 0.3630413 | -0.2166555 | -0.588875297 | 1.32609464 | 0.039484482 | 4.336732212 | 1.108586969 |
| IgG ELISA vs triplicate KK | 4 | 3.852538933 | 0.930226 | 0.6843219 | 2.763101276 | 1.089437657 | 0.6695003 | -1.0036361 | 2.073498862 | 5.490040716 | 1.049764738 | 1.245446694 | 10.09659939 |
| SEA-ELISA vs Urine Micro. | 4 | 3.299073392 | 0.8893851 | 0.7129045 | 2.224698876 | 1.074374517 | 0.581938 | -0.8508314 | 1.105677904 | 3.856205423 | 0.575019333 | 1.805449225 | 1.39023474 |
| SmCTF-RDT vs quadruple KK | 4 | 1.779560015 | 0.8560304 | 0.3548606 | 2.419805098 | -0.640245083 | 0.1084787 | -0.6643744 | 1.316609766 | 0.426412266 | -0.963480779 | 0.967801763 | 6.199561722 |
| Leukocyturia vs Urine | 5 | 0.692161985 | 0.5569908 | 0.5995012 | 0.240191846 | 0.451970139 | 0.1769323 | -0.1236698 | -0.005458151 | 0.665003379 | 0.614551157 | 1.318909875 | 0.949381175 |
| CCA1 cassette vs sextuple KK | 7 | 2.431379665 | 0.8348166 | 0.6786077 | 1.669654331 | 0.761725334 | 0.4216463 | -0.6342885 | 0.28745217 | 2.373255986 | -0.293777726 | 0.414154448 | 0.760280051 |
| CCA1 cassette vs quadruple KK | 11 | 2.299895257 | 0.870482 | 0.5863121 | 1.944011816 | 0.35588344 | 0.328874 | -0.6699578 | 0.803674578 | 2.330029656 | -0.003548522 | 0.582753863 | 0.813372952 |
| CCA1 cassette vs duplicate KK | 17 | 2.199690706 | 0.8547191 | 0.590921 | 1.824340057 | 0.375350649 | 0.3256972 | -0.6296163 | 0.597533855 | 2.055340811 | -0.249527461 | 0.896271582 | 2.0372956 |
| Proteinuria vs Urine | 41 | 1.92224248 | 0.5865564 | 0.8256068 | 0.352611315 | 1.569631165 | 0.5322758 | -0.3025435 | -0.535764931 | 1.837329787 | 0.157712356 | 1.078063695 | 1.708611274 |
| Haematuria vs Urine | 72 | 2.957768287 | 0.7437497 | 0.86777 | 1.069391788 | 1.888376499 | 0.7529811 | -0.5315613 | -0.333182353 | 2.924218646 | 0.104913408 | 0.717026672 | 1.757249332 |
| Grouped test comparisons |  |  |  |  |  |  |  |  |  |  |  |  |  |
| CCA2 cassette vs KK (all) | 2 | 3.058332939 | 0.5175037 | 0.8759044 | 0.098805607 | 2.959527332 | 0.9514956 | -0.3767216 | -1.60215144 | 3.43571232 | -0.004905905 | 8.222717338 | 72.97423906 |
| FLOTAC (all) vs triplicate KK | 3 | 2.807043743 | 0.7936918 | 0.647058 | 1.948108598 | 0.858935145 | 0.5041381 | -0.7149455 | 0.610232926 | 3.131237233 | 0.010433914 | 13.39812001 | 47.28436633 |
| CAA vs KK (all) | 4 | 2.218558705 | 0.7265028 | 0.6807271 | 1.287010346 | 0.931548359 | 0.4243936 | -0.5391142 | 0.055141348 | 2.230624646 | -0.212391379 | 5.109337752 | 2.127542323 |
| CCA1 vs KK (all) | 37 | 2.353420113 | 0.8721674 | 0.6013747 | 1.938457628 | 0.414962485 | 0.342787 | -0.6792903 | 0.707545633 | 2.291879766 | -0.099422966 | 0.63936168 | 1.240019211 |

**Table 5.** Sensitivities and specificities from the bayesian bivariate random effects model for each diagnostic test

| Tests | Number of studies | Sensitivity |  |  | Specificity |  |  |
| --- | --- | --- | --- | --- | --- | --- | --- |
|  |  | Se | LL 2.5% eCI | UL 97.5% eCI | Sp | LL 2.5% eCI | UL 97.5% eCI |
| Anti IgG RDT-Sh vs Urine | 1 | 0.4750171 | 0.0049109 | 0.993815455 | 0.1924764 | 0.001032063 | 0.972541853 |
| CAA vs quadruple KK | 1 | 0.8886117 | 0.0734737 | 0.999774085 | 0.6082348 | 0.010394884 | 0.996604095 |
| CCA1 cassette vs 16KK | 1 | 0.7243738 | 0.0159298 | 0.998315315 | 0.6486684 | 0.011873868 | 0.997336831 |
| CCA1 cassette vs Helmintex | 1 | 0.5432535 | 0.0060719 | 0.995681455 | 0.6527018 | 0.01029001 | 0.997782925 |
| CCA1 cassette vs RT-PCR | 1 | 0.6010372 | 0.0089075 | 0.996671064 | 0.6588832 | 0.010627548 | 0.99735831 |
| CCA1 cassette vs single KK | 1 | 0.7562486 | 0.0200934 | 0.998821622 | 0.6151916 | 0.010061895 | 0.997084593 |
| CCA2 cassette vs double KK | 1 | 0.5911522 | 0.0071908 | 0.99698671 | 0.8780587 | 0.051948794 | 0.99948796 |
| CCA2 cassette vs quadruple KK | 1 | 0.4659715 | 0.004732 | 0.993631655 | 0.8804041 | 0.07237841 | 0.999506175 |
| Colorimetric vs Urine | 1 | 0.5160936 | 0.0059616 | 0.994874188 | 0.6732351 | 0.012315758 | 0.99771679 |
| COPT vs double KK | 1 | 0.7350894 | 0.0183996 | 0.999168593 | 0.8799962 | 0.071662891 | 0.999419096 |
| DDIA vs Urine Microscopy | 1 | 0.5646498 | 0.0062138 | 0.996332453 | 0.5801309 | 0.007168676 | 0.997244909 |
| FLOTAC (10 days) vs tripl. KK | 1 | 0.8833156 | 0.0678575 | 0.999504214 | 0.6429088 | 0.012737015 | 0.997002063 |
| FLOTAC (30 days) vs tripl. KK | 1 | 0.9340129 | 0.1654421 | 0.999859688 | 0.6085309 | 0.011458981 | 0.996324903 |
| FLOTAC (fresh) vs tripl. KK | 1 | 0.645669 | 0.0104418 | 0.997094993 | 0.7001996 | 0.015566698 | 0.998048114 |
| Helmintex vs duplicate KK | 1 | 0.9267815 | 0.1566279 | 0.999850753 | 0.6214142 | 0.010191859 | 0.996552959 |
| Helmintex vs RT-PCR | 1 | 0.6276163 | 0.0096701 | 0.996927567 | 0.828896 | 0.031694518 | 0.999196901 |
| IgG SEA-ELISA vs CCA1 | 1 | 0.8933107 | 0.0707431 | 0.999561351 | 0.5189698 | 0.005307201 | 0.994896931 |
| IgM ELISA vs triplicate KK | 1 | 0.9243813 | 0.1467481 | 0.999836095 | 0.9163419 | 0.125765352 | 0.999715443 |
| IHA vs Urine Microscopy | 1 | 0.6676742 | 0.0116374 | 0.997553444 | 0.6528987 | 0.011296826 | 0.997251355 |
| PCR vs CCA1 | 1 | 0.7629655 | 0.022043 | 0.998669822 | 0.5407684 | 0.006448133 | 0.995846675 |
| rSP13 ELISA vs 27KK | 1 | 0.884508 | 0.0650399 | 0.999537488 | 0.6371709 | 0.010320459 | 0.99722862 |
| RT-PCR vs duplicate KK | 1 | 0.9559431 | 0.235933 | 0.999946861 | 0.5394982 | 0.003054185 | 0.997756114 |
| RT-PCR vs sextuple KK | 1 | 0.8775339 | 0.0637974 | 0.999513105 | 0.6319832 | 0.010406706 | 0.996953866 |
| Sm DNA PCR vs duplicate KK | 1 | 0.9562232 | 0.2298923 | 0.999947103 | 0.316136 | 0.001208631 | 0.996315402 |
| SmCTF-RDT vs Urine Microscopy | 1 | 0.6235238 | 0.0092144 | 0.997516474 | 0.3840932 | 0.003031678 | 0.990146401 |
| SWAP ELISA vs Sextuple KK | 1 | 0.8299689 | 0.0353961 | 0.99911491 | 0.5505388 | 0.006669649 | 0.995520526 |
| AWE-SEA ELISA vs quadruple KK | 2 | 0.9374428 | 0.663782 | 0.998324929 | 0.6417 | 0.185606389 | 0.943594863 |
| IHA vs triplicate KK | 2 | 0.8147975 | 0.0194877 | 0.999352904 | 0.0715095 | 5.5441E-05 | 0.865398383 |
| LAMP vs triplicate KK | 2 | 0.9428532 | 0.7019744 | 0.997337756 | 0.9823006 | 0.879388278 | 0.999949756 |
| LAMP vs Urine Microscopy | 2 | 0.7706132 | 0.2678742 | 0.979544766 | 0.6350087 | 0.040074085 | 0.989892082 |
| CAA vs duplicate KK | 3 | 0.6069626 | 0.024503 | 0.994050292 | 0.6662477 | 0.029325523 | 0.995140491 |
| PCR vs KK | 3 | 0.9578831 | 0.6327875 | 0.999806283 | 0.4692175 | 0.047322874 | 0.918938689 |
| CAA vs Urine Microscopy | 4 | 0.709309 | 0.1968127 | 0.97561546 | 0.7857417 | 0.142936487 | 0.997654471 |
| CCA1 vs Urine Microscopy | 4 | 0.5142618 | 0.1094359 | 0.900281862 | 0.7412583 | 0.279021452 | 0.973293967 |
| IgG ELISA vs triplicate KK | 4 | 0.930226 | 0.8204302 | 0.98412027 | 0.6843219 | 0.076773596 | 0.990898564 |
| SEA-ELISA vs Urine Micro. | 4 | 0.8893851 | 0.7365351 | 0.971267703 | 0.7129045 | 0.272602542 | 0.959936037 |
| SmCTF-RDT vs quadruple KK | 4 | 0.8560304 | 0.3004736 | 0.998556357 | 0.3548606 | 0.140483256 | 0.630656592 |
| Leukocyturia vs Urine | 5 | 0.5569908 | 0.3447937 | 0.7618991 | 0.5995012 | 0.245456227 | 0.883532101 |
| CCA1 cassette vs sextuple KK | 7 | 0.8348166 | 0.7073517 | 0.920219224 | 0.6786077 | 0.548966403 | 0.793295985 |
| CCA1 cassette vs quadruple KK | 11 | 0.870482 | 0.7855841 | 0.930458304 | 0.5863121 | 0.446411666 | 0.717986875 |
| CCA1 cassette vs duplicate KK | 17 | 0.8547191 | 0.7430488 | 0.931016547 | 0.590921 | 0.447902542 | 0.721719083 |
| Proteinuria vs Urine | 41 | 0.5865564 | 0.4968416 | 0.670114249 | 0.8256068 | 0.759632106 | 0.880166945 |
| Haematuria vs Urine | 72 | 0.7437497 | 0.6953608 | 0.788788909 | 0.86777 | 0.834639646 | 0.896729776 |
| Grouped test comparisons |  |  |  |  |  |  |  |
| CCA2 cassette vs KK (all) | 2 | 0.5175037 | 0.0051457 | 0.995304665 | 0.8759044 | 0.049243157 | 0.999494388 |
| FLOTAC (all) vs triplicate KK | 3 | 0.7936918 | 0.0271207 | 0.998856805 | 0.647058 | 0.009599002 | 0.997441116 |
| CAA vs KK (all) | 4 | 0.7265028 | 0.1403461 | 0.98879899 | 0.6807271 | 0.175192484 | 0.969812396 |
| CCA1 vs KK (all) | 37 | 0.8721674 | 0.8186703 | 0.915510917 | 0.6013747 | 0.508044351 | 0.689495314 |

**Table 6.** Number of studies and number of participants for each test comparison

| Test | Studies | Participants |
| --- | --- | --- |
| 1 CCA1 cassette vs single KK | 1 | 217 |
| 2 CCA1 cassette vs duplicate KK | 14 | 4884 |
| 3 CCA1 cassette vs quadruple KK | 10 | 4592 |
| 4 CCA1 cassette vs sextuple KK | 7 | 2325 |
| 5 CCA1 cassette vs 16KK | 1 | 217 |
| 6 CCA2 cassette vs double KK | 1 | 100 |
| 7 CCA2 cassette vs quadruple KK | 1 | 100 |
| 8 CAA vs duplicate KK | 2 | 830 |
| 9 CAA vs quadruple KK | 1 | 377 |
| 10 FLOTAC (fresh) vs triplicate KK | 1 | 112 |
| 11 FLOTAC (10 days) vs triplicate KK | 1 | 112 |
| 12 FLOTAC (30 days) vs triplicate KK | 1 | 112 |
| 13 SmCTF-RDT vs quadruple KK | 3 | 291 |
| 14 SmCTF-RDT vs Urine Microscopy | 1 | 117 |
| 15 Sm DNA PCR vs duplicate KK | 1 | 89 |
| 16 SWAP ELISA vs Sextuple KK | 1 | 482 |
| 17 IgM ELISA vs triplicate KK | 1 | 137 |
| 18 IgG ELISA vs triplicate KK | 4 | 954 |
| 19 rSP13 ELISA vs 27KK | 1 | 1371 |
| 20 AWE-SEA ELISA vs quadruple KK | 2 | 484 |
| 21 IgG SEA-ELISA vs CCA1 | 1 | 258 |
| 22 IgG SEA-ELISA vs Urine Microscopy | 4 | 503 |
| 23 Anti IGg RDT-Sh vs Urine Microscopy | 1 | 160 |
| 24 Haematuria (R strip) vs Urine Microscopy | 71 | 156279 |
| 25 Proteinuria (R strip) vs Urine Microscopy | 40 | 79466 |
| 26 Leukocyturia vs Urine Microscopy | 5 | 1532 |
| 27 LAMP vs triplicate KK | 2 | 493 |
| 28 LAMP vs Urine Microscopy | 2 | 266 |
| 29 IHA vs triplicate KK | 1 | 203 |
| 30 IHA vs Urine Microscopy | 1 | 146 |
| 31 Colorimetric test vs Urine Microscopy | 1 | 1279 |
| 32 COPT vs double KK | 1 | 572 |
| 33 PCR vs KK | 2 | 551 |
| 34 PCR vs CCA1 | 1 | 258 |
| 35 Helmintex vs duplicate KK | 1 | 461 |
| 36 Helmintex vs RT-PCR | 1 | 176 |
| 37 DDIA vs Urine Microscopy | 1 | 146 |
| 38 CCA1 vs Urine Microscopy | 4 | 991 |
| 39 CCA1 vs Helmintex | 1 | 214 |
| 40 CCA1 vs RT-PCR | 1 | 196 |
| 41 CAA vs Urine Microscopy | 4 | 1247 |
| 42 RT-PCR vs sextuple KK | 1 | 198 |
| 43 RT-PCR vs duplicate KK | 1 | 206 |
| 44 CCA1 vs KK | 25 | 12235 |
| 45 CCA2 vs KK | 1 | 200 |
| 46 CAA vs KK | 3 | 1207 |
| 47 FLOTAC vs KK | 1 | 336 |

#### Forest plots and Summary ROC curves

For every test comparison, the forest plot as well as the SROC curve can be found in Figures 4 – 59.

**Figure 4.**
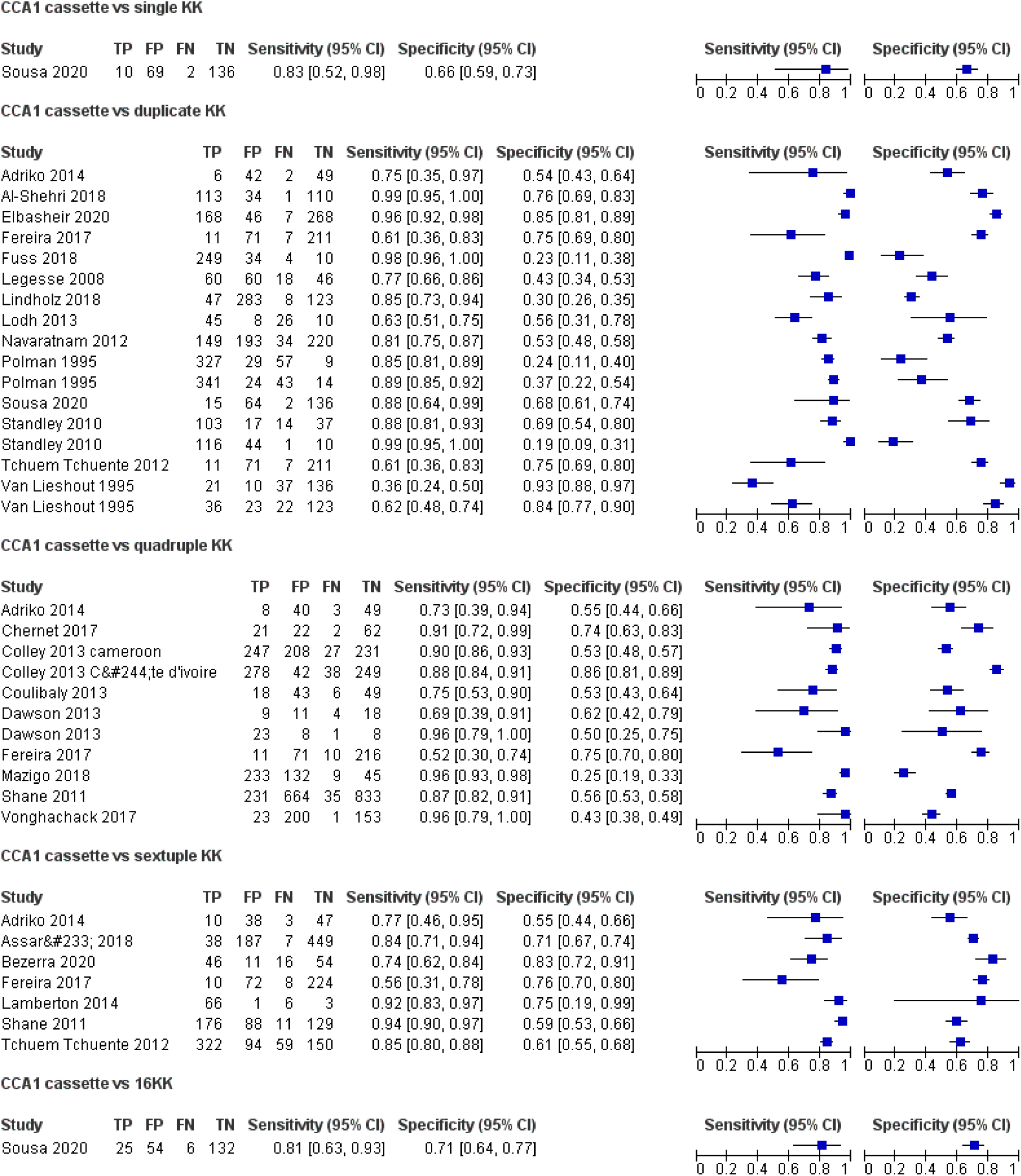
Forest plot of CCA1 cassette vs single KK, double KK, quadruple KK, sextuple KK or 16KK

**Figure 5.**
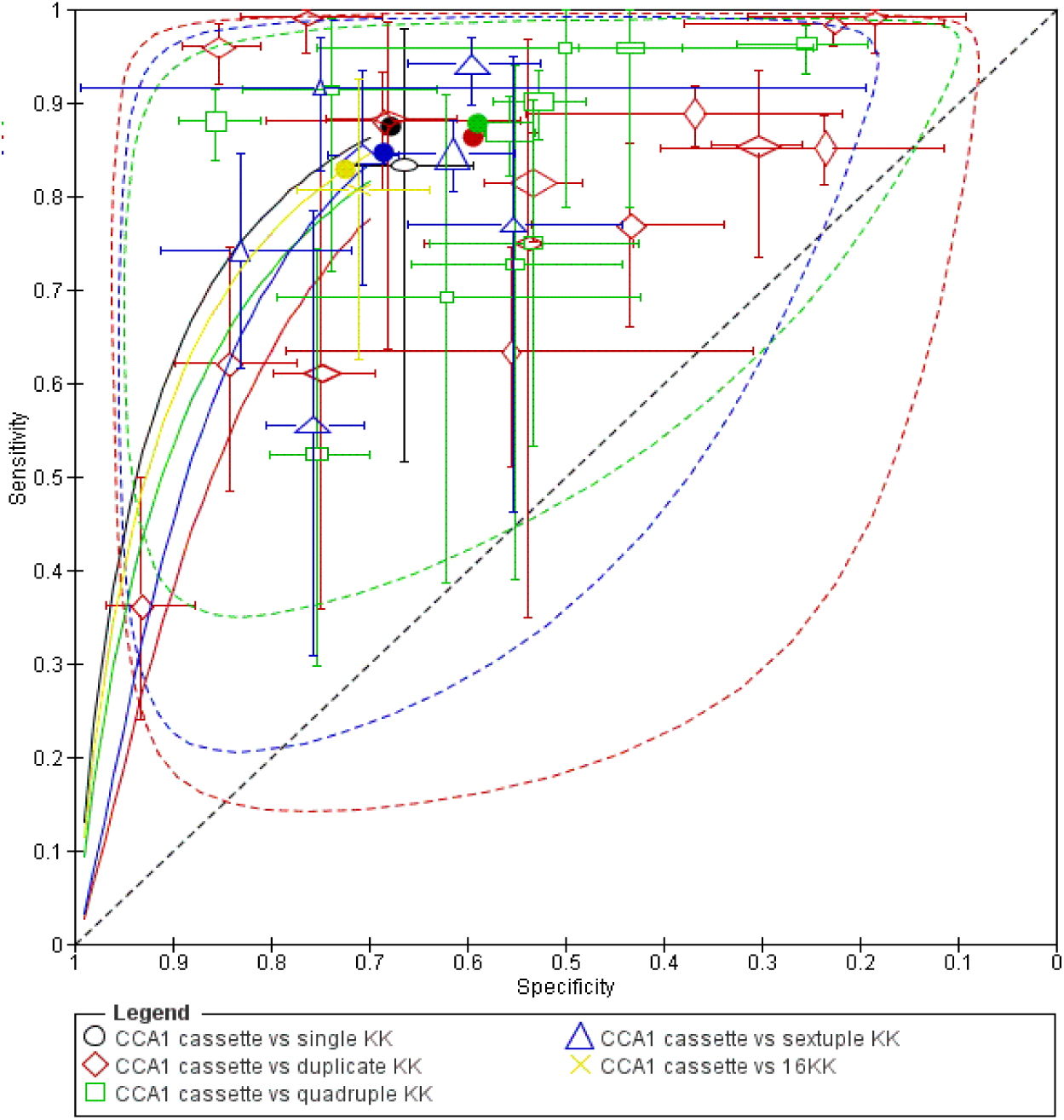
Summary ROC Plot of CCA1 cassette vs single KK, double KK, quadruple KK, sextuple KK or 16KK

**Figure 6.**
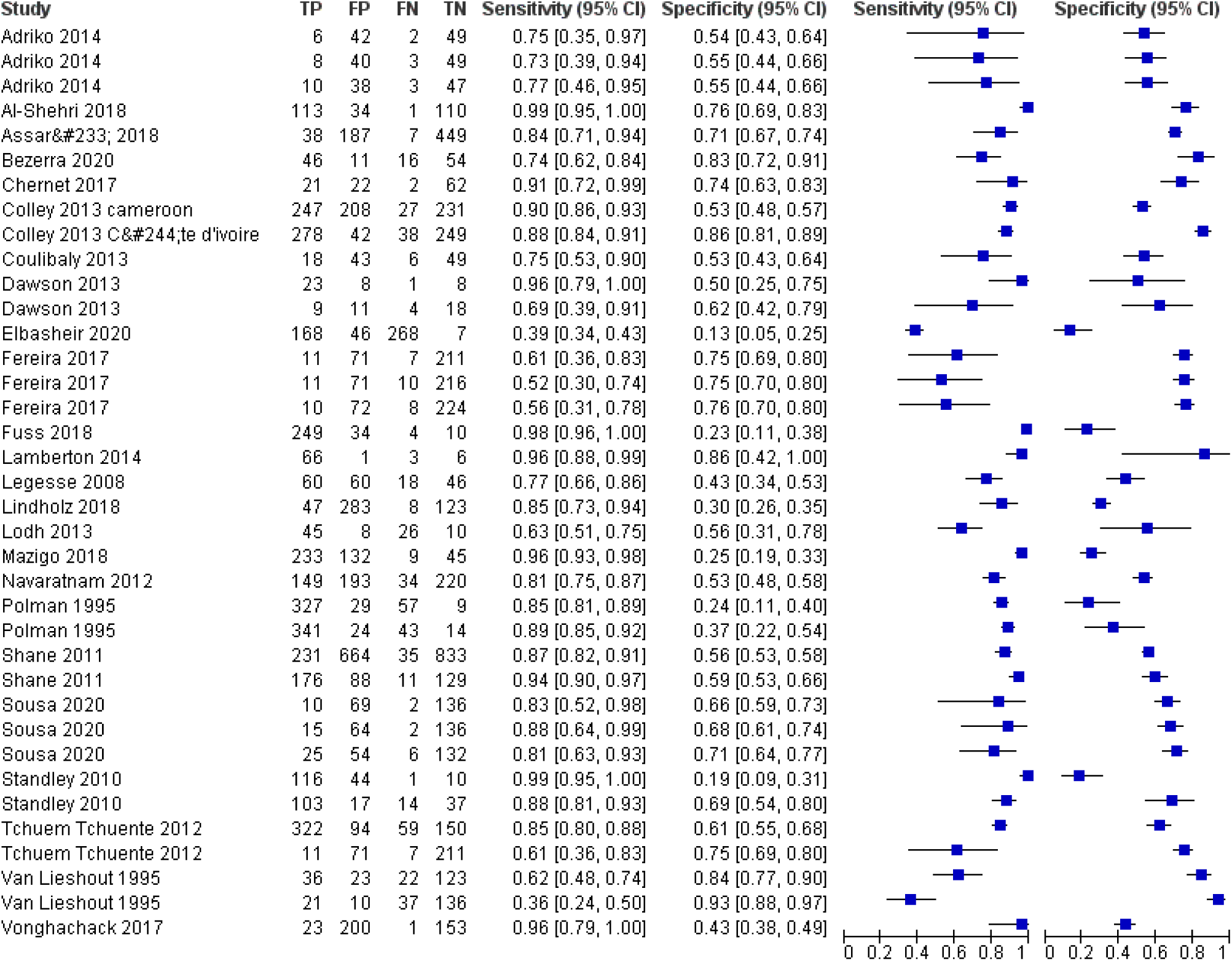
Forest plot of CCA1 cassette vs single, duplicate KK, quadruple KK, sextuple KK and 16KK

**Figure 7.**
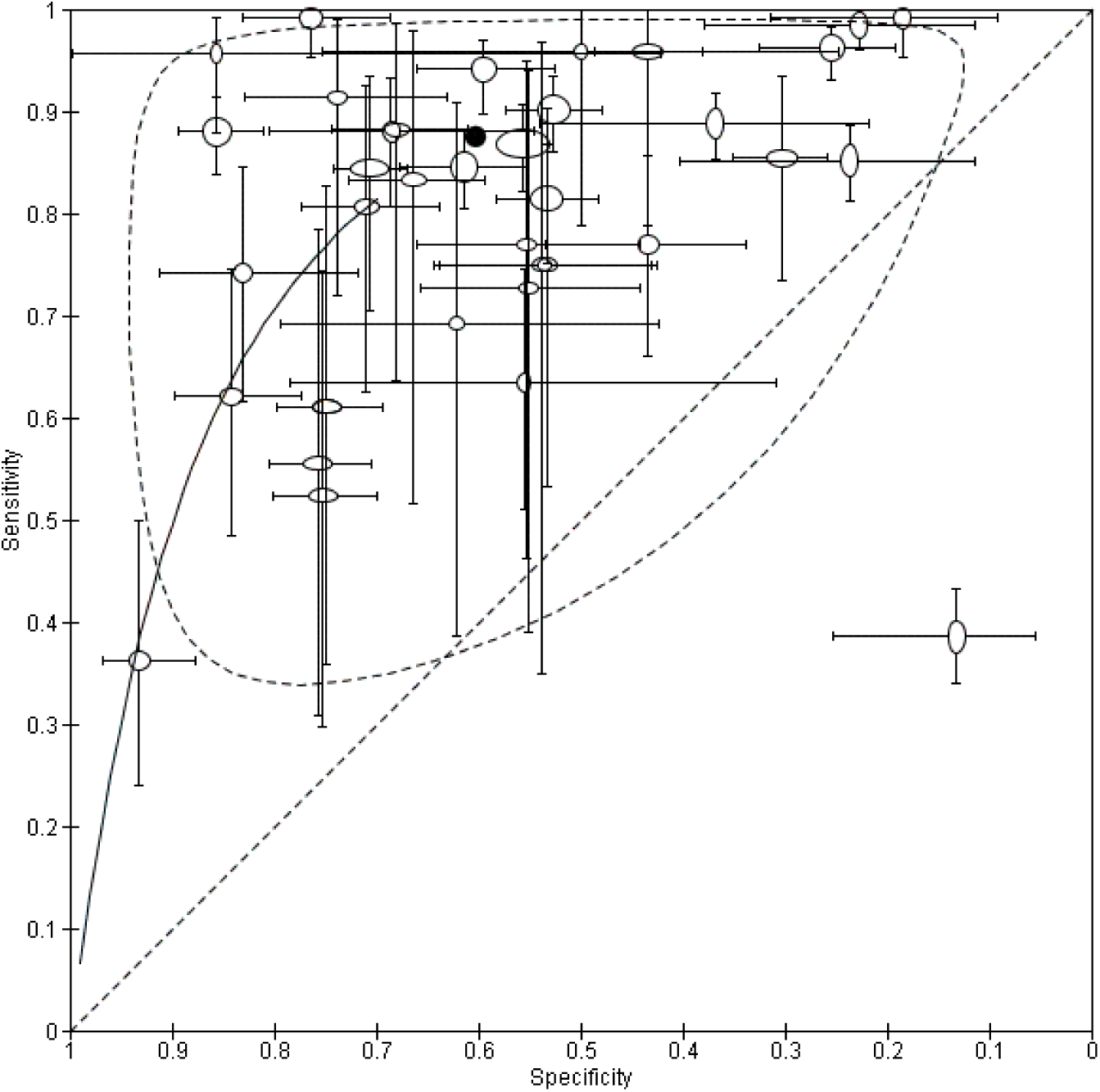
Summary ROC Plot of CCA1 cassette vs single, duplicate KK, quadruple KK, sextuple KK and 16KK

**Figure 8.**
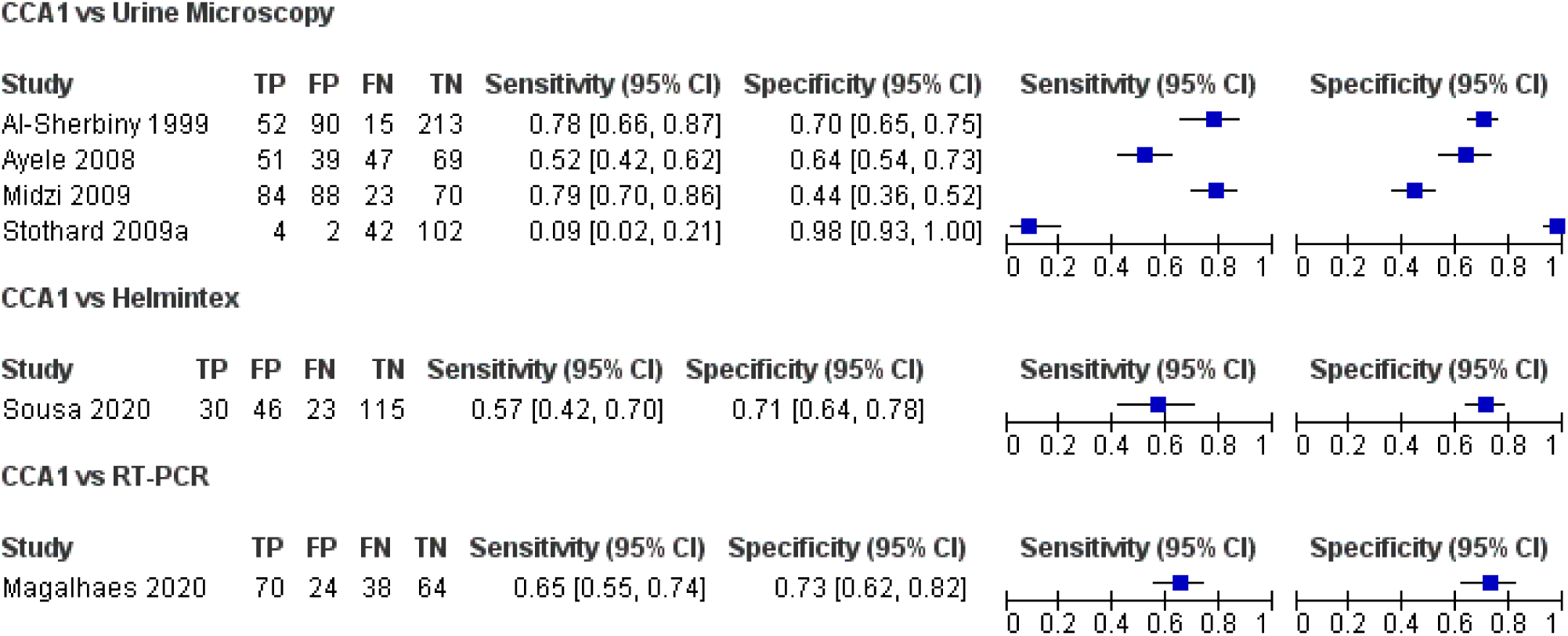
Forest plot of CCA1 cassette vs Urine Microscopy, Helmintex or RTPCR

**Figure 9.**
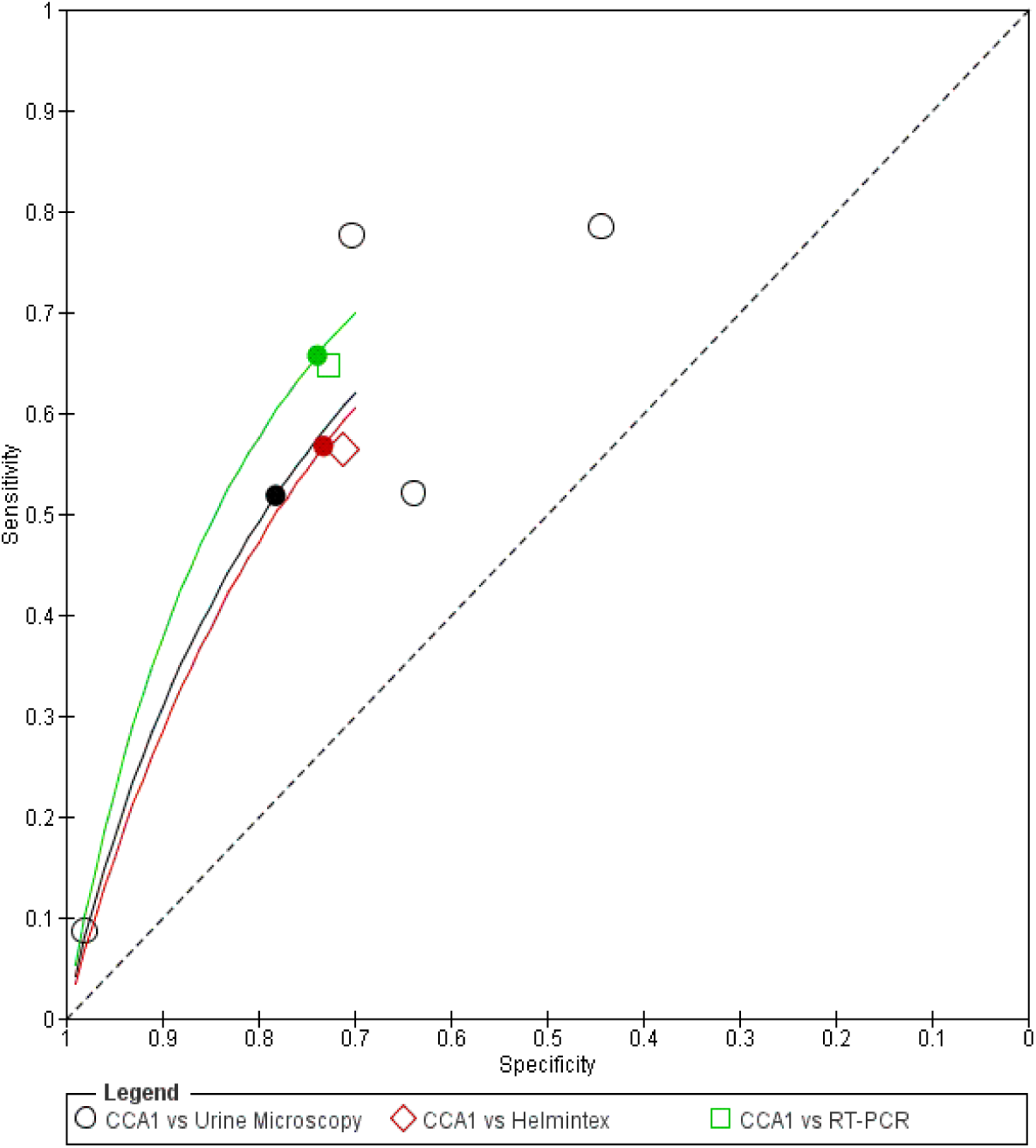
Summary ROC Plot of CCA1 cassette vs Urine Microscopy, Helmintex or RT-PCR

**Figure 10.**
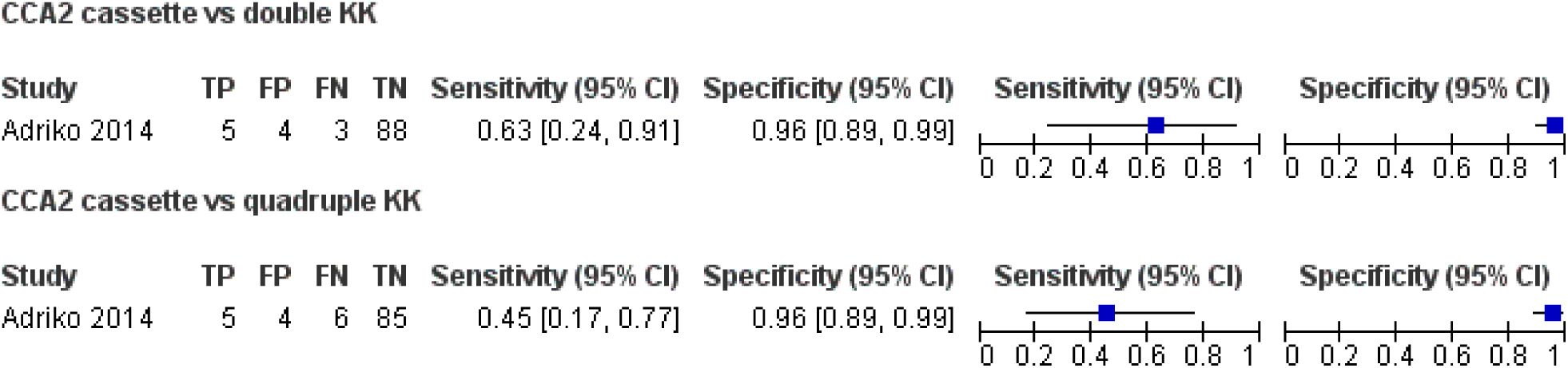
Forest plot of CCA2 cassette vs duplicate KK or quadruple KK

**Figure 11.**
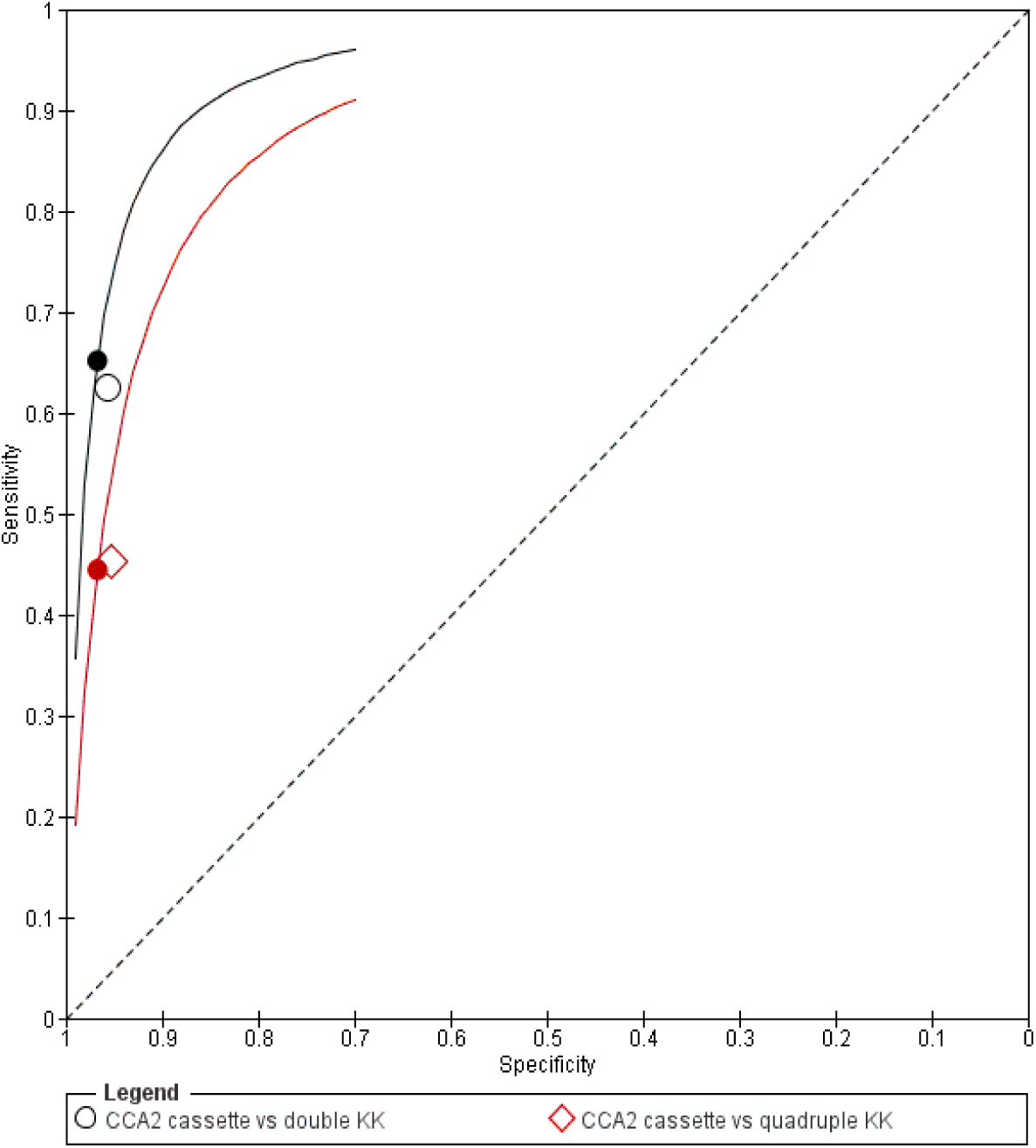
Summary ROC Plot of CCA2 cassette vs duplicate KK or quadruple KK

**Figure 12.**
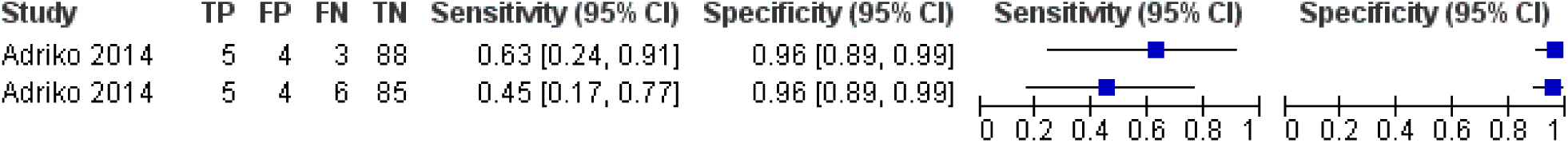
Forest plot of CCA2 cassette vs duplicate KK and quadruple KK

**Figure 13.**
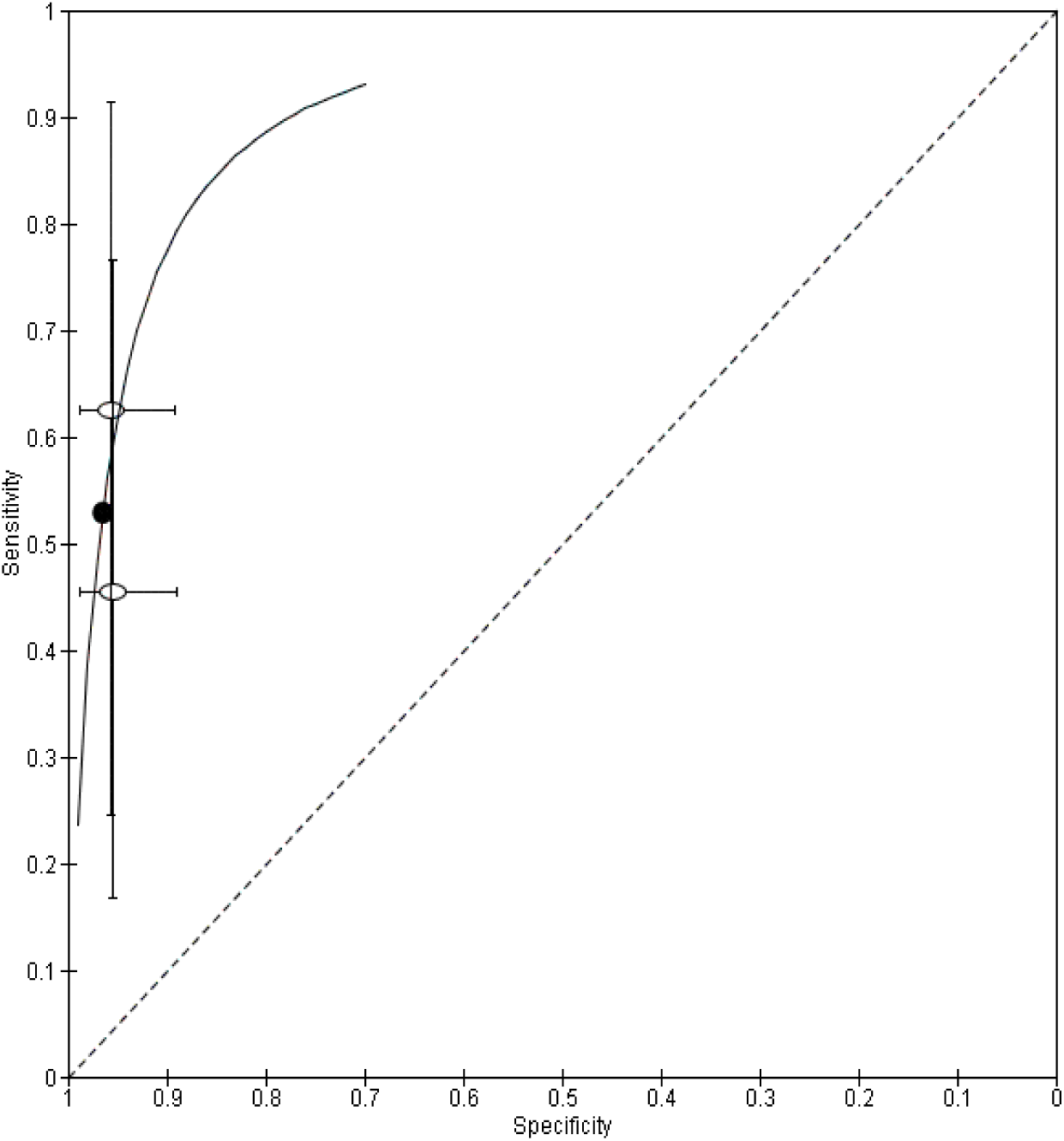
Summary ROC Plot of CCA2 cassette vs duplicate KK and quadruple KK

**Figure 14.**
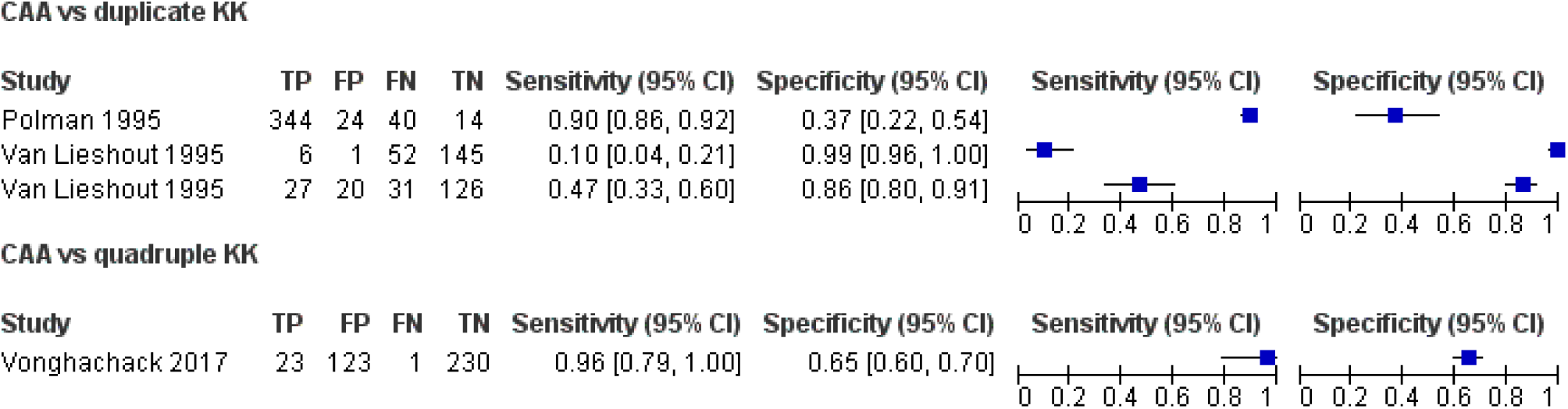
Forest plot of CAA cassette vs duplicate KK or quadruple KK

**Figure 15.**
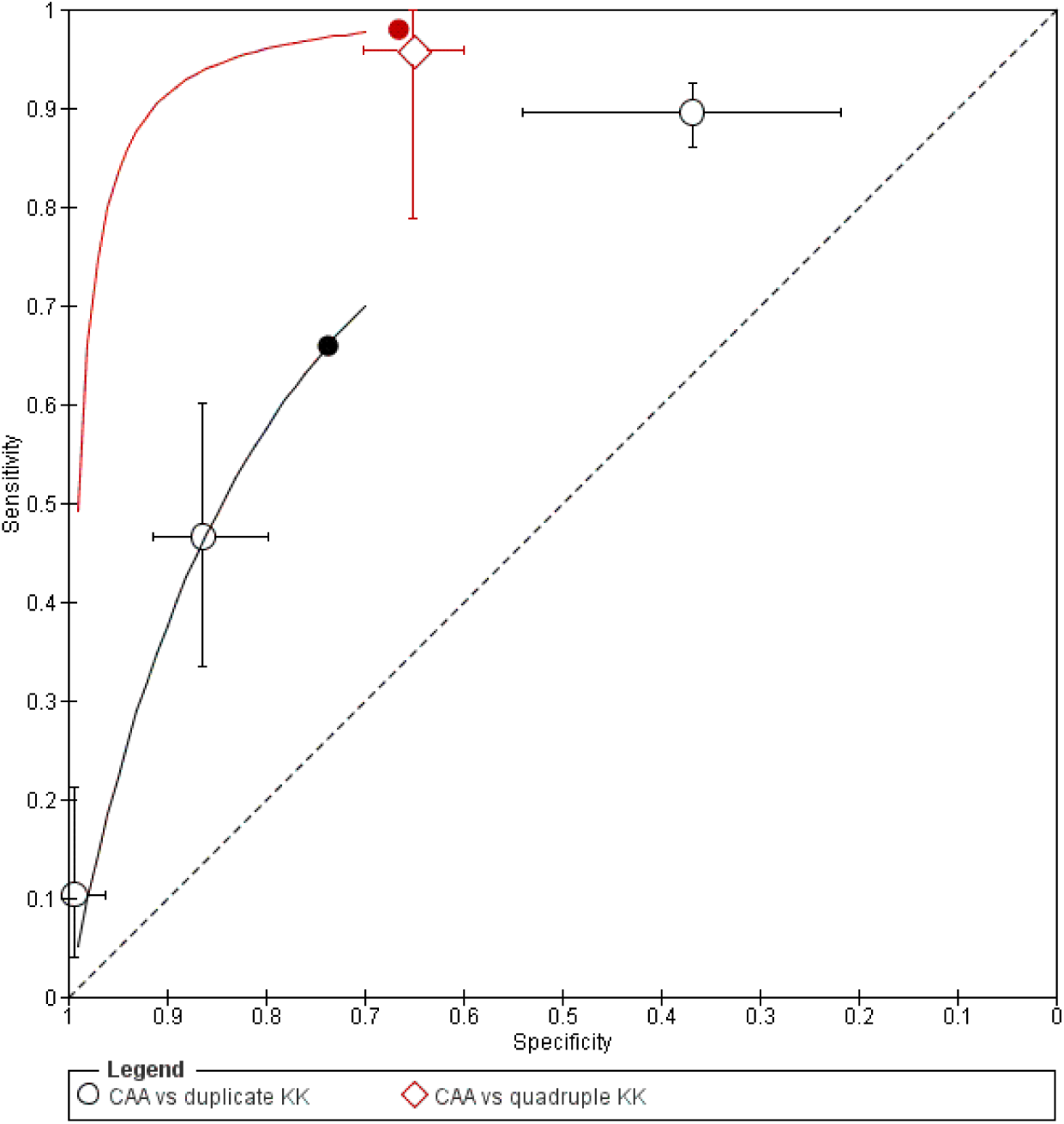
Summary ROC Plot of CAA cassette vs duplicate KK or quadruple KK

**Figure 16.**
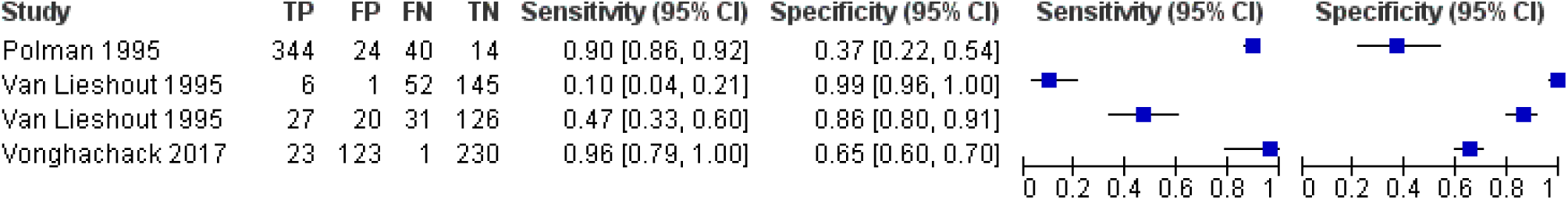
Forest plot of CAA cassette vs duplicate KK and quadruple KK

**Figure 17.**
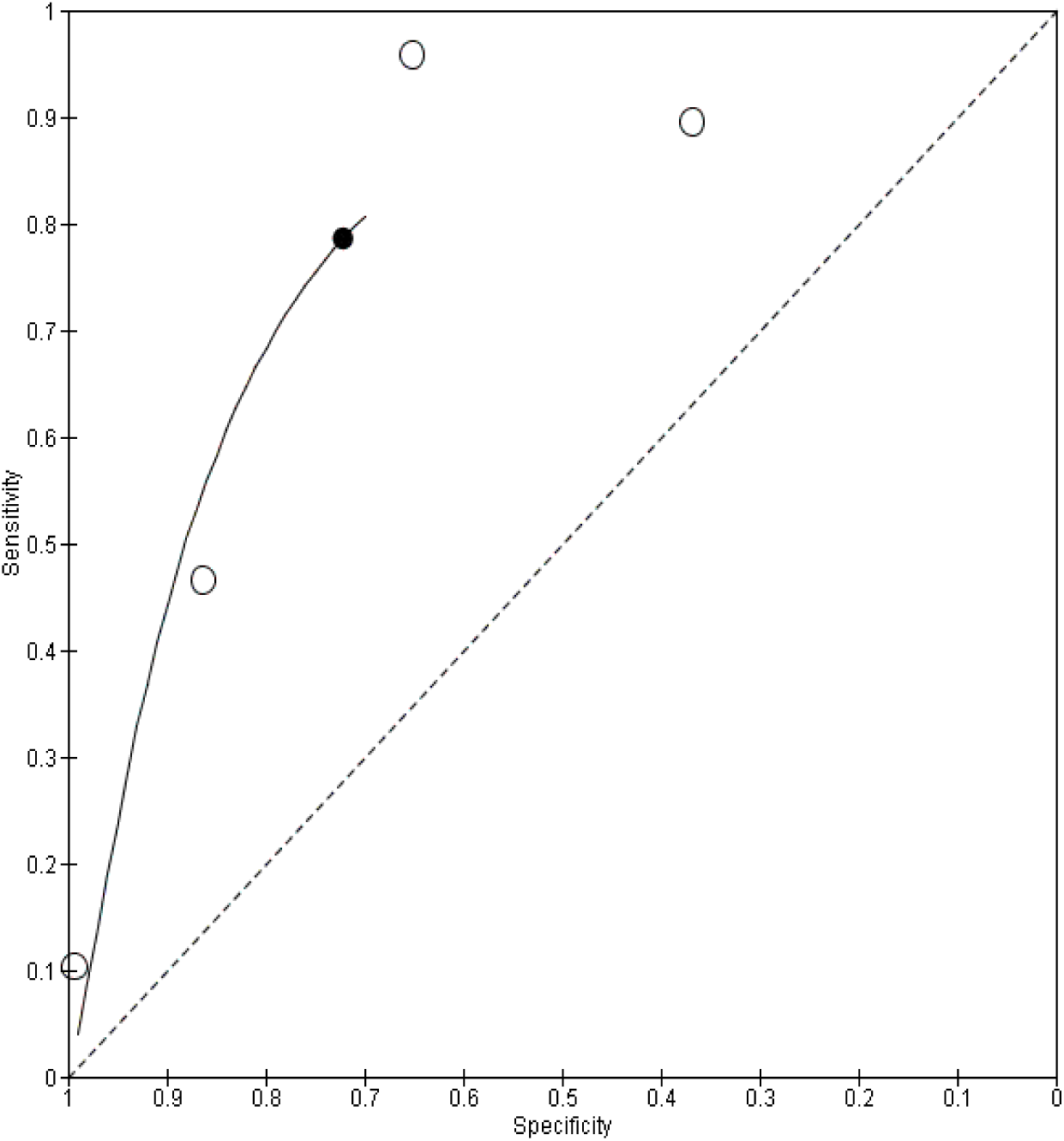
Summary ROC Plot of CAA cassette vs duplicate KK and quadruple KK

**Figure 18.**
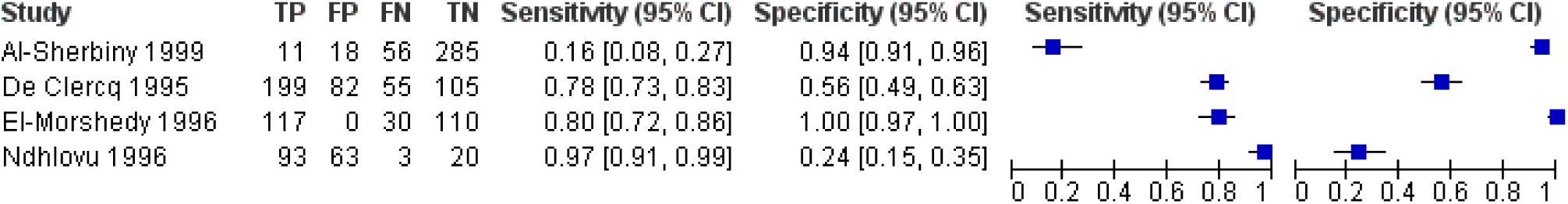
Forest plot of CAA cassette vs Urine Microscopy

**Figure 19.**
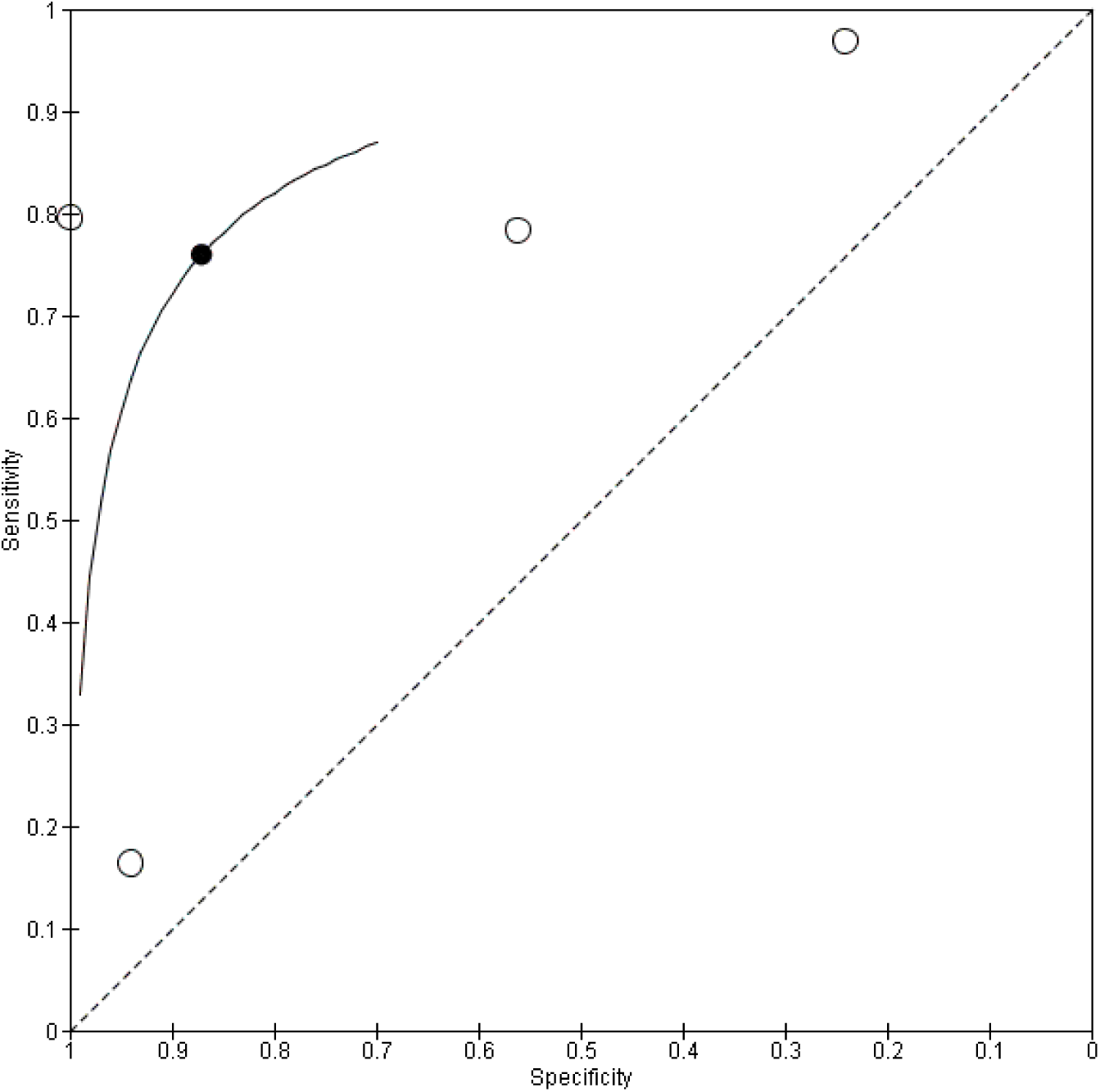
Summary ROC Plot of CAA cassette vs Urine Microscopy

**Figure 20.**
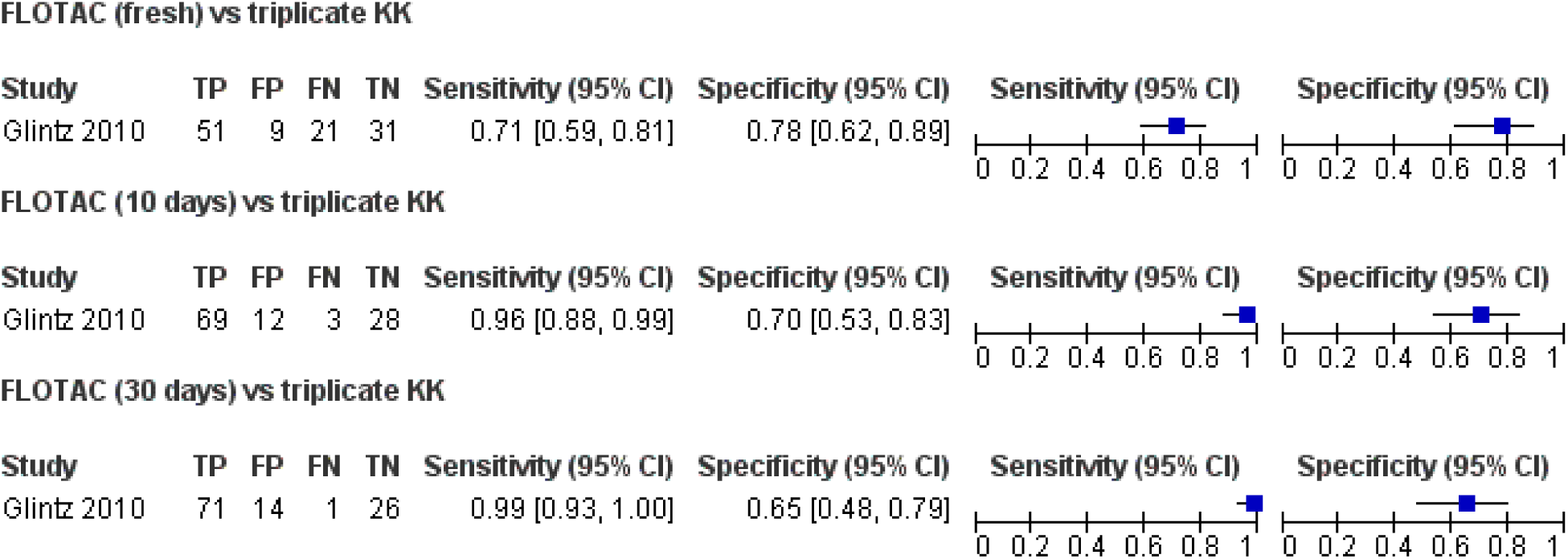
Forest plot of FLOTAC (fresh), FLOTAC (10 days) or FLOTAC (30 days) vs triplicate KK

**Figure 21.**
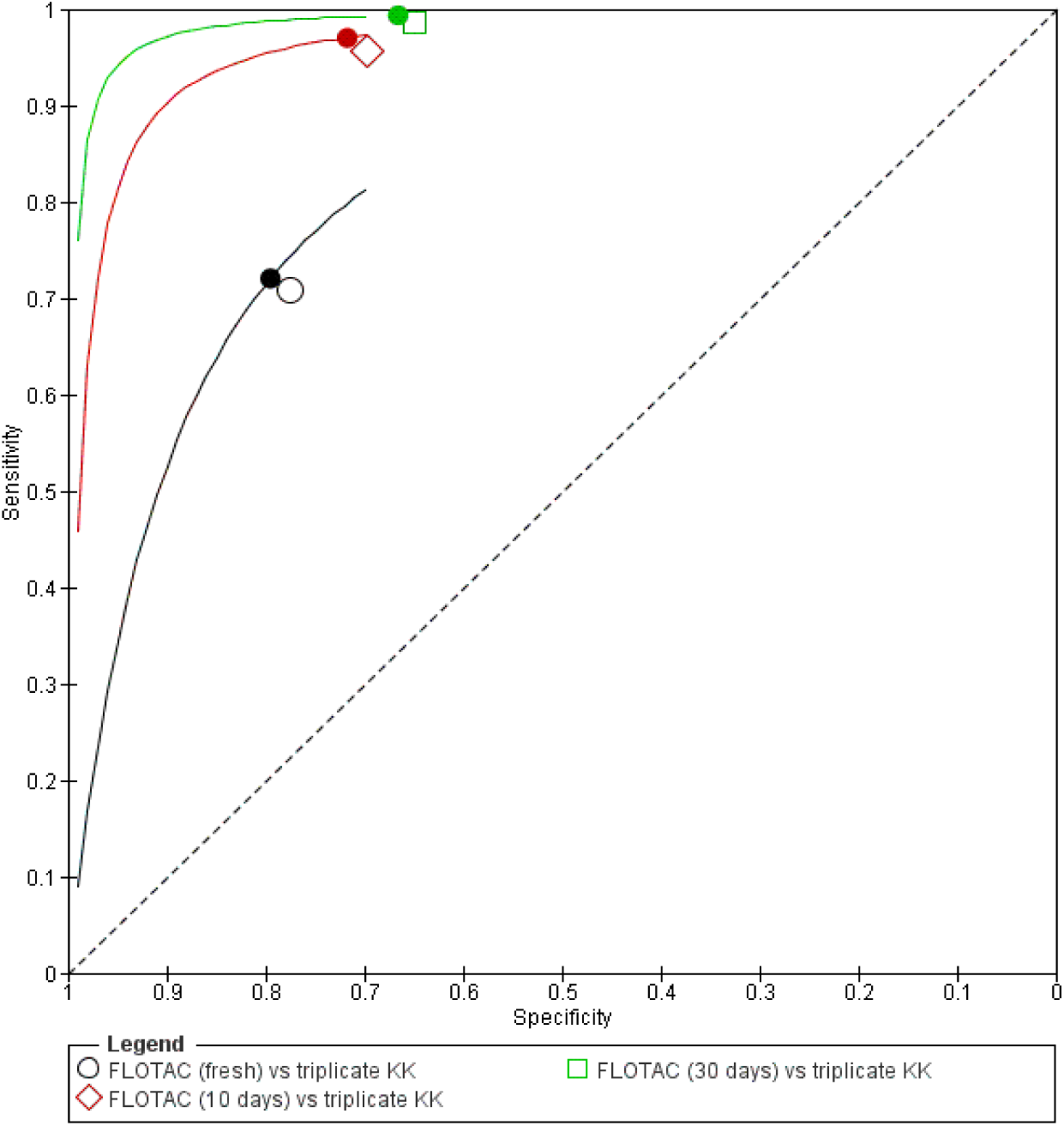
Summary ROC Plot of FLOTAC (fresh), FLOTAC (10 days) or FLOTAC (30 days) vs triplicate KK

**Figure 22.**
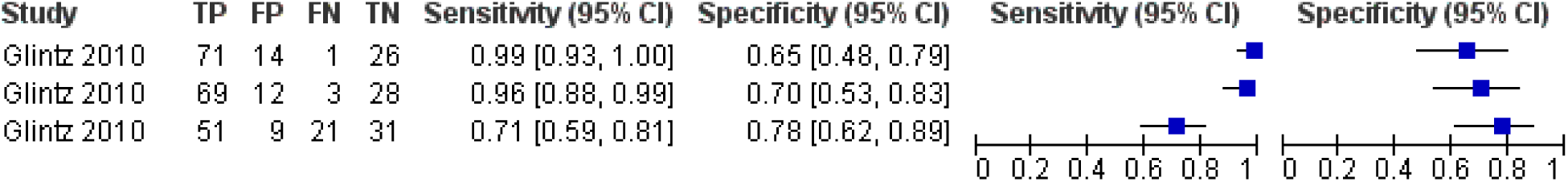
Forest plot of FLOTAC (fresh), FLOTAC (10 days) or FLOTAC (30 days) vs triplicate KK

**Figure 23.**
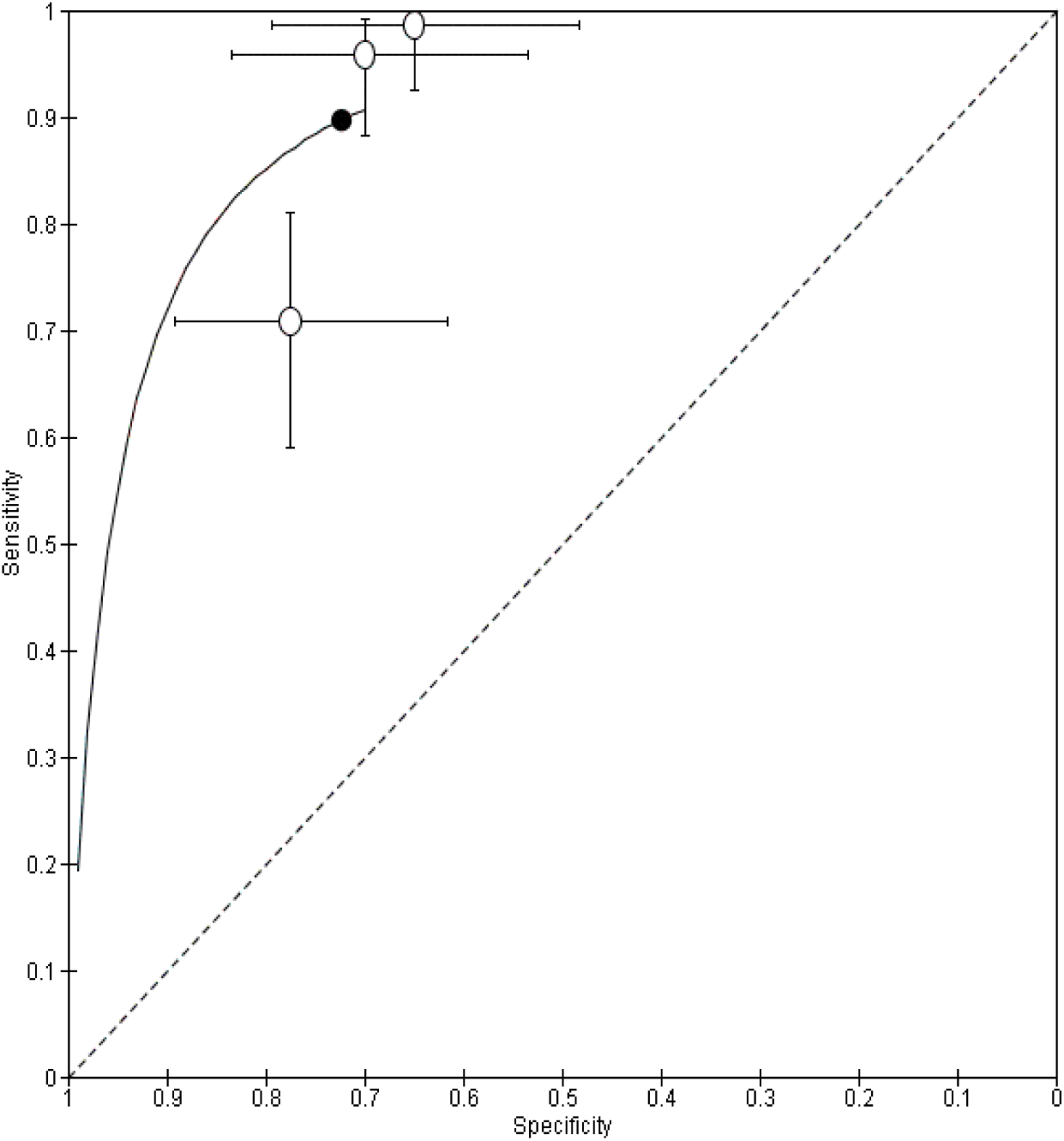
Summary ROC Plot of FLOTAC (fresh), FLOTAC (10 days) and FLOTAC (30 days) vs triplicate KK

**Figure 24.**
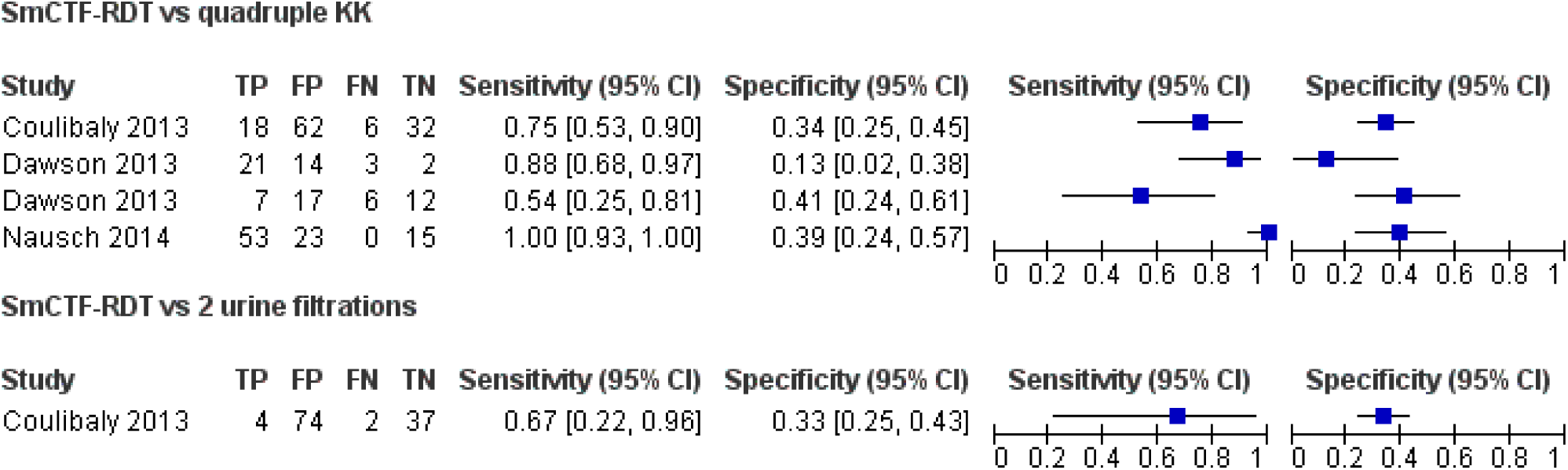
Forest plot of SmCTF-RDT vs quadruple KK or Urine Microscopy

**Figure 25.**
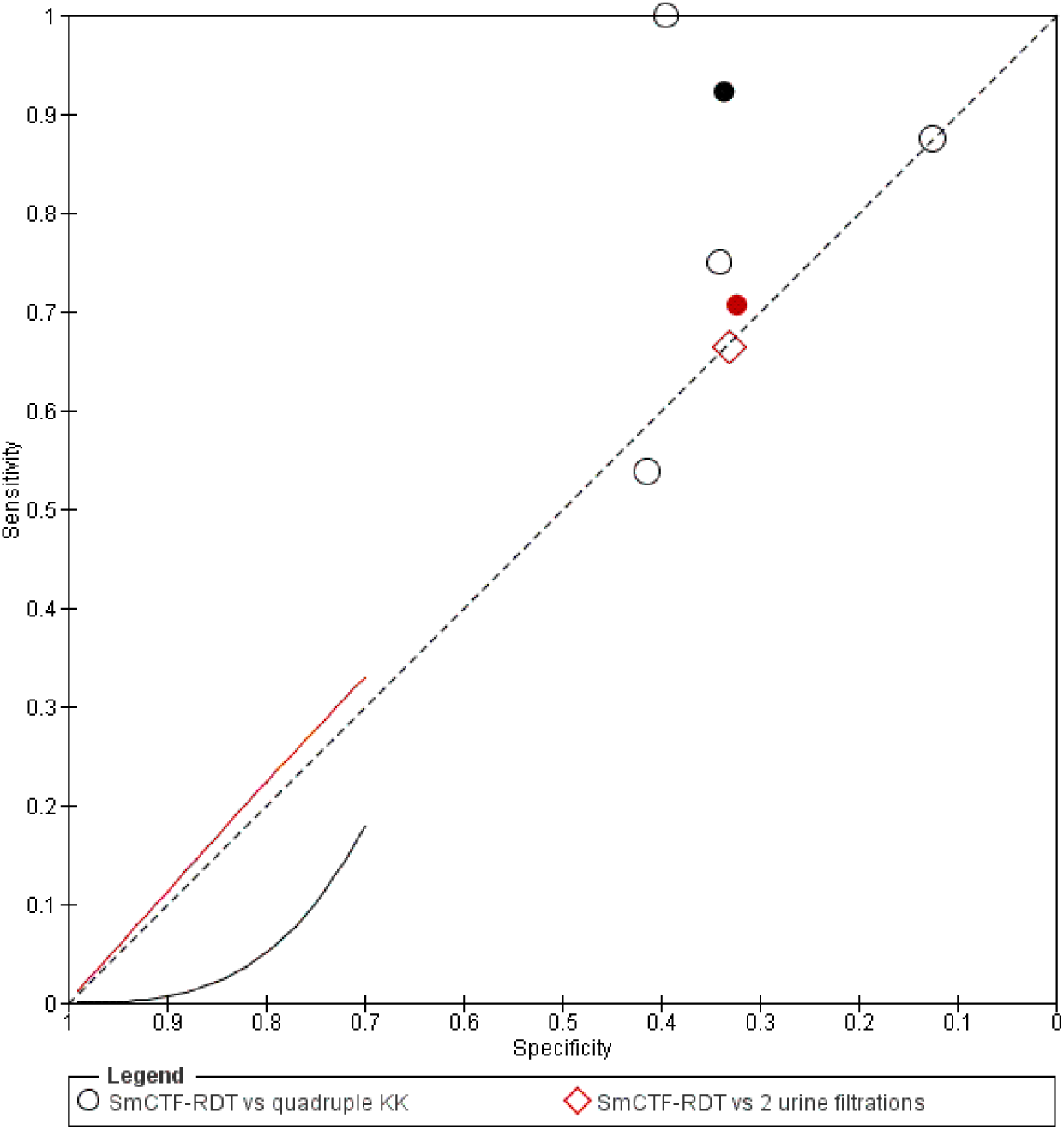
Summary ROC Plot of SmCTF-RDT vs quadruple KK or Urine Microscopy

**Figure 26.**
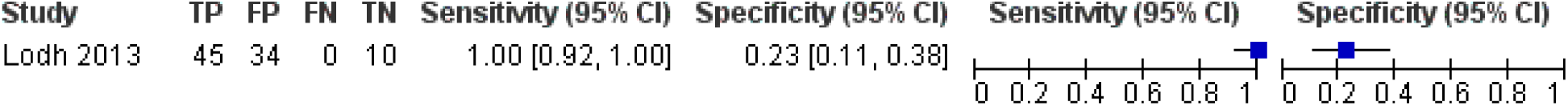
Forest plot of Sm DNA PCR vs duplicate KK

**Figure 27.**
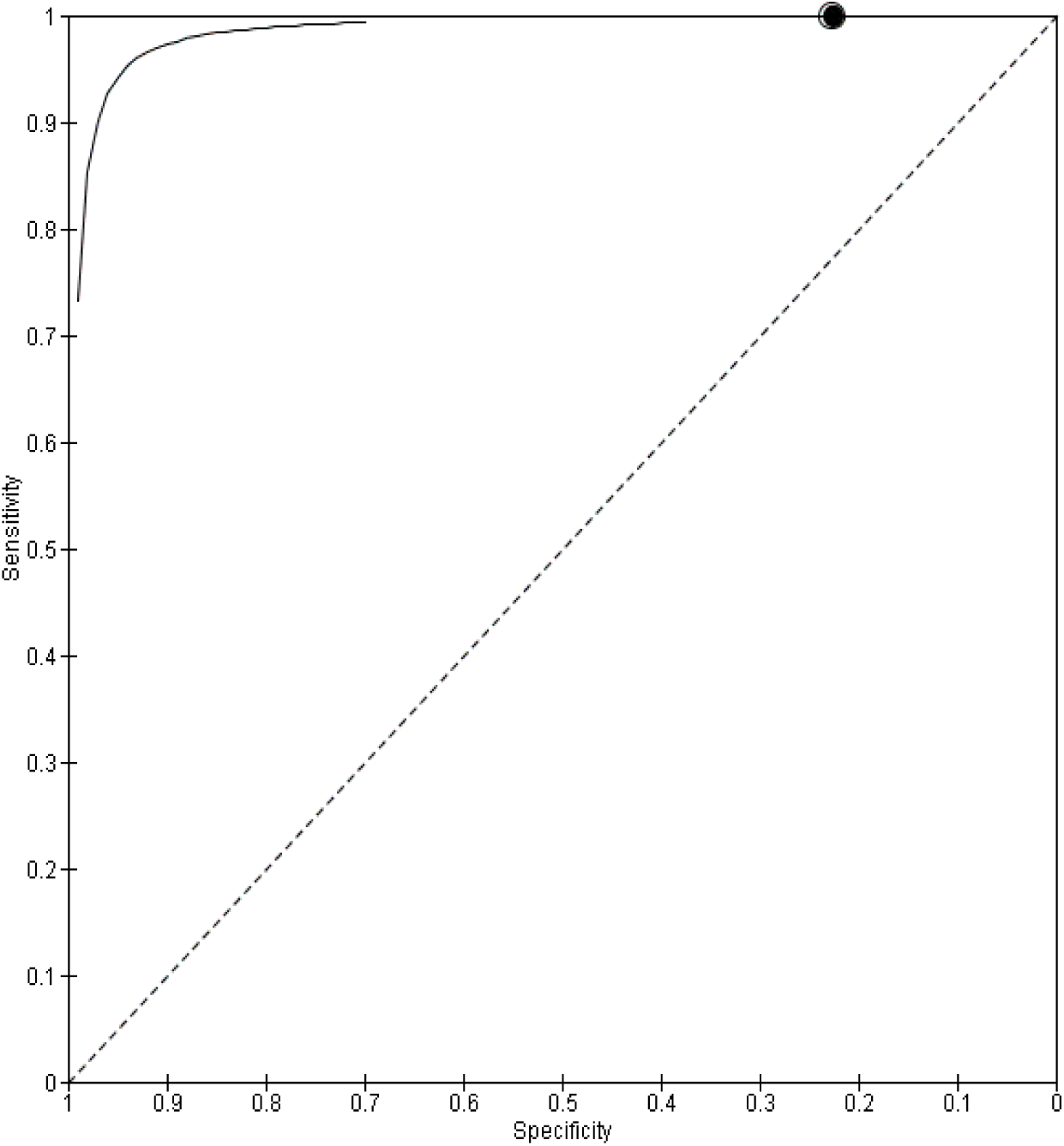
Summary ROC Plot of Sm DNA PCR vs duplicate KK

**Figure 28.**
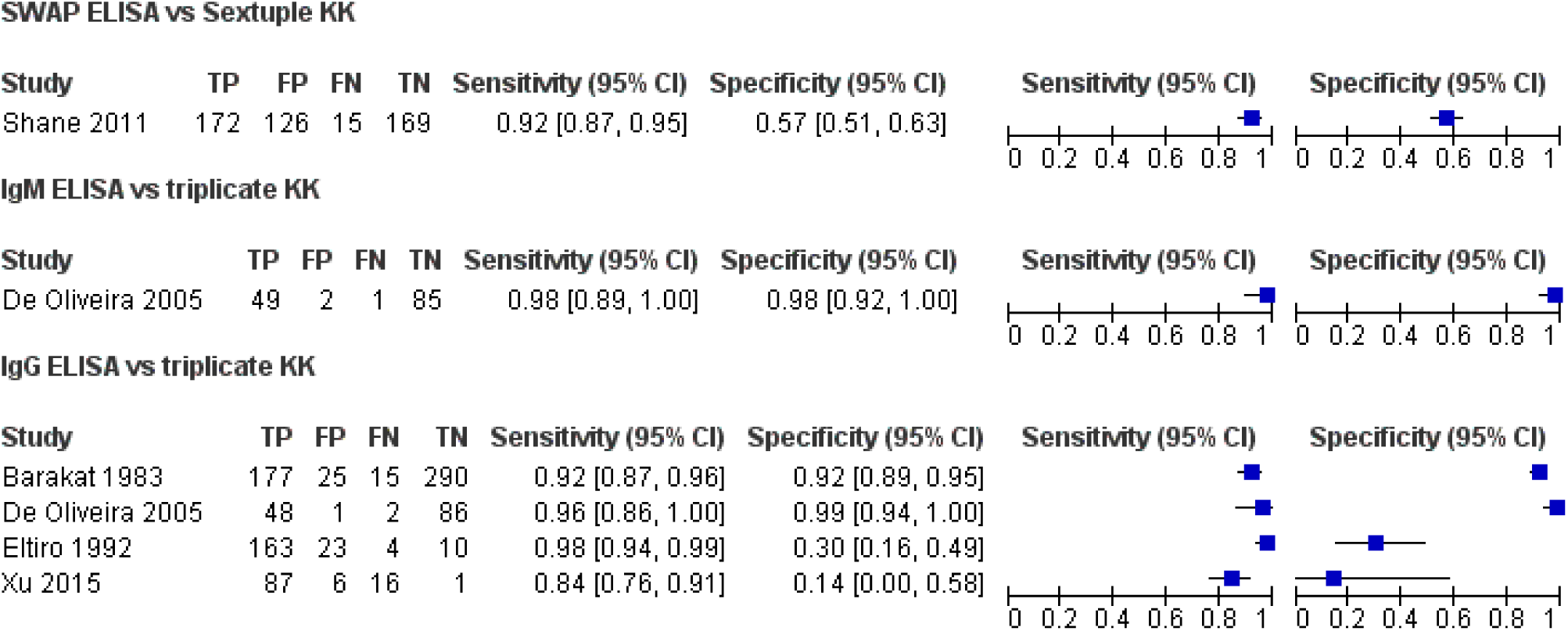
Forest plot of SWAP ELISA vs Sextuple KK, IgM ELISA or IgG ELISA vs triplicate KK

**Figure 29.**
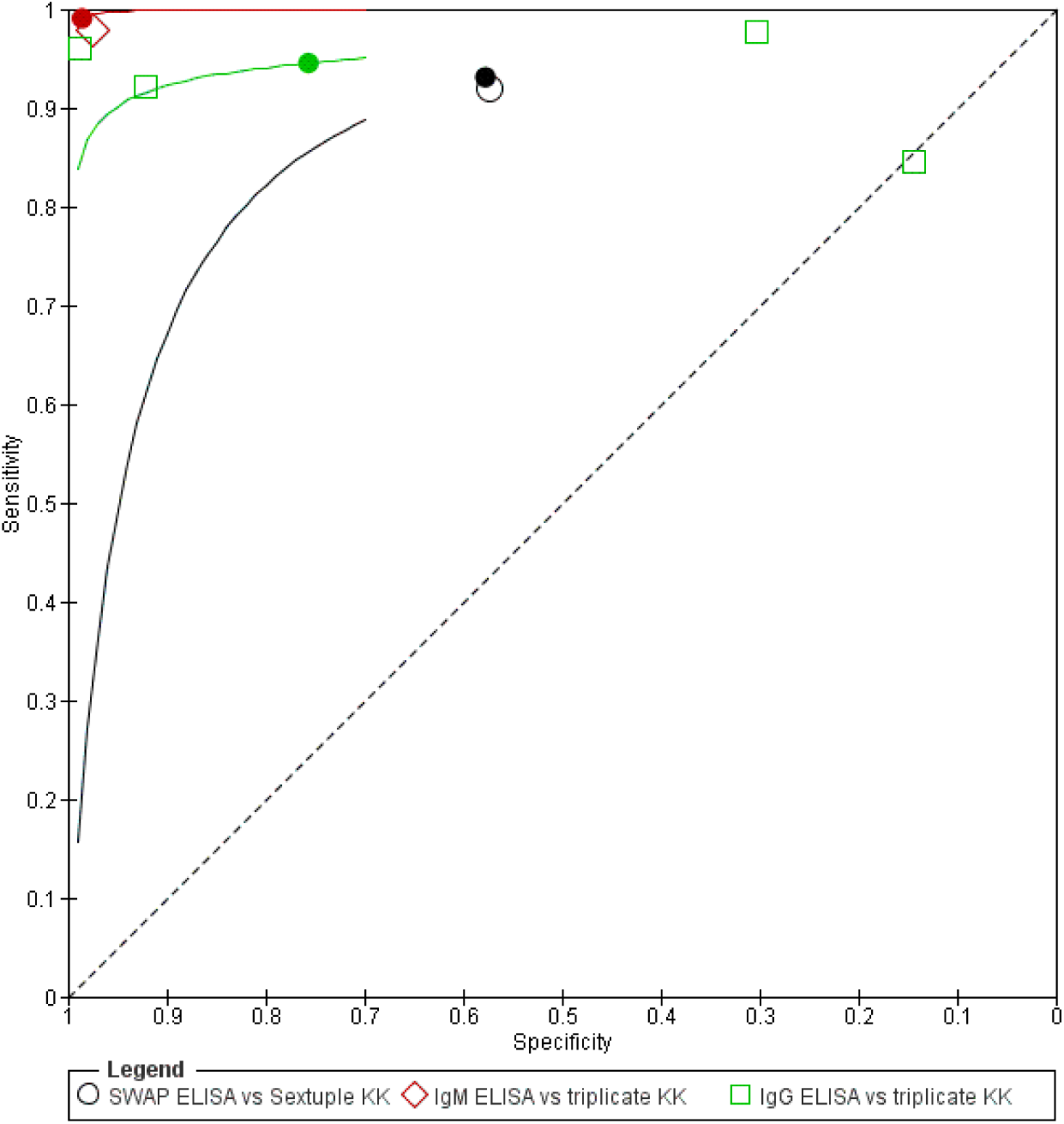
Summary ROC Plot of SWAP ELISA vs Sextuple KK, IgM ELISA or IgG ELISA vs triplicate KK

**Figure 30.**
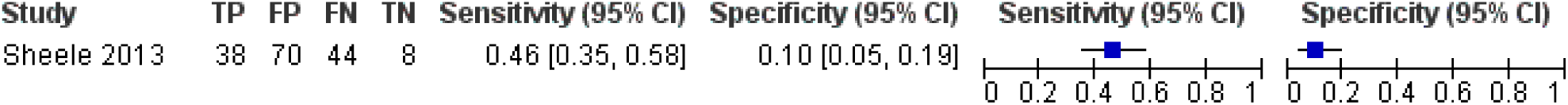
Forest plot of Anti IGg RDT-Sh vs Urine Microscopy

**Figure 31.**
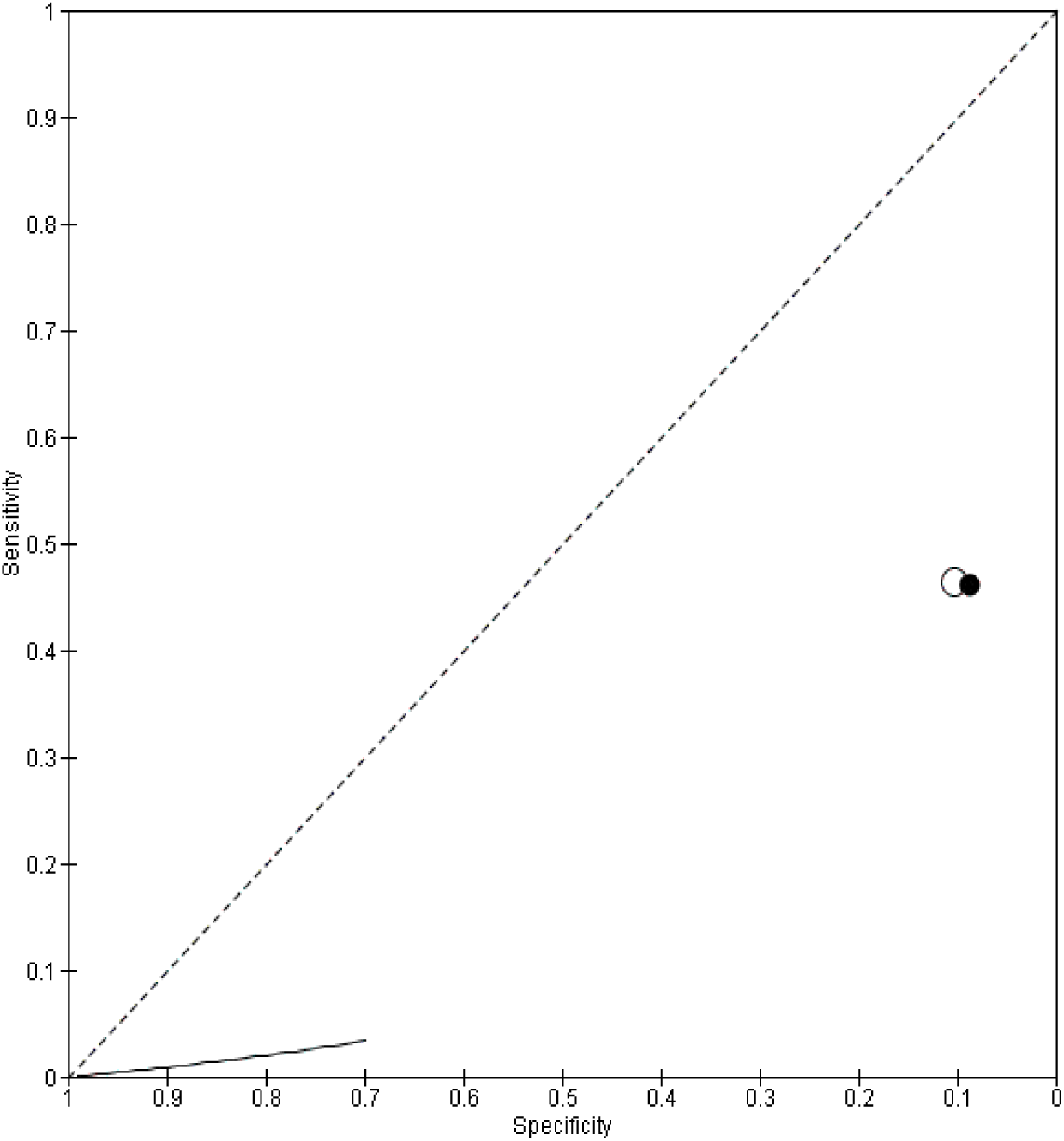
Summary ROC Plot of Anti IGg RDT-Sh vs Urine Microscopy

**Figure 32.**
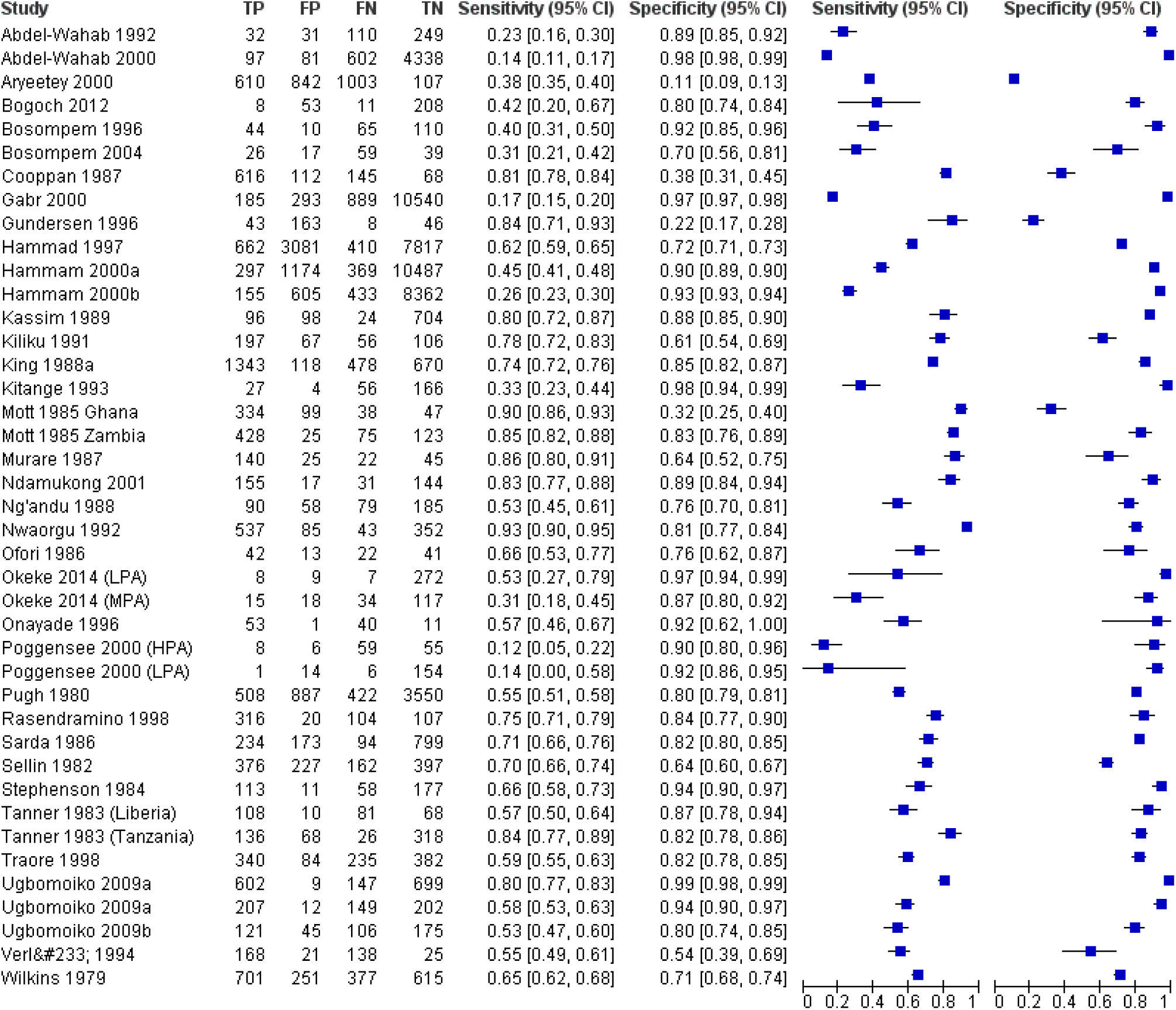
Forest plot of Proteinuria (R strip) vs Urine Microscopy

**Figure 33.**
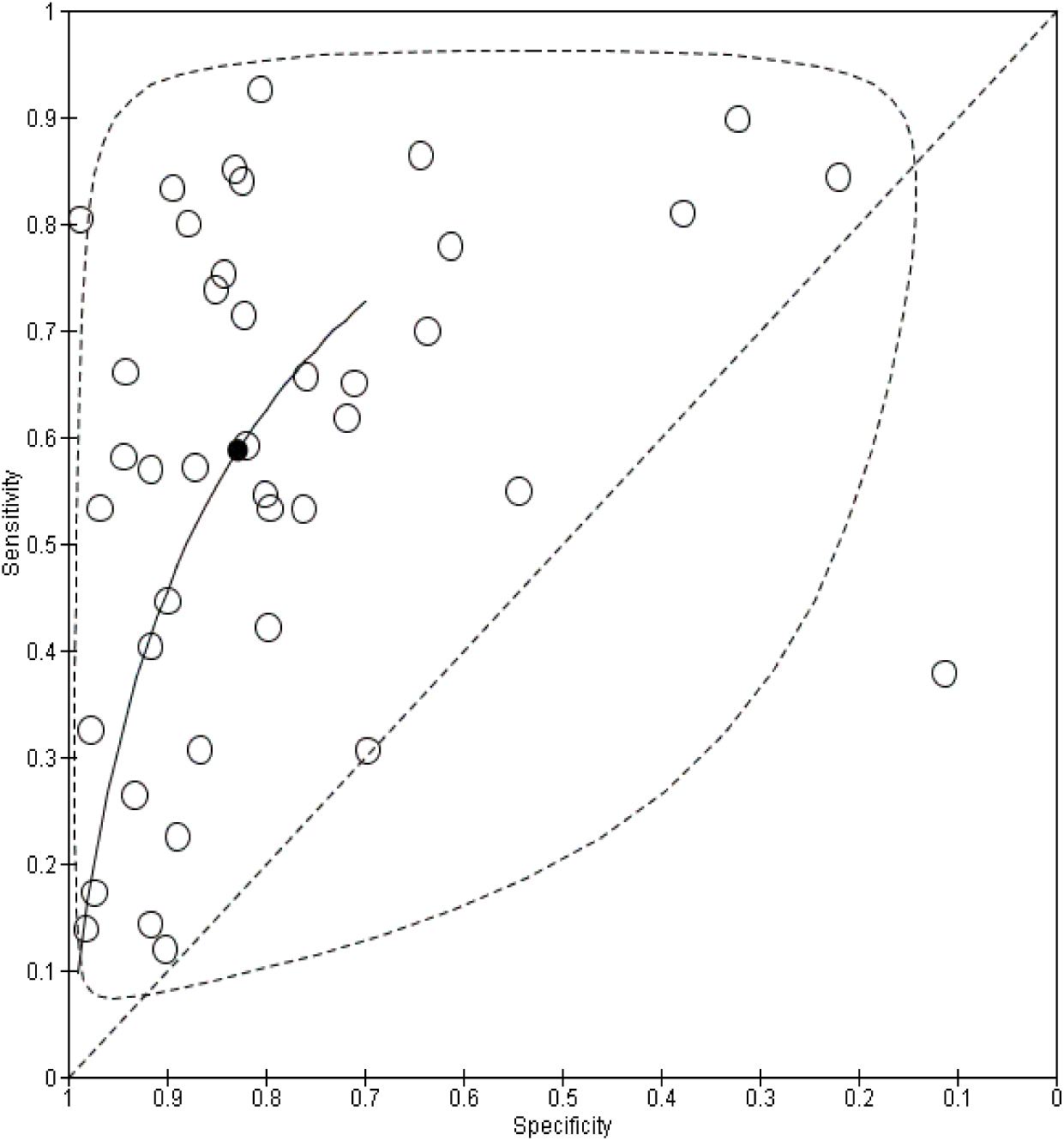
Summary ROC Plot of Proteinuria (R strip) vs Urine Microscopy

**Figure 34.** Forest plot of Haematuria (R strip) vs Urine Microscop

**Figure 35.**
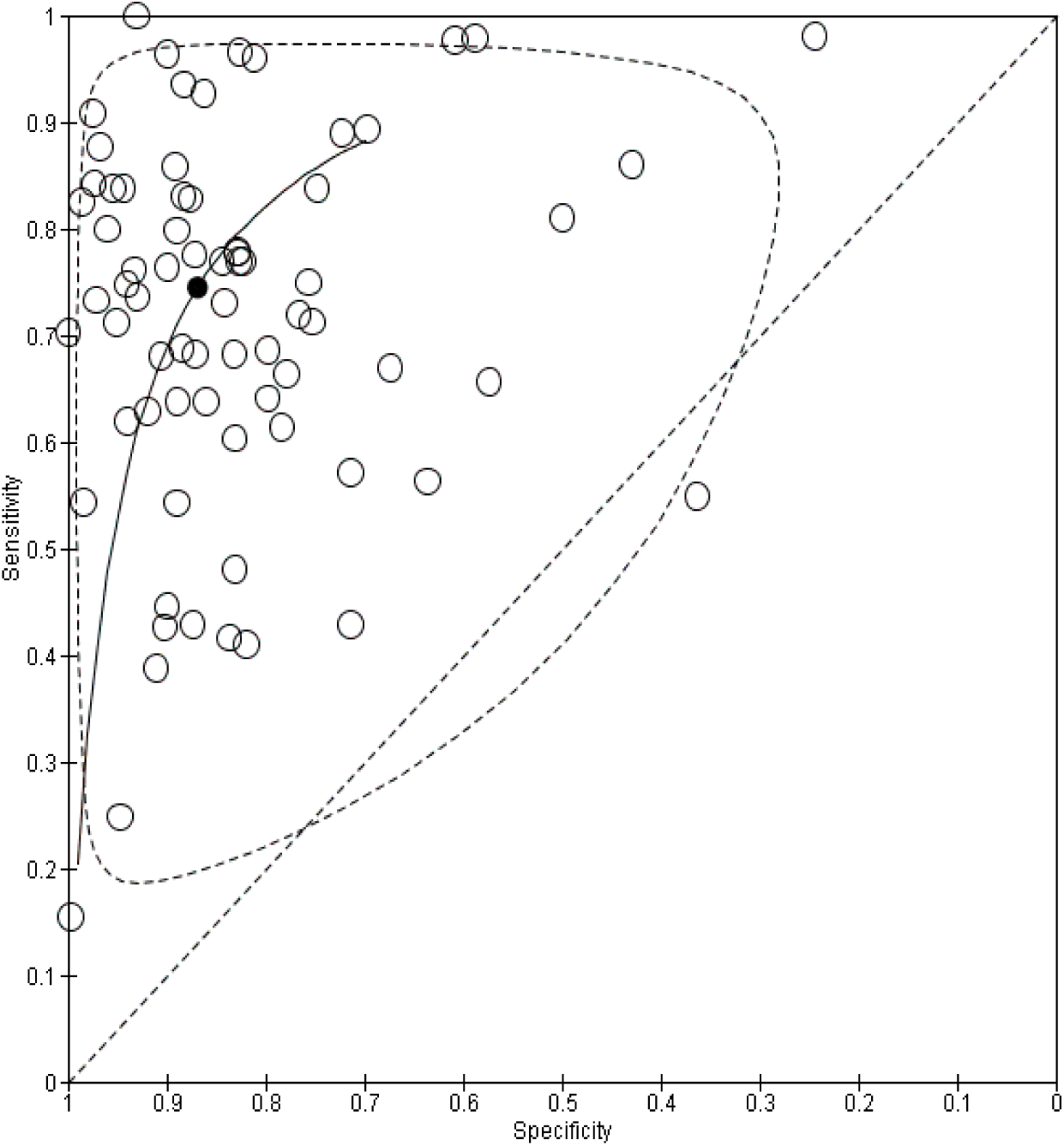
Summary ROC Plot of Haematuria (R strip) vs Urine Microscopy

**Figure 36.**
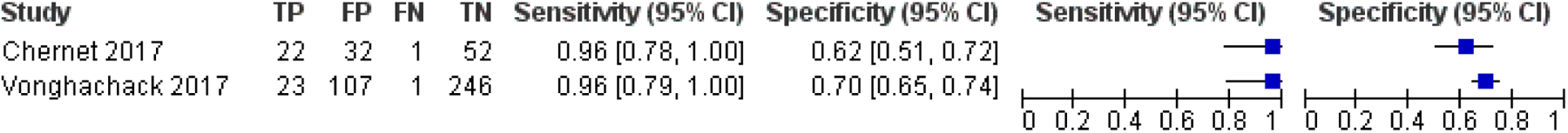
Forest plot of AWE-SEA ELISA vs quadruple KK

**Figure 37.**
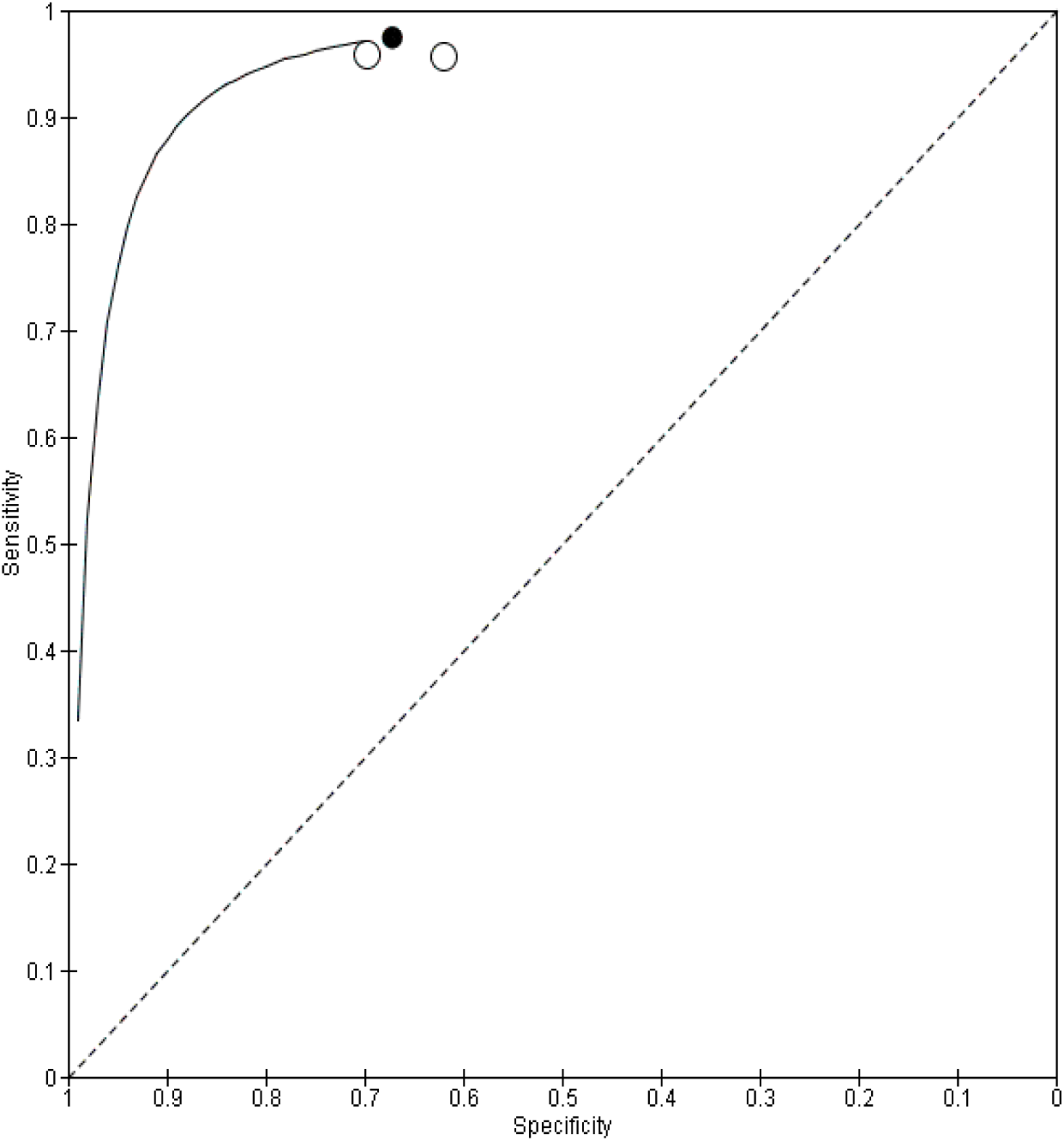
Summary ROC Plot of AWE-SEA ELISA vs quadruple KK

**Figure 38.**
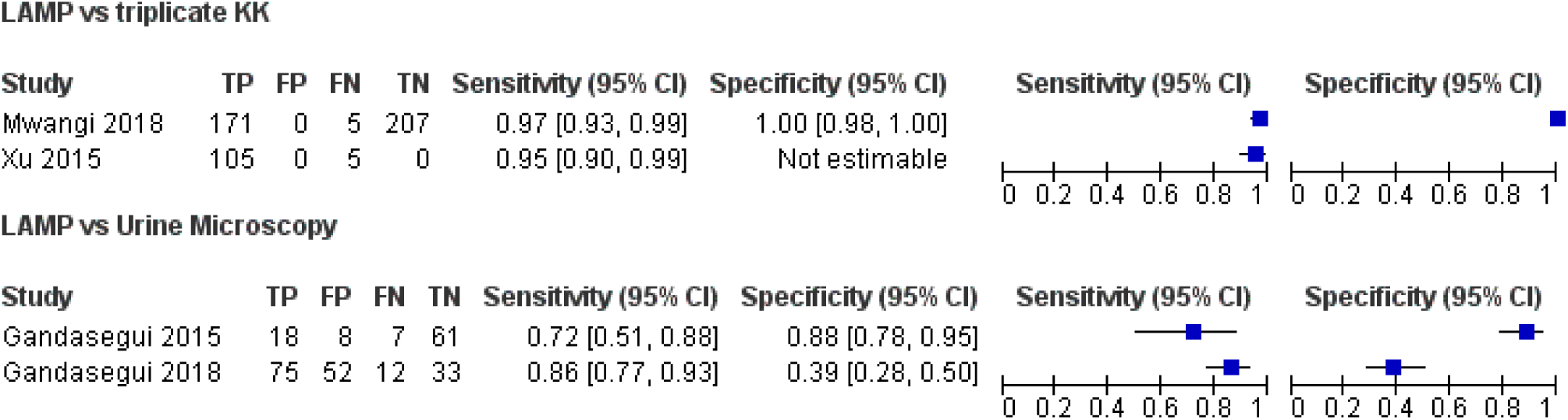
Forest plot of LAMP vs triplicate KK or Urine Microscopy

**Figure 39.**
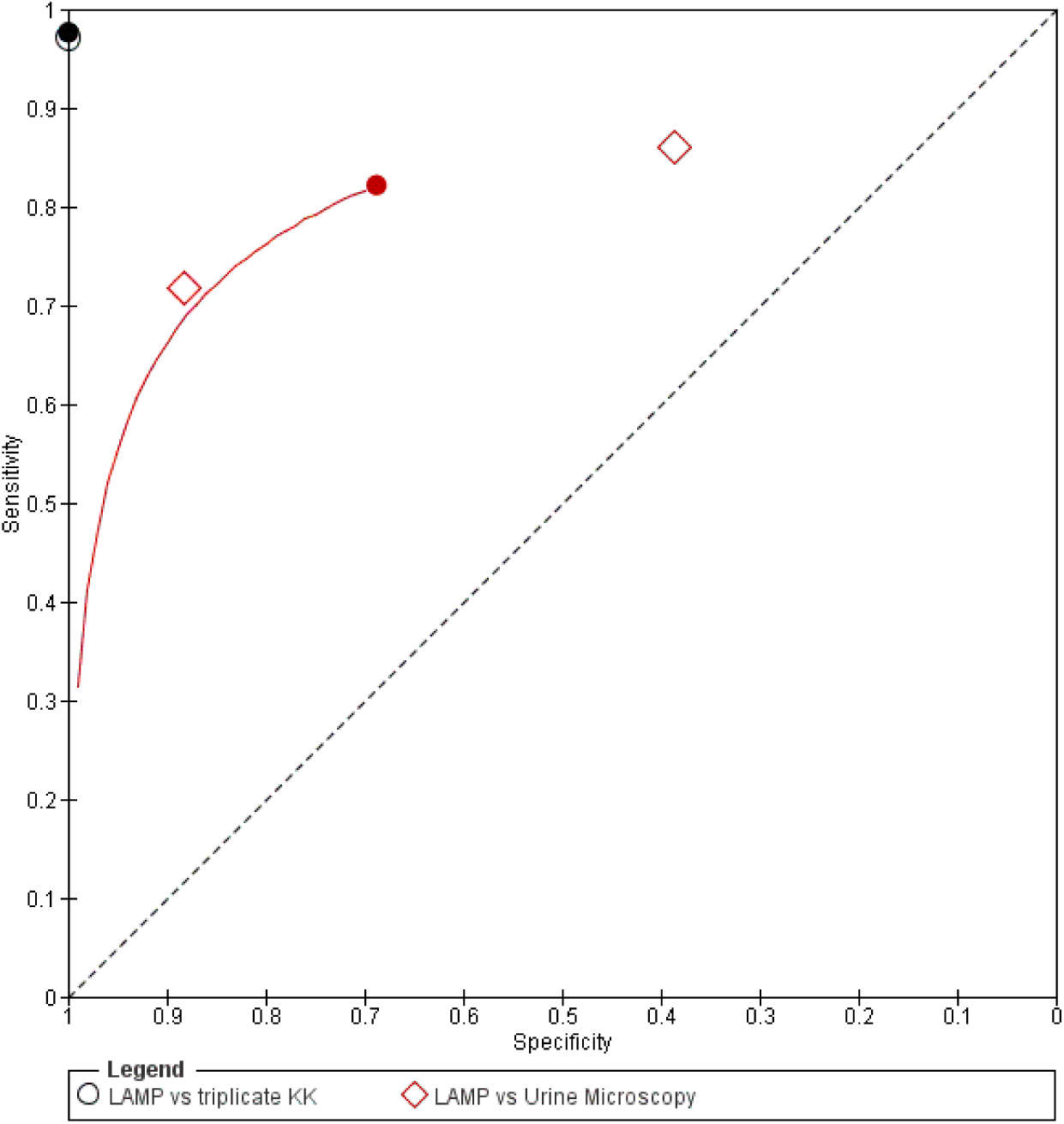
Summary ROC Plot of LAMP vs triplicate KK or Urine Microscopy

**Figure 40.**
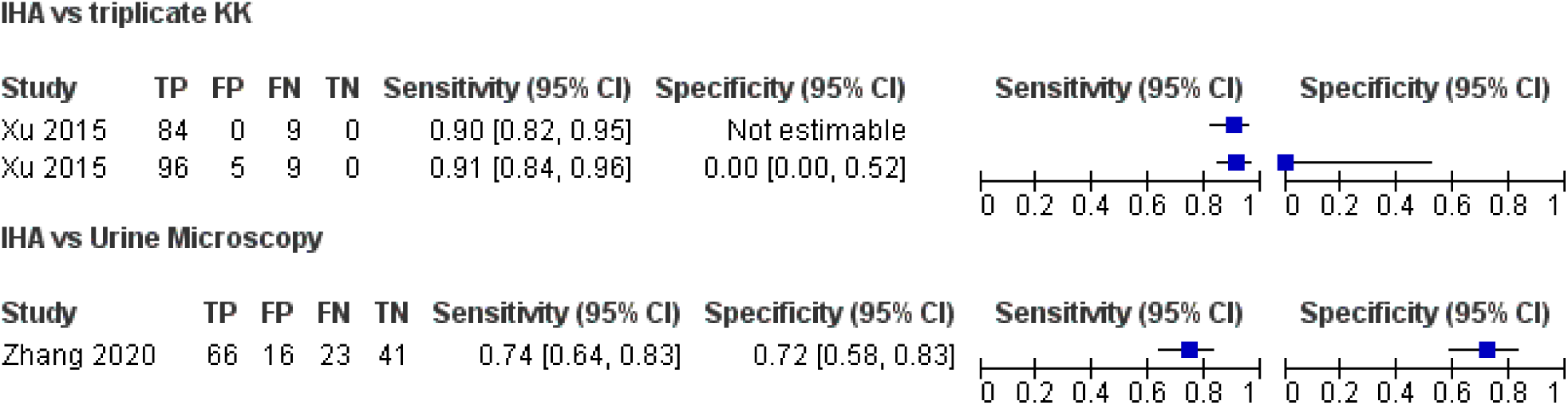
Forest plot of IHA vs triplicate KK or Urine Microscopy.

**Figure 41.**
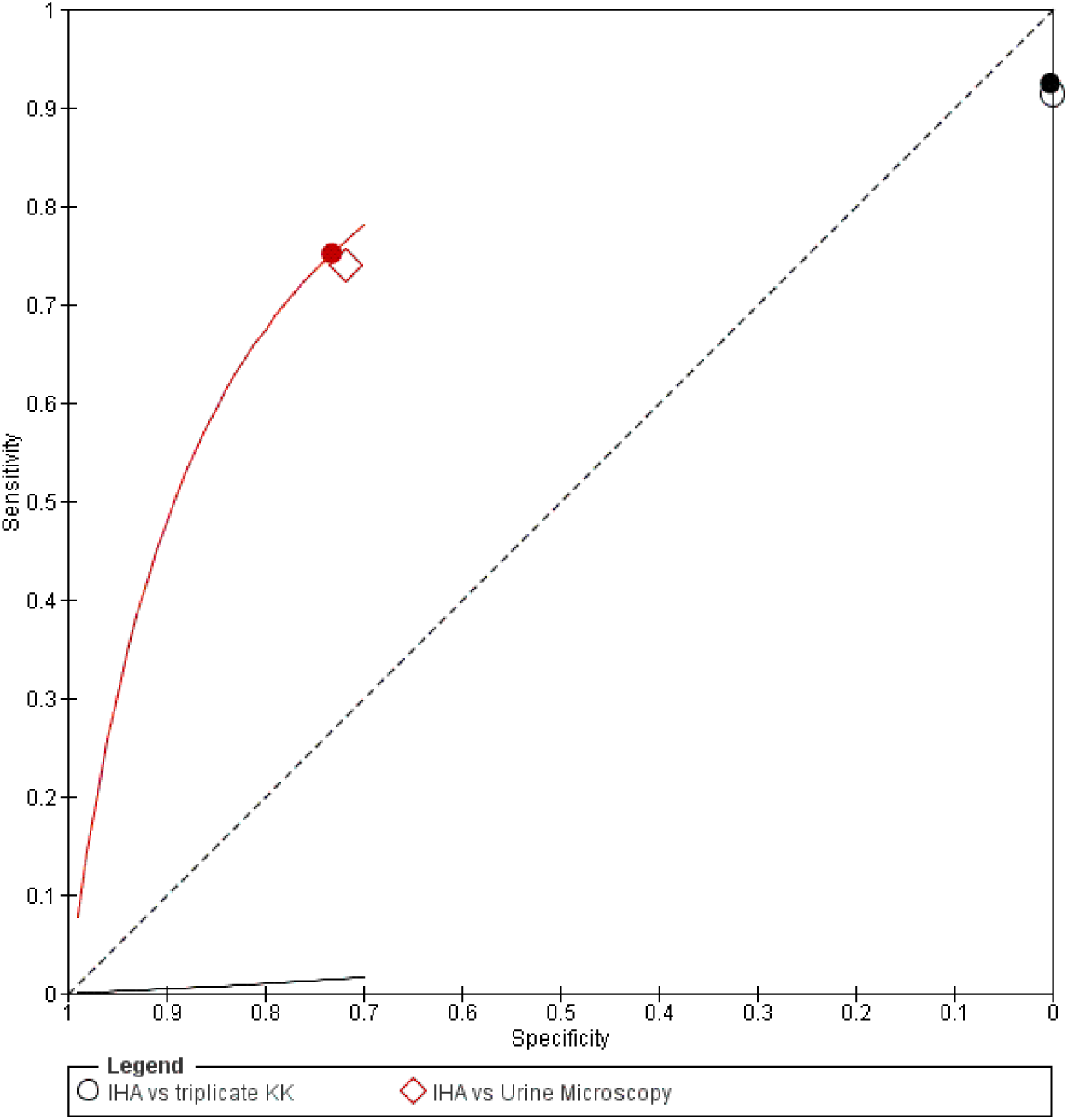
Summary ROC Plot of IHA vs triplicate KK or Urine Microscopy

**Figure 42.**
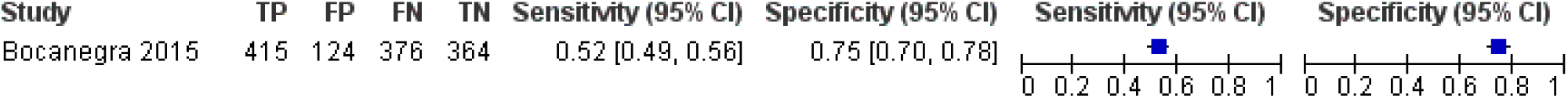
Forest plot of Colorimetric test vs Urine Microscopy

**Figure 43.**
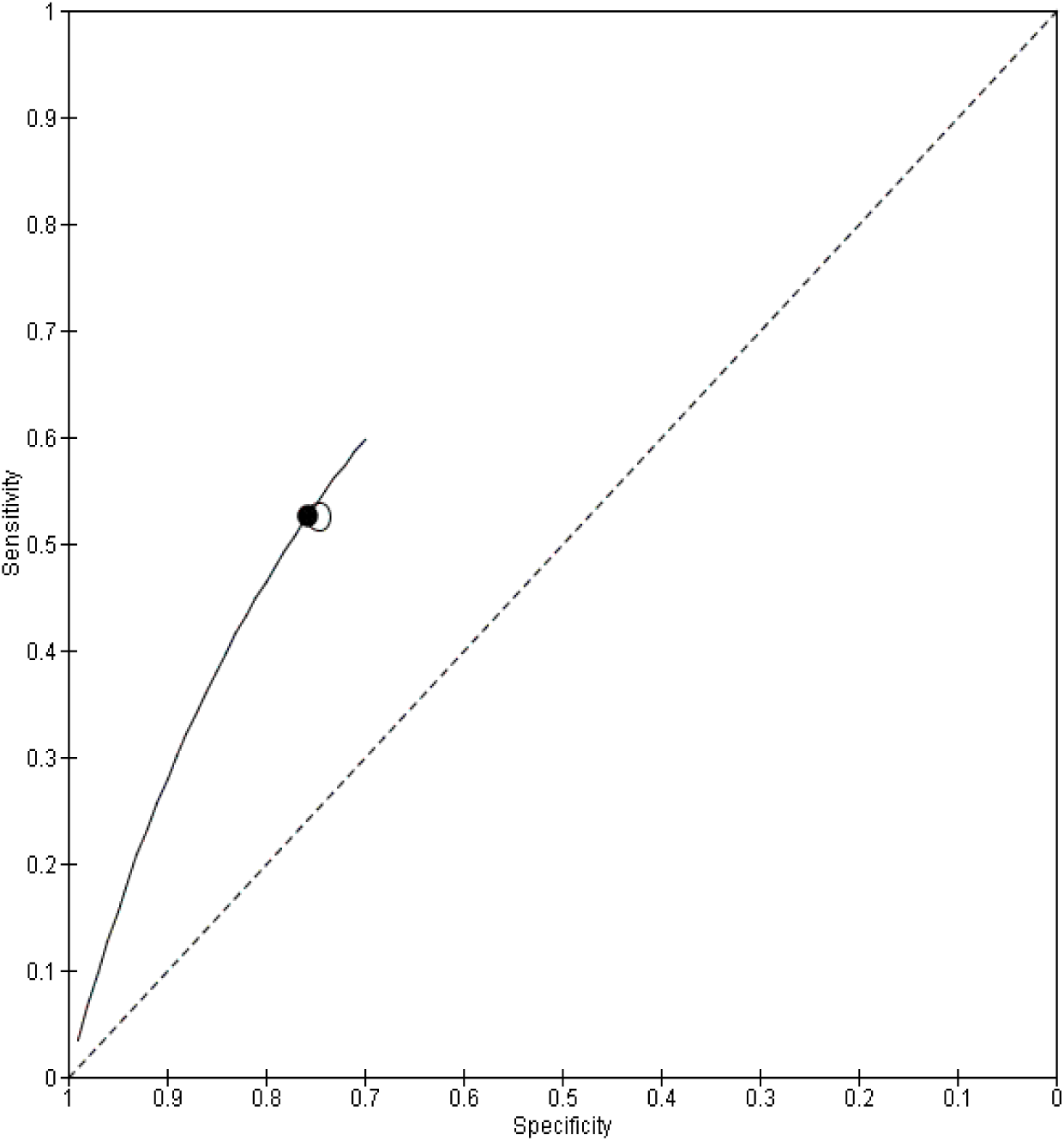
Summary ROC Plot of Colorimetric test vs Urine Microscopy

**Figure 44.**
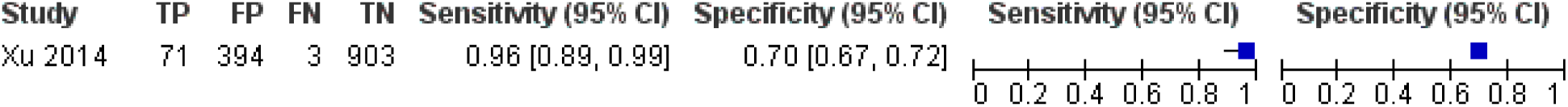
Forest plot of rSP13 ELISA vs 27KK

**Figure 45.**
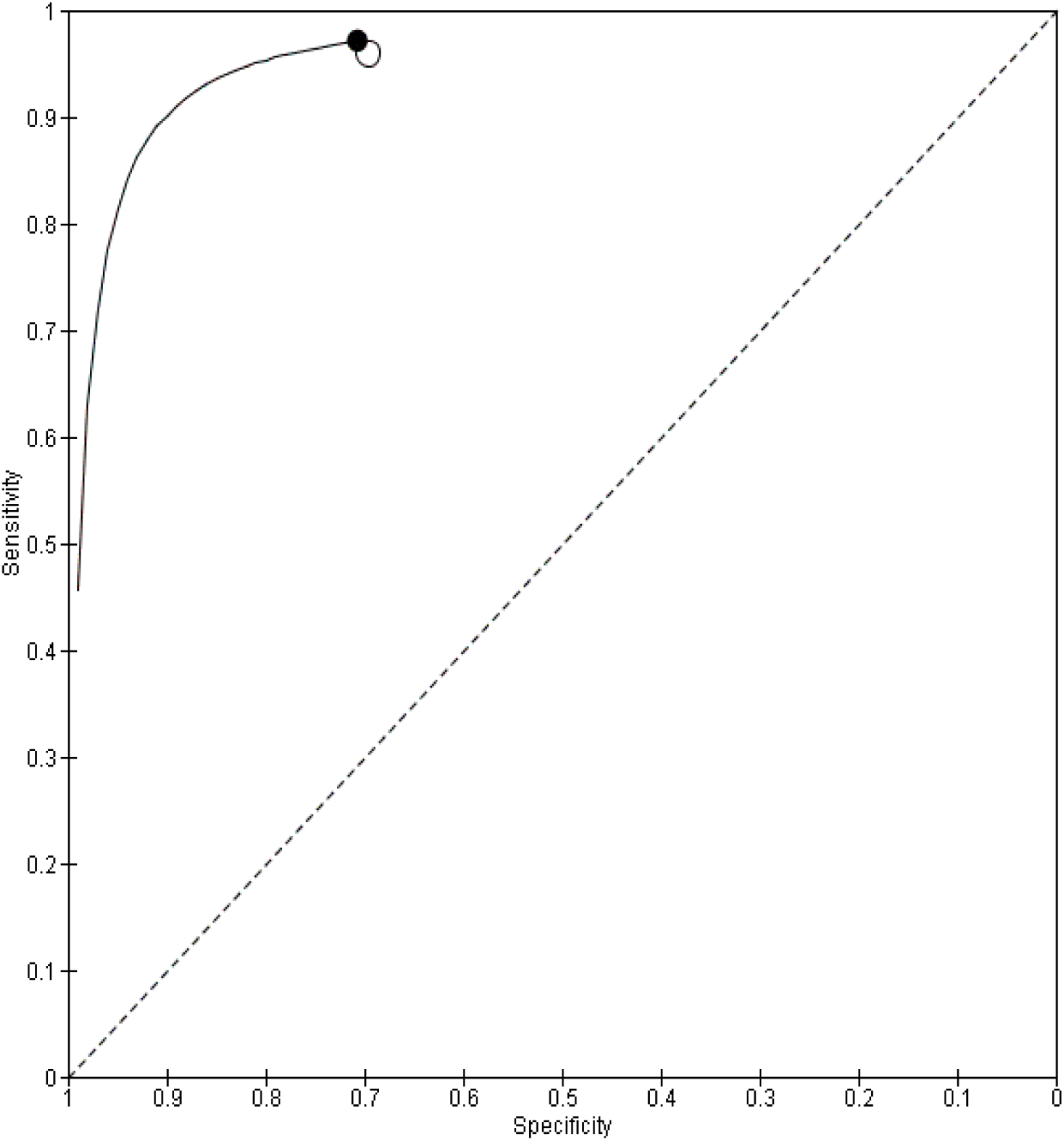
Summary ROC Plot of rSP13 ELISA vs 27KK

**Figure 46.**
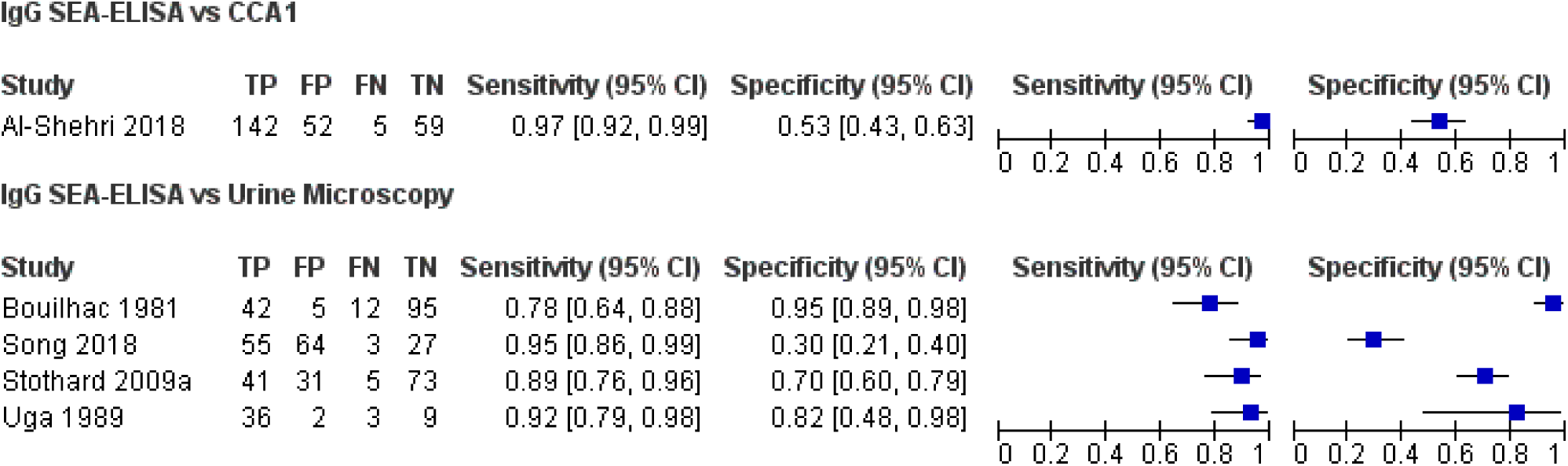
Forest plot of IgG SEA-ELISA vs Urine Microscopy

**Figure 47.**
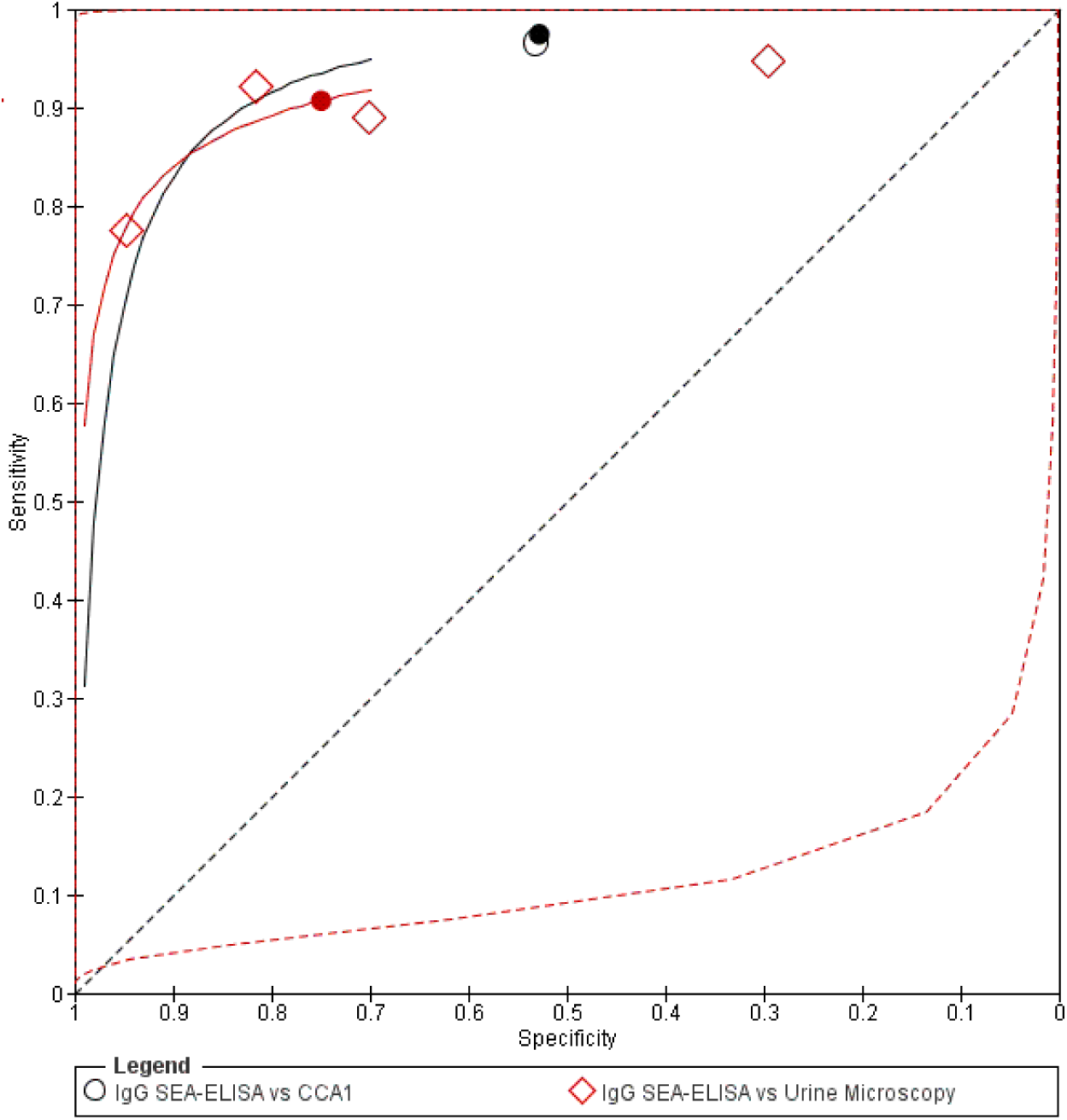
Summary ROC Plot of IgG SEA-ELISA vs Urine Microscopy

**Figure 48.**
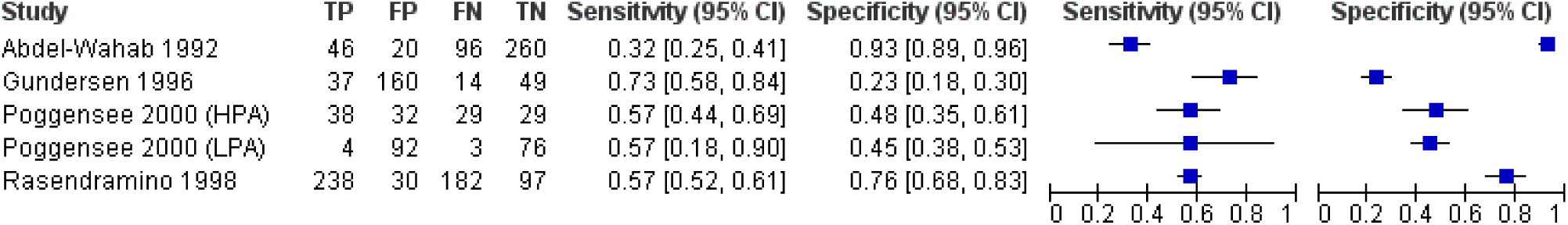
Forest plot of Leukocyturia (reagent strip) vs Urine Microscopy

**Figure 49.**
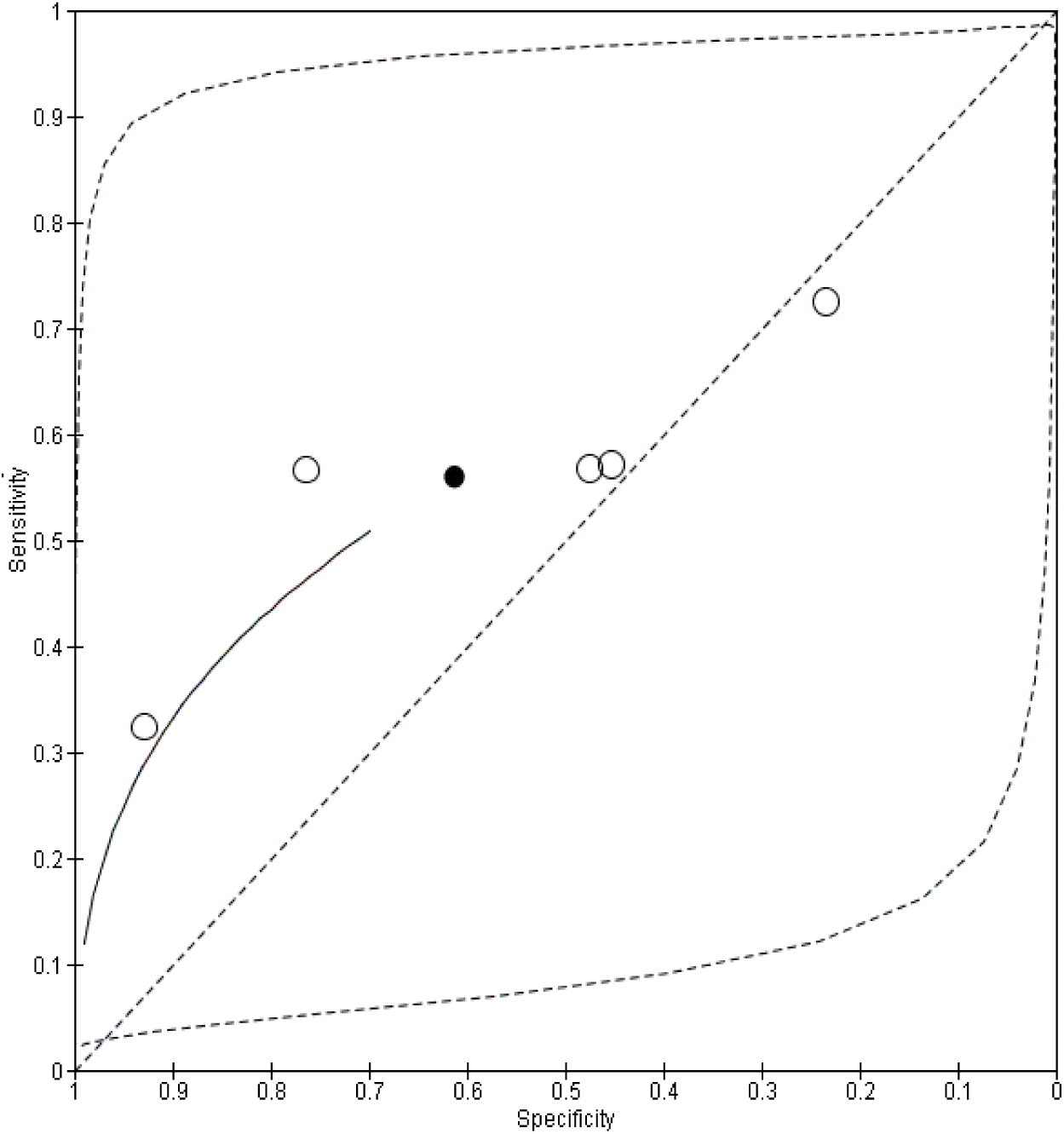
Summary ROC Plot of Leukocyturia (reagent strip) vs Urine Microscopy

**Figure 50.**
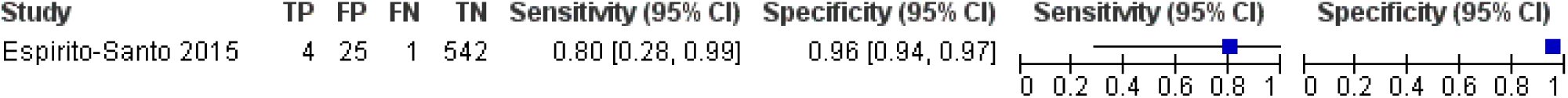
Forest plot of COPT vs duplicate KK

**Figure 51.**
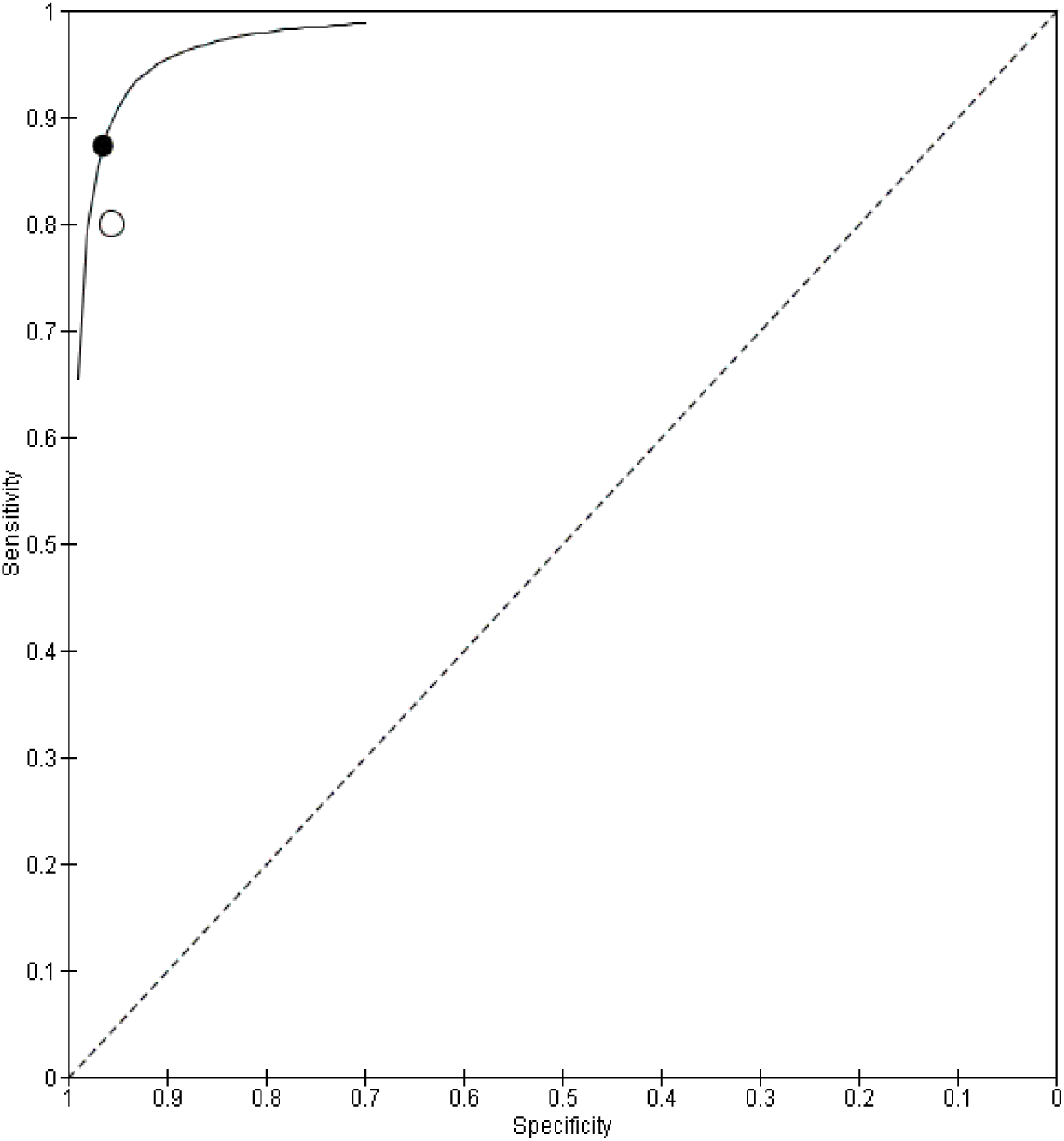
Summary ROC Plot of COPT vs duplicate KK

**Figure 52.**
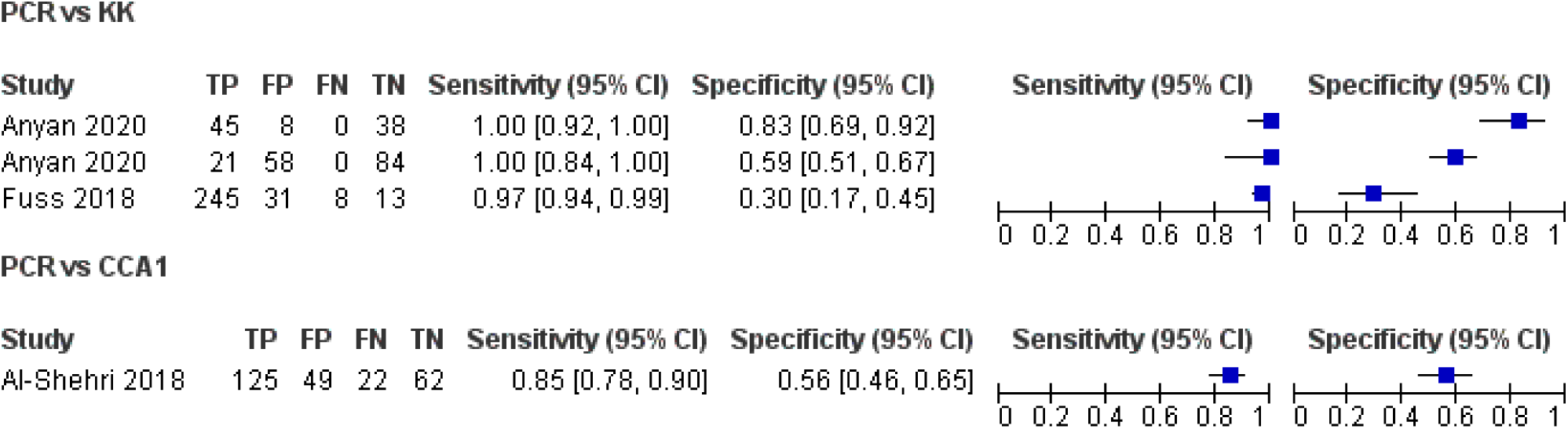
Forest plot of PCR vs KK

**Figure 53.**
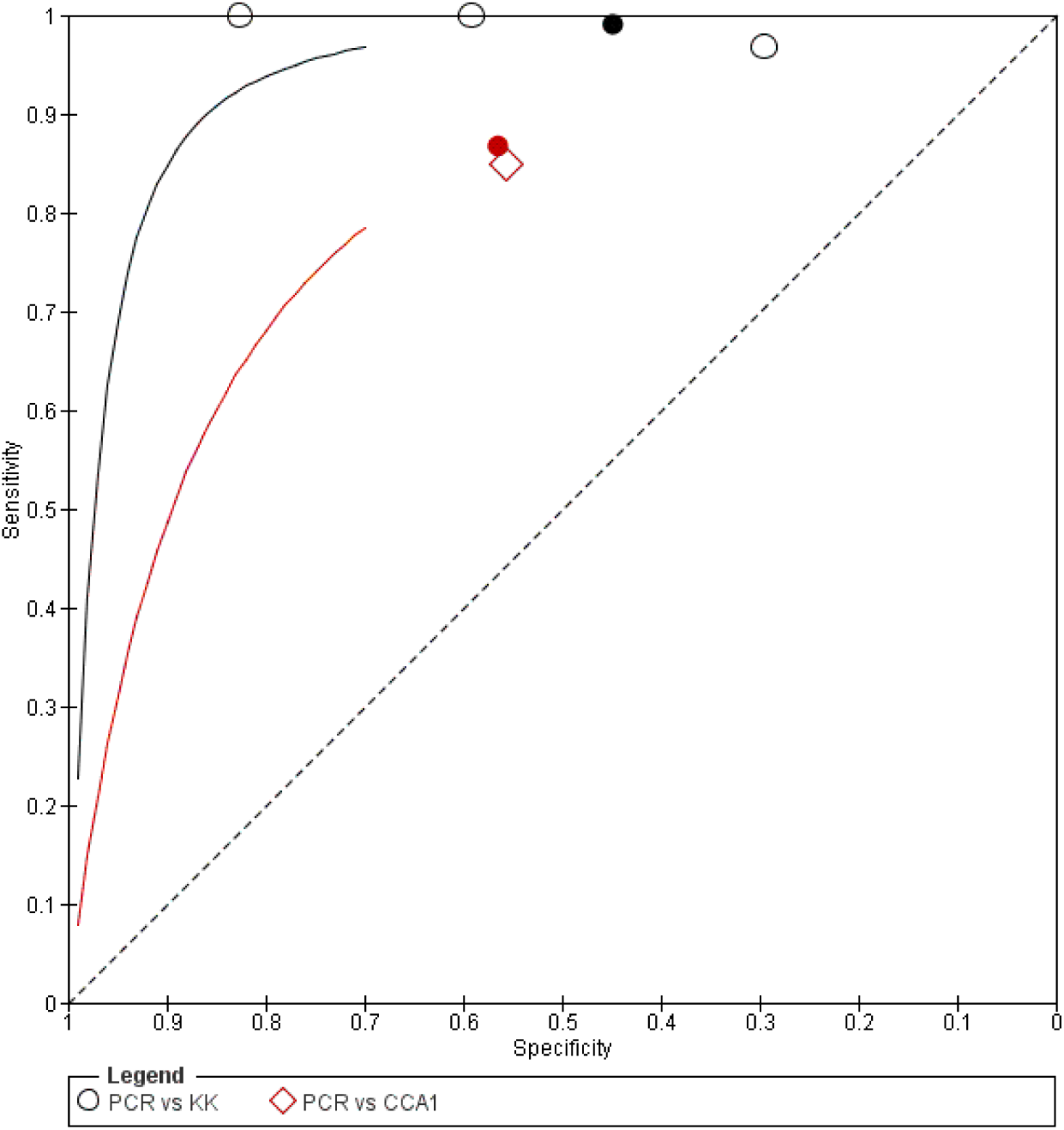
Summary ROC Plot of PCR vs KK

**Figure 54.**
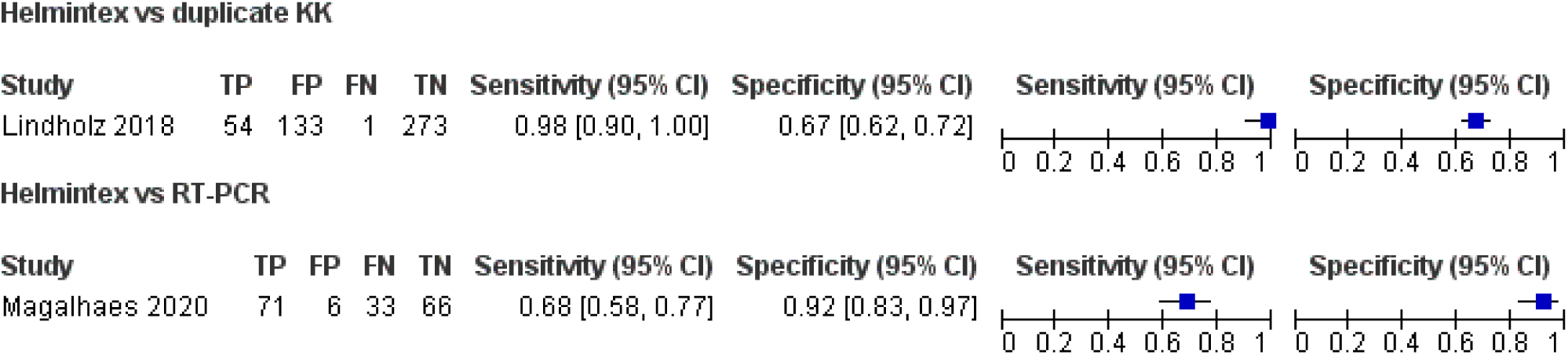
Forest plot of Helmintex vs duplicate KK or RT-PCR

**Figure 55.**
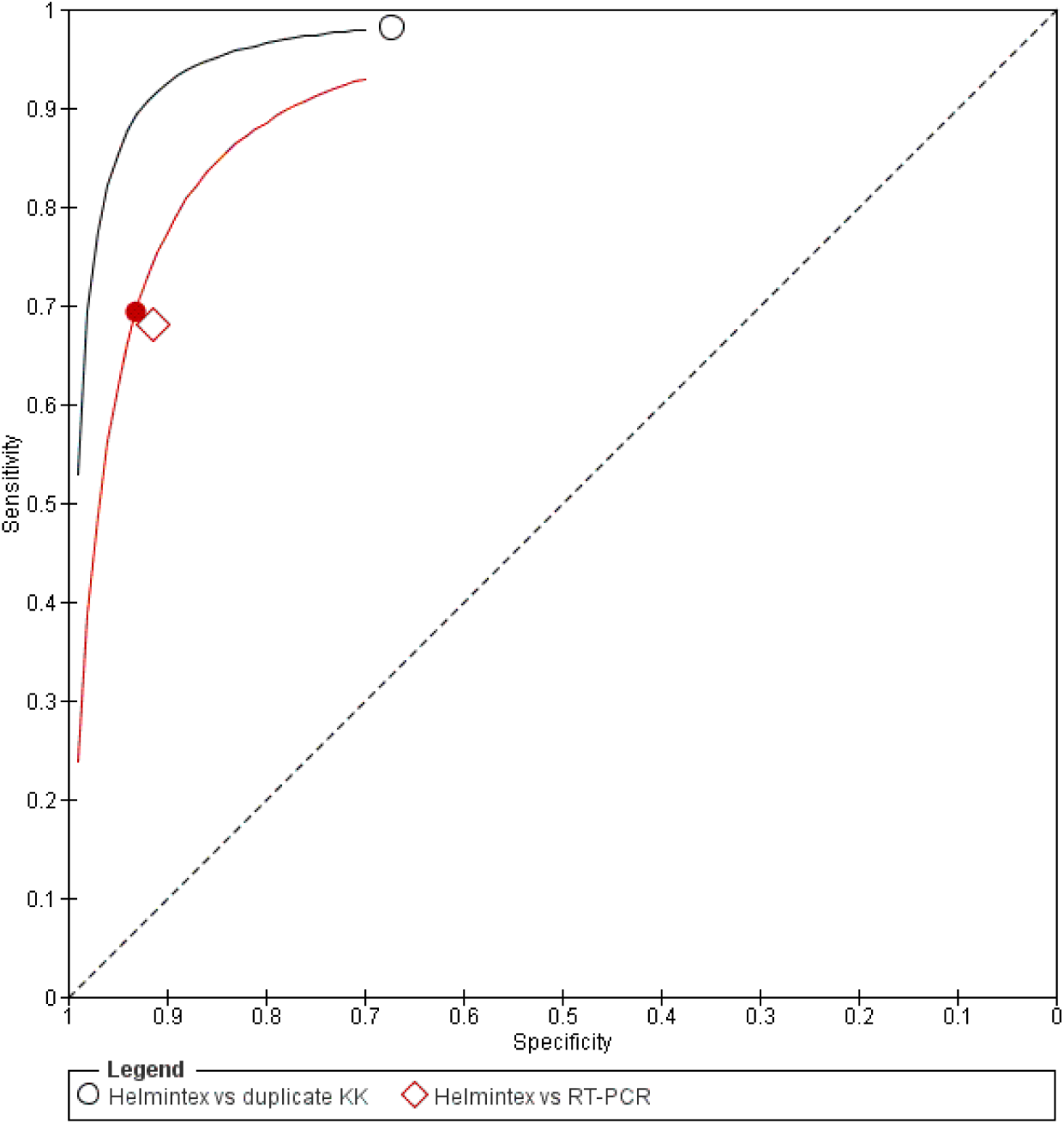
Summary ROC Plot of Helmintex vs duplicate KK or RT-PCR

**Figure 56.**
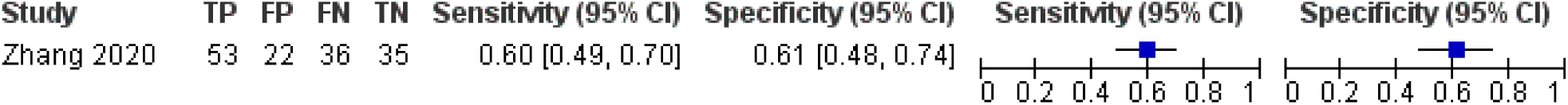
Forest plot of DDIA vs Urine Microscopy

**Figure 57.**
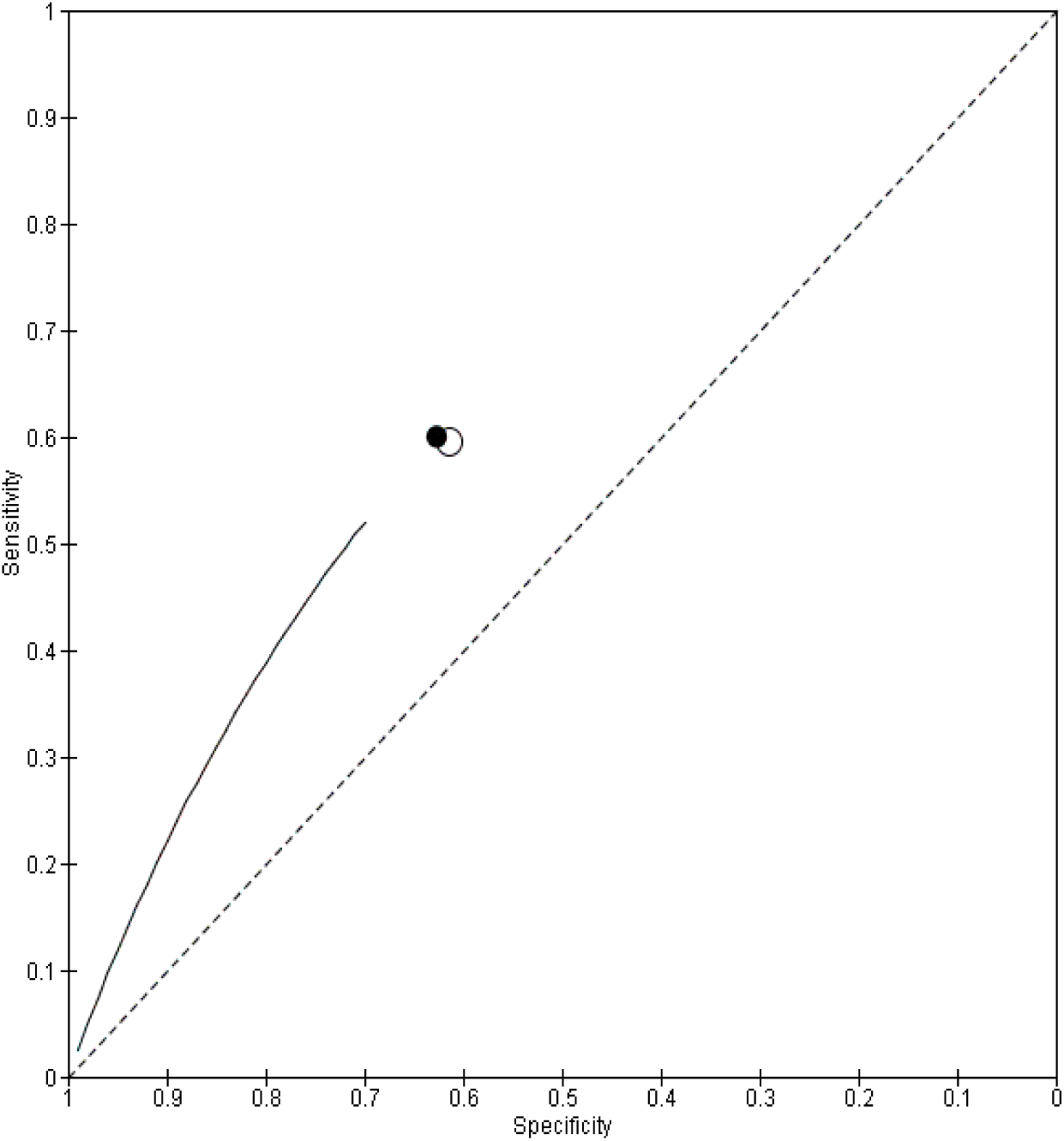
Summary ROC Plot of DDIA vs Urine Microscopy

**Figure 58.**
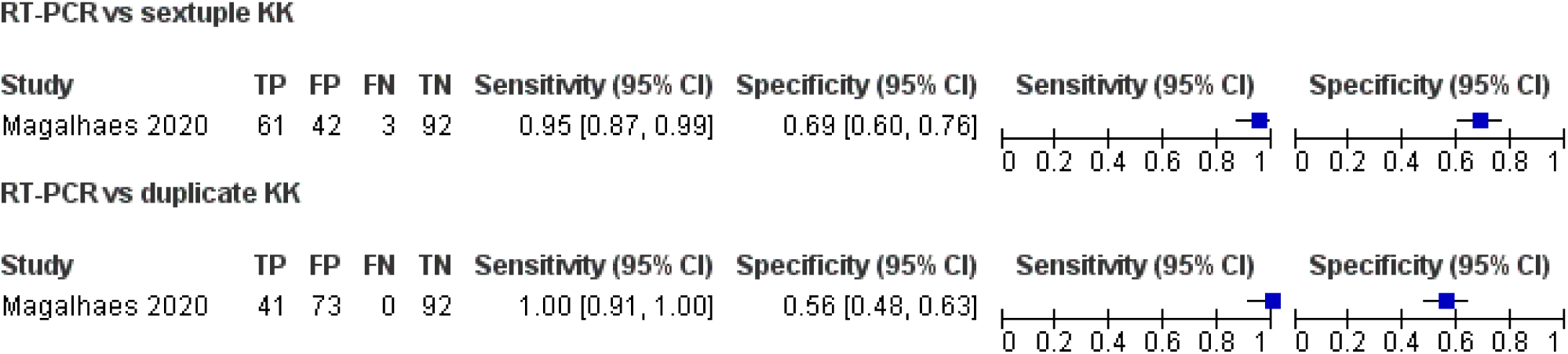
Forest plot of RT-PCR vs duplicate or sextuple KK

**Figure 59.**
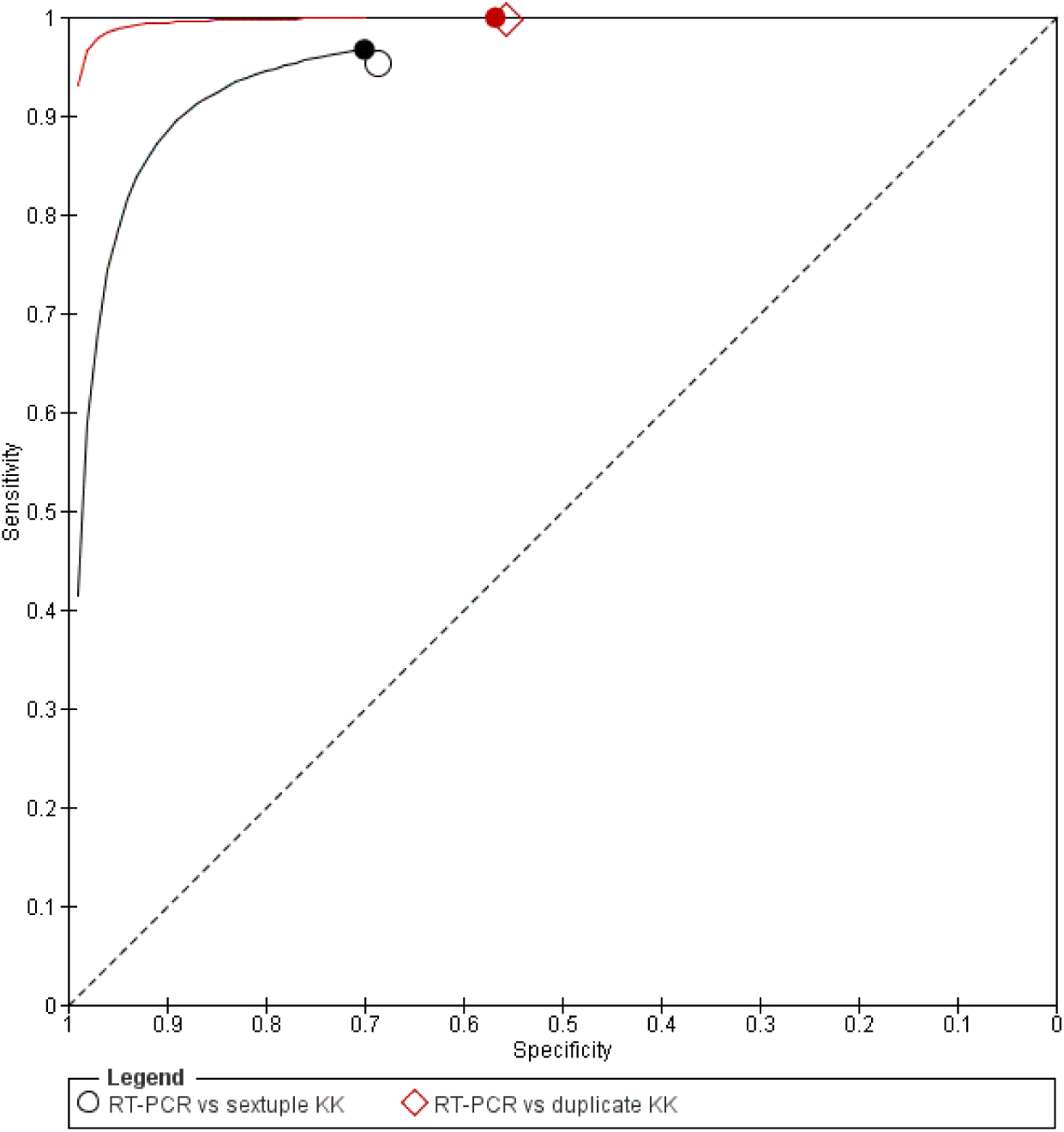
Summary ROC Plot of RT-PCR vs duplicate or sextuple KK

#### Publication Bias & Heterogeneity

For every test comparison, included in at least 4 studies, Publication Bias and Heterogeneity has been assessed by multiple methods.

In order to evaluate publication bias, funnel plots were created by plotting, for every test comparison, log-transformed Diagnostic Odds Ratio against its Standard Error. Additionally, given that standard funnel plots are less appropriate for DTA meta-analyses, Deeks’ funnel plot and asymmetry test have been used.

Heterogeneity was studied by using the *l*^S^ statistic and by performing Cochran’s Q tests on the Diagnostic Odds Ratio (DOR) as well as separately on sensitivity and specificity.

Results of both analyses (for test comparisons included in at least 4 studies) can be found in Appendix D.

#### CCA1

##### CCA1 cassette versus single Kato-Katz smear

1 study with a total of 217 individuals was found for this comparison. Prevalence was 5.35%.

##### CCA1 cassette versus duplicate Kato-Katz smears

14 studies totalling 17 comparisons with a total of 4884 individuals were found for this comparison. Prevalence ranged from 6% to 91%. The forest plot show heterogeneity which is also observed in the HSROC curve.

Meta analytic sensitivity and specificity of data were 85.47% and 59.09% respectively.

##### CCA1 cassette versus quadruplicate Kato-Katz smears

10 studies totalling 11 comparisons with a total of 4592 individuals were found for this comparison. Prevalence ranged from 6.37% to 60%. The forest plot show heterogeneity which is also observed in the HSROC curve.

Meta analytic sensitivity and specificity of data were 87.05% and 58.63% respectively.

##### CCA1 cassette versus sextuplicate Kato-Katz smears

7 studies with a total of 2325 individuals were found for this comparison. Prevalence ranged from 5.73% to 94.74%. The forest plot show heterogeneity which is also observed in the HSROC curve.

Meta analytic sensitivity and specificity of data were 83.48% and 67.86% respectively.

##### CCA1 cassette versus 16 Kato-Katz smears

1 study with a total of 217 individuals was found for this comparison. Prevalence was 14.29%.

##### CCA1 cassette versus single, duplicate, quadruplicate, sextuplicate and 16 Kato-Katz smears

There were 25 studies totalling 37 comparisons against single, duplicate, quadruplicate, sextuplicate or 16 Kato-Katz smears in 12,235 individuals. The prevalence ranged from 5.73% to 91%. The forest plot shows heterogeneity which is also observed in the HSROC curve.

Meta analytic sensitivity and specificity of data were 87.22% and 60.14% respectively.

##### CCA1 cassette versus Urine Microscopy

There were 4 studies with a total of 991 individuals. The prevalence ranged from 18.11% to 57.60%. The forest plot show heterogeneity which is also observed in the HSROC curve.

Meta analytic sensitivity and specificity of data were 51.43% and 74.13% respectively.

##### CCA1 cassette versus Helmintex

1 study with a total of 214 individuals was found for this comparison. Prevalence was 24.77%.

##### CCA1 cassette versus RT-PCR

1 study with a total of 196 individuals was found for this comparison. Prevalence was 55.10%.

#### CCA2

##### CCA2 cassette versus duplicate Kato-Katz smears

1 study with a total of 100 individuals was found for this comparison. Prevalence was 8%.

##### CCA2 cassette versus quadriplicate Kato-Katz smears

1 study with a total of 100 individuals was found for this comparison. Prevalence was 11%.

##### CCA2 cassette versus duplicate and quadruplicate Kato-Katz smears

There was 1 study totalling 2 comparisons against duplicate and quadruplicate Kato-Katz smears in 200 individuals. Prevalence ranged from 8% to 11%. The forest plots show heterogeneity which is also observed in the HSROC curve.

Meta analytic sensitivity and specificity of data were 51.75% and 87.59% respectively.

#### CAA

##### CAA cassette versus duplicate Kato-Katz smears

There were 2 studies totalling 3 comparisons against duplicate Kato-Katz smears in 830 individuals. Prevalence ranged from 28.43% to 91%. The forest plots show heterogeneity which is also observed in the HSROC curve.

Meta analytic sensitivity and specificity of data were 60.70% and 66.63% respectively.

##### CAA cassette versus quadriplicate Kato-Katz smears

1 study with a total of 377 individuals was found for this comparison. Prevalence was 6.37%.

##### CAA cassette versus duplicate and quadruplicate Kato-Katz smears

There were 3 studies totalling 4 comparisons against duplicate or quadruplicate Kato-Katz smears in 1207 individuals. Prevalence ranged from 6.37% to 91%. The forest plots show heterogeneity which is also observed in the HSROC curve.

Meta analytic sensitivity and specificity of data were 72.65% and 68.07% respectively.

##### CAA cassette vs Urine Microscopy

There were 4 studies with a total of 1247 individuals. Prevalence ranged from 18.11% to 57.60%. The forest plots show heterogeneity which is also observed in the HSROC curve.

Meta-analytic sensitivity and specificity of data were 70.93% and 78.57% respectively.

#### FLOTAC

##### FLOTAC (fresh) versus triplicate Kato-Katz smears

1 studies with a total of 112 individuals was found for this comparison. Prevalence was 64.29%.

##### FLOTAC (10 days) versus triplicate Kato-Katz smears

1 studies with a total of 112 individuals was found for this comparison. Prevalence was 64.29%.

##### FLOTAC (30 days) versus triplicate Kato-Katz smears

1 studies with a total of 112 individuals was found for this comparison. Prevalence was 64.29%.

##### FLOTAC (all) versus triplicate Kato-Katz smears

1 study with 3 different comparisons totaling 336 individuals was found for this comparison. Prevalence was 64.29%.

The forest plot show heterogeneity which is also observed in the HSROC curve.

Meta analytic sensitivity and specificity of data were 79.37% and 64.71% respectively.

#### SmCTF-RDT

##### SmCTF-RDT versus quadriplicate Kato-Katz smears

3 studies totalling 4 comparisons with a total of 291 individuals was found for this comparison. Prevalence ranged from 20.34% to 60%.

The forest plot show heterogeneity which is also observed in the HSROC curve.

Meta analytic sensitivity and specificity of data were 85.60% and 35.48% respectively.

##### SmCTF-RDT versus Urine Microscopy

1 study with a total of 117 individuals was found for this comparison. Prevalence was 5.13%.

#### Sm DNA PCR

##### Sm DNA PCR versus duplicate Kato-Katz smears

1 study with a total of 89 individuals was found for this comparison. Prevalence was 50.56%.

#### ELISA

##### SWAP ELISA versus sextuplicate Kato-Katz smears

1 study with a total of 482 individuals was found for this comparison. Prevalence was 38.80%.

##### IgM ELISA versus triplicate Kato-Katz smears

1 study with a total of 137 individuals was found for this comparison. Prevalence was 36.50%.

##### IgG ELISA versus triplicate Kato-Katz smears

4 studies with a total of 954 individuals were found for this comparison. Prevalence ranged from 36.50% to 93.64%.

The forest plot shows heterogeneity which is also observed in the HSROC curve.

Meta analytic sensitivity and specificity of data were 93.02% and 68.43% respectively.

#### Anti IgG RDT-Sh

##### Anti IgG RDT-Sh versus Urine Microscopy

1 study with a total of 160 individuals was found for this comparison. Prevalence was 51.25%.

#### Proteinuria (Reagent strip)

##### Proteinuria (Reagent strip) versus Urine microscopy

40 studies with 41 different test comparisons totalling 79,466 individuals were found for this comparison. Prevalence ranged from 4% to 88.57%.

The forest plots show heterogeneity which is also observed in the HSROC curve. Meta analytic sensitivity and specificity of data were 58.66% and 82.56% respectively.

#### Haematuria (Reagent strip)

##### Haematuria (Reagent strip) versus Urine microscopy

71 studies with 72 different test comparisons totalling 156,279 individuals were found for this comparison. Prevalence ranged from 3.23% to 86.93%.

The forest plot show heterogeneity which is also observed in the HSROC curve.

Meta analytic sensitivity and specificity of data were 74.38% and 86.78% respectively.

#### AWE-SEA ELISA

##### AWE-SEA ELISA versus quadriplicate Kato-Katz smears

2 studies with a total of 484 individuals were found for this comparison. Prevalence ranged from 6.37% to 21.50%. The forest plot did not show great heterogeneity as well as the HSROC curve. Meta analytic sensitivity and specificity of data were 93.74% and 64.17% respectively.

#### LAMP

##### LAMP versus triplicate Kato-Katz smears

2 studies with a total of 493 individuals was found for this comparison. Prevalence ranged from 45.95% to 100%.

Meta-analytic sensitivity and specificity of data were 94.29% and 98.23% respectively.

##### LAMP versus Urine Microscopy

2 studies with a total of 266 individuals was found for this comparison. Prevalence ranged from 26.60% to 50.58%.

Meta-analytic sensitivity and specificity of data were 77.06% and 63.50% respectively.

#### IHA

##### IHA versus triplicate Kato-Katz smears

1 study with a total of 2 comparisons and 203 individuals was found for this comparison. Prevalence ranged from 95.45% to 100%. The forest plot did not show great heterogeneity as well as the HSROC curve.

Meta analytic sensitivity and specificity of data were 81.48% and 7.15% respectively.

##### IHA versus Urine Microscopy

1 studies with a total of 145 individuals was found for this comparison. Prevalence was 60.96%. The forest plot did not show great heterogeneity as well as the HSROC curve.

#### Colorimetric test

##### Colorimetric test versus Urine Microscopy

1 study with a total of 1279 individuals was found for this comparison. Prevalence was 61.85%.

#### rSP13 ELISA

##### rSP13 ELISA versus 27 Kato-Katz smears

1 study with a total of 1371 individuals was found for this comparison. Prevalence was 5.40%.

#### IgG SEA-ELISA

##### IgG SEA-ELISA vs CCA1 cassette

1 study with a total of 258 individuals was found for this comparison. Prevalence was 56.98%.

##### IgG SEA-ELISA vs Urine Microscopy

4 studies with a total of 503 individuals were found for this comparison. Prevalence ranged from 30.67% to 78%. The forest plot did not show great heterogeneity as well as the HSROC curve.

Meta analytic sensitivity and specificity of data were 88.94% and 71.29% respectively.

#### Leukocyturia (reagent strip)

##### Leukocyturia (reagent strip) vs Urine Microscopy

5 studies with a total of 1532 individuals were found for this comparison. Prevalence ranged from 4% to 76.78%. The forest plot did not show great heterogeneity as well as the HSROC curve.

Meta analytic sensitivity and specificity of data were 55.70% and 59.95% respectively.

#### COPT

##### COPT vs duplicate Kato-Katz smears

1 study with a total of 572 individuals was found for this comparison. Prevalence was 0.87%.

#### PCR

##### PCR vs Kato-Katz smears

2 studies with a total of 3 comparisons and 551 individuals were found for this comparison. Prevalence ranged from 12.88% to 85.19%.

Meta analytic sensitivity and specificity of data were 95.79% and 46.92% respectively.

##### PCR vs CCA1 cassette

1 study with a total of 258 individuals was found for this comparison. Prevalence was 56.98%.

#### Helmintex

##### Helmintex vs duplicate KK

1 study with a total of 461 individuals was found for this comparison. Prevalence was 11.93%.

##### Helmintex vs RT-PCR

1 study with a total of 176 individuals was found for this comparison. Prevalence was 59.09%.

#### DDIA

##### DDIA vs Urine Microscopy

1 study with a total of 146 individuals was found for this comparison. Prevalence was 60.96%.

#### RT-PCR

##### RT-PCR vs duplicate KK

1 study with a total of 206 individuals was found for this comparison. Prevalence was 19.90%.

##### RT-PCR vs sextuple KK

1 study with a total of 198 individuals was found for this comparison. Prevalence was 32.32%.

### Analyses with the bayesian bivariate random-effects HSROC model

The Bayesian bivariate random effects hierarchical summary ROC model allowed to identify the sensitivity and specificity for each comparisons previously made. It gave for each of them the meta-analystic estimates of sensitivity and specificity. The results are shown in Table 4 (raw outputs of the model) and Table 5 (cf. Appendix A) with the evaluation of sensitivity and specificity with their 95% Bayesian Credible Intervals.

Results are not valid when the number of studies is 1. In that case the result in the forest plot is to be considered.

## Discussion

### Summary of main results

The review gathered 203 articles stating a diagnostic test for the diagnosis of S. haematobium and S. mansoni out of which 114 entered the analyses. Microscopy of Urine Microscopy or Kato-Katz smears were used as reference standards.

There was a great heterogeneity in the reference standards used over the studies. For S. mansoni, duplicate to sextuplicate Kato-Katz smears and even 16 or 27 smears were used. For S. haematobium Urine Microscopy could be simple or double.

However concerning S. mansoni CCA1 (Se=87%; Sp=60%) seems to show higher superiority in sensitivity and specificity (estimated by the bayesian bivariate model) than CCA2 (Se=52%; Sp=88%) as compared with Kato-Katz smears.

SmCTF-RDT (Se=86%; Sp=36%) has a similar sensitivity but a much lower specificity.

IgG ELISA (Se=93%; Sp=68%) is comparable to CCA1 but there is only 4 studies in this comparison.

AWE-SEA ELISA (Se=94%; Sp=64%) has also a very ability to detect true positive but with only two studies in the comparison.

Based on 4 studies of 1207 individuals CAA cassette was comparable (Se=73%; Sp=68%) to FLOTAC (Se=79%; Sp=65%).

SWAP ELISA evaluated in 1 study of 482 individual showed a sensitivity of 92% and specificity of 57%.

For S. haematobium proteinuria (Se=59%; Sp=83%) and haematuria (Se=74%; Sp=87%) reagent strips showed reasonably high specificities with a considerable better sensitivity for the haematuria test.

The colorimetric test gave interesting results despite low sensitivity (Se=52%; Sp=75%)

The new tests would have better feasibility (i.e. commercialized and available) and should be the other argument for choosing them on top of this analysis. Other laboratory tests (e.g. various ELISAs) that are not ready for public health application are less important in the short term.

## Limitations

- Imperfect standard tests
- Few number of studies and sparse analyses for some test comparisons
- Limited sample size and prevalence not accounted for in the analyses
- While higher eggs means higher sensitivity and population prevalence, there is a relation between sensitivity/specificity and prevalence. This relation was not accounted for in the analysis due the limited sample size

## Evidence to Recommendation

Certainty of the evidence of

- **accuracy**: moderate for CCA, SmCTF-RDT, Proteinuria, Haematuria
- **Values**: Probably no important uncertainty or variability. Prevalences as evaluated by the reference test (Kato-Katz smear or urine microscopy) were always underestimated when compared to the index tests. In general it was below 50% but a few studies showed a prevalence higher than 50%. Only one study in Nigeria (Ugbomoiko 2009) showed lower prevalences with the index test (proteinuria and haematuria against urine microscopy).
- **Balance of effects**: Probably favors the intervention
- **Resources required**: depends on the tests’ prices, no evidence. Concerning S. mansoni PCO test such as CCA1, CCA2, CAA, SmCTF-RDT would be less resource intensive than IgG ELISA, AWE-SEA ELISA, SWAP ELISA which need a local lab or to be sent to a central lab. For S. haematobium proteinuria and haematuria reagent strips can be used on site while the haematuria test cannot. The colorimetric test is also possible in the field.
- **Equity**: Probably increased. The use of a diagnostic test will not provide more equity in itself. A diagnostic test that best detect schistosomiasis pathogen may allow mass deworming toward elimination. However as Helminthiases occurs in poor communities, concurrent interventions oriented against degraded and unhealthy living conditions as well as gender equity to break the cycle of chronic poverty and schistosomiasis infections
- **Acceptability**: Yes. The acceptability of diagnostic test may mostly depends on the cost to be scaled up
- **Feasibility**: Yes if RDT
- Benefits and harms:

Potential benefits: Concerning S. mansoni CCA1 (Se=88%; Sp=59%) seems to show higher superiority in sensitivity and specificity (estimated by the bayesian bivariate model) than CCA2 (Se=52%; Sp=88%) as compared with Kato-Katz smears. SmCTF-RDT (Se=86%; Sp=36%) has a similar sensitivity but a much lower specificity. IgG ELISA (Se=90%; Sp=76%) is comparable to CCA1 but there is only 4 studies in this comparison. AWE-SEA ELISA (Se=94%; Sp=64%) has also a very ability to detect true positive but with only two studies in the comparison. Based on 4 studies of 1207 individuals CAA cassette was comparable (Se=73%; Sp=68%) to FLOTAC (Se=79%; Sp=65%).

SWAP ELISA evaluated in 1 study of 482 individual showed a sensitivity of 92% and specificity of 57%.

For S. haematobium proteinuria (Se=58%; Sp=82%) and haematuria (Se=74%; Sp=86%) reagent strips showed reasonably high specificities with a considerable better sensitivity for the haematuria test. The colorimetric test gave interesting results despite low sensitivity (Se=52%; Sp=75%).

Potential harms: No potential harm

**Type of Recommendation:** Conditional recommendation for CCA. Conventional diagnostic tool in humans of Kato-Katz and CCA for S. mansoni and urine microscopy for S. haematobium have reasonable sensitivity and excellent specificity. Conventional diagnostic tool in humans are well accepted, low-cost, and feasible given their widespread implementation. New diagnostic tools, such as molecular-based and immunologic diagnostics, lack sufficient data on sensitivity and specificity, and their utility is further limited by challenges with feasibility and resource implications.

## Supporting information

Characteristics of studies

Summary of Findings tables

Heterogeneity & publication bias

## Data Availability

Data used in this study are available in the appendices

## Notes

### Competing Interest Statement

The authors have declared no competing interest.

### Clinical Trial

the study was expected to become a paper and since october 2019 PROSPERO register for reviews require that serch is not started at the time of submission

### Funding Statement

The study was funded by a grant of the Department of Control of Neglected Tropical Diseases of the World Health Organisation

### Author Declarations

individual studies received IRB approval

