## Supplementary material for "Diagnostic tests for Schistosomiasis for low prevalence settings: a systematic review and Meta-Analysis": Characteristics of studies

### Characteristics of included studies

#### Abdel-Wahab 1992

##### Patient Selection

| **A. Risk of Bias** | | |
| --- | --- | --- |
| Patient Sampling | cross-sectional study | |
| Was a consecutive or random sample of patients enrolled? | | Unclear |
| Was a case-control design avoided? | | Yes |
| Did the study avoid inappropriate exclusions? | | Unclear |
| **Could the selection of patients have introduced bias?** | | Low risk |

| **B. Concerns regarding applicability** | | |
| --- | --- | --- |
| Patient characteristics and setting | - 422 school children aged between 12 and 16 years  - Country: Egypt  - Setting: field study  - Praziquantel treatment: over half of the children have a history of receiving praziquantel treatment during the preceding 2 years | |
| **Are there concerns that the included patients and setting do not match the review question?** | | Low concern |

##### Index Test

| Index tests | micro-haematuria, proteinuria, leukocyturia by urine reagent strip |
| --- | --- |

##### All tests

| **A. Risk of Bias** | |
| --- | --- |
| Were the index test results interpreted without knowledge of the results of the reference standard? | Unclear |
| If a threshold was used, was it pre-specified? | Unclear |
| **Could the conduct or interpretation of the index test have introduced bias?** | Low risk |

| **B. Concerns regarding applicability** | |
| --- | --- |
| **Are there concerns that the index test, its conduct, or interpretation differ from the review question?** | Low concern |

##### Reference Standard

| **A. Risk of Bias** | | |
| --- | --- | --- |
| Target condition and reference standard(s) | S. haematobium  urine microscopy | |
| Is the reference standards likely to correctly classify the target condition? | | Unclear |
| Were the reference standard results interpreted without knowledge of the results of the index tests? | | Unclear |
| **Could the reference standard, its conduct, or its interpretation have introduced bias?** | | Low risk |

| **B. Concerns regarding applicability** | |
| --- | --- |
| **Are there concerns that the target condition as defined by the reference standard does not match the question?** | Low concern |

##### Flow and Timing

| **A. Risk of Bias** | | |
| --- | --- | --- |
| Flow and timing |  | |
| Was there an appropriate interval between index test and reference standard? | | Unclear |
| Did all patients receive the same reference standard? | | Yes |
| Were all patients included in the analysis? | | Unclear |
| **Could the patient flow have introduced bias?** | | Low risk |

##### Notes

| **Notes** |
| --- |

#### Abdel-Wahab 2000

##### Patient Selection

| **A. Risk of Bias** | | |
| --- | --- | --- |
| Patient Sampling | Cross-sectional study ; multi-stage stratified random sampling | |
| Was a consecutive or random sample of patients enrolled? | | Yes |
| Was a case-control design avoided? | | Yes |
| Did the study avoid inappropriate exclusions? | | Yes |
| **Could the selection of patients have introduced bias?** | | Low risk |

| **B. Concerns regarding applicability** | | |
| --- | --- | --- |
| Patient characteristics and setting | - 5214 residents from 5 villages and 16 ezbas in Fayoum Governorate (aged between 5 and 25 years)  - Country: Egypt | |
| **Are there concerns that the included patients and setting do not match the review question?** | | Low concern |

##### Index Test

| Index tests | Haematuria and proteinuria reagent strips |
| --- | --- |

##### All tests

| **A. Risk of Bias** | |
| --- | --- |
| Were the index test results interpreted without knowledge of the results of the reference standard? | Unclear |
| If a threshold was used, was it pre-specified? | Unclear |
| **Could the conduct or interpretation of the index test have introduced bias?** | Unclear risk |

| **B. Concerns regarding applicability** | |
| --- | --- |
| **Are there concerns that the index test, its conduct, or interpretation differ from the review question?** | Low concern |

##### Reference Standard

| **A. Risk of Bias** | | |
| --- | --- | --- |
| Target condition and reference standard(s) | S. haematobium  Urine microscopy | |
| Is the reference standards likely to correctly classify the target condition? | | Unclear |
| Were the reference standard results interpreted without knowledge of the results of the index tests? | | Unclear |
| **Could the reference standard, its conduct, or its interpretation have introduced bias?** | | Low risk |

| **B. Concerns regarding applicability** | |
| --- | --- |
| **Are there concerns that the target condition as defined by the reference standard does not match the question?** | Low concern |

##### Flow and Timing

| **A. Risk of Bias** | | |
| --- | --- | --- |
| Flow and timing |  | |
| Was there an appropriate interval between index test and reference standard? | | Yes |
| Did all patients receive the same reference standard? | | Yes |
| Were all patients included in the analysis? | | Yes |
| **Could the patient flow have introduced bias?** | | Low risk |

##### Notes

| **Notes** |
| --- |

#### Adriko 2014

##### Patient Selection

| **A. Risk of Bias** | | |
| --- | --- | --- |
| Patient Sampling | cross-sectional study; random selection | |
| Was a consecutive or random sample of patients enrolled? | | Yes |
| Was a case-control design avoided? | | Yes |
| Did the study avoid inappropriate exclusions? | | Yes |
| **Could the selection of patients have introduced bias?** | | Low risk |

| **B. Concerns regarding applicability** | | |
| --- | --- | --- |
| Patient characteristics and setting | Species: S. mansoni  Country: Uganda  Sample size:  Age range: 7–13 years  Participants: schoolchildren  Setting: field study  Praziquantel before the study: Annual mass treatment with praziquantel had been administered in the area for five years before the study begun and the campaign had reduced the endemicity of the disease from high to medium levels | |
| **Are there concerns that the included patients and setting do not match the review question?** | | Low concern |

##### Index Test

| Index tests | CCA1 & CCA2 |
| --- | --- |

##### All tests

| **A. Risk of Bias** | |
| --- | --- |
| Were the index test results interpreted without knowledge of the results of the reference standard? | No |
| If a threshold was used, was it pre-specified? | Yes |
| **Could the conduct or interpretation of the index test have introduced bias?** | Low risk |

| **B. Concerns regarding applicability** | |
| --- | --- |
| **Are there concerns that the index test, its conduct, or interpretation differ from the review question?** | Low concern |

##### Reference Standard

| **A. Risk of Bias** | | |
| --- | --- | --- |
| Target condition and reference standard(s) | Duplicate Kato Katz smear | |
| Is the reference standards likely to correctly classify the target condition? | | No |
| Were the reference standard results interpreted without knowledge of the results of the index tests? | | Yes |
| **Could the reference standard, its conduct, or its interpretation have introduced bias?** | | Low risk |

| **B. Concerns regarding applicability** | |
| --- | --- |
| **Are there concerns that the target condition as defined by the reference standard does not match the question?** | Low concern |

##### Flow and Timing

| **A. Risk of Bias** | | |
| --- | --- | --- |
| Flow and timing |  | |
| Was there an appropriate interval between index test and reference standard? | | Yes |
| Did all patients receive the same reference standard? | | Yes |
| Were all patients included in the analysis? | | Yes |
| **Could the patient flow have introduced bias?** | | Low risk |

##### Notes

| **Notes** |
| --- |

#### Al-Shehri 2018

##### Patient Selection

| **A. Risk of Bias** | | |
| --- | --- | --- |
| Patient Sampling | cross-sectional study ; field sampling | |
| Was a consecutive or random sample of patients enrolled? | | Yes |
| Was a case-control design avoided? | | Yes |
| Did the study avoid inappropriate exclusions? | | Unclear |
| **Could the selection of patients have introduced bias?** | | Low risk |

| **B. Concerns regarding applicability** | | |
| --- | --- | --- |
| Patient characteristics and setting | - 258 children from 5 different primary schools aged between 5 and 10 years  - Period: May 2015  - Country: Uganda  - Setting: field study | |
| **Are there concerns that the included patients and setting do not match the review question?** | | Low concern |

##### Index Test

| Index tests | duplicate KK, ELISA-SEA, DNA TaqMan (real-time PCR) |
| --- | --- |

##### All tests

| **A. Risk of Bias** | |
| --- | --- |
| Were the index test results interpreted without knowledge of the results of the reference standard? | Yes |
| If a threshold was used, was it pre-specified? | Unclear |
| **Could the conduct or interpretation of the index test have introduced bias?** | Low risk |

| **B. Concerns regarding applicability** | |
| --- | --- |
| **Are there concerns that the index test, its conduct, or interpretation differ from the review question?** | Low concern |

##### Reference Standard

| **A. Risk of Bias** | | |
| --- | --- | --- |
| Target condition and reference standard(s) | S. mansoni  urine-CCA dipsticks | |
| Is the reference standards likely to correctly classify the target condition? | | Unclear |
| Were the reference standard results interpreted without knowledge of the results of the index tests? | | Unclear |
| **Could the reference standard, its conduct, or its interpretation have introduced bias?** | | Unclear risk |

| **B. Concerns regarding applicability** | |
| --- | --- |
| **Are there concerns that the target condition as defined by the reference standard does not match the question?** | Low concern |

##### Flow and Timing

| **A. Risk of Bias** | | |
| --- | --- | --- |
| Flow and timing |  | |
| Was there an appropriate interval between index test and reference standard? | | Yes |
| Did all patients receive the same reference standard? | | Yes |
| Were all patients included in the analysis? | | No |
| **Could the patient flow have introduced bias?** | | Low risk |

##### Notes

| **Notes** |
| --- |

#### Al-Sherbiny 1999

##### Patient Selection

| **A. Risk of Bias** | | |
| --- | --- | --- |
| Patient Sampling | cross-sectional study | |
| Was a consecutive or random sample of patients enrolled? | | Yes |
| Was a case-control design avoided? | | Yes |
| Did the study avoid inappropriate exclusions? | | Unclear |
| **Could the selection of patients have introduced bias?** | | Low risk |

| **B. Concerns regarding applicability** | | |
| --- | --- | --- |
| Patient characteristics and setting | - 1173 patients  - Country: Nigeria  - Setting: field study | |
| **Are there concerns that the included patients and setting do not match the review question?** | | Low concern |

##### Index Test

| Index tests | urine CCA ELISA & urine CAA ELISA |
| --- | --- |

##### All tests

| **A. Risk of Bias** | |
| --- | --- |
| Were the index test results interpreted without knowledge of the results of the reference standard? | Unclear |
| If a threshold was used, was it pre-specified? | Yes |
| **Could the conduct or interpretation of the index test have introduced bias?** | Low risk |

| **B. Concerns regarding applicability** | |
| --- | --- |
| **Are there concerns that the index test, its conduct, or interpretation differ from the review question?** | Low concern |

##### Reference Standard

| **A. Risk of Bias** | | |
| --- | --- | --- |
| Target condition and reference standard(s) | S. haematobium  urine microscopy | |
| Is the reference standards likely to correctly classify the target condition? | | Unclear |
| Were the reference standard results interpreted without knowledge of the results of the index tests? | | Unclear |
| **Could the reference standard, its conduct, or its interpretation have introduced bias?** | | Low risk |

| **B. Concerns regarding applicability** | |
| --- | --- |
| **Are there concerns that the target condition as defined by the reference standard does not match the question?** | Low concern |

##### Flow and Timing

| **A. Risk of Bias** | | |
| --- | --- | --- |
| Flow and timing |  | |
| Was there an appropriate interval between index test and reference standard? | | Unclear |
| Did all patients receive the same reference standard? | | Yes |
| Were all patients included in the analysis? | | No |
| **Could the patient flow have introduced bias?** | | Low risk |

##### Notes

| **Notes** |
| --- |

#### Anosike 2001

##### Patient Selection

| **A. Risk of Bias** | | |
| --- | --- | --- |
| Patient Sampling | cross-sectional study ; consecutive sampling | |
| Was a consecutive or random sample of patients enrolled? | | Yes |
| Was a case-control design avoided? | | Yes |
| Did the study avoid inappropriate exclusions? | | Yes |
| **Could the selection of patients have introduced bias?** | | Low risk |

| **B. Concerns regarding applicability** | | |
| --- | --- | --- |
| Patient characteristics and setting | - 1173 participants from households in 7 communities  - Country: Nigeria  - Setting: field study | |
| **Are there concerns that the included patients and setting do not match the review question?** | | Low concern |

##### Index Test

| Index tests | micro-haematuria by urine reagent strip |
| --- | --- |

##### All tests

| **A. Risk of Bias** | |
| --- | --- |
| Were the index test results interpreted without knowledge of the results of the reference standard? | Unclear |
| If a threshold was used, was it pre-specified? | Yes |
| **Could the conduct or interpretation of the index test have introduced bias?** | Unclear risk |

| **B. Concerns regarding applicability** | |
| --- | --- |
| **Are there concerns that the index test, its conduct, or interpretation differ from the review question?** | Low concern |

##### Reference Standard

| **A. Risk of Bias** | | |
| --- | --- | --- |
| Target condition and reference standard(s) | S. haematobium  urine microscopy | |
| Is the reference standards likely to correctly classify the target condition? | | Unclear |
| Were the reference standard results interpreted without knowledge of the results of the index tests? | | Unclear |
| **Could the reference standard, its conduct, or its interpretation have introduced bias?** | | Low risk |

| **B. Concerns regarding applicability** | |
| --- | --- |
| **Are there concerns that the target condition as defined by the reference standard does not match the question?** | Low concern |

##### Flow and Timing

| **A. Risk of Bias** | | |
| --- | --- | --- |
| Flow and timing |  | |
| Was there an appropriate interval between index test and reference standard? | | Yes |
| Did all patients receive the same reference standard? | | Yes |
| Were all patients included in the analysis? | | Yes |
| **Could the patient flow have introduced bias?** | | Low risk |

##### Notes

| **Notes** |
| --- |

#### Anyan 2020

##### Patient Selection

| **A. Risk of Bias** | | |
| --- | --- | --- |
| Patient Sampling | cross-sectional study | |
| Was a consecutive or random sample of patients enrolled? | | Unclear |
| Was a case-control design avoided? | | Yes |
| Did the study avoid inappropriate exclusions? | | Unclear |
| **Could the selection of patients have introduced bias?** | | Unclear risk |

| **B. Concerns regarding applicability** | | |
| --- | --- | --- |
| Patient characteristics and setting | - 163 patients (93 male, 70 female) aged between 3 and 63 years (mostly children)  - Country: Ghana  - Setting: field study | |
| **Are there concerns that the included patients and setting do not match the review question?** | | Low concern |

##### Index Test

| Index tests | urine PCR |
| --- | --- |

##### All tests

| **A. Risk of Bias** | |
| --- | --- |
| Were the index test results interpreted without knowledge of the results of the reference standard? | Unclear |
| If a threshold was used, was it pre-specified? | Unclear |
| **Could the conduct or interpretation of the index test have introduced bias?** | Low risk |

| **B. Concerns regarding applicability** | |
| --- | --- |
| **Are there concerns that the index test, its conduct, or interpretation differ from the review question?** | Low concern |

##### Reference Standard

| **A. Risk of Bias** | | |
| --- | --- | --- |
| Target condition and reference standard(s) | S. mansoni & S. hamatobium  Kati-Katz stool examination | |
| Is the reference standards likely to correctly classify the target condition? | | Unclear |
| Were the reference standard results interpreted without knowledge of the results of the index tests? | | Unclear |
| **Could the reference standard, its conduct, or its interpretation have introduced bias?** | | Low risk |

| **B. Concerns regarding applicability** | |
| --- | --- |
| **Are there concerns that the target condition as defined by the reference standard does not match the question?** | Low concern |

##### Flow and Timing

| **A. Risk of Bias** | | |
| --- | --- | --- |
| Flow and timing |  | |
| Was there an appropriate interval between index test and reference standard? | | Yes |
| Did all patients receive the same reference standard? | | Yes |
| Were all patients included in the analysis? | | Unclear |
| **Could the patient flow have introduced bias?** | | Low risk |

##### Notes

| **Notes** |
| --- |

#### Aryeetey 2000

##### Patient Selection

| **A. Risk of Bias** | | |
| --- | --- | --- |
| Patient Sampling | cross-sectional study | |
| Was a consecutive or random sample of patients enrolled? | | Unclear |
| Was a case-control design avoided? | | Yes |
| Did the study avoid inappropriate exclusions? | | Unclear |
| **Could the selection of patients have introduced bias?** | | Low risk |

| **B. Concerns regarding applicability** | | |
| --- | --- | --- |
| Patient characteristics and setting | - 370 residents from 3 study areas aged older than 5 years  - Country: Ghana  - Setting: field study | |
| **Are there concerns that the included patients and setting do not match the review question?** | | Low concern |

##### Index Test

| Index tests | micro-haematuria & proteinuria reagent strips |
| --- | --- |

##### All tests

| **A. Risk of Bias** | |
| --- | --- |
| Were the index test results interpreted without knowledge of the results of the reference standard? | Yes |
| If a threshold was used, was it pre-specified? | Yes |
| **Could the conduct or interpretation of the index test have introduced bias?** | Low risk |

| **B. Concerns regarding applicability** |
| --- |
| **Are there concerns that the index test, its conduct, or interpretation differ from the review question?** |

##### Reference Standard

| **A. Risk of Bias** | | |
| --- | --- | --- |
| Target condition and reference standard(s) | S. haematobium  urine microscopy | |
| Is the reference standards likely to correctly classify the target condition? | | Unclear |
| Were the reference standard results interpreted without knowledge of the results of the index tests? | | Unclear |
| **Could the reference standard, its conduct, or its interpretation have introduced bias?** | | Unclear risk |

| **B. Concerns regarding applicability** | |
| --- | --- |
| **Are there concerns that the target condition as defined by the reference standard does not match the question?** | Low concern |

##### Flow and Timing

| **A. Risk of Bias** | | |
| --- | --- | --- |
| Flow and timing |  | |
| Was there an appropriate interval between index test and reference standard? | | Yes |
| Did all patients receive the same reference standard? | | Yes |
| Were all patients included in the analysis? | | Unclear |
| **Could the patient flow have introduced bias?** | | Low risk |

##### Notes

| **Notes** |
| --- |

#### Assaré 2018

##### Patient Selection

| **A. Risk of Bias** | | |
| --- | --- | --- |
| Patient Sampling | cross-sectional study | |
| Was a consecutive or random sample of patients enrolled? | | Yes |
| Was a case-control design avoided? | | Yes |
| Did the study avoid inappropriate exclusions? | | Unclear |
| **Could the selection of patients have introduced bias?** | | Low risk |

| **B. Concerns regarding applicability** | | |
| --- | --- | --- |
| Patient characteristics and setting | - 681 children between 9 and 12 years from 14 schoold purposely selected based on low prevalence  - 2 or 4 praziquantel treatments during previous 4 years  - Country: Côte d'Ivoire  - Period: October 2016  - Setting: field study | |
| **Are there concerns that the included patients and setting do not match the review question?** | | Low concern |

##### Index Test

| Index tests | POC-CCA |
| --- | --- |

##### All tests

| **A. Risk of Bias** | |
| --- | --- |
| Were the index test results interpreted without knowledge of the results of the reference standard? | Unclear |
| If a threshold was used, was it pre-specified? | Unclear |
| **Could the conduct or interpretation of the index test have introduced bias?** | Low risk |

| **B. Concerns regarding applicability** | |
| --- | --- |
| **Are there concerns that the index test, its conduct, or interpretation differ from the review question?** | Low concern |

##### Reference Standard

| **A. Risk of Bias** | | |
| --- | --- | --- |
| Target condition and reference standard(s) | S. mansoni  sextuple KK | |
| Is the reference standards likely to correctly classify the target condition? | | Yes |
| Were the reference standard results interpreted without knowledge of the results of the index tests? | | Yes |
| **Could the reference standard, its conduct, or its interpretation have introduced bias?** | | Low risk |

| **B. Concerns regarding applicability** | |
| --- | --- |
| **Are there concerns that the target condition as defined by the reference standard does not match the question?** | Low concern |

##### Flow and Timing

| **A. Risk of Bias** | | |
| --- | --- | --- |
| Flow and timing |  | |
| Was there an appropriate interval between index test and reference standard? | | Yes |
| Did all patients receive the same reference standard? | | Yes |
| Were all patients included in the analysis? | | Unclear |
| **Could the patient flow have introduced bias?** | | Low risk |

##### Notes

| **Notes** |
| --- |

#### Ayele 2008

##### Patient Selection

| **A. Risk of Bias** | | |
| --- | --- | --- |
| Patient Sampling | cross-sectional study | |
| Was a consecutive or random sample of patients enrolled? | | Unclear |
| Was a case-control design avoided? | | Yes |
| Did the study avoid inappropriate exclusions? | | Yes |
| **Could the selection of patients have introduced bias?** | | Low risk |

| **B. Concerns regarding applicability** | | |
| --- | --- | --- |
| Patient characteristics and setting | - 206 school children aged between 4 and 21 years  - Country: Ethiopia  - Setting: field study | |
| **Are there concerns that the included patients and setting do not match the review question?** | | Low concern |

##### Index Test

| Index tests | CCA strip |
| --- | --- |

##### All tests

| **A. Risk of Bias** | |
| --- | --- |
| Were the index test results interpreted without knowledge of the results of the reference standard? | Unclear |
| If a threshold was used, was it pre-specified? | Unclear |
| **Could the conduct or interpretation of the index test have introduced bias?** | Low risk |

| **B. Concerns regarding applicability** | |
| --- | --- |
| **Are there concerns that the index test, its conduct, or interpretation differ from the review question?** | Low concern |

##### Reference Standard

| **A. Risk of Bias** | | |
| --- | --- | --- |
| Target condition and reference standard(s) | S. haematobium  Urine microscopy | |
| Is the reference standards likely to correctly classify the target condition? | | Unclear |
| Were the reference standard results interpreted without knowledge of the results of the index tests? | | Unclear |
| **Could the reference standard, its conduct, or its interpretation have introduced bias?** | | Unclear risk |

| **B. Concerns regarding applicability** | |
| --- | --- |
| **Are there concerns that the target condition as defined by the reference standard does not match the question?** | Low concern |

##### Flow and Timing

| **A. Risk of Bias** | | |
| --- | --- | --- |
| Flow and timing |  | |
| Was there an appropriate interval between index test and reference standard? | | Yes |
| Did all patients receive the same reference standard? | | Yes |
| Were all patients included in the analysis? | | Yes |
| **Could the patient flow have introduced bias?** | | Low risk |

##### Notes

| **Notes** |
| --- |

#### Barakat 1983

##### Patient Selection

| **A. Risk of Bias** | | |
| --- | --- | --- |
| Patient Sampling | cross-sectional study | |
| Was a consecutive or random sample of patients enrolled? | | Unclear |
| Was a case-control design avoided? | | Unclear |
| Did the study avoid inappropriate exclusions? | | Unclear |
| **Could the selection of patients have introduced bias?** | | Unclear risk |

| **B. Concerns regarding applicability** | | |
| --- | --- | --- |
| Patient characteristics and setting | - 507 school children from 2 schools aged between 6 and 12 years  - Country: Egypt  - Setting: field study | |
| **Are there concerns that the included patients and setting do not match the review question?** | | Low concern |

##### Index Test

| Index tests | ELISA IgG |
| --- | --- |

##### All tests

| **A. Risk of Bias** | |
| --- | --- |
| Were the index test results interpreted without knowledge of the results of the reference standard? | Unclear |
| If a threshold was used, was it pre-specified? | Unclear |
| **Could the conduct or interpretation of the index test have introduced bias?** | Low risk |

| **B. Concerns regarding applicability** | |
| --- | --- |
| **Are there concerns that the index test, its conduct, or interpretation differ from the review question?** | Low concern |

##### Reference Standard

| **A. Risk of Bias** | | |
| --- | --- | --- |
| Target condition and reference standard(s) | S. mansoni  triplicate Kato-Katz | |
| Is the reference standards likely to correctly classify the target condition? | | Unclear |
| Were the reference standard results interpreted without knowledge of the results of the index tests? | | Unclear |
| **Could the reference standard, its conduct, or its interpretation have introduced bias?** | | Low risk |

| **B. Concerns regarding applicability** | |
| --- | --- |
| **Are there concerns that the target condition as defined by the reference standard does not match the question?** | Low concern |

##### Flow and Timing

| **A. Risk of Bias** | | |
| --- | --- | --- |
| Flow and timing |  | |
| Was there an appropriate interval between index test and reference standard? | | Yes |
| Did all patients receive the same reference standard? | | Yes |
| Were all patients included in the analysis? | | Unclear |
| **Could the patient flow have introduced bias?** | | Low risk |

##### Notes

| **Notes** |
| --- |

#### Bassiouny 2014

##### Patient Selection

| **A. Risk of Bias** | | |
| --- | --- | --- |
| Patient Sampling | cross-sectional study | |
| Was a consecutive or random sample of patients enrolled? | | Unclear |
| Was a case-control design avoided? | | Yes |
| Did the study avoid inappropriate exclusions? | | Unclear |
| **Could the selection of patients have introduced bias?** | | Low risk |

| **B. Concerns regarding applicability** | | |
| --- | --- | --- |
| Patient characteristics and setting | - Country: Yemen  - 696 primary school children aged between 10 and 16 years | |
| **Are there concerns that the included patients and setting do not match the review question?** | | Low concern |

##### Index Test

| Index tests | Microhaematuria (by dipstick) |
| --- | --- |

##### All tests

| **A. Risk of Bias** | |
| --- | --- |
| Were the index test results interpreted without knowledge of the results of the reference standard? | Yes |
| If a threshold was used, was it pre-specified? | Unclear |
| **Could the conduct or interpretation of the index test have introduced bias?** | Low risk |

| **B. Concerns regarding applicability** | |
| --- | --- |
| **Are there concerns that the index test, its conduct, or interpretation differ from the review question?** | Low concern |

##### Reference Standard

| **A. Risk of Bias** | | |
| --- | --- | --- |
| Target condition and reference standard(s) | S. haematobium  Urine microscopy | |
| Is the reference standards likely to correctly classify the target condition? | | Unclear |
| Were the reference standard results interpreted without knowledge of the results of the index tests? | | Yes |
| **Could the reference standard, its conduct, or its interpretation have introduced bias?** | | Unclear risk |

| **B. Concerns regarding applicability** | |
| --- | --- |
| **Are there concerns that the target condition as defined by the reference standard does not match the question?** | Low concern |

##### Flow and Timing

| **A. Risk of Bias** | | |
| --- | --- | --- |
| Flow and timing |  | |
| Was there an appropriate interval between index test and reference standard? | | Yes |
| Did all patients receive the same reference standard? | | Yes |
| Were all patients included in the analysis? | | No |
| **Could the patient flow have introduced bias?** | | Low risk |

##### Notes

| **Notes** |
| --- |

#### Bezerra 2020

##### Patient Selection

| **A. Risk of Bias** | | |
| --- | --- | --- |
| Patient Sampling | cross-sectional study | |
| Was a consecutive or random sample of patients enrolled? | | Yes |
| Was a case-control design avoided? | | Yes |
| Did the study avoid inappropriate exclusions? | | Unclear |
| **Could the selection of patients have introduced bias?** | | Low risk |

| **B. Concerns regarding applicability** | | |
| --- | --- | --- |
| Patient characteristics and setting | - 127 patients (55 male, 72 female)  - Country: Brazil  - Setting: field study | |
| **Are there concerns that the included patients and setting do not match the review question?** | | Low concern |

##### Index Test

| Index tests | POC-CCA |
| --- | --- |

##### All tests

| **A. Risk of Bias** | |
| --- | --- |
| Were the index test results interpreted without knowledge of the results of the reference standard? | Unclear |
| If a threshold was used, was it pre-specified? | No |
| **Could the conduct or interpretation of the index test have introduced bias?** | Low risk |

| **B. Concerns regarding applicability** | |
| --- | --- |
| **Are there concerns that the index test, its conduct, or interpretation differ from the review question?** | Low concern |

##### Reference Standard

| **A. Risk of Bias** | | |
| --- | --- | --- |
| Target condition and reference standard(s) | S. mansoni  Sextuple Kato-Katz | |
| Is the reference standards likely to correctly classify the target condition? | | Unclear |
| Were the reference standard results interpreted without knowledge of the results of the index tests? | | Unclear |
| **Could the reference standard, its conduct, or its interpretation have introduced bias?** | | Low risk |

| **B. Concerns regarding applicability** | |
| --- | --- |
| **Are there concerns that the target condition as defined by the reference standard does not match the question?** | Low concern |

##### Flow and Timing

| **A. Risk of Bias** | | |
| --- | --- | --- |
| Flow and timing |  | |
| Was there an appropriate interval between index test and reference standard? | | Yes |
| Did all patients receive the same reference standard? | | Yes |
| Were all patients included in the analysis? | | Unclear |
| **Could the patient flow have introduced bias?** | | Low risk |

##### Notes

| **Notes** |
| --- |

#### Birrie 1995 (HPA)

##### Patient Selection

| **A. Risk of Bias** | | |
| --- | --- | --- |
| Patient Sampling | cross-sectional study ; consecutive sampling | |
| Was a consecutive or random sample of patients enrolled? | | Yes |
| Was a case-control design avoided? | | Yes |
| Did the study avoid inappropriate exclusions? | | Unclear |
| **Could the selection of patients have introduced bias?** | | Low risk |

| **B. Concerns regarding applicability** | | |
| --- | --- | --- |
| Patient characteristics and setting | - 224 resdients from High Prevalence Area  - Country: Ethiopia  - Setting: field study | |
| **Are there concerns that the included patients and setting do not match the review question?** | | Low concern |

##### Index Test

| Index tests | micro-haematuria by urine reagent strip (Multistix) |
| --- | --- |

##### All tests

| **A. Risk of Bias** | |
| --- | --- |
| Were the index test results interpreted without knowledge of the results of the reference standard? | Unclear |
| If a threshold was used, was it pre-specified? | No |
| **Could the conduct or interpretation of the index test have introduced bias?** | Low risk |

| **B. Concerns regarding applicability** | |
| --- | --- |
| **Are there concerns that the index test, its conduct, or interpretation differ from the review question?** | Low concern |

##### Reference Standard

| **A. Risk of Bias** | | |
| --- | --- | --- |
| Target condition and reference standard(s) | S. haematobium  urine microscopy | |
| Is the reference standards likely to correctly classify the target condition? | | Unclear |
| Were the reference standard results interpreted without knowledge of the results of the index tests? | | Unclear |
| **Could the reference standard, its conduct, or its interpretation have introduced bias?** | | Low risk |

| **B. Concerns regarding applicability** | |
| --- | --- |
| **Are there concerns that the target condition as defined by the reference standard does not match the question?** | Low concern |

##### Flow and Timing

| **A. Risk of Bias** | | |
| --- | --- | --- |
| Flow and timing |  | |
| Was there an appropriate interval between index test and reference standard? | | Unclear |
| Did all patients receive the same reference standard? | | Yes |
| Were all patients included in the analysis? | | Unclear |
| **Could the patient flow have introduced bias?** | | Low risk |

##### Notes

| **Notes** |
| --- |

#### Birrie 1995 (LPA)

##### Patient Selection

| **A. Risk of Bias** | | |
| --- | --- | --- |
| Patient Sampling | cross-sectional study ; consecutive sampling | |
| Was a consecutive or random sample of patients enrolled? | | Yes |
| Was a case-control design avoided? | | Yes |
| Did the study avoid inappropriate exclusions? | | Unclear |
| **Could the selection of patients have introduced bias?** | | Low risk |

| **B. Concerns regarding applicability** | | |
| --- | --- | --- |
| Patient characteristics and setting | - 156 resdients from Low Prevalence Area  - Country: Ethiopia  - Setting: field study | |
| **Are there concerns that the included patients and setting do not match the review question?** | | Low concern |

##### Index Test

| Index tests | micro-haematuria by urine reagent strip (Multistix) |
| --- | --- |

##### All tests

| **A. Risk of Bias** | |
| --- | --- |
| Were the index test results interpreted without knowledge of the results of the reference standard? | Unclear |
| If a threshold was used, was it pre-specified? | No |
| **Could the conduct or interpretation of the index test have introduced bias?** | Low risk |

| **B. Concerns regarding applicability** | |
| --- | --- |
| **Are there concerns that the index test, its conduct, or interpretation differ from the review question?** | Low concern |

##### Reference Standard

| **A. Risk of Bias** | | |
| --- | --- | --- |
| Target condition and reference standard(s) | S. haematobium  urine microscopy | |
| Is the reference standards likely to correctly classify the target condition? | | Unclear |
| Were the reference standard results interpreted without knowledge of the results of the index tests? | | Unclear |
| **Could the reference standard, its conduct, or its interpretation have introduced bias?** | | Low risk |

| **B. Concerns regarding applicability** | |
| --- | --- |
| **Are there concerns that the target condition as defined by the reference standard does not match the question?** | Low concern |

##### Flow and Timing

| **A. Risk of Bias** | | |
| --- | --- | --- |
| Flow and timing |  | |
| Was there an appropriate interval between index test and reference standard? | | Unclear |
| Did all patients receive the same reference standard? | | Yes |
| Were all patients included in the analysis? | | Unclear |
| **Could the patient flow have introduced bias?** | | Low risk |

##### Notes

| **Notes** |
| --- |

#### Birrie 1995 (MPA)

##### Patient Selection

| **A. Risk of Bias** | | |
| --- | --- | --- |
| Patient Sampling | cross-sectional study ; consecutive sampling | |
| Was a consecutive or random sample of patients enrolled? | | Yes |
| Was a case-control design avoided? | | Yes |
| Did the study avoid inappropriate exclusions? | | Unclear |
| **Could the selection of patients have introduced bias?** | | Low risk |

| **B. Concerns regarding applicability** | | |
| --- | --- | --- |
| Patient characteristics and setting | - 121 resdients from Moderate Prevalence Area  - Country: Ethiopia  - Setting: field study | |
| **Are there concerns that the included patients and setting do not match the review question?** | | Low concern |

##### Index Test

| Index tests | micro-haematuria by urine reagent strip (Multistix) |
| --- | --- |

##### All tests

| **A. Risk of Bias** | |
| --- | --- |
| Were the index test results interpreted without knowledge of the results of the reference standard? | Unclear |
| If a threshold was used, was it pre-specified? | No |
| **Could the conduct or interpretation of the index test have introduced bias?** | Low risk |

| **B. Concerns regarding applicability** | |
| --- | --- |
| **Are there concerns that the index test, its conduct, or interpretation differ from the review question?** | Low concern |

##### Reference Standard

| **A. Risk of Bias** | | |
| --- | --- | --- |
| Target condition and reference standard(s) | S. haematobium  urine microscopy | |
| Is the reference standards likely to correctly classify the target condition? | | Unclear |
| Were the reference standard results interpreted without knowledge of the results of the index tests? | | Unclear |
| **Could the reference standard, its conduct, or its interpretation have introduced bias?** | | Low risk |

| **B. Concerns regarding applicability** | |
| --- | --- |
| **Are there concerns that the target condition as defined by the reference standard does not match the question?** | Low concern |

##### Flow and Timing

| **A. Risk of Bias** | | |
| --- | --- | --- |
| Flow and timing |  | |
| Was there an appropriate interval between index test and reference standard? | | Unclear |
| Did all patients receive the same reference standard? | | Yes |
| Were all patients included in the analysis? | | Unclear |
| **Could the patient flow have introduced bias?** | | Low risk |

##### Notes

| **Notes** |
| --- |

#### Bocanegra 2015

##### Patient Selection

| **A. Risk of Bias** | | |
| --- | --- | --- |
| Patient Sampling | cross-sectional study; random selection one of consecutive pairs of children, in alphabetical order | |
| Was a consecutive or random sample of patients enrolled? | | Unclear |
| Was a case-control design avoided? | | Yes |
| Did the study avoid inappropriate exclusions? | | Yes |
| **Could the selection of patients have introduced bias?** | | Unclear risk |

| **B. Concerns regarding applicability** | | |
| --- | --- | --- |
| Patient characteristics and setting | Species: S. haematobium  Country: Angola  Sample size: 1425  Age range: 15y<  Participants: schoolchildren  Setting: field study  Drug before the study: 65 children (4.5%) reported receiving treatment with praziquantel in the two previous months and 22 (1.5%) with mebendazole or albendazole | |
| **Are there concerns that the included patients and setting do not match the review question?** | | Low concern |

##### Index Test

| Index tests | Haematuria colorimetric test; Haematuria aurine dipstick test (Combi-Screen 11SYS); Haematuria visual examination of urine |
| --- | --- |

##### All tests

| **A. Risk of Bias** | |
| --- | --- |
| Were the index test results interpreted without knowledge of the results of the reference standard? | Yes |
| If a threshold was used, was it pre-specified? | Yes |
| **Could the conduct or interpretation of the index test have introduced bias?** | Low risk |

| **B. Concerns regarding applicability** | |
| --- | --- |
| **Are there concerns that the index test, its conduct, or interpretation differ from the review question?** | Low concern |

##### Reference Standard

| **A. Risk of Bias** | | |
| --- | --- | --- |
| Target condition and reference standard(s) | S haematobium measured by Urine microscopy (filtration method) | |
| Is the reference standards likely to correctly classify the target condition? | | Yes |
| Were the reference standard results interpreted without knowledge of the results of the index tests? | | Yes |
| **Could the reference standard, its conduct, or its interpretation have introduced bias?** | | Low risk |

| **B. Concerns regarding applicability** | |
| --- | --- |
| **Are there concerns that the target condition as defined by the reference standard does not match the question?** | Low concern |

##### Flow and Timing

| **A. Risk of Bias** | | |
| --- | --- | --- |
| Flow and timing |  | |
| Was there an appropriate interval between index test and reference standard? | | Yes |
| Did all patients receive the same reference standard? | | Yes |
| Were all patients included in the analysis? | | Yes |
| **Could the patient flow have introduced bias?** | | Low risk |

##### Notes

| **Notes** |
| --- |

#### Bogoch 2012

##### Patient Selection

| **A. Risk of Bias** | | |
| --- | --- | --- |
| Patient Sampling | cross-sectional survey; | |
| Was a consecutive or random sample of patients enrolled? | | Unclear |
| Was a case-control design avoided? | | Yes |
| Did the study avoid inappropriate exclusions? | | Yes |
| **Could the selection of patients have introduced bias?** | | Unclear risk |

| **B. Concerns regarding applicability** | | |
| --- | --- | --- |
| Patient characteristics and setting | Species: S. haematobium  Country: Ghana  Sample size: 280  Age range: 1–77 years  Participants: district population  Setting: field study  Praziquantel before the study: Empiric therapy with albendazole and ivermectin 4 months prior to the study. | |
| **Are there concerns that the included patients and setting do not match the review question?** | | Low concern |

##### Index Test

| Index tests | modified short questionnaire, in combination with urine reagent strips |
| --- | --- |

##### All tests

| **A. Risk of Bias** | |
| --- | --- |
| Were the index test results interpreted without knowledge of the results of the reference standard? | Yes |
| If a threshold was used, was it pre-specified? | Yes |
| **Could the conduct or interpretation of the index test have introduced bias?** | Low risk |

| **B. Concerns regarding applicability** | |
| --- | --- |
| **Are there concerns that the index test, its conduct, or interpretation differ from the review question?** | Low concern |

##### Reference Standard

| **A. Risk of Bias** | | |
| --- | --- | --- |
| Target condition and reference standard(s) | S haematobium measured by Urine microscopy (filtration method) | |
| Is the reference standards likely to correctly classify the target condition? | | Yes |
| Were the reference standard results interpreted without knowledge of the results of the index tests? | | Yes |
| **Could the reference standard, its conduct, or its interpretation have introduced bias?** | | Low risk |

| **B. Concerns regarding applicability** | |
| --- | --- |
| **Are there concerns that the target condition as defined by the reference standard does not match the question?** | Low concern |

##### Flow and Timing

| **A. Risk of Bias** | | |
| --- | --- | --- |
| Flow and timing |  | |
| Was there an appropriate interval between index test and reference standard? | | Yes |
| Did all patients receive the same reference standard? | | Yes |
| Were all patients included in the analysis? | | Yes |
| **Could the patient flow have introduced bias?** | | Low risk |

##### Notes

| **Notes** |
| --- |

#### Bosompem 1996

##### Patient Selection

| **A. Risk of Bias** | | |
| --- | --- | --- |
| Patient Sampling | cross-sectional study | |
| Was a consecutive or random sample of patients enrolled? | | Unclear |
| Was a case-control design avoided? | | Yes |
| Did the study avoid inappropriate exclusions? | | Unclear |
| **Could the selection of patients have introduced bias?** | | Low risk |

| **B. Concerns regarding applicability** | | |
| --- | --- | --- |
| Patient characteristics and setting | - 229 volunteers aged between 1 and 86 years (114 males, 115 females)  - Country: Ghana  - Setting: field study | |
| **Are there concerns that the included patients and setting do not match the review question?** | | Low concern |

##### Index Test

| Index tests | micro-haematuria & proteinuria by reagent strip |
| --- | --- |

##### All tests

| **A. Risk of Bias** | |
| --- | --- |
| Were the index test results interpreted without knowledge of the results of the reference standard? | Unclear |
| If a threshold was used, was it pre-specified? | Unclear |
| **Could the conduct or interpretation of the index test have introduced bias?** | Low risk |

| **B. Concerns regarding applicability** | |
| --- | --- |
| **Are there concerns that the index test, its conduct, or interpretation differ from the review question?** | Low concern |

##### Reference Standard

| **A. Risk of Bias** | | |
| --- | --- | --- |
| Target condition and reference standard(s) | S. haematobium  urine microscopy | |
| Is the reference standards likely to correctly classify the target condition? | | Unclear |
| Were the reference standard results interpreted without knowledge of the results of the index tests? | | Unclear |
| **Could the reference standard, its conduct, or its interpretation have introduced bias?** | | Unclear risk |

| **B. Concerns regarding applicability** | |
| --- | --- |
| **Are there concerns that the target condition as defined by the reference standard does not match the question?** | Low concern |

##### Flow and Timing

| **A. Risk of Bias** | | |
| --- | --- | --- |
| Flow and timing |  | |
| Was there an appropriate interval between index test and reference standard? | | Unclear |
| Did all patients receive the same reference standard? | | Yes |
| Were all patients included in the analysis? | | Yes |
| **Could the patient flow have introduced bias?** | | Low risk |

##### Notes

| **Notes** |
| --- |

#### Bosompem 2004

##### Patient Selection

| **A. Risk of Bias** | | |
| --- | --- | --- |
| Patient Sampling | cross-sectional study | |
| Was a consecutive or random sample of patients enrolled? | | Unclear |
| Was a case-control design avoided? | | Yes |
| Did the study avoid inappropriate exclusions? | | Unclear |
| **Could the selection of patients have introduced bias?** | | Low risk |

| **B. Concerns regarding applicability** | | |
| --- | --- | --- |
| Patient characteristics and setting | - 141 school children (90 with symptoms, 51 asymptomatic)  - Country: Ghana  - Setting: field study | |
| **Are there concerns that the included patients and setting do not match the review question?** | | Low concern |

##### Index Test

| Index tests | proteinuria & micro-haematuria by urine reagent strips (haemacombrix) |
| --- | --- |

##### All tests

| **A. Risk of Bias** | |
| --- | --- |
| Were the index test results interpreted without knowledge of the results of the reference standard? | Unclear |
| If a threshold was used, was it pre-specified? | Unclear |
| **Could the conduct or interpretation of the index test have introduced bias?** | Low risk |

| **B. Concerns regarding applicability** | |
| --- | --- |
| **Are there concerns that the index test, its conduct, or interpretation differ from the review question?** | Low concern |

##### Reference Standard

| **A. Risk of Bias** | | |
| --- | --- | --- |
| Target condition and reference standard(s) | S. haematobium  urine microscopy | |
| Is the reference standards likely to correctly classify the target condition? | | Unclear |
| Were the reference standard results interpreted without knowledge of the results of the index tests? | | Unclear |
| **Could the reference standard, its conduct, or its interpretation have introduced bias?** | | Unclear risk |

| **B. Concerns regarding applicability** | |
| --- | --- |
| **Are there concerns that the target condition as defined by the reference standard does not match the question?** | Low concern |

##### Flow and Timing

| **A. Risk of Bias** | | |
| --- | --- | --- |
| Flow and timing |  | |
| Was there an appropriate interval between index test and reference standard? | | Yes |
| Did all patients receive the same reference standard? | | Yes |
| Were all patients included in the analysis? | | Unclear |
| **Could the patient flow have introduced bias?** | | Low risk |

##### Notes

| **Notes** |
| --- |

#### Bouilhac 1981

##### Patient Selection

| **A. Risk of Bias** | | |
| --- | --- | --- |
| Patient Sampling | unknown | |
| Was a consecutive or random sample of patients enrolled? | | No |
| Was a case-control design avoided? | | Unclear |
| Did the study avoid inappropriate exclusions? | | Unclear |
| **Could the selection of patients have introduced bias?** | | Unclear risk |

| **B. Concerns regarding applicability** | | |
| --- | --- | --- |
| Patient characteristics and setting | - 186 patients (presumably 86 diseased, 100 non-diseased)  - Country: unknown | |
| **Are there concerns that the included patients and setting do not match the review question?** | | Unclear concern |

##### Index Test

| Index tests | SEA-ELISA |
| --- | --- |

##### All tests

| **A. Risk of Bias** | |
| --- | --- |
| Were the index test results interpreted without knowledge of the results of the reference standard? | Unclear |
| If a threshold was used, was it pre-specified? | Yes |
| **Could the conduct or interpretation of the index test have introduced bias?** | Low risk |

| **B. Concerns regarding applicability** | |
| --- | --- |
| **Are there concerns that the index test, its conduct, or interpretation differ from the review question?** | Low concern |

##### Reference Standard

| **A. Risk of Bias** | | |
| --- | --- | --- |
| Target condition and reference standard(s) | S. haematobium  Urine microscopy | |
| Is the reference standards likely to correctly classify the target condition? | | Unclear |
| Were the reference standard results interpreted without knowledge of the results of the index tests? | | Unclear |
| **Could the reference standard, its conduct, or its interpretation have introduced bias?** | | Unclear risk |

| **B. Concerns regarding applicability** | |
| --- | --- |
| **Are there concerns that the target condition as defined by the reference standard does not match the question?** | Low concern |

##### Flow and Timing

| **A. Risk of Bias** | | |
| --- | --- | --- |
| Flow and timing |  | |
| Was there an appropriate interval between index test and reference standard? | | Yes |
| Did all patients receive the same reference standard? | | Yes |
| Were all patients included in the analysis? | | Unclear |
| **Could the patient flow have introduced bias?** | |  |

##### Notes

| **Notes** |
| --- |

#### Chernet 2017

##### Patient Selection

| **A. Risk of Bias** | | |
| --- | --- | --- |
| Patient Sampling | cross-sectional survey; no random selection | |
| Was a consecutive or random sample of patients enrolled? | | Unclear |
| Was a case-control design avoided? | | Yes |
| Did the study avoid inappropriate exclusions? | | Yes |
| **Could the selection of patients have introduced bias?** | | Unclear risk |

| **B. Concerns regarding applicability** | | |
| --- | --- | --- |
| Patient characteristics and setting | Species: S. mansoni  Country: from Eritrea (Bâle)  Sample size: 121  Age range: ≥16 years  Participants: migrants  Setting: field study  Praziquantel before the study: | |
| **Are there concerns that the included patients and setting do not match the review question?** | | Low concern |

##### Index Test

| Index tests | Schistosoma-specific serology by ELISA for Ab against SEA and AWE, and IFAT; immunochromatographic POC-CCA cassette test (Rapid Medical Diagnostics, Pretoria, South Africa) |
| --- | --- |

##### All tests

| **A. Risk of Bias** | |
| --- | --- |
| Were the index test results interpreted without knowledge of the results of the reference standard? | Yes |
| If a threshold was used, was it pre-specified? | Yes |
| **Could the conduct or interpretation of the index test have introduced bias?** | Low risk |

| **B. Concerns regarding applicability** | |
| --- | --- |
| **Are there concerns that the index test, its conduct, or interpretation differ from the review question?** | Low concern |

##### Reference Standard

| **A. Risk of Bias** | | |
| --- | --- | --- |
| Target condition and reference standard(s) | S mansoni measured by KK stool microscopy | |
| Is the reference standards likely to correctly classify the target condition? | | Yes |
| Were the reference standard results interpreted without knowledge of the results of the index tests? | | Yes |
| **Could the reference standard, its conduct, or its interpretation have introduced bias?** | | Low risk |

| **B. Concerns regarding applicability** | |
| --- | --- |
| **Are there concerns that the target condition as defined by the reference standard does not match the question?** | Low concern |

##### Flow and Timing

| **A. Risk of Bias** |
| --- |
| Flow and timing |
| Was there an appropriate interval between index test and reference standard? |
| Did all patients receive the same reference standard? |
| Were all patients included in the analysis? |
| **Could the patient flow have introduced bias?** |

##### Notes

| **Notes** |
| --- |

#### Colley 2013 cameroon

##### Patient Selection

| **A. Risk of Bias** | | |
| --- | --- | --- |
| Patient Sampling | cross-sectional survey; no random selection | |
| Was a consecutive or random sample of patients enrolled? | | Yes |
| Was a case-control design avoided? | | Yes |
| Did the study avoid inappropriate exclusions? | | Yes |
| **Could the selection of patients have introduced bias?** | | Low risk |

| **B. Concerns regarding applicability** | | |
| --- | --- | --- |
| Patient characteristics and setting | Species: S. mansoni  Country: Cameroun  Sample size: 733  Age range: 9-12y  Participants: schoolchildren  Setting: field study  Praziquantel before the study: | |
| **Are there concerns that the included patients and setting do not match the review question?** | | Low concern |

##### Index Test

| Index tests | CCA1 & CCA2 |
| --- | --- |

##### All tests

| **A. Risk of Bias** | |
| --- | --- |
| Were the index test results interpreted without knowledge of the results of the reference standard? | Yes |
| If a threshold was used, was it pre-specified? | Yes |
| **Could the conduct or interpretation of the index test have introduced bias?** | Low risk |

| **B. Concerns regarding applicability** | |
| --- | --- |
| **Are there concerns that the index test, its conduct, or interpretation differ from the review question?** | Low concern |

##### Reference Standard

| **A. Risk of Bias** | | |
| --- | --- | --- |
| Target condition and reference standard(s) | Triplicate Kato Katz smear | |
| Is the reference standards likely to correctly classify the target condition? | | No |
| Were the reference standard results interpreted without knowledge of the results of the index tests? | | Yes |
| **Could the reference standard, its conduct, or its interpretation have introduced bias?** | | Low risk |

| **B. Concerns regarding applicability** | |
| --- | --- |
| **Are there concerns that the target condition as defined by the reference standard does not match the question?** | Low concern |

##### Flow and Timing

| **A. Risk of Bias** | | |
| --- | --- | --- |
| Flow and timing |  | |
| Was there an appropriate interval between index test and reference standard? | | Yes |
| Did all patients receive the same reference standard? | | Yes |
| Were all patients included in the analysis? | | Yes |
| **Could the patient flow have introduced bias?** | | Low risk |

##### Notes

| **Notes** |
| --- |

#### Colley 2013 Côte d'ivoire

##### Patient Selection

| **A. Risk of Bias** | |
| --- | --- |
| Patient Sampling | cross-sectional survey; no random selection |
| Was a consecutive or random sample of patients enrolled? | |
| Was a case-control design avoided? | |
| Did the study avoid inappropriate exclusions? | |
| **Could the selection of patients have introduced bias?** | |

| **B. Concerns regarding applicability** | |
| --- | --- |
| Patient characteristics and setting | Species: S. mansoni  Country: Côte d'Ivoire  Sample size: 607  Age range: 9-12y  Participants: Schoolchildren  Setting: field study  Praziquantel before the study: |
| **Are there concerns that the included patients and setting do not match the review question?** | |

##### Index Test

| Index tests | CCA1 & CCA2 |
| --- | --- |

##### All tests

| **A. Risk of Bias** | |
| --- | --- |
| Were the index test results interpreted without knowledge of the results of the reference standard? | Yes |
| If a threshold was used, was it pre-specified? | Yes |
| **Could the conduct or interpretation of the index test have introduced bias?** | Low risk |

| **B. Concerns regarding applicability** | |
| --- | --- |
| **Are there concerns that the index test, its conduct, or interpretation differ from the review question?** | Low concern |

##### Reference Standard

| **A. Risk of Bias** | | |
| --- | --- | --- |
| Target condition and reference standard(s) | Triplicate Kato Katz smear | |
| Is the reference standards likely to correctly classify the target condition? | | No |
| Were the reference standard results interpreted without knowledge of the results of the index tests? | | Yes |
| **Could the reference standard, its conduct, or its interpretation have introduced bias?** | | Low risk |

| **B. Concerns regarding applicability** | |
| --- | --- |
| **Are there concerns that the target condition as defined by the reference standard does not match the question?** | Low concern |

##### Flow and Timing

| **A. Risk of Bias** | | |
| --- | --- | --- |
| Flow and timing |  | |
| Was there an appropriate interval between index test and reference standard? | | Yes |
| Did all patients receive the same reference standard? | | Yes |
| Were all patients included in the analysis? | | Yes |
| **Could the patient flow have introduced bias?** | | Low risk |

##### Notes

| **Notes** |
| --- |

#### Cooppan 1987

##### Patient Selection

| **A. Risk of Bias** | | |
| --- | --- | --- |
| Patient Sampling | cross-sectional study | |
| Was a consecutive or random sample of patients enrolled? | | Unclear |
| Was a case-control design avoided? | | Yes |
| Did the study avoid inappropriate exclusions? | | Unclear |
| **Could the selection of patients have introduced bias?** | | Low risk |

| **B. Concerns regarding applicability** | | |
| --- | --- | --- |
| Patient characteristics and setting | - 941 school children aged between 4 and 20 years  - Country: South Africa  - Setting: field study | |
| **Are there concerns that the included patients and setting do not match the review question?** | | Low concern |

##### Index Test

| Index tests | micro-haematuria & proteinuria by urine reagent strip (Labstix) |
| --- | --- |

##### All tests

| **A. Risk of Bias** | |
| --- | --- |
| Were the index test results interpreted without knowledge of the results of the reference standard? | Unclear |
| If a threshold was used, was it pre-specified? | Yes |
| **Could the conduct or interpretation of the index test have introduced bias?** | Low risk |

| **B. Concerns regarding applicability** | |
| --- | --- |
| **Are there concerns that the index test, its conduct, or interpretation differ from the review question?** | Low concern |

##### Reference Standard

| **A. Risk of Bias** | | |
| --- | --- | --- |
| Target condition and reference standard(s) | S. haematobium  urine microscopy | |
| Is the reference standards likely to correctly classify the target condition? | | Unclear |
| Were the reference standard results interpreted without knowledge of the results of the index tests? | | Unclear |
| **Could the reference standard, its conduct, or its interpretation have introduced bias?** | | Unclear risk |

| **B. Concerns regarding applicability** | |
| --- | --- |
| **Are there concerns that the target condition as defined by the reference standard does not match the question?** | Low concern |

##### Flow and Timing

| **A. Risk of Bias** | | |
| --- | --- | --- |
| Flow and timing |  | |
| Was there an appropriate interval between index test and reference standard? | | Unclear |
| Did all patients receive the same reference standard? | | Yes |
| Were all patients included in the analysis? | | Yes |
| **Could the patient flow have introduced bias?** | | Low risk |

##### Notes

| **Notes** |
| --- |

#### Coulibaly 2013

##### Patient Selection

| **A. Risk of Bias** | | |
| --- | --- | --- |
| Patient Sampling | cross-sectional survey; no random selection | |
| Was a consecutive or random sample of patients enrolled? | | Yes |
| Was a case-control design avoided? | | Yes |
| Did the study avoid inappropriate exclusions? | | Yes |
| **Could the selection of patients have introduced bias?** | | Low risk |

| **B. Concerns regarding applicability** | | |
| --- | --- | --- |
| Patient characteristics and setting | Species: S. mansoni and S. haematobium  Country: Côte d'Ivoire  Sample size: 140  Age range: <6y  Participants: Villagers  Setting: field study  Praziquantel before the study:No recent schistosomiasis treatment within the past 6 month | |
| **Are there concerns that the included patients and setting do not match the review question?** | | Low concern |

##### Index Test

| Index tests | CCA1, SmCTF |
| --- | --- |

##### All tests

| **A. Risk of Bias** | |
| --- | --- |
| Were the index test results interpreted without knowledge of the results of the reference standard? | No |
| If a threshold was used, was it pre-specified? | Yes |
| **Could the conduct or interpretation of the index test have introduced bias?** | Low risk |

| **B. Concerns regarding applicability** | |
| --- | --- |
| **Are there concerns that the index test, its conduct, or interpretation differ from the review question?** | Low concern |

##### Reference Standard

| **A. Risk of Bias** | | |
| --- | --- | --- |
| Target condition and reference standard(s) | Quadriplicate Kato Katz smear | |
| Is the reference standards likely to correctly classify the target condition? | | No |
| Were the reference standard results interpreted without knowledge of the results of the index tests? | | Yes |
| **Could the reference standard, its conduct, or its interpretation have introduced bias?** | | Low risk |

| **B. Concerns regarding applicability** | |
| --- | --- |
| **Are there concerns that the target condition as defined by the reference standard does not match the question?** | Low concern |

##### Flow and Timing

| **A. Risk of Bias** | | |
| --- | --- | --- |
| Flow and timing |  | |
| Was there an appropriate interval between index test and reference standard? | | Yes |
| Did all patients receive the same reference standard? | | Yes |
| Were all patients included in the analysis? | | Yes |
| **Could the patient flow have introduced bias?** | | Low risk |

##### Notes

| **Notes** |
| --- |

#### Coulibaly 2016

##### Patient Selection

| **A. Risk of Bias** | | |
| --- | --- | --- |
| Patient Sampling | cross-sectional survey; no random selection | |
| Was a consecutive or random sample of patients enrolled? | | Yes |
| Was a case-control design avoided? | | Yes |
| Did the study avoid inappropriate exclusions? | | Yes |
| **Could the selection of patients have introduced bias?** | | Low risk |

| **B. Concerns regarding applicability** | | |
| --- | --- | --- |
| Patient characteristics and setting | Species: S. mansoni  Country: Côte d'Ivoire  Sample size: 149  Age range: 5-16y  Participants: Villagers  Setting: field study  Praziquantel before the study: | |
| **Are there concerns that the included patients and setting do not match the review question?** | | Low concern |

##### Index Test

| Index tests | Mini FLOTAC |
| --- | --- |

##### All tests

| **A. Risk of Bias** | |
| --- | --- |
| Were the index test results interpreted without knowledge of the results of the reference standard? | Unclear |
| If a threshold was used, was it pre-specified? | Yes |
| **Could the conduct or interpretation of the index test have introduced bias?** | Unclear risk |

| **B. Concerns regarding applicability** | |
| --- | --- |
| **Are there concerns that the index test, its conduct, or interpretation differ from the review question?** | Low concern |

##### Reference Standard

| **A. Risk of Bias** | | |
| --- | --- | --- |
| Target condition and reference standard(s) | Quadriplicate Kato Katz smear | |
| Is the reference standards likely to correctly classify the target condition? | | No |
| Were the reference standard results interpreted without knowledge of the results of the index tests? | | Yes |
| **Could the reference standard, its conduct, or its interpretation have introduced bias?** | | Low risk |

| **B. Concerns regarding applicability** | |
| --- | --- |
| **Are there concerns that the target condition as defined by the reference standard does not match the question?** | Low concern |

##### Flow and Timing

| **A. Risk of Bias** | | |
| --- | --- | --- |
| Flow and timing |  | |
| Was there an appropriate interval between index test and reference standard? | | Yes |
| Did all patients receive the same reference standard? | | Yes |
| Were all patients included in the analysis? | | Yes |
| **Could the patient flow have introduced bias?** | | Low risk |

##### Notes

| **Notes** |
| --- |

#### Dawson 2013

##### Patient Selection

| **A. Risk of Bias** | | |
| --- | --- | --- |
| Patient Sampling | cross-sectional survey; no random selection | |
| Was a consecutive or random sample of patients enrolled? | | Yes |
| Was a case-control design avoided? | | No |
| Did the study avoid inappropriate exclusions? | | Yes |
| **Could the selection of patients have introduced bias?** | | Low risk |

| **B. Concerns regarding applicability** | | |
| --- | --- | --- |
| Patient characteristics and setting | Species: S. haematobium  Country: Uganda  Sample size: 46 + 42  Age range: < 3 years & 3 - 5 years  Participants: pre school-aged children  Setting: field study  Praziquantel before the study: no history of previous treatment of schistosomiasis | |
| **Are there concerns that the included patients and setting do not match the review question?** | | Low concern |

##### Index Test

| Index tests | POC-CCA; SmCTF-RDT; SEA-ELISA, |
| --- | --- |

##### All tests

| **A. Risk of Bias** | |
| --- | --- |
| Were the index test results interpreted without knowledge of the results of the reference standard? | Unclear |
| If a threshold was used, was it pre-specified? | Yes |
| **Could the conduct or interpretation of the index test have introduced bias?** | Unclear risk |

| **B. Concerns regarding applicability** | |
| --- | --- |
| **Are there concerns that the index test, its conduct, or interpretation differ from the review question?** | Low concern |

##### Reference Standard

| **A. Risk of Bias** | | |
| --- | --- | --- |
| Target condition and reference standard(s) | Quadriplicate Kato-Katz thick smears | |
| Is the reference standards likely to correctly classify the target condition? | | Yes |
| Were the reference standard results interpreted without knowledge of the results of the index tests? | | Unclear |
| **Could the reference standard, its conduct, or its interpretation have introduced bias?** | | Low risk |

| **B. Concerns regarding applicability** | |
| --- | --- |
| **Are there concerns that the target condition as defined by the reference standard does not match the question?** | Low concern |

##### Flow and Timing

| **A. Risk of Bias** | | |
| --- | --- | --- |
| Flow and timing |  | |
| Was there an appropriate interval between index test and reference standard? | | No |
| Did all patients receive the same reference standard? | | Yes |
| Were all patients included in the analysis? | | Yes |
| **Could the patient flow have introduced bias?** | | Low risk |

##### Notes

| **Notes** |
| --- |

#### De Clercq 1995

##### Patient Selection

| **A. Risk of Bias** | | |
| --- | --- | --- |
| Patient Sampling | cross-sectional study ; consecutive sampling | |
| Was a consecutive or random sample of patients enrolled? | | Yes |
| Was a case-control design avoided? | | Yes |
| Did the study avoid inappropriate exclusions? | | Unclear |
| **Could the selection of patients have introduced bias?** | | Low risk |

| **B. Concerns regarding applicability** | | |
| --- | --- | --- |
| Patient characteristics and setting | - 441 participants from 2 villages  - Country: Mali  - Setting: field study  - No previous praziquantel treatment | |
| **Are there concerns that the included patients and setting do not match the review question?** | | Low concern |

##### Index Test

| Index tests | CAA ELISA Serum |
| --- | --- |

##### All tests

| **A. Risk of Bias** | |
| --- | --- |
| Were the index test results interpreted without knowledge of the results of the reference standard? | Unclear |
| If a threshold was used, was it pre-specified? | Unclear |
| **Could the conduct or interpretation of the index test have introduced bias?** | Low risk |

| **B. Concerns regarding applicability** | |
| --- | --- |
| **Are there concerns that the index test, its conduct, or interpretation differ from the review question?** | Low concern |

##### Reference Standard

| **A. Risk of Bias** | | |
| --- | --- | --- |
| Target condition and reference standard(s) | S. haematobium  urine microscopy | |
| Is the reference standards likely to correctly classify the target condition? | | Unclear |
| Were the reference standard results interpreted without knowledge of the results of the index tests? | | Yes |
| **Could the reference standard, its conduct, or its interpretation have introduced bias?** | | Unclear risk |

| **B. Concerns regarding applicability** | |
| --- | --- |
| **Are there concerns that the target condition as defined by the reference standard does not match the question?** | Low concern |

##### Flow and Timing

| **A. Risk of Bias** | | |
| --- | --- | --- |
| Flow and timing |  | |
| Was there an appropriate interval between index test and reference standard? | | Yes |
| Did all patients receive the same reference standard? | | Yes |
| Were all patients included in the analysis? | | Yes |
| **Could the patient flow have introduced bias?** | | Low risk |

##### Notes

| **Notes** |
| --- |

#### De Oliveira 2005

##### Patient Selection

| **A. Risk of Bias** | |
| --- | --- |
| Patient Sampling | Serum bank & cross-sectional study; no random selection |
| Was a consecutive or random sample of patients enrolled? | |
| Was a case-control design avoided? | |
| Did the study avoid inappropriate exclusions? | |
| **Could the selection of patients have introduced bias?** | |

| **B. Concerns regarding applicability** | |
| --- | --- |
| Patient characteristics and setting | Species: S. mansoni  Country:Brazil  Sample size: 50 pts + 50 Healthy individuals + 37 participants from a non endemic area  Age range:  Participants:  Setting:  Praziquantel before the study: |
| **Are there concerns that the included patients and setting do not match the review question?** | |

##### Index Test

| Index tests | IgM-ELISA - TCA; IgG-ELISA; IgG-IFT; IgM-IFT |
| --- | --- |

##### All tests

| **A. Risk of Bias** | |
| --- | --- |
| Were the index test results interpreted without knowledge of the results of the reference standard? | Unclear |
| If a threshold was used, was it pre-specified? | Yes |
| **Could the conduct or interpretation of the index test have introduced bias?** | Unclear risk |

| **B. Concerns regarding applicability** | |
| --- | --- |
| **Are there concerns that the index test, its conduct, or interpretation differ from the review question?** | Low concern |

##### Reference Standard

| **A. Risk of Bias** | | |
| --- | --- | --- |
| Target condition and reference standard(s) | triplicate kato katz thick smears | |
| Is the reference standards likely to correctly classify the target condition? | | No |
| Were the reference standard results interpreted without knowledge of the results of the index tests? | | Yes |
| **Could the reference standard, its conduct, or its interpretation have introduced bias?** | | Low risk |

| **B. Concerns regarding applicability** | |
| --- | --- |
| **Are there concerns that the target condition as defined by the reference standard does not match the question?** | Low concern |

##### Flow and Timing

| **A. Risk of Bias** | | |
| --- | --- | --- |
| Flow and timing |  | |
| Was there an appropriate interval between index test and reference standard? | | Unclear |
| Did all patients receive the same reference standard? | | Yes |
| Were all patients included in the analysis? | | Yes |
| **Could the patient flow have introduced bias?** | | Low risk |

##### Notes

| **Notes** |
| --- |

#### Elbasheir 2020

##### Patient Selection

| **A. Risk of Bias** | | |
| --- | --- | --- |
| Patient Sampling | longitudinal survey ; using non-probability convenience sampling | |
| Was a consecutive or random sample of patients enrolled? | | Yes |
| Was a case-control design avoided? | | Yes |
| Did the study avoid inappropriate exclusions? | | Unclear |
| **Could the selection of patients have introduced bias?** | | Low risk |

| **B. Concerns regarding applicability** | | |
| --- | --- | --- |
| Patient characteristics and setting | - 489 school children (291 male, 198 female) from 4 elementary schools aged between 5 and 15 years  - Period: February 2017 - March 2018  - Country: Central Sudan  - Setting: field study | |
| **Are there concerns that the included patients and setting do not match the review question?** | | Low concern |

##### Index Test

| Index tests | CCA strip test |
| --- | --- |

##### All tests

| **A. Risk of Bias** | |
| --- | --- |
| Were the index test results interpreted without knowledge of the results of the reference standard? | Unclear |
| If a threshold was used, was it pre-specified? | Unclear |
| **Could the conduct or interpretation of the index test have introduced bias?** | Low risk |

| **B. Concerns regarding applicability** | |
| --- | --- |
| **Are there concerns that the index test, its conduct, or interpretation differ from the review question?** | Low concern |

##### Reference Standard

| **A. Risk of Bias** | | |
| --- | --- | --- |
| Target condition and reference standard(s) | S. mansoni  2 Kato-Katz thick smears | |
| Is the reference standards likely to correctly classify the target condition? | | Unclear |
| Were the reference standard results interpreted without knowledge of the results of the index tests? | | Unclear |
| **Could the reference standard, its conduct, or its interpretation have introduced bias?** | | Low risk |

| **B. Concerns regarding applicability** | |
| --- | --- |
| **Are there concerns that the target condition as defined by the reference standard does not match the question?** | Low concern |

##### Flow and Timing

| **A. Risk of Bias** | | |
| --- | --- | --- |
| Flow and timing |  | |
| Was there an appropriate interval between index test and reference standard? | | Yes |
| Did all patients receive the same reference standard? | | Yes |
| Were all patients included in the analysis? | | Unclear |
| **Could the patient flow have introduced bias?** | | Low risk |

##### Notes

| **Notes** |
| --- |

#### El-Morshedy 1996

##### Patient Selection

| **A. Risk of Bias** | | |
| --- | --- | --- |
| Patient Sampling | cross-sectional study ; random sampling | |
| Was a consecutive or random sample of patients enrolled? | | Yes |
| Was a case-control design avoided? | | Yes |
| Did the study avoid inappropriate exclusions? | | No |
| **Could the selection of patients have introduced bias?** | | Unclear risk |

| **B. Concerns regarding applicability** | | |
| --- | --- | --- |
| Patient characteristics and setting | - 257 men from military camp in Alexandria aged between 20 and 25 years  - Country: Egypt  - Setting: military camp | |
| **Are there concerns that the included patients and setting do not match the review question?** | | Low concern |

##### Index Test

| Index tests | CAA ELISA Serum |
| --- | --- |

##### All tests

| **A. Risk of Bias** | |
| --- | --- |
| Were the index test results interpreted without knowledge of the results of the reference standard? | Unclear |
| If a threshold was used, was it pre-specified? | Unclear |
| **Could the conduct or interpretation of the index test have introduced bias?** | Low risk |

| **B. Concerns regarding applicability** | |
| --- | --- |
| **Are there concerns that the index test, its conduct, or interpretation differ from the review question?** | Low concern |

##### Reference Standard

| **A. Risk of Bias** | | |
| --- | --- | --- |
| Target condition and reference standard(s) | S. mansoni  duplicate Kato-Katz | |
| Is the reference standards likely to correctly classify the target condition? | | Unclear |
| Were the reference standard results interpreted without knowledge of the results of the index tests? | | Unclear |
| **Could the reference standard, its conduct, or its interpretation have introduced bias?** | | Low risk |

| **B. Concerns regarding applicability** | |
| --- | --- |
| **Are there concerns that the target condition as defined by the reference standard does not match the question?** | Low concern |

##### Flow and Timing

| **A. Risk of Bias** | | |
| --- | --- | --- |
| Flow and timing |  | |
| Was there an appropriate interval between index test and reference standard? | | Yes |
| Did all patients receive the same reference standard? | | Yes |
| Were all patients included in the analysis? | | Yes |
| **Could the patient flow have introduced bias?** | | Low risk |

##### Notes

| **Notes** |
| --- |

#### El-Sayed 1995

##### Patient Selection

| **A. Risk of Bias** | | |
| --- | --- | --- |
| Patient Sampling | cross-sectional study | |
| Was a consecutive or random sample of patients enrolled? | | Unclear |
| Was a case-control design avoided? | | Yes |
| Did the study avoid inappropriate exclusions? | | Unclear |
| **Could the selection of patients have introduced bias?** | | Low risk |

| **B. Concerns regarding applicability** | | |
| --- | --- | --- |
| Patient characteristics and setting | - 280 permanent settlers aged between 4 and 36 years  - Country: Egypt  - Setting: field study | |
| **Are there concerns that the included patients and setting do not match the review question?** | | Low concern |

##### Index Test

| Index tests | haematuria by urine reagent strip (Chemistrip) |
| --- | --- |

##### All tests

| **A. Risk of Bias** | |
| --- | --- |
| Were the index test results interpreted without knowledge of the results of the reference standard? | Unclear |
| If a threshold was used, was it pre-specified? | Unclear |
| **Could the conduct or interpretation of the index test have introduced bias?** | Low risk |

| **B. Concerns regarding applicability** | |
| --- | --- |
| **Are there concerns that the index test, its conduct, or interpretation differ from the review question?** | Low concern |

##### Reference Standard

| **A. Risk of Bias** | | |
| --- | --- | --- |
| Target condition and reference standard(s) | S. haematobium  urine microscopy | |
| Is the reference standards likely to correctly classify the target condition? | | Unclear |
| Were the reference standard results interpreted without knowledge of the results of the index tests? | | Unclear |
| **Could the reference standard, its conduct, or its interpretation have introduced bias?** | | Low risk |

| **B. Concerns regarding applicability** | |
| --- | --- |
| **Are there concerns that the target condition as defined by the reference standard does not match the question?** | Low concern |

##### Flow and Timing

| **A. Risk of Bias** | | |
| --- | --- | --- |
| Flow and timing |  | |
| Was there an appropriate interval between index test and reference standard? | | Unclear |
| Did all patients receive the same reference standard? | | Yes |
| Were all patients included in the analysis? | | Unclear |
| **Could the patient flow have introduced bias?** | | Low risk |

##### Notes

| **Notes** |
| --- |

#### Eltiro 1992

##### Patient Selection

| **A. Risk of Bias** | | |
| --- | --- | --- |
| Patient Sampling | cross-sectional study | |
| Was a consecutive or random sample of patients enrolled? | | Yes |
| Was a case-control design avoided? | | Unclear |
| Did the study avoid inappropriate exclusions? | | Unclear |
| **Could the selection of patients have introduced bias?** | | Low risk |

| **B. Concerns regarding applicability** | | |
| --- | --- | --- |
| Patient characteristics and setting | - 200 school children from endemic area aged between 5 and 14 years (119 male, 81 female)  - Country: Ethiopia  - Setting: field study | |
| **Are there concerns that the included patients and setting do not match the review question?** | | Low concern |

##### Index Test

| Index tests | IgG SEA-ELISA |
| --- | --- |

##### All tests

| **A. Risk of Bias** | |
| --- | --- |
| Were the index test results interpreted without knowledge of the results of the reference standard? | Unclear |
| If a threshold was used, was it pre-specified? | Unclear |
| **Could the conduct or interpretation of the index test have introduced bias?** | Low risk |

| **B. Concerns regarding applicability** | |
| --- | --- |
| **Are there concerns that the index test, its conduct, or interpretation differ from the review question?** | Low concern |

##### Reference Standard

| **A. Risk of Bias** | | |
| --- | --- | --- |
| Target condition and reference standard(s) | S. mansoni  triplicate Kato-Katz | |
| Is the reference standards likely to correctly classify the target condition? | | Unclear |
| Were the reference standard results interpreted without knowledge of the results of the index tests? | | Unclear |
| **Could the reference standard, its conduct, or its interpretation have introduced bias?** | | Low risk |

| **B. Concerns regarding applicability** | |
| --- | --- |
| **Are there concerns that the target condition as defined by the reference standard does not match the question?** | Low concern |

##### Flow and Timing

| **A. Risk of Bias** | | |
| --- | --- | --- |
| Flow and timing |  | |
| Was there an appropriate interval between index test and reference standard? | | Yes |
| Did all patients receive the same reference standard? | | Yes |
| Were all patients included in the analysis? | | Unclear |
| **Could the patient flow have introduced bias?** | | Low risk |

##### Notes

| **Notes** |
| --- |

#### Eltoum 1992

##### Patient Selection

| **A. Risk of Bias** | | |
| --- | --- | --- |
| Patient Sampling | cross-sectional study ; random sampling | |
| Was a consecutive or random sample of patients enrolled? | | Yes |
| Was a case-control design avoided? | | Yes |
| Did the study avoid inappropriate exclusions? | | Unclear |
| **Could the selection of patients have introduced bias?** | | Low risk |

| **B. Concerns regarding applicability** | | |
| --- | --- | --- |
| Patient characteristics and setting | - 425 symptomatic and asymptomatic patients aged between 3 and 39 years  - Country: Sudan  - Setting: field study | |
| **Are there concerns that the included patients and setting do not match the review question?** | | Low concern |

##### Index Test

| Index tests | micro-haematuria by urine reagent strip test (Ames-Miles) |
| --- | --- |

##### All tests

| **A. Risk of Bias** | |
| --- | --- |
| Were the index test results interpreted without knowledge of the results of the reference standard? | Unclear |
| If a threshold was used, was it pre-specified? | Unclear |
| **Could the conduct or interpretation of the index test have introduced bias?** | Low risk |

| **B. Concerns regarding applicability** | |
| --- | --- |
| **Are there concerns that the index test, its conduct, or interpretation differ from the review question?** | Low concern |

##### Reference Standard

| **A. Risk of Bias** | | |
| --- | --- | --- |
| Target condition and reference standard(s) | S. haematobium  urine microscopy | |
| Is the reference standards likely to correctly classify the target condition? | | Unclear |
| Were the reference standard results interpreted without knowledge of the results of the index tests? | | Unclear |
| **Could the reference standard, its conduct, or its interpretation have introduced bias?** | | Unclear risk |

| **B. Concerns regarding applicability** | |
| --- | --- |
| **Are there concerns that the target condition as defined by the reference standard does not match the question?** | Low concern |

##### Flow and Timing

| **A. Risk of Bias** | | |
| --- | --- | --- |
| Flow and timing |  | |
| Was there an appropriate interval between index test and reference standard? | | Unclear |
| Did all patients receive the same reference standard? | | Yes |
| Were all patients included in the analysis? | | No |
| **Could the patient flow have introduced bias?** | | Low risk |

##### Notes

| **Notes** |
| --- |

#### Espirito-Santo 2015

##### Patient Selection

| **A. Risk of Bias** | | |
| --- | --- | --- |
| Patient Sampling | cross-sectional survey; randomization of individuals within 1 out of 6 households | |
| Was a consecutive or random sample of patients enrolled? | | Yes |
| Was a case-control design avoided? | | Yes |
| Did the study avoid inappropriate exclusions? | | Yes |
| **Could the selection of patients have introduced bias?** | | Low risk |

| **B. Concerns regarding applicability** | | |
| --- | --- | --- |
| Patient characteristics and setting | Species: S. mansoni  Country: Brazil  Sample size: 610  Age range: >5y to 50y and over  Participants: city neighborhood  Setting: field study  Praziquantel before the study: no schistosomiasis treatment since one year | |
| **Are there concerns that the included patients and setting do not match the review question?** | | Low concern |

##### Index Test

| Index tests | ELISA-IgG, ELISA-IgM, ITF-IgM,COPT, qPCR-serum,qPCR-feces,KK, andHH |
| --- | --- |

##### All tests

| **A. Risk of Bias** | |
| --- | --- |
| Were the index test results interpreted without knowledge of the results of the reference standard? | Yes |
| If a threshold was used, was it pre-specified? | Yes |
| **Could the conduct or interpretation of the index test have introduced bias?** | Low risk |

| **B. Concerns regarding applicability** | |
| --- | --- |
| **Are there concerns that the index test, its conduct, or interpretation differ from the review question?** | Low concern |

##### Reference Standard

| **A. Risk of Bias** | | |
| --- | --- | --- |
| Target condition and reference standard(s) | Schistosomiasis, KK | |
| Is the reference standards likely to correctly classify the target condition? | | Yes |
| Were the reference standard results interpreted without knowledge of the results of the index tests? | | Yes |
| **Could the reference standard, its conduct, or its interpretation have introduced bias?** | | Low risk |

| **B. Concerns regarding applicability** | |
| --- | --- |
| **Are there concerns that the target condition as defined by the reference standard does not match the question?** | Low concern |

##### Flow and Timing

| **A. Risk of Bias** | | |
| --- | --- | --- |
| Flow and timing |  | |
| Was there an appropriate interval between index test and reference standard? | | Yes |
| Did all patients receive the same reference standard? | | Yes |
| Were all patients included in the analysis? | | Yes |
| **Could the patient flow have introduced bias?** | | Low risk |

##### Notes

| **Notes** |
| --- |

#### Fatiregun 2005

##### Patient Selection

| **A. Risk of Bias** | | |
| --- | --- | --- |
| Patient Sampling | cross-sectional study | |
| Was a consecutive or random sample of patients enrolled? | | Yes |
| Was a case-control design avoided? | | Yes |
| Did the study avoid inappropriate exclusions? | | Yes |
| **Could the selection of patients have introduced bias?** | | Low risk |

| **B. Concerns regarding applicability** | | |
| --- | --- | --- |
| Patient characteristics and setting | - 592 students of junior classes aged between 11 and 20 years  - Country: Nigeria  Setting: field study | |
| **Are there concerns that the included patients and setting do not match the review question?** | | Low concern |

##### Index Test

| Index tests | micro-haematuria chemical reagent strip |
| --- | --- |

##### All tests

| **A. Risk of Bias** | |
| --- | --- |
| Were the index test results interpreted without knowledge of the results of the reference standard? | Unclear |
| If a threshold was used, was it pre-specified? | Unclear |
| **Could the conduct or interpretation of the index test have introduced bias?** | Low risk |

| **B. Concerns regarding applicability** | |
| --- | --- |
| **Are there concerns that the index test, its conduct, or interpretation differ from the review question?** | Low concern |

##### Reference Standard

| **A. Risk of Bias** | | |
| --- | --- | --- |
| Target condition and reference standard(s) | S. haematobium  urine microscopy | |
| Is the reference standards likely to correctly classify the target condition? | | Unclear |
| Were the reference standard results interpreted without knowledge of the results of the index tests? | | Unclear |
| **Could the reference standard, its conduct, or its interpretation have introduced bias?** | | Unclear risk |

| **B. Concerns regarding applicability** | |
| --- | --- |
| **Are there concerns that the target condition as defined by the reference standard does not match the question?** | Low concern |

##### Flow and Timing

| **A. Risk of Bias** | | |
| --- | --- | --- |
| Flow and timing |  | |
| Was there an appropriate interval between index test and reference standard? | | Yes |
| Did all patients receive the same reference standard? | | Yes |
| Were all patients included in the analysis? | | Yes |
| **Could the patient flow have introduced bias?** | | Low risk |

##### Notes

| **Notes** |
| --- |

#### Fereira 2017

##### Patient Selection

| **A. Risk of Bias** | | |
| --- | --- | --- |
| Patient Sampling | cross-sectional survey; randomization of households | |
| Was a consecutive or random sample of patients enrolled? | | Yes |
| Was a case-control design avoided? | | Yes |
| Did the study avoid inappropriate exclusions? | | Yes |
| **Could the selection of patients have introduced bias?** | | Low risk |

| **B. Concerns regarding applicability** | |
| --- | --- |
| Patient characteristics and setting | Species: S. mansoni  Country: Brazil  Sample size: 300  Age range: 7-76 y  Participants: villagers  Setting: field study  Praziquantel before the study: |
| **Are there concerns that the included patients and setting do not match the review question?** | |

##### Index Test

| Index tests | POC-CCA |
| --- | --- |

##### All tests

| **A. Risk of Bias** | |
| --- | --- |
| Were the index test results interpreted without knowledge of the results of the reference standard? | Yes |
| If a threshold was used, was it pre-specified? | Yes |
| **Could the conduct or interpretation of the index test have introduced bias?** | Low risk |

| **B. Concerns regarding applicability** | |
| --- | --- |
| **Are there concerns that the index test, its conduct, or interpretation differ from the review question?** | Low concern |

##### Reference Standard

| **A. Risk of Bias** | | |
| --- | --- | --- |
| Target condition and reference standard(s) | 1 sample/1 KK slide; 1 sample/2 KK slides; the first and second samples/4 KK slides; and the first, second, and third samples/6 KK slides. | |
| Is the reference standards likely to correctly classify the target condition? | | No |
| Were the reference standard results interpreted without knowledge of the results of the index tests? | | Yes |
| **Could the reference standard, its conduct, or its interpretation have introduced bias?** | | Low risk |

| **B. Concerns regarding applicability** | |
| --- | --- |
| **Are there concerns that the target condition as defined by the reference standard does not match the question?** | Low concern |

##### Flow and Timing

| **A. Risk of Bias** | | |
| --- | --- | --- |
| Flow and timing |  | |
| Was there an appropriate interval between index test and reference standard? | | Yes |
| Did all patients receive the same reference standard? | | Yes |
| Were all patients included in the analysis? | | Yes |
| **Could the patient flow have introduced bias?** | | Low risk |

##### Notes

| **Notes** |
| --- |

#### French 2007

##### Patient Selection

| **A. Risk of Bias** | | |
| --- | --- | --- |
| Patient Sampling | cross-sectional study | |
| Was a consecutive or random sample of patients enrolled? | | Unclear |
| Was a case-control design avoided? | | Yes |
| Did the study avoid inappropriate exclusions? | | Unclear |
| **Could the selection of patients have introduced bias?** | | Low risk |

| **B. Concerns regarding applicability** | | |
| --- | --- | --- |
| Patient characteristics and setting | - 1976 school children from 24 sentinel schools aged between 6 and 19 years  - Country: Tanzania  - Setting: field study | |
| **Are there concerns that the included patients and setting do not match the review question?** | | Low concern |

##### Index Test

| Index tests | haematuria urine reagent strips (Hemastix) |
| --- | --- |

##### All tests

| **A. Risk of Bias** | |
| --- | --- |
| Were the index test results interpreted without knowledge of the results of the reference standard? | Unclear |
| If a threshold was used, was it pre-specified? | Yes |
| **Could the conduct or interpretation of the index test have introduced bias?** | Low risk |

| **B. Concerns regarding applicability** | |
| --- | --- |
| **Are there concerns that the index test, its conduct, or interpretation differ from the review question?** | Low concern |

##### Reference Standard

| **A. Risk of Bias** | | |
| --- | --- | --- |
| Target condition and reference standard(s) | S. haematobium  urine microscopy | |
| Is the reference standards likely to correctly classify the target condition? | | Unclear |
| Were the reference standard results interpreted without knowledge of the results of the index tests? | | Unclear |
| **Could the reference standard, its conduct, or its interpretation have introduced bias?** | | Unclear risk |

| **B. Concerns regarding applicability** | |
| --- | --- |
| **Are there concerns that the target condition as defined by the reference standard does not match the question?** | Low concern |

##### Flow and Timing

| **A. Risk of Bias** | | |
| --- | --- | --- |
| Flow and timing |  | |
| Was there an appropriate interval between index test and reference standard? | | Unclear |
| Did all patients receive the same reference standard? | | Yes |
| Were all patients included in the analysis? | | Unclear |
| **Could the patient flow have introduced bias?** | | Low risk |

##### Notes

| **Notes** |
| --- |

#### Fuss 2018

##### Patient Selection

| **A. Risk of Bias** | | |
| --- | --- | --- |
| Patient Sampling | cross-sectional study | |
| Was a consecutive or random sample of patients enrolled? | | Yes |
| Was a case-control design avoided? | | Yes |
| Did the study avoid inappropriate exclusions? | | Unclear |
| **Could the selection of patients have introduced bias?** | | Low risk |

| **B. Concerns regarding applicability** | | |
| --- | --- | --- |
| Patient characteristics and setting | - 305 school children (297 included in study) from two different schools aged between 7 and 16 years (146 female, 151 male)  - Country: Tanzania  - Setting: field study | |
| **Are there concerns that the included patients and setting do not match the review question?** | | Low concern |

##### Index Test

| Index tests | POC-CCA cassette & real-time PCR |
| --- | --- |

##### All tests

| **A. Risk of Bias** | |
| --- | --- |
| Were the index test results interpreted without knowledge of the results of the reference standard? | Unclear |
| If a threshold was used, was it pre-specified? | Unclear |
| **Could the conduct or interpretation of the index test have introduced bias?** | Low risk |

| **B. Concerns regarding applicability** | |
| --- | --- |
| **Are there concerns that the index test, its conduct, or interpretation differ from the review question?** | Low concern |

##### Reference Standard

| **A. Risk of Bias** | | |
| --- | --- | --- |
| Target condition and reference standard(s) | S. mansoni  duplicate KK | |
| Is the reference standards likely to correctly classify the target condition? | | Unclear |
| Were the reference standard results interpreted without knowledge of the results of the index tests? | | Unclear |
| **Could the reference standard, its conduct, or its interpretation have introduced bias?** | | Low risk |

| **B. Concerns regarding applicability** | |
| --- | --- |
| **Are there concerns that the target condition as defined by the reference standard does not match the question?** | Low concern |

##### Flow and Timing

| **A. Risk of Bias** | | |
| --- | --- | --- |
| Flow and timing |  | |
| Was there an appropriate interval between index test and reference standard? | | Yes |
| Did all patients receive the same reference standard? | | Yes |
| Were all patients included in the analysis? | | No |
| **Could the patient flow have introduced bias?** | | Low risk |

##### Notes

| **Notes** |
| --- |

#### Gabr 2000

##### Patient Selection

| **A. Risk of Bias** | | |
| --- | --- | --- |
| Patient Sampling | cross-sectional study ; multistage random sampling | |
| Was a consecutive or random sample of patients enrolled? | | Yes |
| Was a case-control design avoided? | | Yes |
| Did the study avoid inappropriate exclusions? | | Yes |
| **Could the selection of patients have introduced bias?** | | Low risk |

| **B. Concerns regarding applicability** | | |
| --- | --- | --- |
| Patient characteristics and setting | - 12'134 residents  - Country: Egypt  - Setting: field study | |
| **Are there concerns that the included patients and setting do not match the review question?** | | Low concern |

##### Index Test

| Index tests | micro-haematuria & proteinuria by reagent strips |
| --- | --- |

##### All tests

| **A. Risk of Bias** | |
| --- | --- |
| Were the index test results interpreted without knowledge of the results of the reference standard? | Unclear |
| If a threshold was used, was it pre-specified? | Unclear |
| **Could the conduct or interpretation of the index test have introduced bias?** | Low risk |

| **B. Concerns regarding applicability** | |
| --- | --- |
| **Are there concerns that the index test, its conduct, or interpretation differ from the review question?** | Low concern |

##### Reference Standard

| **A. Risk of Bias** | | |
| --- | --- | --- |
| Target condition and reference standard(s) | S. haematobium  urine microscopy | |
| Is the reference standards likely to correctly classify the target condition? | | Unclear |
| Were the reference standard results interpreted without knowledge of the results of the index tests? | | Unclear |
| **Could the reference standard, its conduct, or its interpretation have introduced bias?** | | Low risk |

| **B. Concerns regarding applicability** | |
| --- | --- |
| **Are there concerns that the target condition as defined by the reference standard does not match the question?** | Low concern |

##### Flow and Timing

| **A. Risk of Bias** | | |
| --- | --- | --- |
| Flow and timing |  | |
| Was there an appropriate interval between index test and reference standard? | | Unclear |
| Did all patients receive the same reference standard? | | Yes |
| Were all patients included in the analysis? | | No |
| **Could the patient flow have introduced bias?** | | Low risk |

##### Notes

| **Notes** |
| --- |

#### Gandasegui 2015

##### Patient Selection

| **A. Risk of Bias** | | |
| --- | --- | --- |
| Patient Sampling | sample analysis; no randomization | |
| Was a consecutive or random sample of patients enrolled? | | No |
| Was a case-control design avoided? | | Yes |
| Did the study avoid inappropriate exclusions? | | Yes |
| **Could the selection of patients have introduced bias?** | | Unclear risk |

| **B. Concerns regarding applicability** | | |
| --- | --- | --- |
| Patient characteristics and setting | Species: S. haematobium  Country: Spain  Sample size: 94  Age range:  Participants: Sub-Saharan immigrants  Setting:public health diagnostic activities in hospital  Praziquantel before the study: | |
| **Are there concerns that the included patients and setting do not match the review question?** | | Unclear concern |

##### Index Test

| Index tests | LAMP |
| --- | --- |

##### All tests

| **A. Risk of Bias** | |
| --- | --- |
| Were the index test results interpreted without knowledge of the results of the reference standard? | Unclear |
| If a threshold was used, was it pre-specified? | Yes |
| **Could the conduct or interpretation of the index test have introduced bias?** | Unclear risk |

| **B. Concerns regarding applicability** | |
| --- | --- |
| **Are there concerns that the index test, its conduct, or interpretation differ from the review question?** | Low concern |

##### Reference Standard

| **A. Risk of Bias** | | |
| --- | --- | --- |
| Target condition and reference standard(s) | Urine microscopy | |
| Is the reference standards likely to correctly classify the target condition? | | No |
| Were the reference standard results interpreted without knowledge of the results of the index tests? | | Yes |
| **Could the reference standard, its conduct, or its interpretation have introduced bias?** | | Low risk |

| **B. Concerns regarding applicability** | |
| --- | --- |
| **Are there concerns that the target condition as defined by the reference standard does not match the question?** | Low concern |

##### Flow and Timing

| **A. Risk of Bias** | | |
| --- | --- | --- |
| Flow and timing |  | |
| Was there an appropriate interval between index test and reference standard? | | Yes |
| Did all patients receive the same reference standard? | | Yes |
| Were all patients included in the analysis? | | Yes |
| **Could the patient flow have introduced bias?** | | Low risk |

##### Notes

| **Notes** |
| --- |

#### Gandasegui 2018

##### Patient Selection

| **A. Risk of Bias** | | |
| --- | --- | --- |
| Patient Sampling | cross-sectional study | |
| Was a consecutive or random sample of patients enrolled? | | Yes |
| Was a case-control design avoided? | | Yes |
| Did the study avoid inappropriate exclusions? | | Unclear |
| **Could the selection of patients have introduced bias?** | | Low risk |

| **B. Concerns regarding applicability** | | |
| --- | --- | --- |
| Patient characteristics and setting | - 252 school-aged children from 10 schools aged between 5 and 14 years  - Country: Angola  - Period: February - July 2015  - Setting: field study | |
| **Are there concerns that the included patients and setting do not match the review question?** | | Low concern |

##### Index Test

| Index tests | LAMP & microhaematuria |
| --- | --- |

##### All tests

| **A. Risk of Bias** | |
| --- | --- |
| Were the index test results interpreted without knowledge of the results of the reference standard? | Yes |
| If a threshold was used, was it pre-specified? | Unclear |
| **Could the conduct or interpretation of the index test have introduced bias?** | Low risk |

| **B. Concerns regarding applicability** | |
| --- | --- |
| **Are there concerns that the index test, its conduct, or interpretation differ from the review question?** | Low concern |

##### Reference Standard

| **A. Risk of Bias** | | |
| --- | --- | --- |
| Target condition and reference standard(s) | S. haematobium  urine microscopy | |
| Is the reference standards likely to correctly classify the target condition? | | Unclear |
| Were the reference standard results interpreted without knowledge of the results of the index tests? | | Yes |
| **Could the reference standard, its conduct, or its interpretation have introduced bias?** | | Low risk |

| **B. Concerns regarding applicability** | |
| --- | --- |
| **Are there concerns that the target condition as defined by the reference standard does not match the question?** | Low concern |

##### Flow and Timing

| **A. Risk of Bias** | | |
| --- | --- | --- |
| Flow and timing |  | |
| Was there an appropriate interval between index test and reference standard? | | Yes |
| Did all patients receive the same reference standard? | | Yes |
| Were all patients included in the analysis? | | No |
| **Could the patient flow have introduced bias?** | | Low risk |

##### Notes

| **Notes** |
| --- |

#### Glintz 2010

##### Patient Selection

| **A. Risk of Bias** | | |
| --- | --- | --- |
| Patient Sampling | cross-sectional survey; no random selection | |
| Was a consecutive or random sample of patients enrolled? | | Yes |
| Was a case-control design avoided? | | Yes |
| Did the study avoid inappropriate exclusions? | | Yes |
| **Could the selection of patients have introduced bias?** | | Low risk |

| **B. Concerns regarding applicability** | | |
| --- | --- | --- |
| Patient characteristics and setting | Species: S. mansoni  Country: Côte d'Ivoire  Sample size: 133  Age range: 6–15 y  Participants: Schoolchildren  Setting: field study  Praziquantel before the study: | |
| **Are there concerns that the included patients and setting do not match the review question?** | | Low concern |

##### Index Test

| Index tests | FLOTAC |
| --- | --- |

##### All tests

| **A. Risk of Bias** | |
| --- | --- |
| Were the index test results interpreted without knowledge of the results of the reference standard? | Yes |
| If a threshold was used, was it pre-specified? | Yes |
| **Could the conduct or interpretation of the index test have introduced bias?** | Low risk |

| **B. Concerns regarding applicability** | |
| --- | --- |
| **Are there concerns that the index test, its conduct, or interpretation differ from the review question?** | Low concern |

##### Reference Standard

| **A. Risk of Bias** | | |
| --- | --- | --- |
| Target condition and reference standard(s) | triplicate kato katz smear | |
| Is the reference standards likely to correctly classify the target condition? | | No |
| Were the reference standard results interpreted without knowledge of the results of the index tests? | | Yes |
| **Could the reference standard, its conduct, or its interpretation have introduced bias?** | | Low risk |

| **B. Concerns regarding applicability** | |
| --- | --- |
| **Are there concerns that the target condition as defined by the reference standard does not match the question?** | Low concern |

##### Flow and Timing

| **A. Risk of Bias** | | |
| --- | --- | --- |
| Flow and timing |  | |
| Was there an appropriate interval between index test and reference standard? | | Yes |
| Did all patients receive the same reference standard? | | Yes |
| Were all patients included in the analysis? | | Yes |
| **Could the patient flow have introduced bias?** | | Low risk |

##### Notes

| **Notes** |
| --- |

#### Gundersen 1996

##### Patient Selection

| **A. Risk of Bias** | | |
| --- | --- | --- |
| Patient Sampling | cross-sectional study ; consecutive sampling | |
| Was a consecutive or random sample of patients enrolled? | | Yes |
| Was a case-control design avoided? | | Yes |
| Did the study avoid inappropriate exclusions? | | Yes |
| **Could the selection of patients have introduced bias?** | | Low risk |

| **B. Concerns regarding applicability** | | |
| --- | --- | --- |
| Patient characteristics and setting | - 260 women of childbearing age (aged between 15 and 47 years)  - Country: Malawi  - Setting: outpatient department of hospital | |
| **Are there concerns that the included patients and setting do not match the review question?** | | Low concern |

##### Index Test

| Index tests | haematuria, proteinuria & leukocyturia by reagent strip |
| --- | --- |

##### All tests

| **A. Risk of Bias** | |
| --- | --- |
| Were the index test results interpreted without knowledge of the results of the reference standard? | Unclear |
| If a threshold was used, was it pre-specified? | Yes |
| **Could the conduct or interpretation of the index test have introduced bias?** | Low risk |

| **B. Concerns regarding applicability** | |
| --- | --- |
| **Are there concerns that the index test, its conduct, or interpretation differ from the review question?** | Low concern |

##### Reference Standard

| **A. Risk of Bias** | | |
| --- | --- | --- |
| Target condition and reference standard(s) | S. haematobium  urine microscopy | |
| Is the reference standards likely to correctly classify the target condition? | | Unclear |
| Were the reference standard results interpreted without knowledge of the results of the index tests? | | Unclear |
| **Could the reference standard, its conduct, or its interpretation have introduced bias?** | | Unclear risk |

| **B. Concerns regarding applicability** | |
| --- | --- |
| **Are there concerns that the target condition as defined by the reference standard does not match the question?** | Low concern |

##### Flow and Timing

| **A. Risk of Bias** | | |
| --- | --- | --- |
| Flow and timing |  | |
| Was there an appropriate interval between index test and reference standard? | | Unclear |
| Did all patients receive the same reference standard? | | Yes |
| Were all patients included in the analysis? | | Yes |
| **Could the patient flow have introduced bias?** | | Low risk |

##### Notes

| **Notes** |
| --- |

#### Hammad 1997

##### Patient Selection

| **A. Risk of Bias** | | |
| --- | --- | --- |
| Patient Sampling | Cross-sectional study, random sampling | |
| Was a consecutive or random sample of patients enrolled? | | Yes |
| Was a case-control design avoided? | | Yes |
| Did the study avoid inappropriate exclusions? | | Yes |
| **Could the selection of patients have introduced bias?** | | Low risk |

| **B. Concerns regarding applicability** | | |
| --- | --- | --- |
| Patient characteristics and setting | - Setting: field study  - 11.970 individuals  - Country: Egypt | |
| **Are there concerns that the included patients and setting do not match the review question?** | | Low concern |

##### Index Test

| Index tests | Haematuria reagent strip, Proteinuria reagent strip |
| --- | --- |

##### All tests

| **A. Risk of Bias** | |
| --- | --- |
| Were the index test results interpreted without knowledge of the results of the reference standard? | Unclear |
| If a threshold was used, was it pre-specified? | Unclear |
| **Could the conduct or interpretation of the index test have introduced bias?** | Low risk |

| **B. Concerns regarding applicability** | |
| --- | --- |
| **Are there concerns that the index test, its conduct, or interpretation differ from the review question?** | Low concern |

##### Reference Standard

| **A. Risk of Bias** | | |
| --- | --- | --- |
| Target condition and reference standard(s) | S. haematobium  Urine microscopy | |
| Is the reference standards likely to correctly classify the target condition? | | Unclear |
| Were the reference standard results interpreted without knowledge of the results of the index tests? | | Unclear |
| **Could the reference standard, its conduct, or its interpretation have introduced bias?** | | Unclear risk |

| **B. Concerns regarding applicability** | |
| --- | --- |
| **Are there concerns that the target condition as defined by the reference standard does not match the question?** | Low concern |

##### Flow and Timing

| **A. Risk of Bias** | | |
| --- | --- | --- |
| Flow and timing |  | |
| Was there an appropriate interval between index test and reference standard? | | Yes |
| Did all patients receive the same reference standard? | | Yes |
| Were all patients included in the analysis? | | No |
| **Could the patient flow have introduced bias?** | | Low risk |

##### Notes

| **Notes** |
| --- |

#### Hammam 2000a

##### Patient Selection

| **A. Risk of Bias** | | |
| --- | --- | --- |
| Patient Sampling | cross-sectional study ; population sample selcted by multistage stratified cluster sampling | |
| Was a consecutive or random sample of patients enrolled? | | Unclear |
| Was a case-control design avoided? | | Yes |
| Did the study avoid inappropriate exclusions? | | Unclear |
| **Could the selection of patients have introduced bias?** | | Low risk |

| **B. Concerns regarding applicability** | | |
| --- | --- | --- |
| Patient characteristics and setting | - 12'327 residents from Qena Governorate  - Country: Egypt  - Setting: field study | |
| **Are there concerns that the included patients and setting do not match the review question?** | | Low concern |

##### Index Test

| Index tests | micro-haematuria & proteinuria by reagent strip |
| --- | --- |

##### All tests

| **A. Risk of Bias** | |
| --- | --- |
| Were the index test results interpreted without knowledge of the results of the reference standard? | Unclear |
| If a threshold was used, was it pre-specified? | Unclear |
| **Could the conduct or interpretation of the index test have introduced bias?** | Low risk |

| **B. Concerns regarding applicability** | |
| --- | --- |
| **Are there concerns that the index test, its conduct, or interpretation differ from the review question?** | Low concern |

##### Reference Standard

| **A. Risk of Bias** | | |
| --- | --- | --- |
| Target condition and reference standard(s) | S. haematobium  urine microscopy | |
| Is the reference standards likely to correctly classify the target condition? | | Unclear |
| Were the reference standard results interpreted without knowledge of the results of the index tests? | | Unclear |
| **Could the reference standard, its conduct, or its interpretation have introduced bias?** | | Low risk |

| **B. Concerns regarding applicability** | |
| --- | --- |
| **Are there concerns that the target condition as defined by the reference standard does not match the question?** | Low concern |

##### Flow and Timing

| **A. Risk of Bias** | | |
| --- | --- | --- |
| Flow and timing |  | |
| Was there an appropriate interval between index test and reference standard? | | Unclear |
| Did all patients receive the same reference standard? | | Yes |
| Were all patients included in the analysis? | | Yes |
| **Could the patient flow have introduced bias?** | | Low risk |

##### Notes

| **Notes** |
| --- |

#### Hammam 2000b

##### Patient Selection

| **A. Risk of Bias** | | |
| --- | --- | --- |
| Patient Sampling | cross-sectional study | |
| Was a consecutive or random sample of patients enrolled? | | Unclear |
| Was a case-control design avoided? | | Yes |
| Did the study avoid inappropriate exclusions? | | Unclear |
| **Could the selection of patients have introduced bias?** | | Low risk |

| **B. Concerns regarding applicability** | | |
| --- | --- | --- |
| Patient characteristics and setting | - 9555 residents from Assiut Governorate  - Country: Egypt  - Setting: field study | |
| **Are there concerns that the included patients and setting do not match the review question?** | | Low concern |

##### Index Test

| Index tests | micro-haematuria & proteinuria by reagent strip |
| --- | --- |

##### All tests

| **A. Risk of Bias** | |
| --- | --- |
| Were the index test results interpreted without knowledge of the results of the reference standard? | Unclear |
| If a threshold was used, was it pre-specified? | Unclear |
| **Could the conduct or interpretation of the index test have introduced bias?** | Low risk |

| **B. Concerns regarding applicability** | |
| --- | --- |
| **Are there concerns that the index test, its conduct, or interpretation differ from the review question?** | Low concern |

##### Reference Standard

| **A. Risk of Bias** | | |
| --- | --- | --- |
| Target condition and reference standard(s) | S. haematobium  urine microscopy | |
| Is the reference standards likely to correctly classify the target condition? | | Unclear |
| Were the reference standard results interpreted without knowledge of the results of the index tests? | | Unclear |
| **Could the reference standard, its conduct, or its interpretation have introduced bias?** | | Low risk |

| **B. Concerns regarding applicability** | |
| --- | --- |
| **Are there concerns that the target condition as defined by the reference standard does not match the question?** | Low concern |

##### Flow and Timing

| **A. Risk of Bias** | | |
| --- | --- | --- |
| Flow and timing |  | |
| Was there an appropriate interval between index test and reference standard? | | Unclear |
| Did all patients receive the same reference standard? | | Unclear |
| Were all patients included in the analysis? | | Unclear |
| **Could the patient flow have introduced bias?** | | Low risk |

##### Notes

| **Notes** |
| --- |

#### Kassim 1989

##### Patient Selection

| **A. Risk of Bias** | | |
| --- | --- | --- |
| Patient Sampling | cross-sectional study | |
| Was a consecutive or random sample of patients enrolled? | | Unclear |
| Was a case-control design avoided? | | Yes |
| Did the study avoid inappropriate exclusions? | | Yes |
| **Could the selection of patients have introduced bias?** | | Low risk |

| **B. Concerns regarding applicability** | | |
| --- | --- | --- |
| Patient characteristics and setting | - 922 school children aged between 5 and 14 years  - Country: Nigeria  - Setting: field study | |
| **Are there concerns that the included patients and setting do not match the review question?** | | Low concern |

##### Index Test

| Index tests | micro-haematuria & proteinuria by urine reagent strip |
| --- | --- |

##### All tests

| **A. Risk of Bias** | |
| --- | --- |
| Were the index test results interpreted without knowledge of the results of the reference standard? | Unclear |
| If a threshold was used, was it pre-specified? | Unclear |
| **Could the conduct or interpretation of the index test have introduced bias?** | Low risk |

| **B. Concerns regarding applicability** | |
| --- | --- |
| **Are there concerns that the index test, its conduct, or interpretation differ from the review question?** | Low concern |

##### Reference Standard

| **A. Risk of Bias** | | |
| --- | --- | --- |
| Target condition and reference standard(s) | S. haematobium  urine microscopy | |
| Is the reference standards likely to correctly classify the target condition? | | Unclear |
| Were the reference standard results interpreted without knowledge of the results of the index tests? | | Unclear |
| **Could the reference standard, its conduct, or its interpretation have introduced bias?** | | Low risk |

| **B. Concerns regarding applicability** | |
| --- | --- |
| **Are there concerns that the target condition as defined by the reference standard does not match the question?** | Low concern |

##### Flow and Timing

| **A. Risk of Bias** | | |
| --- | --- | --- |
| Flow and timing |  | |
| Was there an appropriate interval between index test and reference standard? | | Yes |
| Did all patients receive the same reference standard? | | Yes |
| Were all patients included in the analysis? | | Yes |
| **Could the patient flow have introduced bias?** | | Low risk |

##### Notes

| **Notes** |
| --- |

#### Kiliku 1991

##### Patient Selection

| **A. Risk of Bias** | | |
| --- | --- | --- |
| Patient Sampling | cross-sectional study | |
| Was a consecutive or random sample of patients enrolled? | | Unclear |
| Was a case-control design avoided? | | Yes |
| Did the study avoid inappropriate exclusions? | | Unclear |
| **Could the selection of patients have introduced bias?** | | Unclear risk |

| **B. Concerns regarding applicability** | | |
| --- | --- | --- |
| Patient characteristics and setting | - 426 patients  - Country: Kenya  - Setting: field study | |
| **Are there concerns that the included patients and setting do not match the review question?** | | Low concern |

##### Index Test

| Index tests | Micro-haematuria & Proteinuria by urine reagent strip (Uro-Labstix) |
| --- | --- |

##### All tests

| **A. Risk of Bias** | |
| --- | --- |
| Were the index test results interpreted without knowledge of the results of the reference standard? | Unclear |
| If a threshold was used, was it pre-specified? | No |
| **Could the conduct or interpretation of the index test have introduced bias?** | Low risk |

| **B. Concerns regarding applicability** | |
| --- | --- |
| **Are there concerns that the index test, its conduct, or interpretation differ from the review question?** | Low concern |

##### Reference Standard

| **A. Risk of Bias** | | |
| --- | --- | --- |
| Target condition and reference standard(s) | S. haematobium  Urine Microscopy | |
| Is the reference standards likely to correctly classify the target condition? | | Unclear |
| Were the reference standard results interpreted without knowledge of the results of the index tests? | | Unclear |
| **Could the reference standard, its conduct, or its interpretation have introduced bias?** | | Unclear risk |

| **B. Concerns regarding applicability** | |
| --- | --- |
| **Are there concerns that the target condition as defined by the reference standard does not match the question?** | Low concern |

##### Flow and Timing

| **A. Risk of Bias** | | |
| --- | --- | --- |
| Flow and timing |  | |
| Was there an appropriate interval between index test and reference standard? | | Yes |
| Did all patients receive the same reference standard? | | Unclear |
| Were all patients included in the analysis? | | Yes |
| **Could the patient flow have introduced bias?** | | Low risk |

##### Notes

| **Notes** |
| --- |

#### King 1988a

##### Patient Selection

| **A. Risk of Bias** | | |
| --- | --- | --- |
| Patient Sampling | cross-sectional study | |
| Was a consecutive or random sample of patients enrolled? | | Unclear |
| Was a case-control design avoided? | | Yes |
| Did the study avoid inappropriate exclusions? | | Yes |
| **Could the selection of patients have introduced bias?** | | Low risk |

| **B. Concerns regarding applicability** | | |
| --- | --- | --- |
| Patient characteristics and setting | - 2628 students from 5 local primary and secondary schools aged between 4 and 21 years  - Country: Kenya  - Setting: field study | |
| **Are there concerns that the included patients and setting do not match the review question?** | | Low concern |

##### Index Test

| Index tests | micro-haematuria & proteinuria urine reagent strips (Chemstrip) |
| --- | --- |

##### All tests

| **A. Risk of Bias** | |
| --- | --- |
| Were the index test results interpreted without knowledge of the results of the reference standard? | Unclear |
| If a threshold was used, was it pre-specified? | Unclear |
| **Could the conduct or interpretation of the index test have introduced bias?** | Low risk |

| **B. Concerns regarding applicability** | |
| --- | --- |
| **Are there concerns that the index test, its conduct, or interpretation differ from the review question?** | Low concern |

##### Reference Standard

| **A. Risk of Bias** | | |
| --- | --- | --- |
| Target condition and reference standard(s) | S. haematobium  urine microscopy | |
| Is the reference standards likely to correctly classify the target condition? | | Unclear |
| Were the reference standard results interpreted without knowledge of the results of the index tests? | | Unclear |
| **Could the reference standard, its conduct, or its interpretation have introduced bias?** | | Low risk |

| **B. Concerns regarding applicability** | |
| --- | --- |
| **Are there concerns that the target condition as defined by the reference standard does not match the question?** | Low concern |

##### Flow and Timing

| **A. Risk of Bias** | | |
| --- | --- | --- |
| Flow and timing |  | |
| Was there an appropriate interval between index test and reference standard? | | Yes |
| Did all patients receive the same reference standard? | | Yes |
| Were all patients included in the analysis? | | Yes |
| **Could the patient flow have introduced bias?** | | Low risk |

##### Notes

| **Notes** |
| --- |

#### King 1988b

##### Patient Selection

| **A. Risk of Bias** | | |
| --- | --- | --- |
| Patient Sampling | cross-sectional study | |
| Was a consecutive or random sample of patients enrolled? | | Unclear |
| Was a case-control design avoided? | | Yes |
| Did the study avoid inappropriate exclusions? | | Unclear |
| **Could the selection of patients have introduced bias?** | | Low risk |

| **B. Concerns regarding applicability** | | |
| --- | --- | --- |
| Patient characteristics and setting | - 639 residents of village of all ages  - Country: Kenya  - Setting: field study | |
| **Are there concerns that the included patients and setting do not match the review question?** | | Low concern |

##### Index Test

| Index tests | micro-haematuria by urine reagent strip (Combur) |
| --- | --- |

##### All tests

| **A. Risk of Bias** | |
| --- | --- |
| Were the index test results interpreted without knowledge of the results of the reference standard? | Unclear |
| If a threshold was used, was it pre-specified? | Unclear |
| **Could the conduct or interpretation of the index test have introduced bias?** | Low risk |

| **B. Concerns regarding applicability** | |
| --- | --- |
| **Are there concerns that the index test, its conduct, or interpretation differ from the review question?** | Low concern |

##### Reference Standard

| **A. Risk of Bias** | | |
| --- | --- | --- |
| Target condition and reference standard(s) | S. haematobium  urine microscopy | |
| Is the reference standards likely to correctly classify the target condition? | | Unclear |
| Were the reference standard results interpreted without knowledge of the results of the index tests? | | Unclear |
| **Could the reference standard, its conduct, or its interpretation have introduced bias?** | | Low risk |

| **B. Concerns regarding applicability** | |
| --- | --- |
| **Are there concerns that the target condition as defined by the reference standard does not match the question?** | Low concern |

##### Flow and Timing

| **A. Risk of Bias** | | |
| --- | --- | --- |
| Flow and timing |  | |
| Was there an appropriate interval between index test and reference standard? | | Unclear |
| Did all patients receive the same reference standard? | | Yes |
| Were all patients included in the analysis? | | Yes |
| **Could the patient flow have introduced bias?** | | Low risk |

##### Notes

| **Notes** |
| --- |

#### Kitange 1993

##### Patient Selection

| **A. Risk of Bias** | | |
| --- | --- | --- |
| Patient Sampling | cohort study ; random sampling | |
| Was a consecutive or random sample of patients enrolled? | | Unclear |
| Was a case-control design avoided? | | Yes |
| Did the study avoid inappropriate exclusions? | | Unclear |
| **Could the selection of patients have introduced bias?** | | Unclear risk |

| **B. Concerns regarding applicability** | | |
| --- | --- | --- |
| Patient characteristics and setting | - 253 school children aged between 7 and 19 years (130 male, 123 female)  - Country: Tanzania  - Setting: field study  - Praziquantel treatment: 1 year prior to study | |
| **Are there concerns that the included patients and setting do not match the review question?** | | Low concern |

##### Index Test

| Index tests | Micro-haematuria & Proteinuria by urine reagent strip (BM TEST 5L) |
| --- | --- |

##### All tests

| **A. Risk of Bias** | |
| --- | --- |
| Were the index test results interpreted without knowledge of the results of the reference standard? | Unclear |
| If a threshold was used, was it pre-specified? | No |
| **Could the conduct or interpretation of the index test have introduced bias?** | Low risk |

| **B. Concerns regarding applicability** | |
| --- | --- |
| **Are there concerns that the index test, its conduct, or interpretation differ from the review question?** | Low concern |

##### Reference Standard

| **A. Risk of Bias** | | |
| --- | --- | --- |
| Target condition and reference standard(s) | S. haematobium  Urine microscopy | |
| Is the reference standards likely to correctly classify the target condition? | | Unclear |
| Were the reference standard results interpreted without knowledge of the results of the index tests? | | Yes |
| **Could the reference standard, its conduct, or its interpretation have introduced bias?** | | Low risk |

| **B. Concerns regarding applicability** | |
| --- | --- |
| **Are there concerns that the target condition as defined by the reference standard does not match the question?** | Low concern |

##### Flow and Timing

| **A. Risk of Bias** | | |
| --- | --- | --- |
| Flow and timing |  | |
| Was there an appropriate interval between index test and reference standard? | | Unclear |
| Did all patients receive the same reference standard? | | Unclear |
| Were all patients included in the analysis? | | Unclear |
| **Could the patient flow have introduced bias?** | | Low risk |

##### Notes

| **Notes** |
| --- |

#### Knopp 2015

##### Patient Selection

| **A. Risk of Bias** | | |
| --- | --- | --- |
| Patient Sampling | cross-sectional study | |
| Was a consecutive or random sample of patients enrolled? | | Yes |
| Was a case-control design avoided? | | Yes |
| Did the study avoid inappropriate exclusions? | | Unclear |
| **Could the selection of patients have introduced bias?** | | Low risk |

| **B. Concerns regarding applicability** | | |
| --- | --- | --- |
| Patient characteristics and setting | - 1200 children from 16 primary schools aged between 9 and 12 years  - Country: Tanzania  - Praziquantel treatment in June and November 2013  - Period: 2013  - Setting: field study | |
| **Are there concerns that the included patients and setting do not match the review question?** | | Low concern |

##### Index Test

| Index tests | Reagent strip |
| --- | --- |

##### All tests

| **A. Risk of Bias** | |
| --- | --- |
| Were the index test results interpreted without knowledge of the results of the reference standard? | Yes |
| If a threshold was used, was it pre-specified? | Yes |
| **Could the conduct or interpretation of the index test have introduced bias?** | Low risk |

| **B. Concerns regarding applicability** | |
| --- | --- |
| **Are there concerns that the index test, its conduct, or interpretation differ from the review question?** | Low concern |

##### Reference Standard

| **A. Risk of Bias** | | |
| --- | --- | --- |
| Target condition and reference standard(s) | S. haematobium  Urine filtration (with quality control slide) | |
| Is the reference standards likely to correctly classify the target condition? | | Unclear |
| Were the reference standard results interpreted without knowledge of the results of the index tests? | | Unclear |
| **Could the reference standard, its conduct, or its interpretation have introduced bias?** | | Low risk |

| **B. Concerns regarding applicability** | |
| --- | --- |
| **Are there concerns that the target condition as defined by the reference standard does not match the question?** | Low concern |

##### Flow and Timing

| **A. Risk of Bias** | | |
| --- | --- | --- |
| Flow and timing |  | |
| Was there an appropriate interval between index test and reference standard? | | Yes |
| Did all patients receive the same reference standard? | | Yes |
| Were all patients included in the analysis? | | Unclear |
| **Could the patient flow have introduced bias?** | | Low risk |

##### Notes

| **Notes** |
| --- |

#### Knopp 2018

##### Patient Selection

| **A. Risk of Bias** | | |
| --- | --- | --- |
| Patient Sampling | cross-sectional study | |
| Was a consecutive or random sample of patients enrolled? | | Yes |
| Was a case-control design avoided? | | Unclear |
| Did the study avoid inappropriate exclusions? | | Unclear |
| **Could the selection of patients have introduced bias?** | | Low risk |

| **B. Concerns regarding applicability** | |
| --- | --- |
| Patient characteristics and setting | - 39'207 children aged between 9 and 12 years  - 18'155 adults aged between 20 and 55 years  - Country: Zanzibar  - Setting |
| **Are there concerns that the included patients and setting do not match the review question?** | |

##### Index Test

| Index tests | Micro-haematuria by urine reagent strip (Hemastix) |
| --- | --- |

##### All tests

| **A. Risk of Bias** | |
| --- | --- |
| Were the index test results interpreted without knowledge of the results of the reference standard? | Unclear |
| If a threshold was used, was it pre-specified? | Yes |
| **Could the conduct or interpretation of the index test have introduced bias?** | Low risk |

| **B. Concerns regarding applicability** | |
| --- | --- |
| **Are there concerns that the index test, its conduct, or interpretation differ from the review question?** | Low concern |

##### Reference Standard

| **A. Risk of Bias** | | |
| --- | --- | --- |
| Target condition and reference standard(s) | S. haematobium  Urine microscopy | |
| Is the reference standards likely to correctly classify the target condition? | | Unclear |
| Were the reference standard results interpreted without knowledge of the results of the index tests? | | Unclear |
| **Could the reference standard, its conduct, or its interpretation have introduced bias?** | | Low risk |

| **B. Concerns regarding applicability** | |
| --- | --- |
| **Are there concerns that the target condition as defined by the reference standard does not match the question?** | Low concern |

##### Flow and Timing

| **A. Risk of Bias** | | |
| --- | --- | --- |
| Flow and timing |  | |
| Was there an appropriate interval between index test and reference standard? | | Yes |
| Did all patients receive the same reference standard? | | Yes |
| Were all patients included in the analysis? | | Yes |
| **Could the patient flow have introduced bias?** | | Low risk |

##### Notes

| **Notes** |
| --- |

#### Kosinski 2011

##### Patient Selection

| **A. Risk of Bias** | | |
| --- | --- | --- |
| Patient Sampling | cross-sectional survey; no random selection | |
| Was a consecutive or random sample of patients enrolled? | | Yes |
| Was a case-control design avoided? | | Yes |
| Did the study avoid inappropriate exclusions? | | Yes |
| **Could the selection of patients have introduced bias?** | | Low risk |

| **B. Concerns regarding applicability** | | |
| --- | --- | --- |
| Patient characteristics and setting | Species: S. haematobium  Country: Ghana  Sample size: 255  Age range: 8–18 y  Participants: Schollchildren  Setting: field study  Praziquantel before the study: | |
| **Are there concerns that the included patients and setting do not match the review question?** | | Low concern |

##### Index Test

| Index tests | microhematuria via a semi-quantitative dipstick test; |
| --- | --- |

##### All tests

| **A. Risk of Bias** | |
| --- | --- |
| Were the index test results interpreted without knowledge of the results of the reference standard? | Yes |
| If a threshold was used, was it pre-specified? | Yes |
| **Could the conduct or interpretation of the index test have introduced bias?** | Low risk |

| **B. Concerns regarding applicability** | |
| --- | --- |
| **Are there concerns that the index test, its conduct, or interpretation differ from the review question?** | Low concern |

##### Reference Standard

| **A. Risk of Bias** | | |
| --- | --- | --- |
| Target condition and reference standard(s) | Urine microscopy | |
| Is the reference standards likely to correctly classify the target condition? | | Yes |
| Were the reference standard results interpreted without knowledge of the results of the index tests? | | Yes |
| **Could the reference standard, its conduct, or its interpretation have introduced bias?** | | Low risk |

| **B. Concerns regarding applicability** | |
| --- | --- |
| **Are there concerns that the target condition as defined by the reference standard does not match the question?** | Low concern |

##### Flow and Timing

| **A. Risk of Bias** | | |
| --- | --- | --- |
| Flow and timing |  | |
| Was there an appropriate interval between index test and reference standard? | | Yes |
| Did all patients receive the same reference standard? | | Yes |
| Were all patients included in the analysis? | | Yes |
| **Could the patient flow have introduced bias?** | | Low risk |

##### Notes

| **Notes** |
| --- |

#### Lamberton 2014

##### Patient Selection

| **A. Risk of Bias** | | |
| --- | --- | --- |
| Patient Sampling | cross-sectional study | |
| Was a consecutive or random sample of patients enrolled? | | Yes |
| Was a case-control design avoided? | | Yes |
| Did the study avoid inappropriate exclusions? | | Unclear |
| **Could the selection of patients have introduced bias?** | | Low risk |

| **B. Concerns regarding applicability** | | |
| --- | --- | --- |
| Patient characteristics and setting | - 76 schoolchildren  - Country: Uganda  - Setting: field study | |
| **Are there concerns that the included patients and setting do not match the review question?** | | Low concern |

##### Index Test

| Index tests | CCA cassete |
| --- | --- |

##### All tests

| **A. Risk of Bias** | |
| --- | --- |
| Were the index test results interpreted without knowledge of the results of the reference standard? | Unclear |
| If a threshold was used, was it pre-specified? | Yes |
| **Could the conduct or interpretation of the index test have introduced bias?** | Low risk |

| **B. Concerns regarding applicability** | |
| --- | --- |
| **Are there concerns that the index test, its conduct, or interpretation differ from the review question?** | Low concern |

##### Reference Standard

| **A. Risk of Bias** | | |
| --- | --- | --- |
| Target condition and reference standard(s) | S. mansoni  sextuple KK | |
| Is the reference standards likely to correctly classify the target condition? | | Unclear |
| Were the reference standard results interpreted without knowledge of the results of the index tests? | | Yes |
| **Could the reference standard, its conduct, or its interpretation have introduced bias?** | | Low risk |

| **B. Concerns regarding applicability** | |
| --- | --- |
| **Are there concerns that the target condition as defined by the reference standard does not match the question?** | Low concern |

##### Flow and Timing

| **A. Risk of Bias** | | |
| --- | --- | --- |
| Flow and timing |  | |
| Was there an appropriate interval between index test and reference standard? | | Yes |
| Did all patients receive the same reference standard? | | Yes |
| Were all patients included in the analysis? | | No |
| **Could the patient flow have introduced bias?** | | Low risk |

##### Notes

| **Notes** |
| --- |

#### Legesse 2008

##### Patient Selection

| **A. Risk of Bias** | | |
| --- | --- | --- |
| Patient Sampling | cross-sectional study | |
| Was a consecutive or random sample of patients enrolled? | | Yes |
| Was a case-control design avoided? | | Yes |
| Did the study avoid inappropriate exclusions? | | Yes |
| **Could the selection of patients have introduced bias?** | | Low risk |

| **B. Concerns regarding applicability** | | |
| --- | --- | --- |
| Patient characteristics and setting | - 184 primary school children aged between 5 and 22 years (97 boys, 87 girls)  - Country: Ethiopia  - Period: June 2007 | |
| **Are there concerns that the included patients and setting do not match the review question?** | | Low concern |

##### Index Test

| Index tests | CCA by reagent strip test |
| --- | --- |

##### All tests

| **A. Risk of Bias** | |
| --- | --- |
| Were the index test results interpreted without knowledge of the results of the reference standard? | Unclear |
| If a threshold was used, was it pre-specified? | Yes |
| **Could the conduct or interpretation of the index test have introduced bias?** | Low risk |

| **B. Concerns regarding applicability** | |
| --- | --- |
| **Are there concerns that the index test, its conduct, or interpretation differ from the review question?** | Low concern |

##### Reference Standard

| **A. Risk of Bias** | | |
| --- | --- | --- |
| Target condition and reference standard(s) | S. mansoni  stool microscopy (1 Kato-Katz slide) | |
| Is the reference standards likely to correctly classify the target condition? | | Unclear |
| Were the reference standard results interpreted without knowledge of the results of the index tests? | | Yes |
| **Could the reference standard, its conduct, or its interpretation have introduced bias?** | | Unclear risk |

| **B. Concerns regarding applicability** | |
| --- | --- |
| **Are there concerns that the target condition as defined by the reference standard does not match the question?** | Low concern |

##### Flow and Timing

| **A. Risk of Bias** | | |
| --- | --- | --- |
| Flow and timing |  | |
| Was there an appropriate interval between index test and reference standard? | | Yes |
| Did all patients receive the same reference standard? | | Yes |
| Were all patients included in the analysis? | | Yes |
| **Could the patient flow have introduced bias?** | | Low risk |

##### Notes

| **Notes** |
| --- |

#### Lengeler 1993

##### Patient Selection

| **A. Risk of Bias** | | |
| --- | --- | --- |
| Patient Sampling | cross-sectional study | |
| Was a consecutive or random sample of patients enrolled? | | Unclear |
| Was a case-control design avoided? | | Yes |
| Did the study avoid inappropriate exclusions? | | Unclear |
| **Could the selection of patients have introduced bias?** | | Low risk |

| **B. Concerns regarding applicability** | | |
| --- | --- | --- |
| Patient characteristics and setting | - 1208 school children aged between 11 and 15 years  - Country: Tanzania  - Setting: field study | |
| **Are there concerns that the included patients and setting do not match the review question?** | | Low concern |

##### Index Test

| Index tests | micro-haematuria by urine reagent strip (Combur 9 Multistix) |
| --- | --- |

##### All tests

| **A. Risk of Bias** | |
| --- | --- |
| Were the index test results interpreted without knowledge of the results of the reference standard? | Unclear |
| If a threshold was used, was it pre-specified? | Yes |
| **Could the conduct or interpretation of the index test have introduced bias?** | Low risk |

| **B. Concerns regarding applicability** | |
| --- | --- |
| **Are there concerns that the index test, its conduct, or interpretation differ from the review question?** | Low concern |

##### Reference Standard

| **A. Risk of Bias** | | |
| --- | --- | --- |
| Target condition and reference standard(s) | S. haematobium  urine microscopy | |
| Is the reference standards likely to correctly classify the target condition? | | Unclear |
| Were the reference standard results interpreted without knowledge of the results of the index tests? | | Unclear |
| **Could the reference standard, its conduct, or its interpretation have introduced bias?** | | Unclear risk |

| **B. Concerns regarding applicability** | |
| --- | --- |
| **Are there concerns that the target condition as defined by the reference standard does not match the question?** | Low concern |

##### Flow and Timing

| **A. Risk of Bias** | | |
| --- | --- | --- |
| Flow and timing |  | |
| Was there an appropriate interval between index test and reference standard? | | Yes |
| Did all patients receive the same reference standard? | | Yes |
| Were all patients included in the analysis? | | Yes |
| **Could the patient flow have introduced bias?** | | Low risk |

##### Notes

| **Notes** |
| --- |

#### Lindholz 2018

##### Patient Selection

| **A. Risk of Bias** | | |
| --- | --- | --- |
| Patient Sampling | cross-sectional study ; community -and geographically-based study | |
| Was a consecutive or random sample of patients enrolled? | | Yes |
| Was a case-control design avoided? | | Yes |
| Did the study avoid inappropriate exclusions? | | Yes |
| **Could the selection of patients have introduced bias?** | | Low risk |

| **B. Concerns regarding applicability** | | |
| --- | --- | --- |
| Patient characteristics and setting | - 580 patients (from area with approx. 700 inhabitants) aged between 1 and 17 years  - Country: Brazil  - Period: October-November 2015  - Setting: field study | |
| **Are there concerns that the included patients and setting do not match the review question?** | | Low concern |

##### Index Test

| Index tests | POC-CCA & Helmintex |
| --- | --- |

##### All tests

| **A. Risk of Bias** | |
| --- | --- |
| Were the index test results interpreted without knowledge of the results of the reference standard? | Yes |
| If a threshold was used, was it pre-specified? | Unclear |
| **Could the conduct or interpretation of the index test have introduced bias?** | Low risk |

| **B. Concerns regarding applicability** | |
| --- | --- |
| **Are there concerns that the index test, its conduct, or interpretation differ from the review question?** | Low concern |

##### Reference Standard

| **A. Risk of Bias** | | |
| --- | --- | --- |
| Target condition and reference standard(s) | S. mansoni  duplicate KK | |
| Is the reference standards likely to correctly classify the target condition? | | Unclear |
| Were the reference standard results interpreted without knowledge of the results of the index tests? | | Yes |
| **Could the reference standard, its conduct, or its interpretation have introduced bias?** | | Low risk |

| **B. Concerns regarding applicability** | |
| --- | --- |
| **Are there concerns that the target condition as defined by the reference standard does not match the question?** | Low concern |

##### Flow and Timing

| **A. Risk of Bias** | | |
| --- | --- | --- |
| Flow and timing |  | |
| Was there an appropriate interval between index test and reference standard? | | Yes |
| Did all patients receive the same reference standard? | | Yes |
| Were all patients included in the analysis? | | No |
| **Could the patient flow have introduced bias?** | | Low risk |

##### Notes

| **Notes** |
| --- |

#### Lodh 2013

##### Patient Selection

| **A. Risk of Bias** | | |
| --- | --- | --- |
| Patient Sampling | Cross-sectional survey; random selection | |
| Was a consecutive or random sample of patients enrolled? | | Yes |
| Was a case-control design avoided? | | Yes |
| Did the study avoid inappropriate exclusions? | | Yes |
| **Could the selection of patients have introduced bias?** | | Low risk |

| **B. Concerns regarding applicability** | | |
| --- | --- | --- |
| Patient characteristics and setting | Species: S. mansoni  Country: Zambia  Sample size: 100  Age range: 18–50 y  Participants: resident  Setting: field study  Praziquantel before the study: | |
| **Are there concerns that the included patients and setting do not match the review question?** | | Low concern |

##### Index Test

| Index tests | CCA; PCR |
| --- | --- |

##### All tests

| **A. Risk of Bias** | |
| --- | --- |
| Were the index test results interpreted without knowledge of the results of the reference standard? | Yes |
| If a threshold was used, was it pre-specified? | Yes |
| **Could the conduct or interpretation of the index test have introduced bias?** | Low risk |

| **B. Concerns regarding applicability** | |
| --- | --- |
| **Are there concerns that the index test, its conduct, or interpretation differ from the review question?** | Low concern |

##### Reference Standard

| **A. Risk of Bias** | | |
| --- | --- | --- |
| Target condition and reference standard(s) | Duplicate Kato Katz smears | |
| Is the reference standards likely to correctly classify the target condition? | | No |
| Were the reference standard results interpreted without knowledge of the results of the index tests? | | Yes |
| **Could the reference standard, its conduct, or its interpretation have introduced bias?** | | Low risk |

| **B. Concerns regarding applicability** | |
| --- | --- |
| **Are there concerns that the target condition as defined by the reference standard does not match the question?** | Low concern |

##### Flow and Timing

| **A. Risk of Bias** | | |
| --- | --- | --- |
| Flow and timing |  | |
| Was there an appropriate interval between index test and reference standard? | | Yes |
| Did all patients receive the same reference standard? | | Yes |
| Were all patients included in the analysis? | | Yes |
| **Could the patient flow have introduced bias?** | | Low risk |

##### Notes

| **Notes** |
| --- |

#### Mafe 1997

##### Patient Selection

| **A. Risk of Bias** | | |
| --- | --- | --- |
| Patient Sampling | cross-sectional study | |
| Was a consecutive or random sample of patients enrolled? | | Yes |
| Was a case-control design avoided? | | Yes |
| Did the study avoid inappropriate exclusions? | | Unclear |
| **Could the selection of patients have introduced bias?** | | Low risk |

| **B. Concerns regarding applicability** | | |
| --- | --- | --- |
| Patient characteristics and setting | - 1056 patients from 4 riverine villages aged older than 5 years  - no previous specific schistosomiasis treatment administered in these villages  - Country: Nigeria  - Setting: field study | |
| **Are there concerns that the included patients and setting do not match the review question?** | | Low concern |

##### Index Test

| Index tests | Haematuria reagent strip |
| --- | --- |

##### All tests

| **A. Risk of Bias** | |
| --- | --- |
| Were the index test results interpreted without knowledge of the results of the reference standard? | Unclear |
| If a threshold was used, was it pre-specified? | Unclear |
| **Could the conduct or interpretation of the index test have introduced bias?** | Low risk |

| **B. Concerns regarding applicability** | |
| --- | --- |
| **Are there concerns that the index test, its conduct, or interpretation differ from the review question?** | Low concern |

##### Reference Standard

| **A. Risk of Bias** | | |
| --- | --- | --- |
| Target condition and reference standard(s) | S. haematobium  Urine filtration | |
| Is the reference standards likely to correctly classify the target condition? | | Unclear |
| Were the reference standard results interpreted without knowledge of the results of the index tests? | | Unclear |
| **Could the reference standard, its conduct, or its interpretation have introduced bias?** | | Unclear risk |

| **B. Concerns regarding applicability** | |
| --- | --- |
| **Are there concerns that the target condition as defined by the reference standard does not match the question?** | Low concern |

##### Flow and Timing

| **A. Risk of Bias** | | |
| --- | --- | --- |
| Flow and timing |  | |
| Was there an appropriate interval between index test and reference standard? | | Yes |
| Did all patients receive the same reference standard? | | Yes |
| Were all patients included in the analysis? | | Unclear |
| **Could the patient flow have introduced bias?** | | Low risk |

##### Notes

| **Notes** |
| --- |

#### Mafe 2000

##### Patient Selection

| **A. Risk of Bias** | | |
| --- | --- | --- |
| Patient Sampling | cross-sectional study | |
| Was a consecutive or random sample of patients enrolled? | | Yes |
| Was a case-control design avoided? | | Yes |
| Did the study avoid inappropriate exclusions? | | Yes |
| **Could the selection of patients have introduced bias?** | | Low risk |

| **B. Concerns regarding applicability** | | |
| --- | --- | --- |
| Patient characteristics and setting | - 529 school children from 20 schhols in Borgo Local Governement Area  - Country: Nigeria  - Setting: field study | |
| **Are there concerns that the included patients and setting do not match the review question?** | | Low concern |

##### Index Test

| Index tests | micro-haematuria reagent strips |
| --- | --- |

##### All tests

| **A. Risk of Bias** | |
| --- | --- |
| Were the index test results interpreted without knowledge of the results of the reference standard? | Unclear |
| If a threshold was used, was it pre-specified? | Unclear |
| **Could the conduct or interpretation of the index test have introduced bias?** | Low risk |

| **B. Concerns regarding applicability** | |
| --- | --- |
| **Are there concerns that the index test, its conduct, or interpretation differ from the review question?** | Low concern |

##### Reference Standard

| **A. Risk of Bias** | | |
| --- | --- | --- |
| Target condition and reference standard(s) | S. haematobium  urine microscopy | |
| Is the reference standards likely to correctly classify the target condition? | | Unclear |
| Were the reference standard results interpreted without knowledge of the results of the index tests? | | Unclear |
| **Could the reference standard, its conduct, or its interpretation have introduced bias?** | | Low risk |

| **B. Concerns regarding applicability** | |
| --- | --- |
| **Are there concerns that the target condition as defined by the reference standard does not match the question?** | Low concern |

##### Flow and Timing

| **A. Risk of Bias** | | |
| --- | --- | --- |
| Flow and timing |  | |
| Was there an appropriate interval between index test and reference standard? | | Unclear |
| Did all patients receive the same reference standard? | | Yes |
| Were all patients included in the analysis? | | Yes |
| **Could the patient flow have introduced bias?** | | Low risk |

##### Notes

| **Notes** |
| --- |

#### Magalhaes 2020

##### Patient Selection

| **A. Risk of Bias** | | |
| --- | --- | --- |
| Patient Sampling | cross-sectional population-based study | |
| Was a consecutive or random sample of patients enrolled? | | Yes |
| Was a case-control design avoided? | | Unclear |
| Did the study avoid inappropriate exclusions? | | Unclear |
| **Could the selection of patients have introduced bias?** | | Low risk |

| **B. Concerns regarding applicability** | | |
| --- | --- | --- |
| Patient characteristics and setting | - 257 patients aged between 2 and 88 years (47,5% female, 52,5% male)  - Country: Brazil  - Setting: field study | |
| **Are there concerns that the included patients and setting do not match the review question?** | | Low concern |

##### Index Test

| Index tests | 2KK, 6KK, Helmintex, POC-CCA |
| --- | --- |

##### All tests

| **A. Risk of Bias** | |
| --- | --- |
| Were the index test results interpreted without knowledge of the results of the reference standard? | Unclear |
| If a threshold was used, was it pre-specified? | Unclear |
| **Could the conduct or interpretation of the index test have introduced bias?** | Low risk |

| **B. Concerns regarding applicability** | |
| --- | --- |
| **Are there concerns that the index test, its conduct, or interpretation differ from the review question?** | Low concern |

##### Reference Standard

| **A. Risk of Bias** | | |
| --- | --- | --- |
| Target condition and reference standard(s) | S. mansoni  RT-PCR (real-time PCR) | |
| Is the reference standards likely to correctly classify the target condition? | | Unclear |
| Were the reference standard results interpreted without knowledge of the results of the index tests? | | Yes |
| **Could the reference standard, its conduct, or its interpretation have introduced bias?** | | Unclear risk |

| **B. Concerns regarding applicability** | |
| --- | --- |
| **Are there concerns that the target condition as defined by the reference standard does not match the question?** | Low concern |

##### Flow and Timing

| **A. Risk of Bias** | | |
| --- | --- | --- |
| Flow and timing |  | |
| Was there an appropriate interval between index test and reference standard? | | Yes |
| Did all patients receive the same reference standard? | | Yes |
| Were all patients included in the analysis? | | Unclear |
| **Could the patient flow have introduced bias?** | | Low risk |

##### Notes

| **Notes** |
| --- |

#### Magnussen 2001

##### Patient Selection

| **A. Risk of Bias** | | |
| --- | --- | --- |
| Patient Sampling | cohort study | |
| Was a consecutive or random sample of patients enrolled? | | Yes |
| Was a case-control design avoided? | | Yes |
| Did the study avoid inappropriate exclusions? | | Unclear |
| **Could the selection of patients have introduced bias?** | | Low risk |

| **B. Concerns regarding applicability** | | |
| --- | --- | --- |
| Patient characteristics and setting | - 170 children from 5 schools aged between 11 and 17 years  - Country: Tanzania  Setting: field study | |
| **Are there concerns that the included patients and setting do not match the review question?** | | Low concern |

##### Index Test

| Index tests | micro-haematuria urine reagent strip (Haemastix) |
| --- | --- |

##### All tests

| **A. Risk of Bias** | |
| --- | --- |
| Were the index test results interpreted without knowledge of the results of the reference standard? | Unclear |
| If a threshold was used, was it pre-specified? | Unclear |
| **Could the conduct or interpretation of the index test have introduced bias?** | Low risk |

| **B. Concerns regarding applicability** | |
| --- | --- |
| **Are there concerns that the index test, its conduct, or interpretation differ from the review question?** | Low concern |

##### Reference Standard

| **A. Risk of Bias** | | |
| --- | --- | --- |
| Target condition and reference standard(s) | S. haematobium  urine microscopy | |
| Is the reference standards likely to correctly classify the target condition? | | Unclear |
| Were the reference standard results interpreted without knowledge of the results of the index tests? | | Unclear |
| **Could the reference standard, its conduct, or its interpretation have introduced bias?** | | Unclear risk |

| **B. Concerns regarding applicability** | |
| --- | --- |
| **Are there concerns that the target condition as defined by the reference standard does not match the question?** | Low concern |

##### Flow and Timing

| **A. Risk of Bias** | | |
| --- | --- | --- |
| Flow and timing |  | |
| Was there an appropriate interval between index test and reference standard? | | Unclear |
| Did all patients receive the same reference standard? | | Yes |
| Were all patients included in the analysis? | | Unclear |
| **Could the patient flow have introduced bias?** | | Low risk |

##### Notes

| **Notes** |
| --- |

#### Mazigo 2018

##### Patient Selection

| **A. Risk of Bias** | | |
| --- | --- | --- |
| Patient Sampling | lcross-sectional study included in longitudinal study ; random sampling procedure | |
| Was a consecutive or random sample of patients enrolled? | | Yes |
| Was a case-control design avoided? | | Yes |
| Did the study avoid inappropriate exclusions? | | Unclear |
| **Could the selection of patients have introduced bias?** | | Low risk |

| **B. Concerns regarding applicability** | | |
| --- | --- | --- |
| Patient characteristics and setting | - 419 patients enrolled aged between 5 and 55 years  - Country: Tanzania  - No anti-schistosomiasis treatment in the previous 6 months  - Setting: field study | |
| **Are there concerns that the included patients and setting do not match the review question?** | | Low concern |

##### Index Test

| Index tests | POC-CCA |
| --- | --- |

##### All tests

| **A. Risk of Bias** | |
| --- | --- |
| Were the index test results interpreted without knowledge of the results of the reference standard? | Yes |
| If a threshold was used, was it pre-specified? | Unclear |
| **Could the conduct or interpretation of the index test have introduced bias?** | Low risk |

| **B. Concerns regarding applicability** | |
| --- | --- |
| **Are there concerns that the index test, its conduct, or interpretation differ from the review question?** | Low concern |

##### Reference Standard

| **A. Risk of Bias** | | |
| --- | --- | --- |
| Target condition and reference standard(s) | S. mansoni  4 Kato-Katz thick smears | |
| Is the reference standards likely to correctly classify the target condition? | | Unclear |
| Were the reference standard results interpreted without knowledge of the results of the index tests? | | Yes |
| **Could the reference standard, its conduct, or its interpretation have introduced bias?** | | Low risk |

| **B. Concerns regarding applicability** | |
| --- | --- |
| **Are there concerns that the target condition as defined by the reference standard does not match the question?** | Low concern |

##### Flow and Timing

| **A. Risk of Bias** | | |
| --- | --- | --- |
| Flow and timing |  | |
| Was there an appropriate interval between index test and reference standard? | | Yes |
| Did all patients receive the same reference standard? | | Yes |
| Were all patients included in the analysis? | | No |
| **Could the patient flow have introduced bias?** | | Low risk |

##### Notes

| **Notes** |
| --- |

#### Midzi 2009

##### Patient Selection

| **A. Risk of Bias** | | |
| --- | --- | --- |
| Patient Sampling | national survey (control programme); | |
| Was a consecutive or random sample of patients enrolled? | | Yes |
| Was a case-control design avoided? | | Yes |
| Did the study avoid inappropriate exclusions? | | Yes |
| **Could the selection of patients have introduced bias?** | | Low risk |

| **B. Concerns regarding applicability** | | |
| --- | --- | --- |
| Patient characteristics and setting | Species: S. haematobium  Country: Zimbabwe  Sample size: 265  Age range: 2—19 y  Participants: schoolcholdren  Setting: field study  Praziquantel before the study: | |
| **Are there concerns that the included patients and setting do not match the review question?** | | Low concern |

##### Index Test

| Index tests | CCA |
| --- | --- |

##### All tests

| **A. Risk of Bias** | |
| --- | --- |
| Were the index test results interpreted without knowledge of the results of the reference standard? | Yes |
| If a threshold was used, was it pre-specified? | Yes |
| **Could the conduct or interpretation of the index test have introduced bias?** | Low risk |

| **B. Concerns regarding applicability** | |
| --- | --- |
| **Are there concerns that the index test, its conduct, or interpretation differ from the review question?** | Low concern |

##### Reference Standard

| **A. Risk of Bias** | | |
| --- | --- | --- |
| Target condition and reference standard(s) | Urine microscopy | |
| Is the reference standards likely to correctly classify the target condition? | | Yes |
| Were the reference standard results interpreted without knowledge of the results of the index tests? | | Yes |
| **Could the reference standard, its conduct, or its interpretation have introduced bias?** | | Low risk |

| **B. Concerns regarding applicability** | |
| --- | --- |
| **Are there concerns that the target condition as defined by the reference standard does not match the question?** | Low concern |

##### Flow and Timing

| **A. Risk of Bias** | | |
| --- | --- | --- |
| Flow and timing |  | |
| Was there an appropriate interval between index test and reference standard? | | Yes |
| Did all patients receive the same reference standard? | | Yes |
| Were all patients included in the analysis? | | Yes |
| **Could the patient flow have introduced bias?** | | Low risk |

##### Notes

| **Notes** |
| --- |

#### Morenikeji 2014

##### Patient Selection

| **A. Risk of Bias** | | |
| --- | --- | --- |
| Patient Sampling | cross-sectional study | |
| Was a consecutive or random sample of patients enrolled? | | Yes |
| Was a case-control design avoided? | | Yes |
| Did the study avoid inappropriate exclusions? | | Yes |
| **Could the selection of patients have introduced bias?** | | Low risk |

| **B. Concerns regarding applicability** | | |
| --- | --- | --- |
| Patient characteristics and setting | - 541 children of all ages, males and females  - Country: Nigeria  - setting: field study | |
| **Are there concerns that the included patients and setting do not match the review question?** | | Low concern |

##### Index Test

| Index tests | Microhaematuria, Proteinuria |
| --- | --- |

##### All tests

| **A. Risk of Bias** | |
| --- | --- |
| Were the index test results interpreted without knowledge of the results of the reference standard? | No |
| If a threshold was used, was it pre-specified? | Unclear |
| **Could the conduct or interpretation of the index test have introduced bias?** | Low risk |

| **B. Concerns regarding applicability** | |
| --- | --- |
| **Are there concerns that the index test, its conduct, or interpretation differ from the review question?** | Low concern |

##### Reference Standard

| **A. Risk of Bias** | | |
| --- | --- | --- |
| Target condition and reference standard(s) | S. haematobium  Urine microscopy | |
| Is the reference standards likely to correctly classify the target condition? | | Unclear |
| Were the reference standard results interpreted without knowledge of the results of the index tests? | | Unclear |
| **Could the reference standard, its conduct, or its interpretation have introduced bias?** | | Low risk |

| **B. Concerns regarding applicability** | |
| --- | --- |
| **Are there concerns that the target condition as defined by the reference standard does not match the question?** | Low concern |

##### Flow and Timing

| **A. Risk of Bias** | | |
| --- | --- | --- |
| Flow and timing |  | |
| Was there an appropriate interval between index test and reference standard? | | Yes |
| Did all patients receive the same reference standard? | | Yes |
| Were all patients included in the analysis? | | No |
| **Could the patient flow have introduced bias?** | | Low risk |

##### Notes

| **Notes** |
| --- |

#### Mott 1985 Ghana

##### Patient Selection

| **A. Risk of Bias** | | |
| --- | --- | --- |
| Patient Sampling | cohort study ; consecutive sampling | |
| Was a consecutive or random sample of patients enrolled? | | Yes |
| Was a case-control design avoided? | | Yes |
| Did the study avoid inappropriate exclusions? | | Unclear |
| **Could the selection of patients have introduced bias?** | | Low risk |

| **B. Concerns regarding applicability** | | |
| --- | --- | --- |
| Patient characteristics and setting | - 562 children and adults  - Country: Ghana  - Setting: field study | |
| **Are there concerns that the included patients and setting do not match the review question?** | | Low concern |

##### Index Test

| Index tests | micro-haematuria & proteinuria by urine reagent strip |
| --- | --- |

##### All tests

| **A. Risk of Bias** | |
| --- | --- |
| Were the index test results interpreted without knowledge of the results of the reference standard? | Yes |
| If a threshold was used, was it pre-specified? | Yes |
| **Could the conduct or interpretation of the index test have introduced bias?** | Low risk |

| **B. Concerns regarding applicability** | |
| --- | --- |
| **Are there concerns that the index test, its conduct, or interpretation differ from the review question?** | Low concern |

##### Reference Standard

| **A. Risk of Bias** | | |
| --- | --- | --- |
| Target condition and reference standard(s) | S. haematobium  urine microscopy | |
| Is the reference standards likely to correctly classify the target condition? | | Unclear |
| Were the reference standard results interpreted without knowledge of the results of the index tests? | | Unclear |
| **Could the reference standard, its conduct, or its interpretation have introduced bias?** | | Unclear risk |

| **B. Concerns regarding applicability** | |
| --- | --- |
| **Are there concerns that the target condition as defined by the reference standard does not match the question?** | Low concern |

##### Flow and Timing

| **A. Risk of Bias** | | |
| --- | --- | --- |
| Flow and timing |  | |
| Was there an appropriate interval between index test and reference standard? | | Unclear |
| Did all patients receive the same reference standard? | | Yes |
| Were all patients included in the analysis? | | Yes |
| **Could the patient flow have introduced bias?** | | Low risk |

##### Notes

| **Notes** |
| --- |

#### Mott 1985 Zambia

##### Patient Selection

| **A. Risk of Bias** | | |
| --- | --- | --- |
| Patient Sampling | cohort study ; consecutive sampling | |
| Was a consecutive or random sample of patients enrolled? | | Yes |
| Was a case-control design avoided? | | Yes |
| Did the study avoid inappropriate exclusions? | | Unclear |
| **Could the selection of patients have introduced bias?** | | Low risk |

| **B. Concerns regarding applicability** | | |
| --- | --- | --- |
| Patient characteristics and setting | - 562 children and adults  - Country: Ghana  - Setting: field study | |
| **Are there concerns that the included patients and setting do not match the review question?** | | Low concern |

##### Index Test

| Index tests | micro-haematuria & proteinuria by urine reagent strip |
| --- | --- |

##### All tests

| **A. Risk of Bias** | |
| --- | --- |
| Were the index test results interpreted without knowledge of the results of the reference standard? | Yes |
| If a threshold was used, was it pre-specified? | Yes |
| **Could the conduct or interpretation of the index test have introduced bias?** | Low risk |

| **B. Concerns regarding applicability** | |
| --- | --- |
| **Are there concerns that the index test, its conduct, or interpretation differ from the review question?** | Low concern |

##### Reference Standard

| **A. Risk of Bias** | | |
| --- | --- | --- |
| Target condition and reference standard(s) | S. haematobium  urine microscopy | |
| Is the reference standards likely to correctly classify the target condition? | | Unclear |
| Were the reference standard results interpreted without knowledge of the results of the index tests? | | Unclear |
| **Could the reference standard, its conduct, or its interpretation have introduced bias?** | | Unclear risk |

| **B. Concerns regarding applicability** | |
| --- | --- |
| **Are there concerns that the target condition as defined by the reference standard does not match the question?** | Low concern |

##### Flow and Timing

| **A. Risk of Bias** | | |
| --- | --- | --- |
| Flow and timing |  | |
| Was there an appropriate interval between index test and reference standard? | | Unclear |
| Did all patients receive the same reference standard? | | Yes |
| Were all patients included in the analysis? | | Yes |
| **Could the patient flow have introduced bias?** | | Low risk |

##### Notes

| **Notes** |
| --- |

#### Mtasiwa 1996

##### Patient Selection

| **A. Risk of Bias** | | |
| --- | --- | --- |
| Patient Sampling | cross-sectional study | |
| Was a consecutive or random sample of patients enrolled? | | Unclear |
| Was a case-control design avoided? | | Yes |
| Did the study avoid inappropriate exclusions? | | Unclear |
| **Could the selection of patients have introduced bias?** | | Low risk |

| **B. Concerns regarding applicability** | | |
| --- | --- | --- |
| Patient characteristics and setting | - 404 schoolchildren aged between 7 and 15 years  - Country: Tanzania  - Setting: field study | |
| **Are there concerns that the included patients and setting do not match the review question?** | | Low concern |

##### Index Test

| Index tests | Micro-haematuria by urine reagent strip (Sangur) |
| --- | --- |

##### All tests

| **A. Risk of Bias** | |
| --- | --- |
| Were the index test results interpreted without knowledge of the results of the reference standard? | Unclear |
| If a threshold was used, was it pre-specified? | Unclear |
| **Could the conduct or interpretation of the index test have introduced bias?** | Low risk |

| **B. Concerns regarding applicability** | |
| --- | --- |
| **Are there concerns that the index test, its conduct, or interpretation differ from the review question?** | Low concern |

##### Reference Standard

| **A. Risk of Bias** | | |
| --- | --- | --- |
| Target condition and reference standard(s) | S. haematobium  Urine microscopy | |
| Is the reference standards likely to correctly classify the target condition? | | Unclear |
| Were the reference standard results interpreted without knowledge of the results of the index tests? | | Unclear |
| **Could the reference standard, its conduct, or its interpretation have introduced bias?** | | Low risk |

| **B. Concerns regarding applicability** | |
| --- | --- |
| **Are there concerns that the target condition as defined by the reference standard does not match the question?** | Low concern |

##### Flow and Timing

| **A. Risk of Bias** | | |
| --- | --- | --- |
| Flow and timing |  | |
| Was there an appropriate interval between index test and reference standard? | | Unclear |
| Did all patients receive the same reference standard? | | Yes |
| Were all patients included in the analysis? | | Yes |
| **Could the patient flow have introduced bias?** | | Low risk |

##### Notes

| **Notes** |
| --- |

#### Murare 1987

##### Patient Selection

| **A. Risk of Bias** | | |
| --- | --- | --- |
| Patient Sampling | cohort study | |
| Was a consecutive or random sample of patients enrolled? | | Unclear |
| Was a case-control design avoided? | | Yes |
| Did the study avoid inappropriate exclusions? | | Unclear |
| **Could the selection of patients have introduced bias?** | | Low risk |

| **B. Concerns regarding applicability** | | |
| --- | --- | --- |
| Patient characteristics and setting | - 232 school children aged between 9 and 14 years  - Country: Zimbabwe  - Setting: field study | |
| **Are there concerns that the included patients and setting do not match the review question?** | | Low concern |

##### Index Test

| Index tests | micro-haematuria & proteinuria by urine reagent strip |
| --- | --- |

##### All tests

| **A. Risk of Bias** | |
| --- | --- |
| Were the index test results interpreted without knowledge of the results of the reference standard? | Unclear |
| If a threshold was used, was it pre-specified? | Yes |
| **Could the conduct or interpretation of the index test have introduced bias?** | Low risk |

| **B. Concerns regarding applicability** | |
| --- | --- |
| **Are there concerns that the index test, its conduct, or interpretation differ from the review question?** | Low concern |

##### Reference Standard

| **A. Risk of Bias** | | |
| --- | --- | --- |
| Target condition and reference standard(s) | S. haematobium  urine microscopy | |
| Is the reference standards likely to correctly classify the target condition? | | Unclear |
| Were the reference standard results interpreted without knowledge of the results of the index tests? | | Unclear |
| **Could the reference standard, its conduct, or its interpretation have introduced bias?** | | Low risk |

| **B. Concerns regarding applicability** | |
| --- | --- |
| **Are there concerns that the target condition as defined by the reference standard does not match the question?** | Low concern |

##### Flow and Timing

| **A. Risk of Bias** | | |
| --- | --- | --- |
| Flow and timing |  | |
| Was there an appropriate interval between index test and reference standard? | | Yes |
| Did all patients receive the same reference standard? | | Yes |
| Were all patients included in the analysis? | | Yes |
| **Could the patient flow have introduced bias?** | | Low risk |

##### Notes

| **Notes** |
| --- |

#### Mwangi 2018

##### Patient Selection

| **A. Risk of Bias** | | |
| --- | --- | --- |
| Patient Sampling | cross-sectional study | |
| Was a consecutive or random sample of patients enrolled? | | Unclear |
| Was a case-control design avoided? | | Unclear |
| Did the study avoid inappropriate exclusions? | | Unclear |
| **Could the selection of patients have introduced bias?** | | Unclear risk |

| **B. Concerns regarding applicability** | | |
| --- | --- | --- |
| Patient characteristics and setting | - 383 school children  - Country: Kenya  - Setting: field study | |
| **Are there concerns that the included patients and setting do not match the review question?** | | Low concern |

##### Index Test

| Index tests | LAMP |
| --- | --- |

##### All tests

| **A. Risk of Bias** | |
| --- | --- |
| Were the index test results interpreted without knowledge of the results of the reference standard? | Unclear |
| If a threshold was used, was it pre-specified? | Unclear |
| **Could the conduct or interpretation of the index test have introduced bias?** | Low risk |

| **B. Concerns regarding applicability** | |
| --- | --- |
| **Are there concerns that the index test, its conduct, or interpretation differ from the review question?** | Low concern |

##### Reference Standard

| **A. Risk of Bias** | | |
| --- | --- | --- |
| Target condition and reference standard(s) | S. mansoni  Kato-Katz stool examination | |
| Is the reference standards likely to correctly classify the target condition? | | Unclear |
| Were the reference standard results interpreted without knowledge of the results of the index tests? | | Yes |
| **Could the reference standard, its conduct, or its interpretation have introduced bias?** | | Low risk |

| **B. Concerns regarding applicability** | |
| --- | --- |
| **Are there concerns that the target condition as defined by the reference standard does not match the question?** | Low concern |

##### Flow and Timing

| **A. Risk of Bias** | | |
| --- | --- | --- |
| Flow and timing |  | |
| Was there an appropriate interval between index test and reference standard? | | Yes |
| Did all patients receive the same reference standard? | | Yes |
| Were all patients included in the analysis? | | Unclear |
| **Could the patient flow have introduced bias?** | | Low risk |

##### Notes

| **Notes** |
| --- |

#### N'Goran 1989

##### Patient Selection

| **A. Risk of Bias** | | |
| --- | --- | --- |
| Patient Sampling | cross-sectional study | |
| Was a consecutive or random sample of patients enrolled? | | Unclear |
| Was a case-control design avoided? | | Yes |
| Did the study avoid inappropriate exclusions? | | Yes |
| **Could the selection of patients have introduced bias?** | | Low risk |

| **B. Concerns regarding applicability** | | |
| --- | --- | --- |
| Patient characteristics and setting | - 1059 inhabitants of village  - Country: Ivory Coast  - Setting: field study | |
| **Are there concerns that the included patients and setting do not match the review question?** | | Low concern |

##### Index Test

| Index tests | micro-haematuria by urine reagent strip (Hemastix) |
| --- | --- |

##### All tests

| **A. Risk of Bias** | |
| --- | --- |
| Were the index test results interpreted without knowledge of the results of the reference standard? | Unclear |
| If a threshold was used, was it pre-specified? | Yes |
| **Could the conduct or interpretation of the index test have introduced bias?** | Low risk |

| **B. Concerns regarding applicability** | |
| --- | --- |
| **Are there concerns that the index test, its conduct, or interpretation differ from the review question?** | Low concern |

##### Reference Standard

| **A. Risk of Bias** | | |
| --- | --- | --- |
| Target condition and reference standard(s) | S. haematobium  urine microscopy | |
| Is the reference standards likely to correctly classify the target condition? | | No |
| Were the reference standard results interpreted without knowledge of the results of the index tests? | | Unclear |
| **Could the reference standard, its conduct, or its interpretation have introduced bias?** | | Low risk |

| **B. Concerns regarding applicability** | |
| --- | --- |
| **Are there concerns that the target condition as defined by the reference standard does not match the question?** | Low concern |

##### Flow and Timing

| **A. Risk of Bias** | | |
| --- | --- | --- |
| Flow and timing |  | |
| Was there an appropriate interval between index test and reference standard? | | Unclear |
| Did all patients receive the same reference standard? | | Unclear |
| Were all patients included in the analysis? | | Yes |
| **Could the patient flow have introduced bias?** | | Low risk |

##### Notes

| **Notes** |
| --- |

#### Nausch 2014

##### Patient Selection

| **A. Risk of Bias** | | |
| --- | --- | --- |
| Patient Sampling | cross sectional; no random selection | |
| Was a consecutive or random sample of patients enrolled? | | Yes |
| Was a case-control design avoided? | | Yes |
| Did the study avoid inappropriate exclusions? | | Yes |
| **Could the selection of patients have introduced bias?** | | Low risk |

| **B. Concerns regarding applicability** | | |
| --- | --- | --- |
| Patient characteristics and setting | Species: S. haematobium and S. mansoni  Country: Zimbabwe  Sample size:  Age range: 1-12 y  Participants: children  Setting: field study  Praziquantel before the study: no | |
| **Are there concerns that the included patients and setting do not match the review question?** | | Low concern |

##### Index Test

| Index tests | Schistosoma mansoni cercarial transformation fluid (SmCTF); IgM-cercarial antigen preparation (CAP) and IgM-soluble egg antigen (SEA) ELISA |
| --- | --- |

##### All tests

| **A. Risk of Bias** | |
| --- | --- |
| Were the index test results interpreted without knowledge of the results of the reference standard? | Yes |
| If a threshold was used, was it pre-specified? | Yes |
| **Could the conduct or interpretation of the index test have introduced bias?** | Low risk |

| **B. Concerns regarding applicability** | |
| --- | --- |
| **Are there concerns that the index test, its conduct, or interpretation differ from the review question?** | Low concern |

##### Reference Standard

| **A. Risk of Bias** | | |
| --- | --- | --- |
| Target condition and reference standard(s) | quadriplicate Kato Katz smears; urine microscopy | |
| Is the reference standards likely to correctly classify the target condition? | | No |
| Were the reference standard results interpreted without knowledge of the results of the index tests? | | Yes |
| **Could the reference standard, its conduct, or its interpretation have introduced bias?** | | Low risk |

| **B. Concerns regarding applicability** | |
| --- | --- |
| **Are there concerns that the target condition as defined by the reference standard does not match the question?** | Low concern |

##### Flow and Timing

| **A. Risk of Bias** | | |
| --- | --- | --- |
| Flow and timing |  | |
| Was there an appropriate interval between index test and reference standard? | | Yes |
| Did all patients receive the same reference standard? | | Yes |
| Were all patients included in the analysis? | | Yes |
| **Could the patient flow have introduced bias?** | | Low risk |

##### Notes

| **Notes** |
| --- |

#### Navaratnam 2012

##### Patient Selection

| **A. Risk of Bias** | | |
| --- | --- | --- |
| Patient Sampling | cross-sectional study | |
| Was a consecutive or random sample of patients enrolled? | | Yes |
| Was a case-control design avoided? | | Unclear |
| Did the study avoid inappropriate exclusions? | | Unclear |
| **Could the selection of patients have introduced bias?** | | Low risk |

| **B. Concerns regarding applicability** | | |
| --- | --- | --- |
| Patient characteristics and setting | - 569 preschool children from 4 villages aged between 1 and 5 years  - Country: Uganda  - Setting: field study | |
| **Are there concerns that the included patients and setting do not match the review question?** | | Low concern |

##### Index Test

| Index tests | CCA |
| --- | --- |

##### All tests

| **A. Risk of Bias** | |
| --- | --- |
| Were the index test results interpreted without knowledge of the results of the reference standard? | Unclear |
| If a threshold was used, was it pre-specified? | Unclear |
| **Could the conduct or interpretation of the index test have introduced bias?** | Low risk |

| **B. Concerns regarding applicability** | |
| --- | --- |
| **Are there concerns that the index test, its conduct, or interpretation differ from the review question?** | Low concern |

##### Reference Standard

| **A. Risk of Bias** | | |
| --- | --- | --- |
| Target condition and reference standard(s) | S. mansoni  duplicate KK | |
| Is the reference standards likely to correctly classify the target condition? | | Unclear |
| Were the reference standard results interpreted without knowledge of the results of the index tests? | | Unclear |
| **Could the reference standard, its conduct, or its interpretation have introduced bias?** | | Low risk |

| **B. Concerns regarding applicability** | |
| --- | --- |
| **Are there concerns that the target condition as defined by the reference standard does not match the question?** | Low concern |

##### Flow and Timing

| **A. Risk of Bias** | | |
| --- | --- | --- |
| Flow and timing |  | |
| Was there an appropriate interval between index test and reference standard? | | Yes |
| Did all patients receive the same reference standard? | | Yes |
| Were all patients included in the analysis? | | Unclear |
| **Could the patient flow have introduced bias?** | | Low risk |

##### Notes

| **Notes** |
| --- |

#### Ndamukong 2001

##### Patient Selection

| **A. Risk of Bias** | | |
| --- | --- | --- |
| Patient Sampling | cross-sectional study | |
| Was a consecutive or random sample of patients enrolled? | | Unclear |
| Was a case-control design avoided? | | Yes |
| Did the study avoid inappropriate exclusions? | | Unclear |
| **Could the selection of patients have introduced bias?** | | Low risk |

| **B. Concerns regarding applicability** | | |
| --- | --- | --- |
| Patient characteristics and setting | - 347 school children from 6 primary schools aged between 5 and 16 years  - Country: Cameroon  - Setting: field study | |
| **Are there concerns that the included patients and setting do not match the review question?** | | Low concern |

##### Index Test

| Index tests | urine reagent strips:  - Hemastix for haematuria  - Albustix for proteinuria |
| --- | --- |

##### All tests

| **A. Risk of Bias** | |
| --- | --- |
| Were the index test results interpreted without knowledge of the results of the reference standard? | Unclear |
| If a threshold was used, was it pre-specified? | Unclear |
| **Could the conduct or interpretation of the index test have introduced bias?** | Low risk |

| **B. Concerns regarding applicability** | |
| --- | --- |
| **Are there concerns that the index test, its conduct, or interpretation differ from the review question?** | Low concern |

##### Reference Standard

| **A. Risk of Bias** | | |
| --- | --- | --- |
| Target condition and reference standard(s) | S. haematobium  urine microscopy | |
| Is the reference standards likely to correctly classify the target condition? | | Unclear |
| Were the reference standard results interpreted without knowledge of the results of the index tests? | | Unclear |
| **Could the reference standard, its conduct, or its interpretation have introduced bias?** | | Unclear risk |

| **B. Concerns regarding applicability** | |
| --- | --- |
| **Are there concerns that the target condition as defined by the reference standard does not match the question?** | Low concern |

##### Flow and Timing

| **A. Risk of Bias** | | |
| --- | --- | --- |
| Flow and timing |  | |
| Was there an appropriate interval between index test and reference standard? | | Unclear |
| Did all patients receive the same reference standard? | | Yes |
| Were all patients included in the analysis? | | Unclear |
| **Could the patient flow have introduced bias?** | | Low risk |

##### Notes

| **Notes** |
| --- |

#### Ndhlovu 1996

##### Patient Selection

| **A. Risk of Bias** | | |
| --- | --- | --- |
| Patient Sampling | Nested case-control design | |
| Was a consecutive or random sample of patients enrolled? | | Unclear |
| Was a case-control design avoided? | | No |
| Did the study avoid inappropriate exclusions? | | Unclear |
| **Could the selection of patients have introduced bias?** | | Unclear risk |

| **B. Concerns regarding applicability** | | |
| --- | --- | --- |
| Patient characteristics and setting | - 179 patients of all ages  - People with S. mansoni infection excluded from study  - Country: Zimbabwe  - Setting: field study | |
| **Are there concerns that the included patients and setting do not match the review question?** | | Low concern |

##### Index Test

| Index tests | CAA ELISA Serum |
| --- | --- |

##### All tests

| **A. Risk of Bias** | |
| --- | --- |
| Were the index test results interpreted without knowledge of the results of the reference standard? | Unclear |
| If a threshold was used, was it pre-specified? | No |
| **Could the conduct or interpretation of the index test have introduced bias?** | Low risk |

| **B. Concerns regarding applicability** | |
| --- | --- |
| **Are there concerns that the index test, its conduct, or interpretation differ from the review question?** | Low concern |

##### Reference Standard

| **A. Risk of Bias** | | |
| --- | --- | --- |
| Target condition and reference standard(s) | S. haematobium  urine microscopy | |
| Is the reference standards likely to correctly classify the target condition? | | Unclear |
| Were the reference standard results interpreted without knowledge of the results of the index tests? | | Unclear |
| **Could the reference standard, its conduct, or its interpretation have introduced bias?** | | Low risk |

| **B. Concerns regarding applicability** | |
| --- | --- |
| **Are there concerns that the target condition as defined by the reference standard does not match the question?** | Low concern |

##### Flow and Timing

| **A. Risk of Bias** | | |
| --- | --- | --- |
| Flow and timing |  | |
| Was there an appropriate interval between index test and reference standard? | | Unclear |
| Did all patients receive the same reference standard? | | Yes |
| Were all patients included in the analysis? | | No |
| **Could the patient flow have introduced bias?** | | Low risk |

##### Notes

| **Notes** |
| --- |

#### Nduka 1995

##### Patient Selection

| **A. Risk of Bias** | | |
| --- | --- | --- |
| Patient Sampling | cross-sectional study | |
| Was a consecutive or random sample of patients enrolled? | | Unclear |
| Was a case-control design avoided? | | Yes |
| Did the study avoid inappropriate exclusions? | | Unclear |
| **Could the selection of patients have introduced bias?** | | Low risk |

| **B. Concerns regarding applicability** | | |
| --- | --- | --- |
| Patient characteristics and setting | - 1165 school children aged between 6 and 21 years  - Country: Nigeria  - Setting: field study | |
| **Are there concerns that the included patients and setting do not match the review question?** | | Low concern |

##### Index Test

| Index tests | Micro-haematuria by urine reagent strip (Medi-Test Combi-9) |
| --- | --- |

##### All tests

| **A. Risk of Bias** | |
| --- | --- |
| Were the index test results interpreted without knowledge of the results of the reference standard? | Unclear |
| If a threshold was used, was it pre-specified? | Yes |
| **Could the conduct or interpretation of the index test have introduced bias?** | Low risk |

| **B. Concerns regarding applicability** | |
| --- | --- |
| **Are there concerns that the index test, its conduct, or interpretation differ from the review question?** | Low concern |

##### Reference Standard

| **A. Risk of Bias** | | |
| --- | --- | --- |
| Target condition and reference standard(s) | S. haematobium  Urine microscopy | |
| Is the reference standards likely to correctly classify the target condition? | | Unclear |
| Were the reference standard results interpreted without knowledge of the results of the index tests? | | Unclear |
| **Could the reference standard, its conduct, or its interpretation have introduced bias?** | | Low risk |

| **B. Concerns regarding applicability** | |
| --- | --- |
| **Are there concerns that the target condition as defined by the reference standard does not match the question?** | Low concern |

##### Flow and Timing

| **A. Risk of Bias** | | |
| --- | --- | --- |
| Flow and timing |  | |
| Was there an appropriate interval between index test and reference standard? | | Yes |
| Did all patients receive the same reference standard? | | Yes |
| Were all patients included in the analysis? | | Yes |
| **Could the patient flow have introduced bias?** | | Low risk |

##### Notes

| **Notes** |
| --- |

#### Ndyomugyenyi 2001

##### Patient Selection

| **A. Risk of Bias** | | |
| --- | --- | --- |
| Patient Sampling | cross-sectional study | |
| Was a consecutive or random sample of patients enrolled? | | Unclear |
| Was a case-control design avoided? | | Yes |
| Did the study avoid inappropriate exclusions? | | Yes |
| **Could the selection of patients have introduced bias?** | | Low risk |

| **B. Concerns regarding applicability** | | |
| --- | --- | --- |
| Patient characteristics and setting | - 483 children from 3 primary schools aged between 5 and 19 years  - Country: Tanzania  - Setting: field study | |
| **Are there concerns that the included patients and setting do not match the review question?** | | Low concern |

##### Index Test

| Index tests | micro-haematuria urine reagent strip (Multistix) |
| --- | --- |

##### All tests

| **A. Risk of Bias** | |
| --- | --- |
| Were the index test results interpreted without knowledge of the results of the reference standard? | Yes |
| If a threshold was used, was it pre-specified? | Unclear |
| **Could the conduct or interpretation of the index test have introduced bias?** | Low risk |

| **B. Concerns regarding applicability** | |
| --- | --- |
| **Are there concerns that the index test, its conduct, or interpretation differ from the review question?** | Low concern |

##### Reference Standard

| **A. Risk of Bias** | | |
| --- | --- | --- |
| Target condition and reference standard(s) | S. haematobium  urine microscopy | |
| Is the reference standards likely to correctly classify the target condition? | | Unclear |
| Were the reference standard results interpreted without knowledge of the results of the index tests? | | Unclear |
| **Could the reference standard, its conduct, or its interpretation have introduced bias?** | | Unclear risk |

| **B. Concerns regarding applicability** | |
| --- | --- |
| **Are there concerns that the target condition as defined by the reference standard does not match the question?** | Low concern |

##### Flow and Timing

| **A. Risk of Bias** | | |
| --- | --- | --- |
| Flow and timing |  | |
| Was there an appropriate interval between index test and reference standard? | | Yes |
| Did all patients receive the same reference standard? | | Yes |
| Were all patients included in the analysis? | | Yes |
| **Could the patient flow have introduced bias?** | | Low risk |

##### Notes

| **Notes** |
| --- |

#### Ng'andu 1988

##### Patient Selection

| **A. Risk of Bias** | | |
| --- | --- | --- |
| Patient Sampling | cross-sectional study | |
| Was a consecutive or random sample of patients enrolled? | | Unclear |
| Was a case-control design avoided? | | Yes |
| Did the study avoid inappropriate exclusions? | | Yes |
| **Could the selection of patients have introduced bias?** | | Low risk |

| **B. Concerns regarding applicability** | | |
| --- | --- | --- |
| Patient characteristics and setting | - 412 school children from 9 primary schools aged between 4 and 19 years  - Country: Zambia  - Setting: field study | |
| **Are there concerns that the included patients and setting do not match the review question?** | | Low concern |

##### Index Test

| Index tests | micro-haematuria & proteinuria by urine reagent strip |
| --- | --- |

##### All tests

| **A. Risk of Bias** | |
| --- | --- |
| Were the index test results interpreted without knowledge of the results of the reference standard? | Unclear |
| If a threshold was used, was it pre-specified? | Unclear |
| **Could the conduct or interpretation of the index test have introduced bias?** | Unclear risk |

| **B. Concerns regarding applicability** | |
| --- | --- |
| **Are there concerns that the index test, its conduct, or interpretation differ from the review question?** | Low concern |

##### Reference Standard

| **A. Risk of Bias** | | |
| --- | --- | --- |
| Target condition and reference standard(s) | S. haematobium  urine microscopy | |
| Is the reference standards likely to correctly classify the target condition? | | Unclear |
| Were the reference standard results interpreted without knowledge of the results of the index tests? | | Unclear |
| **Could the reference standard, its conduct, or its interpretation have introduced bias?** | | Low risk |

| **B. Concerns regarding applicability** | |
| --- | --- |
| **Are there concerns that the target condition as defined by the reference standard does not match the question?** | Low concern |

##### Flow and Timing

| **A. Risk of Bias** | | |
| --- | --- | --- |
| Flow and timing |  | |
| Was there an appropriate interval between index test and reference standard? | | Unclear |
| Did all patients receive the same reference standard? | | Yes |
| Were all patients included in the analysis? | | No |
| **Could the patient flow have introduced bias?** | | Low risk |

##### Notes

| **Notes** |
| --- |

#### Ngasala 2020 Mta Dam area

##### Patient Selection

| **A. Risk of Bias** | | |
| --- | --- | --- |
| Patient Sampling | cross-sectional study ; multi-stage sampling technique | |
| Was a consecutive or random sample of patients enrolled? | | Yes |
| Was a case-control design avoided? | | Yes |
| Did the study avoid inappropriate exclusions? | | Unclear |
| **Could the selection of patients have introduced bias?** | | Low risk |

| **B. Concerns regarding applicability** | | |
| --- | --- | --- |
| Patient characteristics and setting | - 353 children from 4 primary schools between 5 and 16 years (209 male, 144 female)  - Period: between March and June 2015  - Country: Tanzania  - Setting: field study | |
| **Are there concerns that the included patients and setting do not match the review question?** | | Low concern |

##### Index Test

| Index tests | Urine reagent strips |
| --- | --- |

##### All tests

| **A. Risk of Bias** | |
| --- | --- |
| Were the index test results interpreted without knowledge of the results of the reference standard? | Unclear |
| If a threshold was used, was it pre-specified? | Unclear |
| **Could the conduct or interpretation of the index test have introduced bias?** | Low risk |

| **B. Concerns regarding applicability** | |
| --- | --- |
| **Are there concerns that the index test, its conduct, or interpretation differ from the review question?** | Low concern |

##### Reference Standard

| **A. Risk of Bias** | | |
| --- | --- | --- |
| Target condition and reference standard(s) | S. haematobium  urine microscopy | |
| Is the reference standards likely to correctly classify the target condition? | | Unclear |
| Were the reference standard results interpreted without knowledge of the results of the index tests? | | Unclear |
| **Could the reference standard, its conduct, or its interpretation have introduced bias?** | | Low risk |

| **B. Concerns regarding applicability** | |
| --- | --- |
| **Are there concerns that the target condition as defined by the reference standard does not match the question?** | Low concern |

##### Flow and Timing

| **A. Risk of Bias** | | |
| --- | --- | --- |
| Flow and timing |  | |
| Was there an appropriate interval between index test and reference standard? | | Yes |
| Did all patients receive the same reference standard? | | Yes |
| Were all patients included in the analysis? | | Unclear |
| **Could the patient flow have introduced bias?** | | Low risk |

##### Notes

| **Notes** |
| --- |

#### Ngasala 2020 Uwandani Shehia

##### Patient Selection

| **A. Risk of Bias** | | |
| --- | --- | --- |
| Patient Sampling | cross-sectional study ; multi-stage sampling technique | |
| Was a consecutive or random sample of patients enrolled? | | Yes |
| Was a case-control design avoided? | | Yes |
| Did the study avoid inappropriate exclusions? | | Unclear |
| **Could the selection of patients have introduced bias?** | | Low risk |

| **B. Concerns regarding applicability** | | |
| --- | --- | --- |
| Patient characteristics and setting | - 150 children aged between 7 and 14 years (66 male, 84 female)  - Period: April - May 2016  - Country: Tanzania  - Setting: field study | |
| **Are there concerns that the included patients and setting do not match the review question?** | | Low concern |

##### Index Test

| Index tests | urine reagent strips |
| --- | --- |

##### All tests

| **A. Risk of Bias** | |
| --- | --- |
| Were the index test results interpreted without knowledge of the results of the reference standard? | Unclear |
| If a threshold was used, was it pre-specified? | Unclear |
| **Could the conduct or interpretation of the index test have introduced bias?** | Low risk |

| **B. Concerns regarding applicability** | |
| --- | --- |
| **Are there concerns that the index test, its conduct, or interpretation differ from the review question?** | Low concern |

##### Reference Standard

| **A. Risk of Bias** | | |
| --- | --- | --- |
| Target condition and reference standard(s) | S. haematobium  urine microscopy | |
| Is the reference standards likely to correctly classify the target condition? | | Unclear |
| Were the reference standard results interpreted without knowledge of the results of the index tests? | | Unclear |
| **Could the reference standard, its conduct, or its interpretation have introduced bias?** | | Low risk |

| **B. Concerns regarding applicability** | |
| --- | --- |
| **Are there concerns that the target condition as defined by the reference standard does not match the question?** | Low concern |

##### Flow and Timing

| **A. Risk of Bias** | | |
| --- | --- | --- |
| Flow and timing |  | |
| Was there an appropriate interval between index test and reference standard? | | Yes |
| Did all patients receive the same reference standard? | | Yes |
| Were all patients included in the analysis? | | Unclear |
| **Could the patient flow have introduced bias?** | | Low risk |

##### Notes

| **Notes** |
| --- |

#### Nwaorgu 1992

##### Patient Selection

| **A. Risk of Bias** | | |
| --- | --- | --- |
| Patient Sampling | cross sectional survey; random selectionof households through schoolchildren | |
| Was a consecutive or random sample of patients enrolled? | | Yes |
| Was a case-control design avoided? | | Yes |
| Did the study avoid inappropriate exclusions? | | Yes |
| **Could the selection of patients have introduced bias?** | | Low risk |

| **B. Concerns regarding applicability** | | |
| --- | --- | --- |
| Patient characteristics and setting | Species: S. haematobium  Country: Nigeria  Sample size: 1017  Age range: all  Participants: villagers  Setting: field study  Praziquantel before the study: | |
| **Are there concerns that the included patients and setting do not match the review question?** | | Low concern |

##### Index Test

| Index tests | haematuria; proteinuria |
| --- | --- |

##### All tests

| **A. Risk of Bias** | |
| --- | --- |
| Were the index test results interpreted without knowledge of the results of the reference standard? | Unclear |
| If a threshold was used, was it pre-specified? | Yes |
| **Could the conduct or interpretation of the index test have introduced bias?** | Unclear risk |

| **B. Concerns regarding applicability** | |
| --- | --- |
| **Are there concerns that the index test, its conduct, or interpretation differ from the review question?** | Low concern |

##### Reference Standard

| **A. Risk of Bias** | | |
| --- | --- | --- |
| Target condition and reference standard(s) | Urine microscopy | |
| Is the reference standards likely to correctly classify the target condition? | | Yes |
| Were the reference standard results interpreted without knowledge of the results of the index tests? | | Unclear |
| **Could the reference standard, its conduct, or its interpretation have introduced bias?** | | Unclear risk |

| **B. Concerns regarding applicability** | |
| --- | --- |
| **Are there concerns that the target condition as defined by the reference standard does not match the question?** | Low concern |

##### Flow and Timing

| **A. Risk of Bias** | | |
| --- | --- | --- |
| Flow and timing |  | |
| Was there an appropriate interval between index test and reference standard? | | Yes |
| Did all patients receive the same reference standard? | | Yes |
| Were all patients included in the analysis? | | Yes |
| **Could the patient flow have introduced bias?** | | Low risk |

##### Notes

| **Notes** |
| --- |

#### Ofori 1986

##### Patient Selection

| **A. Risk of Bias** | | |
| --- | --- | --- |
| Patient Sampling | cross-sectional study | |
| Was a consecutive or random sample of patients enrolled? | | Unclear |
| Was a case-control design avoided? | | Yes |
| Did the study avoid inappropriate exclusions? | | Unclear |
| **Could the selection of patients have introduced bias?** | | Low risk |

| **B. Concerns regarding applicability** | | |
| --- | --- | --- |
| Patient characteristics and setting | - 118 pupils  - Country: Ghana  - Setting: field study | |
| **Are there concerns that the included patients and setting do not match the review question?** | | Low concern |

##### Index Test

| Index tests | micro-haematuria & proteinuria by urine reagent strip (N-Multistix SG) |
| --- | --- |

##### All tests

| **A. Risk of Bias** | |
| --- | --- |
| Were the index test results interpreted without knowledge of the results of the reference standard? | Unclear |
| If a threshold was used, was it pre-specified? | Unclear |
| **Could the conduct or interpretation of the index test have introduced bias?** | Low risk |

| **B. Concerns regarding applicability** | |
| --- | --- |
| **Are there concerns that the index test, its conduct, or interpretation differ from the review question?** | Low concern |

##### Reference Standard

| **A. Risk of Bias** | | |
| --- | --- | --- |
| Target condition and reference standard(s) | S. haematobium  urine microscopy | |
| Is the reference standards likely to correctly classify the target condition? | | Unclear |
| Were the reference standard results interpreted without knowledge of the results of the index tests? | | Unclear |
| **Could the reference standard, its conduct, or its interpretation have introduced bias?** | | Low risk |

| **B. Concerns regarding applicability** | |
| --- | --- |
| **Are there concerns that the target condition as defined by the reference standard does not match the question?** | Low concern |

##### Flow and Timing

| **A. Risk of Bias** | | |
| --- | --- | --- |
| Flow and timing |  | |
| Was there an appropriate interval between index test and reference standard? | | Unclear |
| Did all patients receive the same reference standard? | | Yes |
| Were all patients included in the analysis? | | Yes |
| **Could the patient flow have introduced bias?** | | Low risk |

##### Notes

| **Notes** |
| --- |

#### Okeke 2014 (LPA)

##### Patient Selection

| **A. Risk of Bias** | | |
| --- | --- | --- |
| Patient Sampling | cross-sectional study, multi-stage sampling technique | |
| Was a consecutive or random sample of patients enrolled? | | Unclear |
| Was a case-control design avoided? | | Yes |
| Did the study avoid inappropriate exclusions? | | Yes |
| **Could the selection of patients have introduced bias?** | | Low risk |

| **B. Concerns regarding applicability** | | |
| --- | --- | --- |
| Patient characteristics and setting | - children aged between 5 and 13 years  - 296 patients from low prevelance area (LPA)  - Country: Nigeria | |
| **Are there concerns that the included patients and setting do not match the review question?** | | Low concern |

##### Index Test

| Index tests | Micro-haematuria & Proteinuria |
| --- | --- |

##### All tests

| **A. Risk of Bias** | |
| --- | --- |
| Were the index test results interpreted without knowledge of the results of the reference standard? | Yes |
| If a threshold was used, was it pre-specified? | Unclear |
| **Could the conduct or interpretation of the index test have introduced bias?** | Low risk |

| **B. Concerns regarding applicability** | |
| --- | --- |
| **Are there concerns that the index test, its conduct, or interpretation differ from the review question?** | Low concern |

##### Reference Standard

| **A. Risk of Bias** | | |
| --- | --- | --- |
| Target condition and reference standard(s) | S. haematobium  Egg microscopy | |
| Is the reference standards likely to correctly classify the target condition? | | Unclear |
| Were the reference standard results interpreted without knowledge of the results of the index tests? | | Yes |
| **Could the reference standard, its conduct, or its interpretation have introduced bias?** | | Unclear risk |

| **B. Concerns regarding applicability** | |
| --- | --- |
| **Are there concerns that the target condition as defined by the reference standard does not match the question?** | Low concern |

##### Flow and Timing

| **A. Risk of Bias** | | |
| --- | --- | --- |
| Flow and timing |  | |
| Was there an appropriate interval between index test and reference standard? | | Yes |
| Did all patients receive the same reference standard? | | Yes |
| Were all patients included in the analysis? | | No |
| **Could the patient flow have introduced bias?** | | Low risk |

##### Notes

| **Notes** |
| --- |

#### Okeke 2014 (MPA)

##### Patient Selection

| **A. Risk of Bias** | | |
| --- | --- | --- |
| Patient Sampling | cross-sectional study, multi-stage sampling technique | |
| Was a consecutive or random sample of patients enrolled? | | Unclear |
| Was a case-control design avoided? | | Yes |
| Did the study avoid inappropriate exclusions? | | Yes |
| **Could the selection of patients have introduced bias?** | | Low risk |

| **B. Concerns regarding applicability** | | |
| --- | --- | --- |
| Patient characteristics and setting | - children aged between 5 and 13 years  - 184 patients from moderate prevalence area (MPA)  - Country: Nigeria | |
| **Are there concerns that the included patients and setting do not match the review question?** | | Low concern |

##### Index Test

| Index tests | Micro-haematuria & Proteinuria |
| --- | --- |

##### All tests

| **A. Risk of Bias** | |
| --- | --- |
| Were the index test results interpreted without knowledge of the results of the reference standard? | Yes |
| If a threshold was used, was it pre-specified? | Unclear |
| **Could the conduct or interpretation of the index test have introduced bias?** | Low risk |

| **B. Concerns regarding applicability** | |
| --- | --- |
| **Are there concerns that the index test, its conduct, or interpretation differ from the review question?** | Low concern |

##### Reference Standard

| **A. Risk of Bias** | | |
| --- | --- | --- |
| Target condition and reference standard(s) | S. haematobium  Egg microscopy | |
| Is the reference standards likely to correctly classify the target condition? | | Unclear |
| Were the reference standard results interpreted without knowledge of the results of the index tests? | | Yes |
| **Could the reference standard, its conduct, or its interpretation have introduced bias?** | | Unclear risk |

| **B. Concerns regarding applicability** | |
| --- | --- |
| **Are there concerns that the target condition as defined by the reference standard does not match the question?** | Low concern |

##### Flow and Timing

| **A. Risk of Bias** | | |
| --- | --- | --- |
| Flow and timing |  | |
| Was there an appropriate interval between index test and reference standard? | | Yes |
| Did all patients receive the same reference standard? | | Yes |
| Were all patients included in the analysis? | | No |
| **Could the patient flow have introduced bias?** | | Low risk |

##### Notes

| **Notes** |
| --- |

#### Onayade 1996

##### Patient Selection

| **A. Risk of Bias** | | |
| --- | --- | --- |
| Patient Sampling | cross-sectional study ; consecutive sampling | |
| Was a consecutive or random sample of patients enrolled? | | Yes |
| Was a case-control design avoided? | | Yes |
| Did the study avoid inappropriate exclusions? | | Yes |
| **Could the selection of patients have introduced bias?** | | Low risk |

| **B. Concerns regarding applicability** | | |
| --- | --- | --- |
| Patient characteristics and setting | - 105 children aged between 8 and 16 years  - Country: Nigeria  - Setting: field study | |
| **Are there concerns that the included patients and setting do not match the review question?** | | Low concern |

##### Index Test

| Index tests | proteinuria by urine reagent strip |
| --- | --- |

##### All tests

| **A. Risk of Bias** | |
| --- | --- |
| Were the index test results interpreted without knowledge of the results of the reference standard? | Yes |
| If a threshold was used, was it pre-specified? | No |
| **Could the conduct or interpretation of the index test have introduced bias?** | Low risk |

| **B. Concerns regarding applicability** | |
| --- | --- |
| **Are there concerns that the index test, its conduct, or interpretation differ from the review question?** | Low concern |

##### Reference Standard

| **A. Risk of Bias** | | |
| --- | --- | --- |
| Target condition and reference standard(s) | S. haematobium  urine microscopy | |
| Is the reference standards likely to correctly classify the target condition? | | Unclear |
| Were the reference standard results interpreted without knowledge of the results of the index tests? | | Unclear |
| **Could the reference standard, its conduct, or its interpretation have introduced bias?** | | Low risk |

| **B. Concerns regarding applicability** | |
| --- | --- |
| **Are there concerns that the target condition as defined by the reference standard does not match the question?** | Low concern |

##### Flow and Timing

| **A. Risk of Bias** | | |
| --- | --- | --- |
| Flow and timing |  | |
| Was there an appropriate interval between index test and reference standard? | | Yes |
| Did all patients receive the same reference standard? | | Yes |
| Were all patients included in the analysis? | | Yes |
| **Could the patient flow have introduced bias?** | | Low risk |

##### Notes

| **Notes** |
| --- |

#### Poggensee 2000 (HPA)

##### Patient Selection

| **A. Risk of Bias** | | |
| --- | --- | --- |
| Patient Sampling | Cross-sectional study ; non–probability-based sampling procedure | |
| Was a consecutive or random sample of patients enrolled? | | No |
| Was a case-control design avoided? | | Yes |
| Did the study avoid inappropriate exclusions? | | Unclear |
| **Could the selection of patients have introduced bias?** | | Unclear risk |

| **B. Concerns regarding applicability** | | |
| --- | --- | --- |
| Patient characteristics and setting | - 175 childbearing women aged between 15 and 60 years  - Country: Tanzania  - Setting: field study in High Prevelance Area | |
| **Are there concerns that the included patients and setting do not match the review question?** | | Low concern |

##### Index Test

| Index tests | micro-haematuria, proteinuria & leukocyturia by reagent strip |
| --- | --- |

##### All tests

| **A. Risk of Bias** | |
| --- | --- |
| Were the index test results interpreted without knowledge of the results of the reference standard? | Unclear |
| If a threshold was used, was it pre-specified? | Unclear |
| **Could the conduct or interpretation of the index test have introduced bias?** | Low risk |

| **B. Concerns regarding applicability** | |
| --- | --- |
| **Are there concerns that the index test, its conduct, or interpretation differ from the review question?** | Low concern |

##### Reference Standard

| **A. Risk of Bias** | | |
| --- | --- | --- |
| Target condition and reference standard(s) | S. haematobium  urine microscopy | |
| Is the reference standards likely to correctly classify the target condition? | | Unclear |
| Were the reference standard results interpreted without knowledge of the results of the index tests? | | Unclear |
| **Could the reference standard, its conduct, or its interpretation have introduced bias?** | | Low risk |

| **B. Concerns regarding applicability** | |
| --- | --- |
| **Are there concerns that the target condition as defined by the reference standard does not match the question?** | Low concern |

##### Flow and Timing

| **A. Risk of Bias** | | |
| --- | --- | --- |
| Flow and timing |  | |
| Was there an appropriate interval between index test and reference standard? | | Yes |
| Did all patients receive the same reference standard? | | Yes |
| Were all patients included in the analysis? | | Yes |
| **Could the patient flow have introduced bias?** | | Low risk |

##### Notes

| **Notes** |
| --- |

#### Poggensee 2000 (LPA)

##### Patient Selection

| **A. Risk of Bias** | | |
| --- | --- | --- |
| Patient Sampling | Cross-sectional study ; non–probability-based sampling procedure | |
| Was a consecutive or random sample of patients enrolled? | | No |
| Was a case-control design avoided? | | Yes |
| Did the study avoid inappropriate exclusions? | | Unclear |
| **Could the selection of patients have introduced bias?** | | Unclear risk |

| **B. Concerns regarding applicability** | | |
| --- | --- | --- |
| Patient characteristics and setting | - 175 childbearing women aged between 15 and 60 years  - Country: Tanzania  - Setting: field study in High Prevelance Area | |
| **Are there concerns that the included patients and setting do not match the review question?** | | Low concern |

##### Index Test

| Index tests | micro-haematuria, proteinuria & leukocyturia by reagent strip |
| --- | --- |

##### All tests

| **A. Risk of Bias** | |
| --- | --- |
| Were the index test results interpreted without knowledge of the results of the reference standard? | Unclear |
| If a threshold was used, was it pre-specified? | Unclear |
| **Could the conduct or interpretation of the index test have introduced bias?** | Low risk |

| **B. Concerns regarding applicability** | |
| --- | --- |
| **Are there concerns that the index test, its conduct, or interpretation differ from the review question?** | Low concern |

##### Reference Standard

| **A. Risk of Bias** | | |
| --- | --- | --- |
| Target condition and reference standard(s) | S. haematobium  urine microscopy | |
| Is the reference standards likely to correctly classify the target condition? | | Unclear |
| Were the reference standard results interpreted without knowledge of the results of the index tests? | | Unclear |
| **Could the reference standard, its conduct, or its interpretation have introduced bias?** | | Low risk |

| **B. Concerns regarding applicability** | |
| --- | --- |
| **Are there concerns that the target condition as defined by the reference standard does not match the question?** | Low concern |

##### Flow and Timing

| **A. Risk of Bias** | | |
| --- | --- | --- |
| Flow and timing |  | |
| Was there an appropriate interval between index test and reference standard? | | Yes |
| Did all patients receive the same reference standard? | | Yes |
| Were all patients included in the analysis? | | Yes |
| **Could the patient flow have introduced bias?** | | Low risk |

##### Notes

| **Notes** |
| --- |

#### Polman 1995

##### Patient Selection

| **A. Risk of Bias** | | |
| --- | --- | --- |
| Patient Sampling | cross-sectional study ; random sampling | |
| Was a consecutive or random sample of patients enrolled? | | Yes |
| Was a case-control design avoided? | | Yes |
| Did the study avoid inappropriate exclusions? | | Unclear |
| **Could the selection of patients have introduced bias?** | | Low risk |

| **B. Concerns regarding applicability** | | |
| --- | --- | --- |
| Patient characteristics and setting | - 422 individuals aged between 0 and 77 years  - Country: Senegal  - Setting: field study | |
| **Are there concerns that the included patients and setting do not match the review question?** | | Low concern |

##### Index Test

| Index tests | CAA ELISA Serum  CCA ELISA Serum & Urine |
| --- | --- |

##### All tests

| **A. Risk of Bias** | |
| --- | --- |
| Were the index test results interpreted without knowledge of the results of the reference standard? | Unclear |
| If a threshold was used, was it pre-specified? | Unclear |
| **Could the conduct or interpretation of the index test have introduced bias?** | Low risk |

| **B. Concerns regarding applicability** | |
| --- | --- |
| **Are there concerns that the index test, its conduct, or interpretation differ from the review question?** | Low concern |

##### Reference Standard

| **A. Risk of Bias** | | |
| --- | --- | --- |
| Target condition and reference standard(s) | S. mansoni  duplicate Kato-Katz | |
| Is the reference standards likely to correctly classify the target condition? | | Unclear |
| Were the reference standard results interpreted without knowledge of the results of the index tests? | | Unclear |
| **Could the reference standard, its conduct, or its interpretation have introduced bias?** | | Low risk |

| **B. Concerns regarding applicability** | |
| --- | --- |
| **Are there concerns that the target condition as defined by the reference standard does not match the question?** | Low concern |

##### Flow and Timing

| **A. Risk of Bias** | | |
| --- | --- | --- |
| Flow and timing |  | |
| Was there an appropriate interval between index test and reference standard? | | Yes |
| Did all patients receive the same reference standard? | | Yes |
| Were all patients included in the analysis? | | Unclear |
| **Could the patient flow have introduced bias?** | | Low risk |

##### Notes

| **Notes** |
| --- |

#### Pugh 1980

##### Patient Selection

| **A. Risk of Bias** | | |
| --- | --- | --- |
| Patient Sampling | cross-sectional study | |
| Was a consecutive or random sample of patients enrolled? | | Unclear |
| Was a case-control design avoided? | | Yes |
| Did the study avoid inappropriate exclusions? | | Unclear |
| **Could the selection of patients have introduced bias?** | | Low risk |

| **B. Concerns regarding applicability** | | |
| --- | --- | --- |
| Patient characteristics and setting | - 5367 males from 3 villages aged between 5 and 25 years  - Country: Nigeria  - Setting: field study | |
| **Are there concerns that the included patients and setting do not match the review question?** | | Low concern |

##### Index Test

| Index tests | micro-haematuria & proteinuria by urine reagent strip (Labstix) |
| --- | --- |

##### All tests

| **A. Risk of Bias** | |
| --- | --- |
| Were the index test results interpreted without knowledge of the results of the reference standard? | Unclear |
| If a threshold was used, was it pre-specified? | Yes |
| **Could the conduct or interpretation of the index test have introduced bias?** | Low risk |

| **B. Concerns regarding applicability** | |
| --- | --- |
| **Are there concerns that the index test, its conduct, or interpretation differ from the review question?** | Low concern |

##### Reference Standard

| **A. Risk of Bias** | | |
| --- | --- | --- |
| Target condition and reference standard(s) | s. haematobium  urine microscopy | |
| Is the reference standards likely to correctly classify the target condition? | | Unclear |
| Were the reference standard results interpreted without knowledge of the results of the index tests? | | Unclear |
| **Could the reference standard, its conduct, or its interpretation have introduced bias?** | | Low risk |

| **B. Concerns regarding applicability** | |
| --- | --- |
| **Are there concerns that the target condition as defined by the reference standard does not match the question?** | Low concern |

##### Flow and Timing

| **A. Risk of Bias** | | |
| --- | --- | --- |
| Flow and timing |  | |
| Was there an appropriate interval between index test and reference standard? | | Unclear |
| Did all patients receive the same reference standard? | | Yes |
| Were all patients included in the analysis? | | No |
| **Could the patient flow have introduced bias?** | | Low risk |

##### Notes

| **Notes** |
| --- |

#### Rasendramino 1998

##### Patient Selection

| **A. Risk of Bias** | | |
| --- | --- | --- |
| Patient Sampling | cross-sectional study | |
| Was a consecutive or random sample of patients enrolled? | | Yes |
| Was a case-control design avoided? | | Yes |
| Did the study avoid inappropriate exclusions? | | Unclear |
| **Could the selection of patients have introduced bias?** | | Low risk |

| **B. Concerns regarding applicability** | | |
| --- | --- | --- |
| Patient characteristics and setting | - 574 patients older than 5 years  - Country: Madagascar  - Setting: field study | |
| **Are there concerns that the included patients and setting do not match the review question?** | | Low concern |

##### Index Test

| Index tests | Micro-haematuria, Proteinuria & Leukocyturia by urine reagent strip (Nephur 7) |
| --- | --- |

##### All tests

| **A. Risk of Bias** | |
| --- | --- |
| Were the index test results interpreted without knowledge of the results of the reference standard? | Unclear |
| If a threshold was used, was it pre-specified? | Unclear |
| **Could the conduct or interpretation of the index test have introduced bias?** | Low risk |

| **B. Concerns regarding applicability** | |
| --- | --- |
| **Are there concerns that the index test, its conduct, or interpretation differ from the review question?** | Low concern |

##### Reference Standard

| **A. Risk of Bias** | | |
| --- | --- | --- |
| Target condition and reference standard(s) | S. haematobium  Urine microscopy | |
| Is the reference standards likely to correctly classify the target condition? | | Unclear |
| Were the reference standard results interpreted without knowledge of the results of the index tests? | | Unclear |
| **Could the reference standard, its conduct, or its interpretation have introduced bias?** | | Low risk |

| **B. Concerns regarding applicability** | |
| --- | --- |
| **Are there concerns that the target condition as defined by the reference standard does not match the question?** | Low concern |

##### Flow and Timing

| **A. Risk of Bias** | | |
| --- | --- | --- |
| Flow and timing |  | |
| Was there an appropriate interval between index test and reference standard? | | Yes |
| Did all patients receive the same reference standard? | | Yes |
| Were all patients included in the analysis? | | Yes |
| **Could the patient flow have introduced bias?** | | Low risk |

##### Notes

| **Notes** |
| --- |

#### Robinson 2009

##### Patient Selection

| **A. Risk of Bias** | | |
| --- | --- | --- |
| Patient Sampling | quasi-random two-stage cluster sampling method ; random selection within communities | |
| Was a consecutive or random sample of patients enrolled? | | Unclear |
| Was a case-control design avoided? | | No |
| Did the study avoid inappropriate exclusions? | | Unclear |
| **Could the selection of patients have introduced bias?** | | Low risk |

| **B. Concerns regarding applicability** | | |
| --- | --- | --- |
| Patient characteristics and setting | - 4901 children from 74 communities (aged between 5 and 16 years)  - Country: Southern Sudan  - Period: February to May 2009 | |
| **Are there concerns that the included patients and setting do not match the review question?** | | Low concern |

##### Index Test

| Index tests | haematuria reagent strips (Hemastix) |
| --- | --- |

##### All tests

| **A. Risk of Bias** | |
| --- | --- |
| Were the index test results interpreted without knowledge of the results of the reference standard? | Yes |
| If a threshold was used, was it pre-specified? | Unclear |
| **Could the conduct or interpretation of the index test have introduced bias?** | Low risk |

| **B. Concerns regarding applicability** | |
| --- | --- |
| **Are there concerns that the index test, its conduct, or interpretation differ from the review question?** | Low concern |

##### Reference Standard

| **A. Risk of Bias** | | |
| --- | --- | --- |
| Target condition and reference standard(s) | S. haematobium  Urine microscopy | |
| Is the reference standards likely to correctly classify the target condition? | | Unclear |
| Were the reference standard results interpreted without knowledge of the results of the index tests? | | Unclear |
| **Could the reference standard, its conduct, or its interpretation have introduced bias?** | | Unclear risk |

| **B. Concerns regarding applicability** | |
| --- | --- |
| **Are there concerns that the target condition as defined by the reference standard does not match the question?** | Low concern |

##### Flow and Timing

| **A. Risk of Bias** | | |
| --- | --- | --- |
| Flow and timing |  | |
| Was there an appropriate interval between index test and reference standard? | | Yes |
| Did all patients receive the same reference standard? | | Yes |
| Were all patients included in the analysis? | | Yes |
| **Could the patient flow have introduced bias?** | | Low risk |

##### Notes

| **Notes** |
| --- |

#### Rollinson 2005

##### Patient Selection

| **A. Risk of Bias** | | |
| --- | --- | --- |
| Patient Sampling | cross-sectional study ; random sampling | |
| Was a consecutive or random sample of patients enrolled? | | Yes |
| Was a case-control design avoided? | | Yes |
| Did the study avoid inappropriate exclusions? | | Yes |
| **Could the selection of patients have introduced bias?** | | Low risk |

| **B. Concerns regarding applicability** | | |
| --- | --- | --- |
| Patient characteristics and setting | - 280 school children from 2 schools aged between 10 and 22 years  - Country: Tanzania  - Setting: field study | |
| **Are there concerns that the included patients and setting do not match the review question?** | | Low concern |

##### Index Test

| Index tests | micro-haematuria ny urine reagent strip (Hemastix) |
| --- | --- |

##### All tests

| **A. Risk of Bias** | |
| --- | --- |
| Were the index test results interpreted without knowledge of the results of the reference standard? | Unclear |
| If a threshold was used, was it pre-specified? | No |
| **Could the conduct or interpretation of the index test have introduced bias?** | Low risk |

| **B. Concerns regarding applicability** | |
| --- | --- |
| **Are there concerns that the index test, its conduct, or interpretation differ from the review question?** | Low concern |

##### Reference Standard

| **A. Risk of Bias** | | |
| --- | --- | --- |
| Target condition and reference standard(s) | S. haematobium  urine microscopy | |
| Is the reference standards likely to correctly classify the target condition? | | Unclear |
| Were the reference standard results interpreted without knowledge of the results of the index tests? | | Unclear |
| **Could the reference standard, its conduct, or its interpretation have introduced bias?** | | Unclear risk |

| **B. Concerns regarding applicability** | |
| --- | --- |
| **Are there concerns that the target condition as defined by the reference standard does not match the question?** | Low concern |

##### Flow and Timing

| **A. Risk of Bias** | | |
| --- | --- | --- |
| Flow and timing |  | |
| Was there an appropriate interval between index test and reference standard? | | Yes |
| Did all patients receive the same reference standard? | | Yes |
| Were all patients included in the analysis? | | No |
| **Could the patient flow have introduced bias?** | | Low risk |

##### Notes

| **Notes** |
| --- |

#### Sarda 1986

##### Patient Selection

| **A. Risk of Bias** | | |
| --- | --- | --- |
| Patient Sampling | cross-sectional study | |
| Was a consecutive or random sample of patients enrolled? | | Unclear |
| Was a case-control design avoided? | | Yes |
| Did the study avoid inappropriate exclusions? | | Yes |
| **Could the selection of patients have introduced bias?** | | Low risk |

| **B. Concerns regarding applicability** | | |
| --- | --- | --- |
| Patient characteristics and setting | - 1300 school children aged between 6 and 19 years  - Country: Tanzania  - Setting: field study | |
| **Are there concerns that the included patients and setting do not match the review question?** | | Low concern |

##### Index Test

| Index tests | Micro-haematuria & Proteinuria by urine reagent strip (N-Multistix) |
| --- | --- |

##### All tests

| **A. Risk of Bias** | |
| --- | --- |
| Were the index test results interpreted without knowledge of the results of the reference standard? | Unclear |
| If a threshold was used, was it pre-specified? | Unclear |
| **Could the conduct or interpretation of the index test have introduced bias?** | Low risk |

| **B. Concerns regarding applicability** | |
| --- | --- |
| **Are there concerns that the index test, its conduct, or interpretation differ from the review question?** | Low concern |

##### Reference Standard

| **A. Risk of Bias** | | |
| --- | --- | --- |
| Target condition and reference standard(s) | S. haematobium  Urine microscopy | |
| Is the reference standards likely to correctly classify the target condition? | | Unclear |
| Were the reference standard results interpreted without knowledge of the results of the index tests? | | Yes |
| **Could the reference standard, its conduct, or its interpretation have introduced bias?** | | Low risk |

| **B. Concerns regarding applicability** | |
| --- | --- |
| **Are there concerns that the target condition as defined by the reference standard does not match the question?** | Low concern |

##### Flow and Timing

| **A. Risk of Bias** | | |
| --- | --- | --- |
| Flow and timing |  | |
| Was there an appropriate interval between index test and reference standard? | | Unclear |
| Did all patients receive the same reference standard? | | Yes |
| Were all patients included in the analysis? | | Yes |
| **Could the patient flow have introduced bias?** | | Low risk |

##### Notes

| **Notes** |
| --- |

#### Savioli 1990

##### Patient Selection

| **A. Risk of Bias** | | |
| --- | --- | --- |
| Patient Sampling | cross-sectional study | |
| Was a consecutive or random sample of patients enrolled? | | Unclear |
| Was a case-control design avoided? | | Yes |
| Did the study avoid inappropriate exclusions? | | Unclear |
| **Could the selection of patients have introduced bias?** | | Low risk |

| **B. Concerns regarding applicability** | | |
| --- | --- | --- |
| Patient characteristics and setting | - 879 children aged between 5 and 19 years  - Country: Tanzania  - Setting: field study | |
| **Are there concerns that the included patients and setting do not match the review question?** | | Low concern |

##### Index Test

| Index tests | micro-haematuria by urine reagent strip (Hemastix) |
| --- | --- |

##### All tests

| **A. Risk of Bias** | |
| --- | --- |
| Were the index test results interpreted without knowledge of the results of the reference standard? | Unclear |
| If a threshold was used, was it pre-specified? | Unclear |
| **Could the conduct or interpretation of the index test have introduced bias?** | Low risk |

| **B. Concerns regarding applicability** | |
| --- | --- |
| **Are there concerns that the index test, its conduct, or interpretation differ from the review question?** | Low concern |

##### Reference Standard

| **A. Risk of Bias** | | |
| --- | --- | --- |
| Target condition and reference standard(s) | S. haematobium  urine microscopy | |
| Is the reference standards likely to correctly classify the target condition? | | Unclear |
| Were the reference standard results interpreted without knowledge of the results of the index tests? | | Unclear |
| **Could the reference standard, its conduct, or its interpretation have introduced bias?** | | Low risk |

| **B. Concerns regarding applicability** | |
| --- | --- |
| **Are there concerns that the target condition as defined by the reference standard does not match the question?** | Low concern |

##### Flow and Timing

| **A. Risk of Bias** | | |
| --- | --- | --- |
| Flow and timing |  | |
| Was there an appropriate interval between index test and reference standard? | | Yes |
| Did all patients receive the same reference standard? | | Yes |
| Were all patients included in the analysis? | | No |
| **Could the patient flow have introduced bias?** | | Low risk |

##### Notes

| **Notes** |
| --- |

#### Sellin 1982

##### Patient Selection

| **A. Risk of Bias** | | |
| --- | --- | --- |
| Patient Sampling | cross-sectional study | |
| Was a consecutive or random sample of patients enrolled? | | Unclear |
| Was a case-control design avoided? | | Yes |
| Did the study avoid inappropriate exclusions? | | Unclear |
| **Could the selection of patients have introduced bias?** | | Low risk |

| **B. Concerns regarding applicability** | | |
| --- | --- | --- |
| Patient characteristics and setting | - 1162 participants from a high endemic village  - Country: Burkina Faso  - Setting: field study | |
| **Are there concerns that the included patients and setting do not match the review question?** | | Low concern |

##### Index Test

| Index tests | micro-haematuria & proteinuria by urine reagent strips |
| --- | --- |

##### All tests

| **A. Risk of Bias** | |
| --- | --- |
| Were the index test results interpreted without knowledge of the results of the reference standard? | Unclear |
| If a threshold was used, was it pre-specified? | Yes |
| **Could the conduct or interpretation of the index test have introduced bias?** | Low risk |

| **B. Concerns regarding applicability** | |
| --- | --- |
| **Are there concerns that the index test, its conduct, or interpretation differ from the review question?** | Low concern |

##### Reference Standard

| **A. Risk of Bias** | | |
| --- | --- | --- |
| Target condition and reference standard(s) | S. haematobium  urine microscopy | |
| Is the reference standards likely to correctly classify the target condition? | | No |
| Were the reference standard results interpreted without knowledge of the results of the index tests? | | Unclear |
| **Could the reference standard, its conduct, or its interpretation have introduced bias?** | | Low risk |

| **B. Concerns regarding applicability** | |
| --- | --- |
| **Are there concerns that the target condition as defined by the reference standard does not match the question?** | Low concern |

##### Flow and Timing

| **A. Risk of Bias** | | |
| --- | --- | --- |
| Flow and timing |  | |
| Was there an appropriate interval between index test and reference standard? | | Unclear |
| Did all patients receive the same reference standard? | | Yes |
| Were all patients included in the analysis? | | Yes |
| **Could the patient flow have introduced bias?** | | Low risk |

##### Notes

| **Notes** |
| --- |

#### Shane 2011

##### Patient Selection

| **A. Risk of Bias** | | |
| --- | --- | --- |
| Patient Sampling | cross-sectional survey; no random selection | |
| Was a consecutive or random sample of patients enrolled? | | Yes |
| Was a case-control design avoided? | | Yes |
| Did the study avoid inappropriate exclusions? | | Yes |
| **Could the selection of patients have introduced bias?** | | Low risk |

| **B. Concerns regarding applicability** | | |
| --- | --- | --- |
| Patient characteristics and setting | Species: S. mansoni  Country: Kenya  Sample size:  Age range: 1 - 15 y  Participants: children  Setting: field study  Praziquantel before the study: no mass drug administration to treat schistosomiasis in this area | |
| **Are there concerns that the included patients and setting do not match the review question?** | | Low concern |

##### Index Test

| Index tests | CCA1 & CCA2 |
| --- | --- |

##### All tests

| **A. Risk of Bias** | |
| --- | --- |
| Were the index test results interpreted without knowledge of the results of the reference standard? | Yes |
| If a threshold was used, was it pre-specified? | Yes |
| **Could the conduct or interpretation of the index test have introduced bias?** | Low risk |

| **B. Concerns regarding applicability** | |
| --- | --- |
| **Are there concerns that the index test, its conduct, or interpretation differ from the review question?** | Low concern |

##### Reference Standard

| **A. Risk of Bias** | | |
| --- | --- | --- |
| Target condition and reference standard(s) | Duplicate Kato Katz smear | |
| Is the reference standards likely to correctly classify the target condition? | | No |
| Were the reference standard results interpreted without knowledge of the results of the index tests? | | Yes |
| **Could the reference standard, its conduct, or its interpretation have introduced bias?** | | Low risk |

| **B. Concerns regarding applicability** | |
| --- | --- |
| **Are there concerns that the target condition as defined by the reference standard does not match the question?** | Low concern |

##### Flow and Timing

| **A. Risk of Bias** | | |
| --- | --- | --- |
| Flow and timing |  | |
| Was there an appropriate interval between index test and reference standard? | | Yes |
| Did all patients receive the same reference standard? | | Yes |
| Were all patients included in the analysis? | | Yes |
| **Could the patient flow have introduced bias?** | | Low risk |

##### Notes

| **Notes** |
| --- |

#### Shaw 1998

##### Patient Selection

| **A. Risk of Bias** | | |
| --- | --- | --- |
| Patient Sampling | cohort study , random sampling | |
| Was a consecutive or random sample of patients enrolled? | | Yes |
| Was a case-control design avoided? | | Yes |
| Did the study avoid inappropriate exclusions? | | Unclear |
| **Could the selection of patients have introduced bias?** | | Low risk |

| **B. Concerns regarding applicability** | | |
| --- | --- | --- |
| Patient characteristics and setting | - 857 participants from 4 villages of all ages  - Country: Senegal  - Setting: field study | |
| **Are there concerns that the included patients and setting do not match the review question?** | | Low concern |

##### Index Test

| Index tests | micro-haematuria by urine reagent strip |
| --- | --- |

##### All tests

| **A. Risk of Bias** | |
| --- | --- |
| Were the index test results interpreted without knowledge of the results of the reference standard? | Unclear |
| If a threshold was used, was it pre-specified? | Unclear |
| **Could the conduct or interpretation of the index test have introduced bias?** | Low risk |

| **B. Concerns regarding applicability** | |
| --- | --- |
| **Are there concerns that the index test, its conduct, or interpretation differ from the review question?** | Low concern |

##### Reference Standard

| **A. Risk of Bias** | | |
| --- | --- | --- |
| Target condition and reference standard(s) | S. haematobium  urine microscopy | |
| Is the reference standards likely to correctly classify the target condition? | | Unclear |
| Were the reference standard results interpreted without knowledge of the results of the index tests? | | Unclear |
| **Could the reference standard, its conduct, or its interpretation have introduced bias?** | | Unclear risk |

| **B. Concerns regarding applicability** | |
| --- | --- |
| **Are there concerns that the target condition as defined by the reference standard does not match the question?** | Low concern |

##### Flow and Timing

| **A. Risk of Bias** | | |
| --- | --- | --- |
| Flow and timing |  | |
| Was there an appropriate interval between index test and reference standard? | | Yes |
| Did all patients receive the same reference standard? | | Yes |
| Were all patients included in the analysis? | | Yes |
| **Could the patient flow have introduced bias?** | | Low risk |

##### Notes

| **Notes** |
| --- |

#### Sheele 2013

##### Patient Selection

| **A. Risk of Bias** | | |
| --- | --- | --- |
| Patient Sampling | cross-sectional survey; no random selection | |
| Was a consecutive or random sample of patients enrolled? | | Yes |
| Was a case-control design avoided? | | Yes |
| Did the study avoid inappropriate exclusions? | | Yes |
| **Could the selection of patients have introduced bias?** | | Low risk |

| **B. Concerns regarding applicability** | | |
| --- | --- | --- |
| Patient characteristics and setting | Species: S. haematobium  Country: Kenya  Sample size: 160  Age range: 8-17y  Participants: schoolchildren  Setting: fiels study  Praziquantel before the study: | |
| **Are there concerns that the included patients and setting do not match the review question?** | | Low concern |

##### Index Test

| Index tests | RDT-Sh |
| --- | --- |

##### All tests

| **A. Risk of Bias** | |
| --- | --- |
| Were the index test results interpreted without knowledge of the results of the reference standard? | Yes |
| If a threshold was used, was it pre-specified? | Yes |
| **Could the conduct or interpretation of the index test have introduced bias?** | Low risk |

| **B. Concerns regarding applicability** | |
| --- | --- |
| **Are there concerns that the index test, its conduct, or interpretation differ from the review question?** | Low concern |

##### Reference Standard

| **A. Risk of Bias** | | |
| --- | --- | --- |
| Target condition and reference standard(s) | urine microscopy | |
| Is the reference standards likely to correctly classify the target condition? | | No |
| Were the reference standard results interpreted without knowledge of the results of the index tests? | | Yes |
| **Could the reference standard, its conduct, or its interpretation have introduced bias?** | | Low risk |

| **B. Concerns regarding applicability** | |
| --- | --- |
| **Are there concerns that the target condition as defined by the reference standard does not match the question?** | Low concern |

##### Flow and Timing

| **A. Risk of Bias** | | |
| --- | --- | --- |
| Flow and timing |  | |
| Was there an appropriate interval between index test and reference standard? | | Yes |
| Did all patients receive the same reference standard? | | Yes |
| Were all patients included in the analysis? | | Yes |
| **Could the patient flow have introduced bias?** | | Low risk |

##### Notes

| **Notes** |
| --- |

#### Song 2018

##### Patient Selection

| **A. Risk of Bias** | | |
| --- | --- | --- |
| Patient Sampling | cross-sectional study | |
| Was a consecutive or random sample of patients enrolled? | | Yes |
| Was a case-control design avoided? | | Unclear |
| Did the study avoid inappropriate exclusions? | | Unclear |
| **Could the selection of patients have introduced bias?** | | Low risk |

| **B. Concerns regarding applicability** | | |
| --- | --- | --- |
| Patient characteristics and setting | - 149 patients (mostly children aged under 16 years)  - Country: Sudan  - Setting: field study | |
| **Are there concerns that the included patients and setting do not match the review question?** | | Low concern |

##### Index Test

| Index tests | ELISA-SEA |
| --- | --- |

##### All tests

| **A. Risk of Bias** | |
| --- | --- |
| Were the index test results interpreted without knowledge of the results of the reference standard? | Unclear |
| If a threshold was used, was it pre-specified? | Yes |
| **Could the conduct or interpretation of the index test have introduced bias?** | Low risk |

| **B. Concerns regarding applicability** | |
| --- | --- |
| **Are there concerns that the index test, its conduct, or interpretation differ from the review question?** | Low concern |

##### Reference Standard

| **A. Risk of Bias** | | |
| --- | --- | --- |
| Target condition and reference standard(s) | S. haematobium  Urine microscopy | |
| Is the reference standards likely to correctly classify the target condition? | | Unclear |
| Were the reference standard results interpreted without knowledge of the results of the index tests? | | Unclear |
| **Could the reference standard, its conduct, or its interpretation have introduced bias?** | | Low risk |

| **B. Concerns regarding applicability** | |
| --- | --- |
| **Are there concerns that the target condition as defined by the reference standard does not match the question?** | Low concern |

##### Flow and Timing

| **A. Risk of Bias** | | |
| --- | --- | --- |
| Flow and timing |  | |
| Was there an appropriate interval between index test and reference standard? | | Yes |
| Did all patients receive the same reference standard? | | Yes |
| Were all patients included in the analysis? | | No |
| **Could the patient flow have introduced bias?** | | Low risk |

##### Notes

| **Notes** |
| --- |

#### Sousa 2020

##### Patient Selection

| **A. Risk of Bias** | | |
| --- | --- | --- |
| Patient Sampling | cross-sectional study | |
| Was a consecutive or random sample of patients enrolled? | | Yes |
| Was a case-control design avoided? | | Unclear |
| Did the study avoid inappropriate exclusions? | | Yes |
| **Could the selection of patients have introduced bias?** | | Low risk |

| **B. Concerns regarding applicability** | | |
| --- | --- | --- |
| Patient characteristics and setting | - 217 patients from village in Amazona region (111 male, 106 female)  - Country: Brazil  - Setting: field study | |
| **Are there concerns that the included patients and setting do not match the review question?** | | Low concern |

##### Index Test

| Index tests | POC-CCA |
| --- | --- |

##### All tests

| **A. Risk of Bias** | |
| --- | --- |
| Were the index test results interpreted without knowledge of the results of the reference standard? | Yes |
| If a threshold was used, was it pre-specified? | Unclear |
| **Could the conduct or interpretation of the index test have introduced bias?** | Low risk |

| **B. Concerns regarding applicability** | |
| --- | --- |
| **Are there concerns that the index test, its conduct, or interpretation differ from the review question?** | Low concern |

##### Reference Standard

| **A. Risk of Bias** | | |
| --- | --- | --- |
| Target condition and reference standard(s) | - S. mansoni  - 1KK, 2KK, 16KK, Helmintex | |
| Is the reference standards likely to correctly classify the target condition? | | Unclear |
| Were the reference standard results interpreted without knowledge of the results of the index tests? | | Yes |
| **Could the reference standard, its conduct, or its interpretation have introduced bias?** | | Low risk |

| **B. Concerns regarding applicability** | |
| --- | --- |
| **Are there concerns that the target condition as defined by the reference standard does not match the question?** | Low concern |

##### Flow and Timing

| **A. Risk of Bias** | | |
| --- | --- | --- |
| Flow and timing |  | |
| Was there an appropriate interval between index test and reference standard? | | Yes |
| Did all patients receive the same reference standard? | | Yes |
| Were all patients included in the analysis? | | No |
| **Could the patient flow have introduced bias?** | | Low risk |

##### Notes

| **Notes** |
| --- |

#### Standley 2010

##### Patient Selection

| **A. Risk of Bias** | | |
| --- | --- | --- |
| Patient Sampling | cross-sectional study | |
| Was a consecutive or random sample of patients enrolled? | | Unclear |
| Was a case-control design avoided? | | Yes |
| Did the study avoid inappropriate exclusions? | | Yes |
| **Could the selection of patients have introduced bias?** | | Low risk |

| **B. Concerns regarding applicability** | | |
| --- | --- | --- |
| Patient characteristics and setting | - 171 school children from 11 schools  - Tanzania & Kenya | |
| **Are there concerns that the included patients and setting do not match the review question?** | | Low concern |

##### Index Test

| Index tests | CCA-POC |
| --- | --- |

##### All tests

| **A. Risk of Bias** | |
| --- | --- |
| Were the index test results interpreted without knowledge of the results of the reference standard? | Yes |
| If a threshold was used, was it pre-specified? | Unclear |
| **Could the conduct or interpretation of the index test have introduced bias?** | Low risk |

| **B. Concerns regarding applicability** | |
| --- | --- |
| **Are there concerns that the index test, its conduct, or interpretation differ from the review question?** | Low concern |

##### Reference Standard

| **A. Risk of Bias** | | |
| --- | --- | --- |
| Target condition and reference standard(s) | S. mansoni  duplicate Kato-Katz | |
| Is the reference standards likely to correctly classify the target condition? | | Unclear |
| Were the reference standard results interpreted without knowledge of the results of the index tests? | | Yes |
| **Could the reference standard, its conduct, or its interpretation have introduced bias?** | | Low risk |

| **B. Concerns regarding applicability** | |
| --- | --- |
| **Are there concerns that the target condition as defined by the reference standard does not match the question?** | Low concern |

##### Flow and Timing

| **A. Risk of Bias** | | |
| --- | --- | --- |
| Flow and timing |  | |
| Was there an appropriate interval between index test and reference standard? | | Yes |
| Did all patients receive the same reference standard? | | Yes |
| Were all patients included in the analysis? | | No |
| **Could the patient flow have introduced bias?** | | Low risk |

##### Notes

| **Notes** |
| --- |

#### Stephenson 1984

##### Patient Selection

| **A. Risk of Bias** | | |
| --- | --- | --- |
| Patient Sampling | cross-sectional study | |
| Was a consecutive or random sample of patients enrolled? | | Yes |
| Was a case-control design avoided? | | Yes |
| Did the study avoid inappropriate exclusions? | | Unclear |
| **Could the selection of patients have introduced bias?** | | Low risk |

| **B. Concerns regarding applicability** | | |
| --- | --- | --- |
| Patient characteristics and setting | - 359 schoolchildren from 4 primary schools aged between 6 and 16 years  - Country: Kenya  - Setting: field study | |
| **Are there concerns that the included patients and setting do not match the review question?** | | Low concern |

##### Index Test

| Index tests | Microhaematuria & Proteinuria by urine reagent strip |
| --- | --- |

##### All tests

| **A. Risk of Bias** | |
| --- | --- |
| Were the index test results interpreted without knowledge of the results of the reference standard? | Unclear |
| If a threshold was used, was it pre-specified? | Yes |
| **Could the conduct or interpretation of the index test have introduced bias?** | Low risk |

| **B. Concerns regarding applicability** | |
| --- | --- |
| **Are there concerns that the index test, its conduct, or interpretation differ from the review question?** | Low concern |

##### Reference Standard

| **A. Risk of Bias** | | |
| --- | --- | --- |
| Target condition and reference standard(s) | S. haematobium  Urine microscopy | |
| Is the reference standards likely to correctly classify the target condition? | | Unclear |
| Were the reference standard results interpreted without knowledge of the results of the index tests? | | Unclear |
| **Could the reference standard, its conduct, or its interpretation have introduced bias?** | | Low risk |

| **B. Concerns regarding applicability** | |
| --- | --- |
| **Are there concerns that the target condition as defined by the reference standard does not match the question?** | Low concern |

##### Flow and Timing

| **A. Risk of Bias** | | |
| --- | --- | --- |
| Flow and timing |  | |
| Was there an appropriate interval between index test and reference standard? | | Yes |
| Did all patients receive the same reference standard? | | Yes |
| Were all patients included in the analysis? | | Unclear |
| **Could the patient flow have introduced bias?** | | Low risk |

##### Notes

| **Notes** |
| --- |

#### Stothard 2009a

##### Patient Selection

| **A. Risk of Bias** | | |
| --- | --- | --- |
| Patient Sampling | cross-sectional study | |
| Was a consecutive or random sample of patients enrolled? | | Unclear |
| Was a case-control design avoided? | | Yes |
| Did the study avoid inappropriate exclusions? | | Yes |
| **Could the selection of patients have introduced bias?** | | Low risk |

| **B. Concerns regarding applicability** | | |
| --- | --- | --- |
| Patient characteristics and setting | - 150 school children aged between 8 and 14 years (75 girls, 75 boys)  - Country: Tanzania  - Setting: field study | |
| **Are there concerns that the included patients and setting do not match the review question?** | | Low concern |

##### Index Test

| Index tests | CCA & SEA-ELISA |
| --- | --- |

##### All tests

| **A. Risk of Bias** | |
| --- | --- |
| Were the index test results interpreted without knowledge of the results of the reference standard? | Unclear |
| If a threshold was used, was it pre-specified? | Unclear |
| **Could the conduct or interpretation of the index test have introduced bias?** | Low risk |

| **B. Concerns regarding applicability** | |
| --- | --- |
| **Are there concerns that the index test, its conduct, or interpretation differ from the review question?** | Low concern |

##### Reference Standard

| **A. Risk of Bias** | | |
| --- | --- | --- |
| Target condition and reference standard(s) | S. haematobium  urine microscopy | |
| Is the reference standards likely to correctly classify the target condition? | | Unclear |
| Were the reference standard results interpreted without knowledge of the results of the index tests? | | Unclear |
| **Could the reference standard, its conduct, or its interpretation have introduced bias?** | | Unclear risk |

| **B. Concerns regarding applicability** | |
| --- | --- |
| **Are there concerns that the target condition as defined by the reference standard does not match the question?** | Low concern |

##### Flow and Timing

| **A. Risk of Bias** | | |
| --- | --- | --- |
| Flow and timing |  | |
| Was there an appropriate interval between index test and reference standard? | | Yes |
| Did all patients receive the same reference standard? | | Yes |
| Were all patients included in the analysis? | | Yes |
| **Could the patient flow have introduced bias?** | | Low risk |

##### Notes

| **Notes** |
| --- |

#### Stothard 2009b

##### Patient Selection

| **A. Risk of Bias** | | |
| --- | --- | --- |
| Patient Sampling | cross-sectional study | |
| Was a consecutive or random sample of patients enrolled? | | Unclear |
| Was a case-control design avoided? | | Yes |
| Did the study avoid inappropriate exclusions? | | Unclear |
| **Could the selection of patients have introduced bias?** | | Low risk |

| **B. Concerns regarding applicability** | | |
| --- | --- | --- |
| Patient characteristics and setting | - 66 school children aged between 9 and 15 years  - country: Tanzania  - Setting: field study | |
| **Are there concerns that the included patients and setting do not match the review question?** | | Low concern |

##### Index Test

| Index tests | urine reagent strips (Hemastix) |
| --- | --- |

##### All tests

| **A. Risk of Bias** | |
| --- | --- |
| Were the index test results interpreted without knowledge of the results of the reference standard? | Unclear |
| If a threshold was used, was it pre-specified? | Unclear |
| **Could the conduct or interpretation of the index test have introduced bias?** | Low risk |

| **B. Concerns regarding applicability** | |
| --- | --- |
| **Are there concerns that the index test, its conduct, or interpretation differ from the review question?** | Low concern |

##### Reference Standard

| **A. Risk of Bias** | | |
| --- | --- | --- |
| Target condition and reference standard(s) | S. haematobium  Urine microscopy | |
| Is the reference standards likely to correctly classify the target condition? | | Unclear |
| Were the reference standard results interpreted without knowledge of the results of the index tests? | | Unclear |
| **Could the reference standard, its conduct, or its interpretation have introduced bias?** | | Unclear risk |

| **B. Concerns regarding applicability** | |
| --- | --- |
| **Are there concerns that the target condition as defined by the reference standard does not match the question?** | Low concern |

##### Flow and Timing

| **A. Risk of Bias** | | |
| --- | --- | --- |
| Flow and timing |  | |
| Was there an appropriate interval between index test and reference standard? | | Yes |
| Did all patients receive the same reference standard? | | Yes |
| Were all patients included in the analysis? | | Yes |
| **Could the patient flow have introduced bias?** | | Low risk |

##### Notes

| **Notes** |
| --- |

#### Tanner 1983 (Liberia)

##### Patient Selection

| **A. Risk of Bias** | | |
| --- | --- | --- |
| Patient Sampling | cross-sectional study ; random sampling | |
| Was a consecutive or random sample of patients enrolled? | | Yes |
| Was a case-control design avoided? | | Yes |
| Did the study avoid inappropriate exclusions? | | Yes |
| **Could the selection of patients have introduced bias?** | | Low risk |

| **B. Concerns regarding applicability** | | |
| --- | --- | --- |
| Patient characteristics and setting | - 267 school children from 3 villages aged between 0 and 15 years  - Country: Liberia  - Setting: field study | |
| **Are there concerns that the included patients and setting do not match the review question?** | | Low concern |

##### Index Test

| Index tests | Micro-haematuria & Proteinuria by urine reagent strips |
| --- | --- |

##### All tests

| **A. Risk of Bias** | |
| --- | --- |
| Were the index test results interpreted without knowledge of the results of the reference standard? | Unclear |
| If a threshold was used, was it pre-specified? | Yes |
| **Could the conduct or interpretation of the index test have introduced bias?** | Low risk |

| **B. Concerns regarding applicability** | |
| --- | --- |
| **Are there concerns that the index test, its conduct, or interpretation differ from the review question?** | Low concern |

##### Reference Standard

| **A. Risk of Bias** | | |
| --- | --- | --- |
| Target condition and reference standard(s) | S. haematobium  Urine microscopy | |
| Is the reference standards likely to correctly classify the target condition? | | Unclear |
| Were the reference standard results interpreted without knowledge of the results of the index tests? | | Unclear |
| **Could the reference standard, its conduct, or its interpretation have introduced bias?** | | Low risk |

| **B. Concerns regarding applicability** | |
| --- | --- |
| **Are there concerns that the target condition as defined by the reference standard does not match the question?** | Low concern |

##### Flow and Timing

| **A. Risk of Bias** | | |
| --- | --- | --- |
| Flow and timing |  | |
| Was there an appropriate interval between index test and reference standard? | | Yes |
| Did all patients receive the same reference standard? | | Yes |
| Were all patients included in the analysis? | | Yes |
| **Could the patient flow have introduced bias?** | | Low risk |

##### Notes

| **Notes** |
| --- |

#### Tanner 1983 (Tanzania)

##### Patient Selection

| **A. Risk of Bias** | | |
| --- | --- | --- |
| Patient Sampling | cross-sectional study ; random sampling | |
| Was a consecutive or random sample of patients enrolled? | | Yes |
| Was a case-control design avoided? | | Yes |
| Did the study avoid inappropriate exclusions? | | Yes |
| **Could the selection of patients have introduced bias?** | | Low risk |

| **B. Concerns regarding applicability** | | |
| --- | --- | --- |
| Patient characteristics and setting | - 548 children from single village aged between 0 and 15 years  - Country: Tanzania  - Setting: field study | |
| **Are there concerns that the included patients and setting do not match the review question?** | | Low concern |

##### Index Test

| Index tests | Micro-haematuria & Proteinuria by urine reagent strips |
| --- | --- |

##### All tests

| **A. Risk of Bias** | |
| --- | --- |
| Were the index test results interpreted without knowledge of the results of the reference standard? | Unclear |
| If a threshold was used, was it pre-specified? | Yes |
| **Could the conduct or interpretation of the index test have introduced bias?** | Low risk |

| **B. Concerns regarding applicability** | |
| --- | --- |
| **Are there concerns that the index test, its conduct, or interpretation differ from the review question?** | Low concern |

##### Reference Standard

| **A. Risk of Bias** | | |
| --- | --- | --- |
| Target condition and reference standard(s) | S. haematobium  Urine microscopy | |
| Is the reference standards likely to correctly classify the target condition? | | Unclear |
| Were the reference standard results interpreted without knowledge of the results of the index tests? | | Unclear |
| **Could the reference standard, its conduct, or its interpretation have introduced bias?** | | Low risk |

| **B. Concerns regarding applicability** | |
| --- | --- |
| **Are there concerns that the target condition as defined by the reference standard does not match the question?** | Low concern |

##### Flow and Timing

| **A. Risk of Bias** | | |
| --- | --- | --- |
| Flow and timing |  | |
| Was there an appropriate interval between index test and reference standard? | | Yes |
| Did all patients receive the same reference standard? | | Yes |
| Were all patients included in the analysis? | | Yes |
| **Could the patient flow have introduced bias?** | | Low risk |

##### Notes

| **Notes** |
| --- |

#### Tchuem Tchuente 2012

##### Patient Selection

| **A. Risk of Bias** | | |
| --- | --- | --- |
| Patient Sampling | cross-sectional survey; no random selection | |
| Was a consecutive or random sample of patients enrolled? | | Yes |
| Was a case-control design avoided? | | Yes |
| Did the study avoid inappropriate exclusions? | | Yes |
| **Could the selection of patients have introduced bias?** | | Low risk |

| **B. Concerns regarding applicability** | | |
| --- | --- | --- |
| Patient characteristics and setting | Species: S. mansoni and S. haematobium  Country: Cameroon  Sample size: 750  Age range: 8–12 y  Participants: schoolchildren  Setting: field study  Praziquantel before the study: | |
| **Are there concerns that the included patients and setting do not match the review question?** | | Low concern |

##### Index Test

| Index tests | CCA1 & CCA2 |
| --- | --- |

##### All tests

| **A. Risk of Bias** | |
| --- | --- |
| Were the index test results interpreted without knowledge of the results of the reference standard? | Yes |
| If a threshold was used, was it pre-specified? | Yes |
| **Could the conduct or interpretation of the index test have introduced bias?** | Low risk |

| **B. Concerns regarding applicability** | |
| --- | --- |
| **Are there concerns that the index test, its conduct, or interpretation differ from the review question?** | Low concern |

##### Reference Standard

| **A. Risk of Bias** | | |
| --- | --- | --- |
| Target condition and reference standard(s) | Duplicate Kato Katz smear ; urine microscopy | |
| Is the reference standards likely to correctly classify the target condition? | | No |
| Were the reference standard results interpreted without knowledge of the results of the index tests? | | Yes |
| **Could the reference standard, its conduct, or its interpretation have introduced bias?** | | Low risk |

| **B. Concerns regarding applicability** | |
| --- | --- |
| **Are there concerns that the target condition as defined by the reference standard does not match the question?** | Low concern |

##### Flow and Timing

| **A. Risk of Bias** | | |
| --- | --- | --- |
| Flow and timing |  | |
| Was there an appropriate interval between index test and reference standard? | | Yes |
| Did all patients receive the same reference standard? | | Yes |
| Were all patients included in the analysis? | | Yes |
| **Could the patient flow have introduced bias?** | | Low risk |

##### Notes

| **Notes** |
| --- |

#### Traore 1998

##### Patient Selection

| **A. Risk of Bias** | | |
| --- | --- | --- |
| Patient Sampling | cross-sectional study | |
| Was a consecutive or random sample of patients enrolled? | | Yes |
| Was a case-control design avoided? | | Yes |
| Did the study avoid inappropriate exclusions? | | Yes |
| **Could the selection of patients have introduced bias?** | | Low risk |

| **B. Concerns regarding applicability** | | |
| --- | --- | --- |
| Patient characteristics and setting | - 1041 village inhabitants older than 2 years  - Country: Mali  - Setting: field study | |
| **Are there concerns that the included patients and setting do not match the review question?** | | Low concern |

##### Index Test

| Index tests | micro-haematuria & proteinuria reagent strips |
| --- | --- |

##### All tests

| **A. Risk of Bias** | |
| --- | --- |
| Were the index test results interpreted without knowledge of the results of the reference standard? | Unclear |
| If a threshold was used, was it pre-specified? | Unclear |
| **Could the conduct or interpretation of the index test have introduced bias?** | Low risk |

| **B. Concerns regarding applicability** | |
| --- | --- |
| **Are there concerns that the index test, its conduct, or interpretation differ from the review question?** | Low concern |

##### Reference Standard

| **A. Risk of Bias** | | |
| --- | --- | --- |
| Target condition and reference standard(s) | S. haematobium  urine microscopy | |
| Is the reference standards likely to correctly classify the target condition? | | Unclear |
| Were the reference standard results interpreted without knowledge of the results of the index tests? | | Yes |
| **Could the reference standard, its conduct, or its interpretation have introduced bias?** | | Low risk |

| **B. Concerns regarding applicability** | |
| --- | --- |
| **Are there concerns that the target condition as defined by the reference standard does not match the question?** | Low concern |

##### Flow and Timing

| **A. Risk of Bias** | | |
| --- | --- | --- |
| Flow and timing |  | |
| Was there an appropriate interval between index test and reference standard? | | Yes |
| Did all patients receive the same reference standard? | | Yes |
| Were all patients included in the analysis? | | No |
| **Could the patient flow have introduced bias?** | | Low risk |

##### Notes

| **Notes** |
| --- |

#### Uga 1989

##### Patient Selection

| **A. Risk of Bias** | | |
| --- | --- | --- |
| Patient Sampling | Unclear | |
| Was a consecutive or random sample of patients enrolled? | | Unclear |
| Was a case-control design avoided? | | Unclear |
| Did the study avoid inappropriate exclusions? | | Unclear |
| **Could the selection of patients have introduced bias?** | | Unclear risk |

| **B. Concerns regarding applicability** | | |
| --- | --- | --- |
| Patient characteristics and setting | - 50 patients  - Country: Kenya | |
| **Are there concerns that the included patients and setting do not match the review question?** | | Unclear concern |

##### Index Test

| Index tests | IgG SEA-ELISA |
| --- | --- |

##### All tests

| **A. Risk of Bias** | |
| --- | --- |
| Were the index test results interpreted without knowledge of the results of the reference standard? | Unclear |
| If a threshold was used, was it pre-specified? | Yes |
| **Could the conduct or interpretation of the index test have introduced bias?** | Low risk |

| **B. Concerns regarding applicability** | |
| --- | --- |
| **Are there concerns that the index test, its conduct, or interpretation differ from the review question?** | Low concern |

##### Reference Standard

| **A. Risk of Bias** | | |
| --- | --- | --- |
| Target condition and reference standard(s) | S. haematobium  Urine microscopy | |
| Is the reference standards likely to correctly classify the target condition? | | Unclear |
| Were the reference standard results interpreted without knowledge of the results of the index tests? | | Unclear |
| **Could the reference standard, its conduct, or its interpretation have introduced bias?** | | Low risk |

| **B. Concerns regarding applicability** | |
| --- | --- |
| **Are there concerns that the target condition as defined by the reference standard does not match the question?** | Low concern |

##### Flow and Timing

| **A. Risk of Bias** | | |
| --- | --- | --- |
| Flow and timing |  | |
| Was there an appropriate interval between index test and reference standard? | | Yes |
| Did all patients receive the same reference standard? | | Yes |
| Were all patients included in the analysis? | | Unclear |
| **Could the patient flow have introduced bias?** | | Low risk |

##### Notes

| **Notes** |
| --- |

#### Ugbomoiko 2009a

##### Patient Selection

| **A. Risk of Bias** | | |
| --- | --- | --- |
| Patient Sampling | cross-sectional survey; no random selection | |
| Was a consecutive or random sample of patients enrolled? | | Yes |
| Was a case-control design avoided? | | Yes |
| Did the study avoid inappropriate exclusions? | | Yes |
| **Could the selection of patients have introduced bias?** | | Low risk |

| **B. Concerns regarding applicability** | | |
| --- | --- | --- |
| Patient characteristics and setting | Species: S. haematobium  Country: Nigeria  Sample size: 566 and 1457  Age range:  Participants: communities  Setting: field study  Praziquantel before the study: | |
| **Are there concerns that the included patients and setting do not match the review question?** | | Low concern |

##### Index Test

| Index tests | haematuria; proteinuria |
| --- | --- |

##### All tests

| **A. Risk of Bias** | |
| --- | --- |
| Were the index test results interpreted without knowledge of the results of the reference standard? | Yes |
| If a threshold was used, was it pre-specified? | Yes |
| **Could the conduct or interpretation of the index test have introduced bias?** | Low risk |

| **B. Concerns regarding applicability** | |
| --- | --- |
| **Are there concerns that the index test, its conduct, or interpretation differ from the review question?** | Low concern |

##### Reference Standard

| **A. Risk of Bias** | | |
| --- | --- | --- |
| Target condition and reference standard(s) | Urine microscopy | |
| Is the reference standards likely to correctly classify the target condition? | | No |
| Were the reference standard results interpreted without knowledge of the results of the index tests? | | Yes |
| **Could the reference standard, its conduct, or its interpretation have introduced bias?** | | Unclear risk |

| **B. Concerns regarding applicability** | |
| --- | --- |
| **Are there concerns that the target condition as defined by the reference standard does not match the question?** | Low concern |

##### Flow and Timing

| **A. Risk of Bias** | | |
| --- | --- | --- |
| Flow and timing |  | |
| Was there an appropriate interval between index test and reference standard? | | Yes |
| Did all patients receive the same reference standard? | | Yes |
| Were all patients included in the analysis? | | Yes |
| **Could the patient flow have introduced bias?** | | Low risk |

##### Notes

| **Notes** |
| --- |

#### Ugbomoiko 2009b

##### Patient Selection

| **A. Risk of Bias** | | |
| --- | --- | --- |
| Patient Sampling | cross-sectional study | |
| Was a consecutive or random sample of patients enrolled? | | Unclear |
| Was a case-control design avoided? | | Yes |
| Did the study avoid inappropriate exclusions? | | Unclear |
| **Could the selection of patients have introduced bias?** | | Low risk |

| **B. Concerns regarding applicability** | | |
| --- | --- | --- |
| Patient characteristics and setting | - 447 school children aged between 3 and 17 years  - except girls with menstruation during previous 5 days  - Country: Nigeria | |
| **Are there concerns that the included patients and setting do not match the review question?** | | Low concern |

##### Index Test

| Index tests | microhaematuria & proteinuria (by reagent strip) |
| --- | --- |

##### All tests

| **A. Risk of Bias** | |
| --- | --- |
| Were the index test results interpreted without knowledge of the results of the reference standard? | Yes |
| If a threshold was used, was it pre-specified? | Unclear |
| **Could the conduct or interpretation of the index test have introduced bias?** | Low risk |

| **B. Concerns regarding applicability** | |
| --- | --- |
| **Are there concerns that the index test, its conduct, or interpretation differ from the review question?** | Low concern |

##### Reference Standard

| **A. Risk of Bias** | | |
| --- | --- | --- |
| Target condition and reference standard(s) | S. haematobium  Urine microscopy | |
| Is the reference standards likely to correctly classify the target condition? | | Unclear |
| Were the reference standard results interpreted without knowledge of the results of the index tests? | | Yes |
| **Could the reference standard, its conduct, or its interpretation have introduced bias?** | | Unclear risk |

| **B. Concerns regarding applicability** | |
| --- | --- |
| **Are there concerns that the target condition as defined by the reference standard does not match the question?** | Low concern |

##### Flow and Timing

| **A. Risk of Bias** | | |
| --- | --- | --- |
| Flow and timing |  | |
| Was there an appropriate interval between index test and reference standard? | | Yes |
| Did all patients receive the same reference standard? | | Yes |
| Were all patients included in the analysis? | | No |
| **Could the patient flow have introduced bias?** | | Low risk |

##### Notes

| **Notes** |
| --- |

#### Van Lieshout 1995

##### Patient Selection

| **A. Risk of Bias** | | |
| --- | --- | --- |
| Patient Sampling | cross-sectional study | |
| Was a consecutive or random sample of patients enrolled? | | Unclear |
| Was a case-control design avoided? | | Yes |
| Did the study avoid inappropriate exclusions? | | Unclear |
| **Could the selection of patients have introduced bias?** | | Low risk |

| **B. Concerns regarding applicability** | | |
| --- | --- | --- |
| Patient characteristics and setting | - 389 inhabitants of a village aged older than 1 year  - Country: Surinam  - Setting: field study | |
| **Are there concerns that the included patients and setting do not match the review question?** | | Low concern |

##### Index Test

| Index tests | urine-CAA & urine-CCA  serum-CAA & serum-CCA |
| --- | --- |

##### All tests

| **A. Risk of Bias** | |
| --- | --- |
| Were the index test results interpreted without knowledge of the results of the reference standard? | Unclear |
| If a threshold was used, was it pre-specified? | Yes |
| **Could the conduct or interpretation of the index test have introduced bias?** | Low risk |

| **B. Concerns regarding applicability** | |
| --- | --- |
| **Are there concerns that the index test, its conduct, or interpretation differ from the review question?** | Low concern |

##### Reference Standard

| **A. Risk of Bias** | | |
| --- | --- | --- |
| Target condition and reference standard(s) | S. mansoni  duplicate Kato-Katz | |
| Is the reference standards likely to correctly classify the target condition? | | Unclear |
| Were the reference standard results interpreted without knowledge of the results of the index tests? | | Unclear |
| **Could the reference standard, its conduct, or its interpretation have introduced bias?** | | Low risk |

| **B. Concerns regarding applicability** | |
| --- | --- |
| **Are there concerns that the target condition as defined by the reference standard does not match the question?** | Low concern |

##### Flow and Timing

| **A. Risk of Bias** | | |
| --- | --- | --- |
| Flow and timing |  | |
| Was there an appropriate interval between index test and reference standard? | | Yes |
| Did all patients receive the same reference standard? | | Yes |
| Were all patients included in the analysis? | | Yes |
| **Could the patient flow have introduced bias?** | | Low risk |

##### Notes

| **Notes** |
| --- |

#### Verlé 1994

##### Patient Selection

| **A. Risk of Bias** | | |
| --- | --- | --- |
| Patient Sampling | cross-sectional study ; consecutive sampling | |
| Was a consecutive or random sample of patients enrolled? | | Yes |
| Was a case-control design avoided? | | Yes |
| Did the study avoid inappropriate exclusions? | | Yes |
| **Could the selection of patients have introduced bias?** | | Low risk |

| **B. Concerns regarding applicability** | | |
| --- | --- | --- |
| Patient characteristics and setting | - 352 patients of all ages  - Country: Senegal  - Setting: field study | |
| **Are there concerns that the included patients and setting do not match the review question?** | | Low concern |

##### Index Test

| Index tests | micro-haematuria & proteinuria by urine reagent strips (Multistix) |
| --- | --- |

##### All tests

| **A. Risk of Bias** | |
| --- | --- |
| Were the index test results interpreted without knowledge of the results of the reference standard? | Unclear |
| If a threshold was used, was it pre-specified? | Yes |
| **Could the conduct or interpretation of the index test have introduced bias?** | Low risk |

| **B. Concerns regarding applicability** | |
| --- | --- |
| **Are there concerns that the index test, its conduct, or interpretation differ from the review question?** | Low concern |

##### Reference Standard

| **A. Risk of Bias** | | |
| --- | --- | --- |
| Target condition and reference standard(s) | S. haematobium  urine microscopy | |
| Is the reference standards likely to correctly classify the target condition? | | Unclear |
| Were the reference standard results interpreted without knowledge of the results of the index tests? | | Unclear |
| **Could the reference standard, its conduct, or its interpretation have introduced bias?** | | Low risk |

| **B. Concerns regarding applicability** | |
| --- | --- |
| **Are there concerns that the target condition as defined by the reference standard does not match the question?** | Low concern |

##### Flow and Timing

| **A. Risk of Bias** | | |
| --- | --- | --- |
| Flow and timing |  | |
| Was there an appropriate interval between index test and reference standard? | | Yes |
| Did all patients receive the same reference standard? | | Yes |
| Were all patients included in the analysis? | | Yes |
| **Could the patient flow have introduced bias?** | | Low risk |

##### Notes

| **Notes** |
| --- |

#### Vonghachack 2017

##### Patient Selection

| **A. Risk of Bias** | | |
| --- | --- | --- |
| Patient Sampling | cross-sectional survey; no random selection | |
| Was a consecutive or random sample of patients enrolled? | | Yes |
| Was a case-control design avoided? | | Yes |
| Did the study avoid inappropriate exclusions? | | Yes |
| **Could the selection of patients have introduced bias?** | | Low risk |

| **B. Concerns regarding applicability** | | |
| --- | --- | --- |
| Patient characteristics and setting | Species: S. mekongi  Country: Lao People’s Democratic Republic ; Cambodia  Sample size: 377  Age range: >6y (12–44y)  Participants: villagers  Setting: field study  Praziquantel before the study: | |
| **Are there concerns that the included patients and setting do not match the review question?** | | Low concern |

##### Index Test

| Index tests | AWE-ELISA; SEA-ELISA; CCA; CAA |
| --- | --- |

##### All tests

| **A. Risk of Bias** | |
| --- | --- |
| Were the index test results interpreted without knowledge of the results of the reference standard? | Yes |
| If a threshold was used, was it pre-specified? | Yes |
| **Could the conduct or interpretation of the index test have introduced bias?** | Low risk |

| **B. Concerns regarding applicability** | |
| --- | --- |
| **Are there concerns that the index test, its conduct, or interpretation differ from the review question?** | Low concern |

##### Reference Standard

| **A. Risk of Bias** | | |
| --- | --- | --- |
| Target condition and reference standard(s) | duplicate Kato Katz smears per each of the 3 stool sample | |
| Is the reference standards likely to correctly classify the target condition? | | No |
| Were the reference standard results interpreted without knowledge of the results of the index tests? | | Yes |
| **Could the reference standard, its conduct, or its interpretation have introduced bias?** | | Low risk |

| **B. Concerns regarding applicability** | |
| --- | --- |
| **Are there concerns that the target condition as defined by the reference standard does not match the question?** | Low concern |

##### Flow and Timing

| **A. Risk of Bias** | | |
| --- | --- | --- |
| Flow and timing |  | |
| Was there an appropriate interval between index test and reference standard? | | Yes |
| Did all patients receive the same reference standard? | | Yes |
| Were all patients included in the analysis? | | Yes |
| **Could the patient flow have introduced bias?** | | Low risk |

##### Notes

| **Notes** |
| --- |

#### Wilkins 1979

##### Patient Selection

| **A. Risk of Bias** | | |
| --- | --- | --- |
| Patient Sampling | cross-sectional study | |
| Was a consecutive or random sample of patients enrolled? | | Unclear |
| Was a case-control design avoided? | | Yes |
| Did the study avoid inappropriate exclusions? | | Unclear |
| **Could the selection of patients have introduced bias?** | | Low risk |

| **B. Concerns regarding applicability** | | |
| --- | --- | --- |
| Patient characteristics and setting | - 1944 participants from several small villages  - Country: Gambia  - Setting: field study | |
| **Are there concerns that the included patients and setting do not match the review question?** | | Low concern |

##### Index Test

| Index tests | micro-haematuria & proteinuria by urine reagent strips (Labstic) |
| --- | --- |

##### All tests

| **A. Risk of Bias** | |
| --- | --- |
| Were the index test results interpreted without knowledge of the results of the reference standard? | Unclear |
| If a threshold was used, was it pre-specified? | Unclear |
| **Could the conduct or interpretation of the index test have introduced bias?** | Low risk |

| **B. Concerns regarding applicability** | |
| --- | --- |
| **Are there concerns that the index test, its conduct, or interpretation differ from the review question?** | Low concern |

##### Reference Standard

| **A. Risk of Bias** | | |
| --- | --- | --- |
| Target condition and reference standard(s) | S. haematobium  urine microscopy | |
| Is the reference standards likely to correctly classify the target condition? | | Unclear |
| Were the reference standard results interpreted without knowledge of the results of the index tests? | | Unclear |
| **Could the reference standard, its conduct, or its interpretation have introduced bias?** | | Unclear risk |

| **B. Concerns regarding applicability** | |
| --- | --- |
| **Are there concerns that the target condition as defined by the reference standard does not match the question?** | Low concern |

##### Flow and Timing

| **A. Risk of Bias** | | |
| --- | --- | --- |
| Flow and timing |  | |
| Was there an appropriate interval between index test and reference standard? | | Unclear |
| Did all patients receive the same reference standard? | | Yes |
| Were all patients included in the analysis? | | Yes |
| **Could the patient flow have introduced bias?** | | Low risk |

##### Notes

| **Notes** |
| --- |

### Xu 2014

##### Patient Selection

| **A. Risk of Bias** | | |
| --- | --- | --- |
| Patient Sampling | cross-sectional survey; no random selection | |
| Was a consecutive or random sample of patients enrolled? | | Yes |
| Was a case-control design avoided? | | Yes |
| Did the study avoid inappropriate exclusions? | | Yes |
| **Could the selection of patients have introduced bias?** | | Low risk |

| **B. Concerns regarding applicability** | | |
| --- | --- | --- |
| Patient characteristics and setting | Species: S. japonicum  Country: China  Sample size: 1371  Age range: 5-78y  Participants: villagers  Setting: field study  Praziquantel before the study: | |
| **Are there concerns that the included patients and setting do not match the review question?** | | Low concern |

##### Index Test

| Index tests | SjSP-13-based ELISA; SEA-ELISA |
| --- | --- |

##### All tests

| **A. Risk of Bias** | |
| --- | --- |
| Were the index test results interpreted without knowledge of the results of the reference standard? | Yes |
| If a threshold was used, was it pre-specified? | Yes |
| **Could the conduct or interpretation of the index test have introduced bias?** | Low risk |

| **B. Concerns regarding applicability** | |
| --- | --- |
| **Are there concerns that the index test, its conduct, or interpretation differ from the review question?** | Low concern |

##### Reference Standard

| **A. Risk of Bias** | | |
| --- | --- | --- |
| Target condition and reference standard(s) | nine Kato-Katz thick smear per each 3 stool sample | |
| Is the reference standards likely to correctly classify the target condition? | | Yes |
| Were the reference standard results interpreted without knowledge of the results of the index tests? | | Yes |
| **Could the reference standard, its conduct, or its interpretation have introduced bias?** | | Low risk |

| **B. Concerns regarding applicability** | |
| --- | --- |
| **Are there concerns that the target condition as defined by the reference standard does not match the question?** | Low concern |

##### Flow and Timing

| **A. Risk of Bias** |
| --- |
| Flow and timing |
| Was there an appropriate interval between index test and reference standard? |
| Did all patients receive the same reference standard? |
| Were all patients included in the analysis? |
| **Could the patient flow have introduced bias?** |

##### Notes

| **Notes** |
| --- |

### Xu 2015

##### Patient Selection

| **A. Risk of Bias** | | |
| --- | --- | --- |
| Patient Sampling | cross-sectional survey; no random selection | |
| Was a consecutive or random sample of patients enrolled? | | Yes |
| Was a case-control design avoided? | | Yes |
| Did the study avoid inappropriate exclusions? | | Yes |
| **Could the selection of patients have introduced bias?** | | Low risk |

| **B. Concerns regarding applicability** | | |
| --- | --- | --- |
| Patient characteristics and setting | Species: S. japonicum  Country: China  Sample size: 110  Age range:  Participants: villagers  Setting: field study  Praziquantel before the study: | |
| **Are there concerns that the included patients and setting do not match the review question?** | | Low concern |

##### Index Test

| Index tests | LAMP |
| --- | --- |

##### All tests

| **A. Risk of Bias** | |
| --- | --- |
| Were the index test results interpreted without knowledge of the results of the reference standard? | Yes |
| If a threshold was used, was it pre-specified? | Yes |
| **Could the conduct or interpretation of the index test have introduced bias?** | Low risk |

| **B. Concerns regarding applicability** | |
| --- | --- |
| **Are there concerns that the index test, its conduct, or interpretation differ from the review question?** | Low concern |

##### Reference Standard

| **A. Risk of Bias** | | |
| --- | --- | --- |
| Target condition and reference standard(s) | triplicate Kato-Katz smears | |
| Is the reference standards likely to correctly classify the target condition? | | No |
| Were the reference standard results interpreted without knowledge of the results of the index tests? | | Yes |
| **Could the reference standard, its conduct, or its interpretation have introduced bias?** | | Low risk |

| **B. Concerns regarding applicability** | |
| --- | --- |
| **Are there concerns that the target condition as defined by the reference standard does not match the question?** | Low concern |

##### Flow and Timing

| **A. Risk of Bias** | | |
| --- | --- | --- |
| Flow and timing |  | |
| Was there an appropriate interval between index test and reference standard? | | Yes |
| Did all patients receive the same reference standard? | | Yes |
| Were all patients included in the analysis? | | Yes |
| **Could the patient flow have introduced bias?** | | Low risk |

##### Notes

| **Notes** |
| --- |

#### Zhang 2020

##### Patient Selection

| **A. Risk of Bias** | | |
| --- | --- | --- |
| Patient Sampling | 148 school children | |
| Was a consecutive or random sample of patients enrolled? | | Yes |
| Was a case-control design avoided? | | Yes |
| Did the study avoid inappropriate exclusions? | | Unclear |
| **Could the selection of patients have introduced bias?** | | Low risk |

| **B. Concerns regarding applicability** | | |
| --- | --- | --- |
| Patient characteristics and setting | - 147 children aged between 7 and 14 years (69 female, 78 male)  - Country: Zambia  - Setting: field study | |
| **Are there concerns that the included patients and setting do not match the review question?** | | Low concern |

##### Index Test

| Index tests | DDIA & IHA |
| --- | --- |

##### All tests

| **A. Risk of Bias** | |
| --- | --- |
| Were the index test results interpreted without knowledge of the results of the reference standard? | Unclear |
| If a threshold was used, was it pre-specified? | Unclear |
| **Could the conduct or interpretation of the index test have introduced bias?** | Low risk |

| **B. Concerns regarding applicability** | |
| --- | --- |
| **Are there concerns that the index test, its conduct, or interpretation differ from the review question?** | Low concern |

##### Reference Standard

| **A. Risk of Bias** | | |
| --- | --- | --- |
| Target condition and reference standard(s) | S. haematobium  urine microscopy | |
| Is the reference standards likely to correctly classify the target condition? | | Unclear |
| Were the reference standard results interpreted without knowledge of the results of the index tests? | | Unclear |
| **Could the reference standard, its conduct, or its interpretation have introduced bias?** | | Low risk |

| **B. Concerns regarding applicability** | |
| --- | --- |
| **Are there concerns that the target condition as defined by the reference standard does not match the question?** | Low concern |

##### Flow and Timing

| **A. Risk of Bias** | | |
| --- | --- | --- |
| Flow and timing |  | |
| Was there an appropriate interval between index test and reference standard? | | Yes |
| Did all patients receive the same reference standard? | | Yes |
| Were all patients included in the analysis? | | Unclear |
| **Could the patient flow have introduced bias?** | | Low risk |

##### Notes

| **Notes** |
| --- |

#### Zumstein 1983

##### Patient Selection

| **A. Risk of Bias** | | |
| --- | --- | --- |
| Patient Sampling | cross-sectional study | |
| Was a consecutive or random sample of patients enrolled? | | Unclear |
| Was a case-control design avoided? | | Unclear |
| Did the study avoid inappropriate exclusions? | | Unclear |
| **Could the selection of patients have introduced bias?** | | Low risk |

| **B. Concerns regarding applicability** | | |
| --- | --- | --- |
| Patient characteristics and setting | - 3478 school children from 15 schools aged between 6 and 19 years  - Country: Tanzania  - Setting: field study | |
| **Are there concerns that the included patients and setting do not match the review question?** | | Low concern |

##### Index Test

| Index tests | micro-haematuria by urine reagent test |
| --- | --- |

##### All tests

| **A. Risk of Bias** | |
| --- | --- |
| Were the index test results interpreted without knowledge of the results of the reference standard? | Unclear |
| If a threshold was used, was it pre-specified? | Unclear |
| **Could the conduct or interpretation of the index test have introduced bias?** | Low risk |

| **B. Concerns regarding applicability** | |
| --- | --- |
| **Are there concerns that the index test, its conduct, or interpretation differ from the review question?** | Low concern |

##### Reference Standard

| **A. Risk of Bias** | | |
| --- | --- | --- |
| Target condition and reference standard(s) | S. haematobium  urine microscopy | |
| Is the reference standards likely to correctly classify the target condition? | | Unclear |
| Were the reference standard results interpreted without knowledge of the results of the index tests? | | Unclear |
| **Could the reference standard, its conduct, or its interpretation have introduced bias?** | | Unclear risk |

| **B. Concerns regarding applicability** | |
| --- | --- |
| **Are there concerns that the target condition as defined by the reference standard does not match the question?** | Low concern |

##### Flow and Timing

| **A. Risk of Bias** | | |
| --- | --- | --- |
| Flow and timing |  | |
| Was there an appropriate interval between index test and reference standard? | | Yes |
| Did all patients receive the same reference standard? | | Yes |
| Were all patients included in the analysis? | | No |
| **Could the patient flow have introduced bias?** | | Low risk |

##### Notes

| **Notes** |
| --- |

###### Footnotes

### Characteristics of excluded studies

#### Ahmed 1993

| **Reason for exclusion** | combined gold standard |
| --- | --- |

#### Alarcon 1997

| **Reason for exclusion** | no indication regarding the reference test |
| --- | --- |

#### Allam 2021

| **Reason for exclusion** | wrong type of study |
| --- | --- |

#### Ansell 2002

| **Reason for exclusion** | wrong type of study |
| --- | --- |

#### Armoo 2020

| **Reason for exclusion** | combined gold standard |
| --- | --- |

#### Bärenbold 2018

| **Reason for exclusion** | statistical analysis |
| --- | --- |

#### Becker 2015

| **Reason for exclusion** | wrong type of study |
| --- | --- |

#### Belo 2009

| **Reason for exclusion** | Article not retrievable |
| --- | --- |

#### Buonfrate 2018

| **Reason for exclusion** | wrong type of study |
| --- | --- |

#### Chen 2011

| **Reason for exclusion** | article in Chinese |
| --- | --- |

#### Clements 2017

| **Reason for exclusion** | statistical analysis |
| --- | --- |

#### Clements 2018

| **Reason for exclusion** | empirical data not available |
| --- | --- |

#### Corstjens 2015

| **Reason for exclusion** | combined gold standard |
| --- | --- |

#### Diab 2019

| **Reason for exclusion** | paper retracted |
| --- | --- |

#### Diab 2021

| **Reason for exclusion** | empirical data not available (only LCA results) |
| --- | --- |

#### Downs 2015

| **Reason for exclusion** | wrong type of study (first samples collected in 2005, rest of samples collected in 2012) |
| --- | --- |

#### Elhag 2011

| **Reason for exclusion** | wrong type of study |
| --- | --- |

#### Enk 2012

| **Reason for exclusion** | combined gold standard |
| --- | --- |

#### Ephraim 2015

| **Reason for exclusion** | wrong type of study |
| --- | --- |

#### Espirito-Santo 2014

| **Reason for exclusion** | combined gold standard |
| --- | --- |

#### Etard 2004

| **Reason for exclusion** | empirical data not available |
| --- | --- |

#### Evengard 1990

| **Reason for exclusion** | wrong type of study |
| --- | --- |

#### Fenta 2020

| **Reason for exclusion** | combined gold standard |
| --- | --- |

#### Ferrer 2020

| **Reason for exclusion** | combined gold standard |
| --- | --- |

#### Fofana 2017

| **Reason for exclusion** | Article not retrievable |
| --- | --- |

#### Fofana 2019

| **Reason for exclusion** | wrong type of study (wrong disease --> Giardia intestinalis) |
| --- | --- |

#### Galappaththi-Arachchige 2018

| **Reason for exclusion** | combined gold standard |
| --- | --- |

#### Gargioni 2008

| **Reason for exclusion** | article in Portuguese |
| --- | --- |

#### Graeff-Teixeira 2020

| **Reason for exclusion** | meta-analysis |
| --- | --- |

#### Grenfell 2014

| **Reason for exclusion** | combined gold standard |
| --- | --- |

#### Guegan 2019

| **Reason for exclusion** | wrong type of study (no cross-sectional field study) |
| --- | --- |

#### Hall 1999

| **Reason for exclusion** | wrong type of publication (Short Report) |
| --- | --- |

#### Hamidu 2018

| **Reason for exclusion** | Article not retrievable |
| --- | --- |

#### Homsana 2020

| **Reason for exclusion** | wrong type of study (Schistosoma mekongi) |
| --- | --- |

#### Honkpehedji 2020

| **Reason for exclusion** | ongoing study |
| --- | --- |

#### Ibidapo 2005

| **Reason for exclusion** | wrong type of study |
| --- | --- |

#### Kanamura 1992

| **Reason for exclusion** | wrong type of study |
| --- | --- |

#### Kanamura 1998

| **Reason for exclusion** | wrong type of study |
| --- | --- |

#### Kanamura 2002

| **Reason for exclusion** | wrong type of study (wrong sampling method) |
| --- | --- |

#### Knight 1976

| **Reason for exclusion** | wrong type of study |
| --- | --- |

#### Kobayashi 1994

| **Reason for exclusion** | wrong type of study |
| --- | --- |

#### Koukounari 2009

| **Reason for exclusion** | statistical analysis |
| --- | --- |

#### Lee 2011

| **Reason for exclusion** | Article not retrievable |
| --- | --- |

#### Lier 2009

| **Reason for exclusion** | combined gold standard |
| --- | --- |

#### Lima 1998

| **Reason for exclusion** | wrong type of study |
| --- | --- |

#### Lodh 2014

| **Reason for exclusion** | Article not retrievable |
| --- | --- |

#### Long 1982

| **Reason for exclusion** | wrong type of study (antigen study) |
| --- | --- |

#### Marti 2020

| **Reason for exclusion** | no indication regarding the reference test |
| --- | --- |

#### McManus 2018

| **Reason for exclusion** | Article not retrievable |
| --- | --- |

#### Midzi 2003

| **Reason for exclusion** | wrong type of study (antigen study on female urinary schistosomiasis) |
| --- | --- |

#### Midzi 2020

| **Reason for exclusion** | statistical analysis |
| --- | --- |

#### Montenegro 1999

| **Reason for exclusion** | wrong type of study (study on matrices for ELISA) |
| --- | --- |

### Mu 2020

| **Reason for exclusion** | wrong type of study (biomarker study) |
| --- | --- |

#### Nakamura 2017

| **Reason for exclusion** | wrong type of study (case report) |
| --- | --- |

#### Neumayr 2019

| **Reason for exclusion** | wrong type of study |
| --- | --- |

#### Ngasala 2015

| **Reason for exclusion** | Article not retrievable |
| --- | --- |

#### Ochodo 2015

| **Reason for exclusion** | meta-analysis |
| --- | --- |

#### Ochola 2012

| **Reason for exclusion** |
| --- |

#### Oliveira 2018

| **Reason for exclusion** | combined gold standard |
| --- | --- |

#### Pieron 1980

| **Reason for exclusion** | combined gold standard |
| --- | --- |

#### Rostron 2019

| **Reason for exclusion** | wrong type of study |
| --- | --- |

#### Ruppel 1990

| **Reason for exclusion** | wrong type of study (sero-epidemiological study) |
| --- | --- |

#### Sarda 1985

| **Reason for exclusion** | empirical data not available |
| --- | --- |

#### Sarhan 2014

| **Reason for exclusion** | wrong type of study |
| --- | --- |

#### Sayed 2000

| **Reason for exclusion** | combined gold standard |
| --- | --- |

#### Schwarz 2014

| **Reason for exclusion** | wrong type of study |
| --- | --- |

#### Sheele 2016

| **Reason for exclusion** | wrong type of study |
| --- | --- |

#### Silva-Moraes 2019

| **Reason for exclusion** | combined gold standard |
| --- | --- |

#### Siziya 1996

| **Reason for exclusion** | Article not retrievable |
| --- | --- |

#### Stothard 2006

| **Reason for exclusion** | empirical data not available |
| --- | --- |

#### Stothard 2020

| **Reason for exclusion** | wrong type of study |
| --- | --- |

#### Taylor 1982

| **Reason for exclusion** | wrong type of study |
| --- | --- |

#### Teesdale 1985

| **Reason for exclusion** | wrong type of study |
| --- | --- |

#### Utzinger 2000

| **Reason for exclusion** | wrong type of study |
| --- | --- |

#### Utzinger 2008

| **Reason for exclusion** | wrong type of study |
| --- | --- |

#### Van Lieshout 1992

| **Reason for exclusion** | combined gold standard |
| --- | --- |

#### Viana 2019

| **Reason for exclusion** | wrong type of study |
| --- | --- |

#### Vinkeles Melchers 2014

| **Reason for exclusion** | combined gold standard |
| --- | --- |

#### Wang 1992

| **Reason for exclusion** | article in Chinese |
| --- | --- |

#### Wang 2011

| **Reason for exclusion** | article in Chinese |
| --- | --- |

#### Wang 2014

| **Reason for exclusion** | wrong type of study (A review) |
| --- | --- |

#### Weerakoon 2017

| **Reason for exclusion** | wrong type of study (PCR test comparison) |
| --- | --- |

### Xu 2018

| **Reason for exclusion** | wrong type of study (antigen study) |
| --- | --- |

#### Zhang 2000

| **Reason for exclusion** | article in Chinese |
| --- | --- |

#### Zheng 2005

| **Reason for exclusion** | article in Chinese |
| --- | --- |

#### Zhou 2014

| **Reason for exclusion** | wrong type of study (Schistosoma japonicum) |
| --- | --- |

#### Zhu 2000

| **Reason for exclusion** | wrong type of study (antigen study) |
| --- | --- |

#### Zhu 2010

| **Reason for exclusion** | meta-analysis |
| --- | --- |

###### Footnotes
