## Supplementary material for "Diagnostic tests for Schistosomiasis for low prevalence settings: a systematic review and Meta-Analysis": Summary of Findings tables

### Question: Should CAA vs. duplicate KK be used to diagnose Schistosomiasis in low endemic regions?

| CAA | | duplicate KK | |
| --- | --- | --- | --- |
| Sensitivity | 0.10 to 0.90 | Sensitivity | -- to -- |
| Specificity | 0.37 to 0.99 | Specificity | -- to -- |

| Prevalences | 5% | 10% | 20% |
| --- | --- | --- | --- |

| Outcome | № of studies (№ of patients) | Study design | Factors that may decrease certainty of evidence | | | | | Effect per 1,000 patients tested | | | | | | Test accuracy CoE |
| --- | --- | --- | --- | --- | --- | --- | --- | --- | --- | --- | --- | --- | --- | --- |
|  |  |  |  |  |  |  |  | pre-test probability of 5% | | pre-test probability of 10% | | pre-test probability of 20% | |  |
|  |  |  | Risk of bias | Indirectness | Inconsistency | Imprecision | Publication bias | CAA | duplicate KK | CAA | duplicate KK | CAA | duplicate KK |  |
| **True positives** (patients with Schistosomiasis) | 2 studies 500 patients | cross-sectional (cohort type accuracy study) | not serious | not serious | not serious | serious ^a^ | none | 5 to 45 | 0 to 0 | 10 to 90 | 0 to 0 | 20 to 180 | 0 to 0 | ⨁⨁⨁◯ MODERATE |
|  |  |  |  |  |  |  |  | **5 more to 45 more TP in CAA** | | **10 more to 90 more TP in CAA** | | **20 more to 180 more TP in CAA** | |  |
| **False negatives** (patients incorrectly classified as not having Schistosomiasis) |  |  |  |  |  |  |  | 5 to 45 | 50 to 50 | 10 to 90 | 100 to 100 | 20 to 180 | 200 to 200 |  |
|  |  |  |  |  |  |  |  | **5 fewer to 45 fewer FN in CAA** | | **10 fewer to 90 fewer FN in CAA** | | **20 fewer to 180 fewer FN in CAA** | |  |
| **True negatives** (patients without Schistosomiasis) | 2 studies 330 patients | cross-sectional (cohort type accuracy study) | not serious | not serious | not serious | serious ^a^ | none | 352 to 941 | 0 to 0 | 333 to 891 | 0 to 0 | 296 to 792 | 0 to 0 | ⨁⨁⨁◯ MODERATE |
|  |  |  |  |  |  |  |  | **352 more to 941 more TN in CAA** | | **333 more to 891 more TN in CAA** | | **296 more to 792 more TN in CAA** | |  |
| **False positives** (patients incorrectly classified as having Schistosomiasis) |  |  |  |  |  |  |  | 9 to 598 | 950 to 950 | 9 to 567 | 900 to 900 | 8 to 504 | 800 to 800 |  |
|  |  |  |  |  |  |  |  | **352 fewer to 941 fewer FP in CAA** | | **333 fewer to 891 fewer FP in CAA** | | **296 fewer to 792 fewer FP in CAA** | |  |

##### Explanations

a. imperfect reference test

### Question: Should CAA vs. quadruple KK be used to diagnose Schistosomiasis in low endemic regions?

| CAA | | quadruple KK | |
| --- | --- | --- | --- |
| Sensitivity | 0.96 (95% CI: 0.79 to 1.00) | Sensitivity | -- (95% CI: -- to --) |
| Specificity | 0.65 (95% CI: 0.60 to 0.70) | Specificity | -- (95% CI: -- to --) |

| Prevalences | 5% | 10% | 20% |
| --- | --- | --- | --- |

| Outcome | № of studies (№ of patients) | Study design | Factors that may decrease certainty of evidence | | | | | Effect per 1,000 patients tested | | | | | | Test accuracy CoE |
| --- | --- | --- | --- | --- | --- | --- | --- | --- | --- | --- | --- | --- | --- | --- |
|  |  |  |  |  |  |  |  | pre-test probability of 5% | | pre-test probability of 10% | | pre-test probability of 20% | |  |
|  |  |  | Risk of bias | Indirectness | Inconsistency | Imprecision | Publication bias | CAA | quadruple KK | CAA | quadruple KK | CAA | quadruple KK |  |
| **True positives** (patients with Schistosomiasis) | 1 studies 24 patients | cross-sectional (cohort type accuracy study) | not serious | not serious | not serious | serious ^a^ | none | 48 (40 to 50) | 0 (0 to 0) | 96 (79 to 100) | 0 (0 to 0) | 192 (158 to 200) | 0 (0 to 0) | ⨁⨁⨁◯ MODERATE |
|  |  |  |  |  |  |  |  | **48 more TP in CAA** | | **96 more TP in CAA** | | **192 more TP in CAA** | |  |
| **False negatives** (patients incorrectly classified as not having Schistosomiasis) |  |  |  |  |  |  |  | 2 (0 to 10) | 50 (50 to 50) | 4 (0 to 21) | 100 (100 to 100) | 8 (0 to 42) | 200 (200 to 200) |  |
|  |  |  |  |  |  |  |  | **48 fewer FN in CAA** | | **96 fewer FN in CAA** | | **192 fewer FN in CAA** | |  |
| **True negatives** (patients without Schistosomiasis) | 1 studies 353 patients | cross-sectional (cohort type accuracy study) | not serious | not serious | not serious | serious ^a^ | none | 617 (570 to 665) | 0 (0 to 0) | 585 (540 to 630) | 0 (0 to 0) | 520 (480 to 560) | 0 (0 to 0) | ⨁⨁⨁◯ MODERATE |
|  |  |  |  |  |  |  |  | **617 more TN in CAA** | | **585 more TN in CAA** | | **520 more TN in CAA** | |  |
| **False positives** (patients incorrectly classified as having Schistosomiasis) |  |  |  |  |  |  |  | 333 (285 to 380) | 950 (950 to 950) | 315 (270 to 360) | 900 (900 to 900) | 280 (240 to 320) | 800 (800 to 800) |  |
|  |  |  |  |  |  |  |  | **617 fewer FP in CAA** | | **585 fewer FP in CAA** | | **520 fewer FP in CAA** | |  |

##### Explanations

a. imperfect reference test

### Question: Should CCA1 cassette vs. duplicate KK be used to diagnose Schistosomiasis in low endemic regions?

| CCA1 cassette | | duplicate KK | |
| --- | --- | --- | --- |
| Sensitivity | 0.36 to 0.99 | Sensitivity | -- to -- |
| Specificity | 0.19 to 0.93 | Specificity | -- to -- |

| Prevalences | 5% | 10% | 20% |
| --- | --- | --- | --- |

| Outcome | № of studies (№ of patients) | Study design | Factors that may decrease certainty of evidence | | | | | Effect per 1,000 patients tested | | | | | | Test accuracy CoE |
| --- | --- | --- | --- | --- | --- | --- | --- | --- | --- | --- | --- | --- | --- | --- |
|  |  |  |  |  |  |  |  | pre-test probability of 5% | | pre-test probability of 10% | | pre-test probability of 20% | |  |
|  |  |  | Risk of bias | Indirectness | Inconsistency | Imprecision | Publication bias | CCA1 cassette | duplicate KK | CCA1 cassette | duplicate KK | CCA1 cassette | duplicate KK |  |
| **True positives** (patients with Schistosomiasis) | 18 studies 2108 patients | cross-sectional (cohort type accuracy study) | not serious | not serious | not serious | serious ^a^ | none | 18 to 50 | 0 to 0 | 36 to 99 | 0 to 0 | 72 to 198 | 0 to 0 | ⨁⨁⨁◯ MODERATE |
|  |  |  |  |  |  |  |  | **18 more to 50 more TP in CCA1 cassette** | | **36 more to 99 more TP in CCA1 cassette** | | **72 more to 198 more TP in CCA1 cassette** | |  |
| **False negatives** (patients incorrectly classified as not having Schistosomiasis) |  |  |  |  |  |  |  | 0 to 32 | 50 to 50 | 1 to 64 | 100 to 100 | 2 to 128 | 200 to 200 |  |
|  |  |  |  |  |  |  |  | **18 fewer to 50 fewer FN in CCA1 cassette** | | **36 fewer to 99 fewer FN in CCA1 cassette** | | **72 fewer to 198 fewer FN in CCA1 cassette** | |  |
| **True negatives** (patients without Schistosomiasis) | 18 studies 2776 patients | cross-sectional (cohort type accuracy study) | not serious | not serious | not serious | serious ^a^ | none | 181 to 884 | 0 to 0 | 171 to 837 | 0 to 0 | 152 to 744 | 0 to 0 | ⨁⨁⨁◯ MODERATE |
|  |  |  |  |  |  |  |  | **181 more to 884 more TN in CCA1 cassette** | | **171 more to 837 more TN in CCA1 cassette** | | **152 more to 744 more TN in CCA1 cassette** | |  |
| **False positives** (patients incorrectly classified as having Schistosomiasis) |  |  |  |  |  |  |  | 66 to 769 | 950 to 950 | 63 to 729 | 900 to 900 | 56 to 648 | 800 to 800 |  |
|  |  |  |  |  |  |  |  | **181 fewer to 884 fewer FP in CCA1 cassette** | | **171 fewer to 837 fewer FP in CCA1 cassette** | | **152 fewer to 744 fewer FP in CCA1 cassette** | |  |

##### Explanations

a. imperfect reference test

### Question: Should CCA1 cassette vs. quadruple KK be used to diagnose Schistosomiasis in low endemic regions?

| CCA1 cassette | | quadruple KK | |
| --- | --- | --- | --- |
| Sensitivity | 0.52 to 0.96 | Sensitivity | -- to -- |
| Specificity | 0.25 to 0.86 | Specificity | -- to -- |

| Prevalences | 5% | 10% | 20% |
| --- | --- | --- | --- |

| Outcome | № of studies (№ of patients) | Study design | Factors that may decrease certainty of evidence | | | | | Effect per 1,000 patients tested | | | | | | Test accuracy CoE |
| --- | --- | --- | --- | --- | --- | --- | --- | --- | --- | --- | --- | --- | --- | --- |
|  |  |  |  |  |  |  |  | pre-test probability of 5% | | pre-test probability of 10% | | pre-test probability of 20% | |  |
|  |  |  | Risk of bias | Indirectness | Inconsistency | Imprecision | Publication bias | CCA1 cassette | quadruple KK | CCA1 cassette | quadruple KK | CCA1 cassette | quadruple KK |  |
| **True positives** (patients with Schistosomiasis) | 11 studies 1238 patients | cross-sectional (cohort type accuracy study) | not serious | not serious | not serious | serious ^a^ | none | 26 to 48 | 0 to 0 | 52 to 96 | 0 to 0 | 104 to 192 | 0 to 0 | ⨁⨁⨁◯ MODERATE |
|  |  |  |  |  |  |  |  | **26 more to 48 more TP in CCA1 cassette** | | **52 more to 96 more TP in CCA1 cassette** | | **104 more to 192 more TP in CCA1 cassette** | |  |
| **False negatives** (patients incorrectly classified as not having Schistosomiasis) |  |  |  |  |  |  |  | 2 to 24 | 50 to 50 | 4 to 48 | 100 to 100 | 8 to 96 | 200 to 200 |  |
|  |  |  |  |  |  |  |  | **26 fewer to 48 fewer FN in CCA1 cassette** | | **52 fewer to 96 fewer FN in CCA1 cassette** | | **104 fewer to 192 fewer FN in CCA1 cassette** | |  |
| **True negatives** (patients without Schistosomiasis) | 11 studies 3354 patients | cross-sectional (cohort type accuracy study) | not serious | not serious | not serious | serious ^a^ | none | 238 to 817 | 0 to 0 | 225 to 774 | 0 to 0 | 200 to 688 | 0 to 0 | ⨁⨁⨁◯ MODERATE |
|  |  |  |  |  |  |  |  | **238 more to 817 more TN in CCA1 cassette** | | **225 more to 774 more TN in CCA1 cassette** | | **200 more to 688 more TN in CCA1 cassette** | |  |
| **False positives** (patients incorrectly classified as having Schistosomiasis) |  |  |  |  |  |  |  | 133 to 712 | 950 to 950 | 126 to 675 | 900 to 900 | 112 to 600 | 800 to 800 |  |
|  |  |  |  |  |  |  |  | **238 fewer to 817 fewer FP in CCA1 cassette** | | **225 fewer to 774 fewer FP in CCA1 cassette** | | **200 fewer to 688 fewer FP in CCA1 cassette** | |  |

##### Explanations

a. imperfect reference test

### Question: Should CCA1 cassette vs. sextuple KK be used to diagnose Schistosomiasis in low endemic regions?

| CCA1 cassette | | sextuple KK | |
| --- | --- | --- | --- |
| Sensitivity | 0.56 to 0.94 | Sensitivity | -- to -- |
| Specificity | 0.55 to 0.83 | Specificity | -- to -- |

| Prevalences | 5% | 10% | 20% |
| --- | --- | --- | --- |

| Outcome | № of studies (№ of patients) | Study design | Factors that may decrease certainty of evidence | | | | | Effect per 1,000 patients tested | | | | | | Test accuracy CoE |
| --- | --- | --- | --- | --- | --- | --- | --- | --- | --- | --- | --- | --- | --- | --- |
|  |  |  |  |  |  |  |  | pre-test probability of 5% | | pre-test probability of 10% | | pre-test probability of 20% | |  |
|  |  |  | Risk of bias | Indirectness | Inconsistency | Imprecision | Publication bias | CCA1 cassette | sextuple KK | CCA1 cassette | sextuple KK | CCA1 cassette | sextuple KK |  |
| **True positives** (patients with Schistosomiasis) | 7 studies 778 patients | cross-sectional (cohort type accuracy study) | not serious | not serious | not serious | serious ^a^ | none | 28 to 47 | 0 to 0 | 56 to 94 | 0 to 0 | 112 to 188 | 0 to 0 | ⨁⨁⨁◯ MODERATE |
|  |  |  |  |  |  |  |  | **28 more to 47 more TP in CCA1 cassette** | | **56 more to 94 more TP in CCA1 cassette** | | **112 more to 188 more TP in CCA1 cassette** | |  |
| **False negatives** (patients incorrectly classified as not having Schistosomiasis) |  |  |  |  |  |  |  | 3 to 22 | 50 to 50 | 6 to 44 | 100 to 100 | 12 to 88 | 200 to 200 |  |
|  |  |  |  |  |  |  |  | **28 fewer to 47 fewer FN in CCA1 cassette** | | **56 fewer to 94 fewer FN in CCA1 cassette** | | **112 fewer to 188 fewer FN in CCA1 cassette** | |  |
| **True negatives** (patients without Schistosomiasis) | 7 studies 1547 patients | cross-sectional (cohort type accuracy study) | not serious | not serious | not serious | serious ^a^ | none | 523 to 789 | 0 to 0 | 495 to 747 | 0 to 0 | 440 to 664 | 0 to 0 | ⨁⨁⨁◯ MODERATE |
|  |  |  |  |  |  |  |  | **523 more to 789 more TN in CCA1 cassette** | | **495 more to 747 more TN in CCA1 cassette** | | **440 more to 664 more TN in CCA1 cassette** | |  |
| **False positives** (patients incorrectly classified as having Schistosomiasis) |  |  |  |  |  |  |  | 161 to 427 | 950 to 950 | 153 to 405 | 900 to 900 | 136 to 360 | 800 to 800 |  |
|  |  |  |  |  |  |  |  | **523 fewer to 789 fewer FP in CCA1 cassette** | | **495 fewer to 747 fewer FP in CCA1 cassette** | | **440 fewer to 664 fewer FP in CCA1 cassette** | |  |

##### Explanations

a. imperfect reference test

### Question: Should CCA2 cassette vs. duplicate KK be used to diagnose Schistosomiasis in low endemic regions?

| CCA2 cassette | | duplicate KK | |
| --- | --- | --- | --- |
| Sensitivity | 0.63 (95% CI: 0.24 to 0.91) | Sensitivity | -- (95% CI: -- to --) |
| Specificity | 0.96 (95% CI: 0.89 to 0.99) | Specificity | -- (95% CI: -- to --) |

| Prevalences | 5% | 10% | 20% |
| --- | --- | --- | --- |

| Outcome | № of studies (№ of patients) | Study design | Factors that may decrease certainty of evidence | | | | | Effect per 1,000 patients tested | | | | | | Test accuracy CoE |
| --- | --- | --- | --- | --- | --- | --- | --- | --- | --- | --- | --- | --- | --- | --- |
|  |  |  |  |  |  |  |  | pre-test probability of 5% | | pre-test probability of 10% | | pre-test probability of 20% | |  |
|  |  |  | Risk of bias | Indirectness | Inconsistency | Imprecision | Publication bias | CCA2 cassette | duplicate KK | CCA2 cassette | duplicate KK | CCA2 cassette | duplicate KK |  |
| **True positives** (patients with Schistosomiasis) | 1 studies 8 patients | cross-sectional (cohort type accuracy study) | not serious | not serious | not serious | serious ^a^ | none | 32 (12 to 46) | 0 (0 to 0) | 63 (24 to 91) | 0 (0 to 0) | 126 (48 to 182) | 0 (0 to 0) | ⨁⨁⨁◯ MODERATE |
|  |  |  |  |  |  |  |  | **32 more TP in CCA2 cassette** | | **63 more TP in CCA2 cassette** | | **126 more TP in CCA2 cassette** | |  |
| **False negatives** (patients incorrectly classified as not having Schistosomiasis) |  |  |  |  |  |  |  | 18 (4 to 38) | 50 (50 to 50) | 37 (9 to 76) | 100 (100 to 100) | 74 (18 to 152) | 200 (200 to 200) |  |
|  |  |  |  |  |  |  |  | **32 fewer FN in CCA2 cassette** | | **63 fewer FN in CCA2 cassette** | | **126 fewer FN in CCA2 cassette** | |  |
| **True negatives** (patients without Schistosomiasis) | 1 studies 92 patients | cross-sectional (cohort type accuracy study) | not serious | not serious | not serious | serious ^a^ | none | 912 (845 to 941) | 0 (0 to 0) | 864 (801 to 891) | 0 (0 to 0) | 768 (712 to 792) | 0 (0 to 0) | ⨁⨁⨁◯ MODERATE |
|  |  |  |  |  |  |  |  | **912 more TN in CCA2 cassette** | | **864 more TN in CCA2 cassette** | | **768 more TN in CCA2 cassette** | |  |
| **False positives** (patients incorrectly classified as having Schistosomiasis) |  |  |  |  |  |  |  | 38 (9 to 105) | 950 (950 to 950) | 36 (9 to 99) | 900 (900 to 900) | 32 (8 to 88) | 800 (800 to 800) |  |
|  |  |  |  |  |  |  |  | **912 fewer FP in CCA2 cassette** | | **864 fewer FP in CCA2 cassette** | | **768 fewer FP in CCA2 cassette** | |  |

##### Explanations

a. imperfect reference test

### Question: Should CCA2 cassette vs. quadruple KK be used to diagnose Schistosomiasis in low endemic regions?

| CCA2 cassette | | quadruple KK | |
| --- | --- | --- | --- |
| Sensitivity | 0.45 (95% CI: 0.17 to 0.77) | Sensitivity | -- (95% CI: -- to --) |
| Specificity | 0.96 (95% CI: 0.89 to 0.99) | Specificity | -- (95% CI: -- to --) |

| Prevalences | 5% | 10% | 20% |
| --- | --- | --- | --- |

| Outcome | № of studies (№ of patients) | Study design | Factors that may decrease certainty of evidence | | | | | Effect per 1,000 patients tested | | | | | | Test accuracy CoE |
| --- | --- | --- | --- | --- | --- | --- | --- | --- | --- | --- | --- | --- | --- | --- |
|  |  |  |  |  |  |  |  | pre-test probability of 5% | | pre-test probability of 10% | | pre-test probability of 20% | |  |
|  |  |  | Risk of bias | Indirectness | Inconsistency | Imprecision | Publication bias | CCA2 cassette | quadruple KK | CCA2 cassette | quadruple KK | CCA2 cassette | quadruple KK |  |
| **True positives** (patients with Schistosomiasis) | 1 studies 11 patients | cross-sectional (cohort type accuracy study) | not serious | not serious | not serious | serious ^a^ | none | 23 (9 to 39) | 0 (0 to 0) | 45 (17 to 77) | 0 (0 to 0) | 90 (34 to 154) | 0 (0 to 0) | ⨁⨁⨁◯ MODERATE |
|  |  |  |  |  |  |  |  | **23 more TP in CCA2 cassette** | | **45 more TP in CCA2 cassette** | | **90 more TP in CCA2 cassette** | |  |
| **False negatives** (patients incorrectly classified as not having Schistosomiasis) |  |  |  |  |  |  |  | 27 (11 to 41) | 50 (50 to 50) | 55 (23 to 83) | 100 (100 to 100) | 110 (46 to 166) | 200 (200 to 200) |  |
|  |  |  |  |  |  |  |  | **23 fewer FN in CCA2 cassette** | | **45 fewer FN in CCA2 cassette** | | **90 fewer FN in CCA2 cassette** | |  |
| **True negatives** (patients without Schistosomiasis) | 1 studies 89 patients | cross-sectional (cohort type accuracy study) | not serious | not serious | not serious | serious ^a^ | none | 912 (845 to 941) | 0 (0 to 0) | 864 (801 to 891) | 0 (0 to 0) | 768 (712 to 792) | 0 (0 to 0) | ⨁⨁⨁◯ MODERATE |
|  |  |  |  |  |  |  |  | **912 more TN in CCA2 cassette** | | **864 more TN in CCA2 cassette** | | **768 more TN in CCA2 cassette** | |  |
| **False positives** (patients incorrectly classified as having Schistosomiasis) |  |  |  |  |  |  |  | 38 (9 to 105) | 950 (950 to 950) | 36 (9 to 99) | 900 (900 to 900) | 32 (8 to 88) | 800 (800 to 800) |  |
|  |  |  |  |  |  |  |  | **912 fewer FP in CCA2 cassette** | | **864 fewer FP in CCA2 cassette** | | **768 fewer FP in CCA2 cassette** | |  |

##### Explanations

a. imperfect reference test

### Question: Should FLOTAC (10 days) vs. triplicate KK be used to diagnose Schistosomiasis in low endemic regions?

| FLOTAC (10 days) | | triplicate KK | |
| --- | --- | --- | --- |
| Sensitivity | 0.96 (95% CI: 0.88 to 0.99) | Sensitivity | -- (95% CI: -- to --) |
| Specificity | 0.70 (95% CI: 0.53 to 0.83) | Specificity | -- (95% CI: -- to --) |

| Prevalences | 5% | 10% | 20% |
| --- | --- | --- | --- |

| Outcome | № of studies (№ of patients) | Study design | Factors that may decrease certainty of evidence | | | | | Effect per 1,000 patients tested | | | | | | Test accuracy CoE |
| --- | --- | --- | --- | --- | --- | --- | --- | --- | --- | --- | --- | --- | --- | --- |
|  |  |  |  |  |  |  |  | pre-test probability of 5% | | pre-test probability of 10% | | pre-test probability of 20% | |  |
|  |  |  | Risk of bias | Indirectness | Inconsistency | Imprecision | Publication bias | FLOTAC (10 days) | triplicate KK | FLOTAC (10 days) | triplicate KK | FLOTAC (10 days) | triplicate KK |  |
| **True positives** (patients with Schistosomiasis) | 1 studies 72 patients | cross-sectional (cohort type accuracy study) | not serious | not serious | not serious | serious ^a^ | none | 48 (44 to 50) | 0 (0 to 0) | 96 (88 to 99) | 0 (0 to 0) | 192 (176 to 198) | 0 (0 to 0) | ⨁⨁⨁◯ MODERATE |
|  |  |  |  |  |  |  |  | **48 more TP in FLOTAC (10 days)** | | **96 more TP in FLOTAC (10 days)** | | **192 more TP in FLOTAC (10 days)** | |  |
| **False negatives** (patients incorrectly classified as not having Schistosomiasis) |  |  |  |  |  |  |  | 2 (0 to 6) | 50 (50 to 50) | 4 (1 to 12) | 100 (100 to 100) | 8 (2 to 24) | 200 (200 to 200) |  |
|  |  |  |  |  |  |  |  | **48 fewer FN in FLOTAC (10 days)** | | **96 fewer FN in FLOTAC (10 days)** | | **192 fewer FN in FLOTAC (10 days)** | |  |
| **True negatives** (patients without Schistosomiasis) | 1 studies 40 patients | cross-sectional (cohort type accuracy study) | not serious | not serious | not serious | serious ^a^ | none | 665 (503 to 789) | 0 (0 to 0) | 630 (477 to 747) | 0 (0 to 0) | 560 (424 to 664) | 0 (0 to 0) | ⨁⨁⨁◯ MODERATE |
|  |  |  |  |  |  |  |  | **665 more TN in FLOTAC (10 days)** | | **630 more TN in FLOTAC (10 days)** | | **560 more TN in FLOTAC (10 days)** | |  |
| **False positives** (patients incorrectly classified as having Schistosomiasis) |  |  |  |  |  |  |  | 285 (161 to 447) | 950 (950 to 950) | 270 (153 to 423) | 900 (900 to 900) | 240 (136 to 376) | 800 (800 to 800) |  |
|  |  |  |  |  |  |  |  | **665 fewer FP in FLOTAC (10 days)** | | **630 fewer FP in FLOTAC (10 days)** | | **560 fewer FP in FLOTAC (10 days)** | |  |

##### Explanations

a. imperfect reference test

### Question: Should FLOTAC (30 days) vs. triplicate KK be used to diagnose Schistosomiasis in low endemic regions?

| FLOTAC (30 days) | | triplicate KK | |
| --- | --- | --- | --- |
| Sensitivity | 0.99 (95% CI: 0.93 to 1.00) | Sensitivity | -- (95% CI: -- to --) |
| Specificity | 0.65 (95% CI: 0.48 to 0.79) | Specificity | -- (95% CI: -- to --) |

| Prevalences | 5% | 10% | 20% |
| --- | --- | --- | --- |

| Outcome | № of studies (№ of patients) | Study design | Factors that may decrease certainty of evidence | | | | | Effect per 1,000 patients tested | | | | | | Test accuracy CoE |
| --- | --- | --- | --- | --- | --- | --- | --- | --- | --- | --- | --- | --- | --- | --- |
|  |  |  |  |  |  |  |  | pre-test probability of 5% | | pre-test probability of 10% | | pre-test probability of 20% | |  |
|  |  |  | Risk of bias | Indirectness | Inconsistency | Imprecision | Publication bias | FLOTAC (30 days) | triplicate KK | FLOTAC (30 days) | triplicate KK | FLOTAC (30 days) | triplicate KK |  |
| **True positives** (patients with Schistosomiasis) | 1 studies 72 patients | cross-sectional (cohort type accuracy study) | not serious | not serious | not serious | serious ^a^ | none | 50 (47 to 50) | 0 (0 to 0) | 99 (93 to 100) | 0 (0 to 0) | 198 (186 to 200) | 0 (0 to 0) | ⨁⨁⨁◯ MODERATE |
|  |  |  |  |  |  |  |  | **50 more TP in FLOTAC (30 days)** | | **99 more TP in FLOTAC (30 days)** | | **198 more TP in FLOTAC (30 days)** | |  |
| **False negatives** (patients incorrectly classified as not having Schistosomiasis) |  |  |  |  |  |  |  | 0 (0 to 3) | 50 (50 to 50) | 1 (0 to 7) | 100 (100 to 100) | 2 (0 to 14) | 200 (200 to 200) |  |
|  |  |  |  |  |  |  |  | **50 fewer FN in FLOTAC (30 days)** | | **99 fewer FN in FLOTAC (30 days)** | | **198 fewer FN in FLOTAC (30 days)** | |  |
| **True negatives** (patients without Schistosomiasis) | 1 studies 40 patients | cross-sectional (cohort type accuracy study) | not serious | not serious | not serious | serious ^a^ | none | 617 (456 to 751) | 0 (0 to 0) | 585 (432 to 711) | 0 (0 to 0) | 520 (384 to 632) | 0 (0 to 0) | ⨁⨁⨁◯ MODERATE |
|  |  |  |  |  |  |  |  | **617 more TN in FLOTAC (30 days)** | | **585 more TN in FLOTAC (30 days)** | | **520 more TN in FLOTAC (30 days)** | |  |
| **False positives** (patients incorrectly classified as having Schistosomiasis) |  |  |  |  |  |  |  | 333 (199 to 494) | 950 (950 to 950) | 315 (189 to 468) | 900 (900 to 900) | 280 (168 to 416) | 800 (800 to 800) |  |
|  |  |  |  |  |  |  |  | **617 fewer FP in FLOTAC (30 days)** | | **585 fewer FP in FLOTAC (30 days)** | | **520 fewer FP in FLOTAC (30 days)** | |  |

##### Explanations

a. imperfect reference test

### Question: Should FLOTAC (fresh) vs. triplicate KK be used to diagnose Schistosomiasis in low endemic regions?

| FLOTAC (fresh) | | triplicate KK | |
| --- | --- | --- | --- |
| Sensitivity | 0.71 (95% CI: 0.59 to 0.81) | Sensitivity | -- (95% CI: -- to --) |
| Specificity | 0.78 (95% CI: 0.62 to 0.89) | Specificity | -- (95% CI: -- to --) |

| Prevalences | 5% | 10% | 20% |
| --- | --- | --- | --- |

| Outcome | № of studies (№ of patients) | Study design | Factors that may decrease certainty of evidence | | | | | Effect per 1,000 patients tested | | | | | | Test accuracy CoE |
| --- | --- | --- | --- | --- | --- | --- | --- | --- | --- | --- | --- | --- | --- | --- |
|  |  |  |  |  |  |  |  | pre-test probability of 5% | | pre-test probability of 10% | | pre-test probability of 20% | |  |
|  |  |  | Risk of bias | Indirectness | Inconsistency | Imprecision | Publication bias | FLOTAC (fresh) | triplicate KK | FLOTAC (fresh) | triplicate KK | FLOTAC (fresh) | triplicate KK |  |
| **True positives** (patients with Schistosomiasis) | 1 studies 72 patients | cross-sectional (cohort type accuracy study) | not serious | not serious | not serious | serious ^a^ | none | 36 (30 to 41) | 0 (0 to 0) | 71 (59 to 81) | 0 (0 to 0) | 142 (118 to 162) | 0 (0 to 0) | ⨁⨁⨁◯ MODERATE |
|  |  |  |  |  |  |  |  | **36 more TP in FLOTAC (fresh)** | | **71 more TP in FLOTAC (fresh)** | | **142 more TP in FLOTAC (fresh)** | |  |
| **False negatives** (patients incorrectly classified as not having Schistosomiasis) |  |  |  |  |  |  |  | 14 (9 to 20) | 50 (50 to 50) | 29 (19 to 41) | 100 (100 to 100) | 58 (38 to 82) | 200 (200 to 200) |  |
|  |  |  |  |  |  |  |  | **36 fewer FN in FLOTAC (fresh)** | | **71 fewer FN in FLOTAC (fresh)** | | **142 fewer FN in FLOTAC (fresh)** | |  |
| **True negatives** (patients without Schistosomiasis) | 1 studies 40 patients | cross-sectional (cohort type accuracy study) | not serious | not serious | not serious | serious ^a^ | none | 741 (589 to 845) | 0 (0 to 0) | 702 (558 to 801) | 0 (0 to 0) | 624 (496 to 712) | 0 (0 to 0) | ⨁⨁⨁◯ MODERATE |
|  |  |  |  |  |  |  |  | **741 more TN in FLOTAC (fresh)** | | **702 more TN in FLOTAC (fresh)** | | **624 more TN in FLOTAC (fresh)** | |  |
| **False positives** (patients incorrectly classified as having Schistosomiasis) |  |  |  |  |  |  |  | 209 (105 to 361) | 950 (950 to 950) | 198 (99 to 342) | 900 (900 to 900) | 176 (88 to 304) | 800 (800 to 800) |  |
|  |  |  |  |  |  |  |  | **741 fewer FP in FLOTAC (fresh)** | | **702 fewer FP in FLOTAC (fresh)** | | **624 fewer FP in FLOTAC (fresh)** | |  |

##### Explanations

a. imperfect reference test

### Question: Should SmCTF-RDT vs. quadruple KK be used to diagnose Schistosomiasis in low endemic regions?

| SmCTF-RDT | | quadruple KK | |
| --- | --- | --- | --- |
| Sensitivity | 0.54 to 1.00 | Sensitivity | -- to -- |
| Specificity | 0.13 to 0.41 | Specificity | -- to -- |

| Prevalences | 5% | 10% | 20% |
| --- | --- | --- | --- |

| Outcome | № of studies (№ of patients) | Study design | Factors that may decrease certainty of evidence | | | | | Effect per 1,000 patients tested | | | | | | Test accuracy CoE |
| --- | --- | --- | --- | --- | --- | --- | --- | --- | --- | --- | --- | --- | --- | --- |
|  |  |  |  |  |  |  |  | pre-test probability of 5% | | pre-test probability of 10% | | pre-test probability of 20% | |  |
|  |  |  | Risk of bias | Indirectness | Inconsistency | Imprecision | Publication bias | SmCTF-RDT | quadruple KK | SmCTF-RDT | quadruple KK | SmCTF-RDT | quadruple KK |  |
| **True positives** (patients with Schistosomiasis) | 4 studies 114 patients | cross-sectional (cohort type accuracy study) | not serious | not serious | not serious | serious ^a^ | none | 27 to 50 | 0 to 0 | 54 to 100 | 0 to 0 | 108 to 200 | 0 to 0 | ⨁⨁⨁◯ MODERATE |
|  |  |  |  |  |  |  |  | **27 more to 50 more TP in SmCTF-RDT** | | **54 more to 100 more TP in SmCTF-RDT** | | **108 more to 200 more TP in SmCTF-RDT** | |  |
| **False negatives** (patients incorrectly classified as not having Schistosomiasis) |  |  |  |  |  |  |  | 0 to 23 | 50 to 50 | 0 to 46 | 100 to 100 | 0 to 92 | 200 to 200 |  |
|  |  |  |  |  |  |  |  | **27 fewer to 50 fewer FN in SmCTF-RDT** | | **54 fewer to 100 fewer FN in SmCTF-RDT** | | **108 fewer to 200 fewer FN in SmCTF-RDT** | |  |
| **True negatives** (patients without Schistosomiasis) | 4 studies 177 patients | cross-sectional (cohort type accuracy study) | not serious | not serious | not serious | serious ^a^ | none | 124 to 389 | 0 to 0 | 117 to 369 | 0 to 0 | 104 to 328 | 0 to 0 | ⨁⨁⨁◯ MODERATE |
|  |  |  |  |  |  |  |  | **124 more to 389 more TN in SmCTF-RDT** | | **117 more to 369 more TN in SmCTF-RDT** | | **104 more to 328 more TN in SmCTF-RDT** | |  |
| **False positives** (patients incorrectly classified as having Schistosomiasis) |  |  |  |  |  |  |  | 561 to 826 | 950 to 950 | 531 to 783 | 900 to 900 | 472 to 696 | 800 to 800 |  |
|  |  |  |  |  |  |  |  | **124 fewer to 389 fewer FP in SmCTF-RDT** | | **117 fewer to 369 fewer FP in SmCTF-RDT** | | **104 fewer to 328 fewer FP in SmCTF-RDT** | |  |

##### Explanations

a. imperfect reference test

### Question: Should SmCTF-RDT vs. Urine Microscopy be used to diagnose Schistosomiasis in low endemic regions?

| SmCTF-RDT | | Urine Microscopy | |
| --- | --- | --- | --- |
| Sensitivity | 0.67 (95% CI: 0.22 to 0.96) | Sensitivity | -- (95% CI: -- to --) |
| Specificity | 0.33 (95% CI: 0.25 to 0.43) | Specificity | -- (95% CI: -- to --) |

| Prevalences | 5% | 10% | 20% |
| --- | --- | --- | --- |

| Outcome | № of studies (№ of patients) | Study design | Factors that may decrease certainty of evidence | | | | | Effect per 1,000 patients tested | | | | | | Test accuracy CoE |
| --- | --- | --- | --- | --- | --- | --- | --- | --- | --- | --- | --- | --- | --- | --- |
|  |  |  |  |  |  |  |  | pre-test probability of 5% | | pre-test probability of 10% | | pre-test probability of 20% | |  |
|  |  |  | Risk of bias | Indirectness | Inconsistency | Imprecision | Publication bias | SmCTF-RDT | Urine Microscopy | SmCTF-RDT | Urine Microscopy | SmCTF-RDT | Urine Microscopy |  |
| **True positives** (patients with Schistosomiasis) | 1 studies 6 patients | cross-sectional (cohort type accuracy study) | not serious | not serious | not serious | serious ^a^ | none | 34 (11 to 48) | 0 (0 to 0) | 67 (22 to 96) | 0 (0 to 0) | 134 (44 to 192) | 0 (0 to 0) | ⨁⨁⨁◯ MODERATE |
|  |  |  |  |  |  |  |  | **34 more TP in SmCTF-RDT** | | **67 more TP in SmCTF-RDT** | | **134 more TP in SmCTF-RDT** | |  |
| **False negatives** (patients incorrectly classified as not having Schistosomiasis) |  |  |  |  |  |  |  | 16 (2 to 39) | 50 (50 to 50) | 33 (4 to 78) | 100 (100 to 100) | 66 (8 to 156) | 200 (200 to 200) |  |
|  |  |  |  |  |  |  |  | **34 fewer FN in SmCTF-RDT** | | **67 fewer FN in SmCTF-RDT** | | **134 fewer FN in SmCTF-RDT** | |  |
| **True negatives** (patients without Schistosomiasis) | 1 studies 111 patients | cross-sectional (cohort type accuracy study) | not serious | not serious | not serious | serious ^a^ | none | 314 (238 to 409) | 0 (0 to 0) | 297 (225 to 387) | 0 (0 to 0) | 264 (200 to 344) | 0 (0 to 0) | ⨁⨁⨁◯ MODERATE |
|  |  |  |  |  |  |  |  | **314 more TN in SmCTF-RDT** | | **297 more TN in SmCTF-RDT** | | **264 more TN in SmCTF-RDT** | |  |
| **False positives** (patients incorrectly classified as having Schistosomiasis) |  |  |  |  |  |  |  | 636 (541 to 712) | 950 (950 to 950) | 603 (513 to 675) | 900 (900 to 900) | 536 (456 to 600) | 800 (800 to 800) |  |
|  |  |  |  |  |  |  |  | **314 fewer FP in SmCTF-RDT** | | **297 fewer FP in SmCTF-RDT** | | **264 fewer FP in SmCTF-RDT** | |  |

##### Explanations

a. imperfect reference test

### Question: Should Anti IGg RDt-Sh vs. Urine Microscopy be used to diagnose Schistosomiasis in low endemic regions?

| Anti IGg RDt-Sh | | Urine Microscopy | |
| --- | --- | --- | --- |
| Sensitivity | 0.46 (95% CI: 0.35 to 0.58) | Sensitivity | -- (95% CI: -- to --) |
| Specificity | 0.10 (95% CI: 0.05 to 0.19) | Specificity | -- (95% CI: -- to --) |

| Prevalences | 5% | 10% | 20% |
| --- | --- | --- | --- |

| Outcome | № of studies (№ of patients) | Study design | Factors that may decrease certainty of evidence | | | | | Effect per 1,000 patients tested | | | | | | Test accuracy CoE |
| --- | --- | --- | --- | --- | --- | --- | --- | --- | --- | --- | --- | --- | --- | --- |
|  |  |  |  |  |  |  |  | pre-test probability of 5% | | pre-test probability of 10% | | pre-test probability of 20% | |  |
|  |  |  | Risk of bias | Indirectness | Inconsistency | Imprecision | Publication bias | Anti IGg RDt-Sh | Urine Microscopy | Anti IGg RDt-Sh | Urine Microscopy | Anti IGg RDt-Sh | Urine Microscopy |  |
| **True positives** (patients with Schistosomiasis) | 1 studies 82 patients | cross-sectional (cohort type accuracy study) | not serious | not serious | not serious | serious ^a^ | none | 23 (17 to 29) | 0 (0 to 0) | 46 (35 to 58) | 0 (0 to 0) | 92 (70 to 116) | 0 (0 to 0) | ⨁⨁⨁◯ MODERATE |
|  |  |  |  |  |  |  |  | **23 more TP in Anti IGg RDt-Sh** | | **46 more TP in Anti IGg RDt-Sh** | | **92 more TP in Anti IGg RDt-Sh** | |  |
| **False negatives** (patients incorrectly classified as not having Schistosomiasis) |  |  |  |  |  |  |  | 27 (21 to 33) | 50 (50 to 50) | 54 (42 to 65) | 100 (100 to 100) | 108 (84 to 130) | 200 (200 to 200) |  |
|  |  |  |  |  |  |  |  | **23 fewer FN in Anti IGg RDt-Sh** | | **46 fewer FN in Anti IGg RDt-Sh** | | **92 fewer FN in Anti IGg RDt-Sh** | |  |
| **True negatives** (patients without Schistosomiasis) | 1 studies 78 patients | cross-sectional (cohort type accuracy study) | not serious | not serious | not serious | serious ^a^ | none | 95 (48 to 181) | 0 (0 to 0) | 90 (45 to 171) | 0 (0 to 0) | 80 (40 to 152) | 0 (0 to 0) | ⨁⨁⨁◯ MODERATE |
|  |  |  |  |  |  |  |  | **95 more TN in Anti IGg RDt-Sh** | | **90 more TN in Anti IGg RDt-Sh** | | **80 more TN in Anti IGg RDt-Sh** | |  |
| **False positives** (patients incorrectly classified as having Schistosomiasis) |  |  |  |  |  |  |  | 855 (769 to 902) | 950 (950 to 950) | 810 (729 to 855) | 900 (900 to 900) | 720 (648 to 760) | 800 (800 to 800) |  |
|  |  |  |  |  |  |  |  | **95 fewer FP in Anti IGg RDt-Sh** | | **90 fewer FP in Anti IGg RDt-Sh** | | **80 fewer FP in Anti IGg RDt-Sh** | |  |

##### Explanations

a. imperfect reference test

### Question: Should AWE-SEA ELISA vs. quadruple KK be used to diagnose Schistosomiasis in low endemic regions?

| AWE-SEA ELISA | | quadruple KK | |
| --- | --- | --- | --- |
| Sensitivity | 0.69 to 0.69 | Sensitivity | -- to -- |
| Specificity | 0.62 to 0.70 | Specificity | -- to -- |

| Prevalences | 5% | 10% | 20% |
| --- | --- | --- | --- |

| Outcome | № of studies (№ of patients) | Study design | Factors that may decrease certainty of evidence | | | | | Effect per 1,000 patients tested | | | | | | Test accuracy CoE |
| --- | --- | --- | --- | --- | --- | --- | --- | --- | --- | --- | --- | --- | --- | --- |
|  |  |  |  |  |  |  |  | pre-test probability of 5% | | pre-test probability of 10% | | pre-test probability of 20% | |  |
|  |  |  | Risk of bias | Indirectness | Inconsistency | Imprecision | Publication bias | AWE-SEA ELISA | quadruple KK | AWE-SEA ELISA | quadruple KK | AWE-SEA ELISA | quadruple KK |  |
| **True positives** (patients with Schistosomiasis) | 2 studies 47 patients | cross-sectional (cohort type accuracy study) | not serious | not serious | not serious | serious ^a^ | none | 34 to 35 | 0 to 0 | 69 to 69 | 0 to 0 | 138 to 138 | 0 to 0 | ⨁⨁⨁◯ MODERATE |
|  |  |  |  |  |  |  |  | **34 more to 35 more TP in AWE-SEA ELISA** | | **69 more to 69 more TP in AWE-SEA ELISA** | | **138 more to 138 more TP in AWE-SEA ELISA** | |  |
| **False negatives** (patients incorrectly classified as not having Schistosomiasis) |  |  |  |  |  |  |  | 15 to 16 | 50 to 50 | 31 to 31 | 100 to 100 | 62 to 62 | 200 to 200 |  |
|  |  |  |  |  |  |  |  | **34 fewer to 35 fewer FN in AWE-SEA ELISA** | | **69 fewer to 69 fewer FN in AWE-SEA ELISA** | | **138 fewer to 138 fewer FN in AWE-SEA ELISA** | |  |
| **True negatives** (patients without Schistosomiasis) | 2 studies 473 patients | cross-sectional (cohort type accuracy study) | not serious | not serious | not serious | serious ^a^ | none | 589 to 665 | 0 to 0 | 558 to 630 | 0 to 0 | 496 to 560 | 0 to 0 | ⨁⨁⨁◯ MODERATE |
|  |  |  |  |  |  |  |  | **589 more to 665 more TN in AWE-SEA ELISA** | | **558 more to 630 more TN in AWE-SEA ELISA** | | **496 more to 560 more TN in AWE-SEA ELISA** | |  |
| **False positives** (patients incorrectly classified as having Schistosomiasis) |  |  |  |  |  |  |  | 285 to 361 | 950 to 950 | 270 to 342 | 900 to 900 | 240 to 304 | 800 to 800 |  |
|  |  |  |  |  |  |  |  | **589 fewer to 665 fewer FP in AWE-SEA ELISA** | | **558 fewer to 630 fewer FP in AWE-SEA ELISA** | | **496 fewer to 560 fewer FP in AWE-SEA ELISA** | |  |

##### Explanations

a. imperfect reference test

### Question: Should Haematuria (R strip) vs. Urine Microscopy be used to diagnose Schistosomiasis in low endemic regions?

| Haematuria (R strip) | | Urine Microscopy | |
| --- | --- | --- | --- |
| Sensitivity | 0.16 to 1.00 | Sensitivity | -- to -- |
| Specificity | 0.24 to 1.00 | Specificity | -- to -- |

| Prevalences | 5% | 10% | 20% |
| --- | --- | --- | --- |

| Outcome | № of studies (№ of patients) | Study design | Factors that may decrease certainty of evidence | | | | | Effect per 1,000 patients tested | | | | | | Test accuracy CoE |
| --- | --- | --- | --- | --- | --- | --- | --- | --- | --- | --- | --- | --- | --- | --- |
|  |  |  |  |  |  |  |  | pre-test probability of 5% | | pre-test probability of 10% | | pre-test probability of 20% | |  |
|  |  |  | Risk of bias | Indirectness | Inconsistency | Imprecision | Publication bias | Haematuria (R strip) | Urine Microscopy | Haematuria (R strip) | Urine Microscopy | Haematuria (R strip) | Urine Microscopy |  |
| **True positives** (patients with Schistosomiasis) | 72 studies 26614 patients | cross-sectional (cohort type accuracy study) | not serious | not serious | not serious | serious ^a^ | none | 8 to 50 | 0 to 0 | 16 to 100 | 0 to 0 | 32 to 200 | 0 to 0 | ⨁⨁⨁◯ MODERATE |
|  |  |  |  |  |  |  |  | **8 more to 50 more TP in Haematuria (R strip)** | | **16 more to 100 more TP in Haematuria (R strip)** | | **32 more to 200 more TP in Haematuria (R strip)** | |  |
| **False negatives** (patients incorrectly classified as not having Schistosomiasis) |  |  |  |  |  |  |  | 0 to 42 | 50 to 50 | 0 to 84 | 100 to 100 | 0 to 168 | 200 to 200 |  |
|  |  |  |  |  |  |  |  | **8 fewer to 50 fewer FN in Haematuria (R strip)** | | **16 fewer to 100 fewer FN in Haematuria (R strip)** | | **32 fewer to 200 fewer FN in Haematuria (R strip)** | |  |
| **True negatives** (patients without Schistosomiasis) | 72 studies 129665 patients | cross-sectional (cohort type accuracy study) | not serious | not serious | not serious | serious ^a^ | none | 228 to 950 | 0 to 0 | 216 to 900 | 0 to 0 | 192 to 800 | 0 to 0 | ⨁⨁⨁◯ MODERATE |
|  |  |  |  |  |  |  |  | **228 more to 950 more TN in Haematuria (R strip)** | | **216 more to 900 more TN in Haematuria (R strip)** | | **192 more to 800 more TN in Haematuria (R strip)** | |  |
| **False positives** (patients incorrectly classified as having Schistosomiasis) |  |  |  |  |  |  |  | 0 to 722 | 950 to 950 | 0 to 684 | 900 to 900 | 0 to 608 | 800 to 800 |  |
|  |  |  |  |  |  |  |  | **228 fewer to 950 fewer FP in Haematuria (R strip)** | | **216 fewer to 900 fewer FP in Haematuria (R strip)** | | **192 fewer to 800 fewer FP in Haematuria (R strip)** | |  |

##### Explanations

a. imperfect reference test

### Question: Should IgG ELISA vs. triplicate KK be used to diagnose Schistosomiasis in low endemic regions?

| IgG ELISA | | triplicate KK | |
| --- | --- | --- | --- |
| Sensitivity | 0.84 to 0.98 | Sensitivity | -- to -- |
| Specificity | 0.14 to 0.99 | Specificity | -- to -- |

| Prevalences | 5% | 10% | 20% |
| --- | --- | --- | --- |

| Outcome | № of studies (№ of patients) | Study design | Factors that may decrease certainty of evidence | | | | | Effect per 1,000 patients tested | | | | | | Test accuracy CoE |
| --- | --- | --- | --- | --- | --- | --- | --- | --- | --- | --- | --- | --- | --- | --- |
|  |  |  |  |  |  |  |  | pre-test probability of 5% | | pre-test probability of 10% | | pre-test probability of 20% | |  |
|  |  |  | Risk of bias | Indirectness | Inconsistency | Imprecision | Publication bias | IgG ELISA | triplicate KK | IgG ELISA | triplicate KK | IgG ELISA | triplicate KK |  |
| **True positives** (patients with Schistosomiasis) | 4 studies 512 patients | cross-sectional (cohort type accuracy study) | not serious | not serious | not serious | serious ^a^ | none | 42 to 49 | 0 to 0 | 84 to 98 | 0 to 0 | 168 to 196 | 0 to 0 | ⨁⨁⨁◯ MODERATE |
|  |  |  |  |  |  |  |  | **42 more to 49 more TP in IgG ELISA** | | **84 more to 98 more TP in IgG ELISA** | | **168 more to 196 more TP in IgG ELISA** | |  |
| **False negatives** (patients incorrectly classified as not having Schistosomiasis) |  |  |  |  |  |  |  | 1 to 8 | 50 to 50 | 2 to 16 | 100 to 100 | 4 to 32 | 200 to 200 |  |
|  |  |  |  |  |  |  |  | **42 fewer to 49 fewer FN in IgG ELISA** | | **84 fewer to 98 fewer FN in IgG ELISA** | | **168 fewer to 196 fewer FN in IgG ELISA** | |  |
| **True negatives** (patients without Schistosomiasis) | 4 studies 442 patients | cross-sectional (cohort type accuracy study) | not serious | not serious | not serious | serious ^a^ | none | 133 to 941 | 0 to 0 | 126 to 891 | 0 to 0 | 112 to 792 | 0 to 0 | ⨁⨁⨁◯ MODERATE |
|  |  |  |  |  |  |  |  | **133 more to 941 more TN in IgG ELISA** | | **126 more to 891 more TN in IgG ELISA** | | **112 more to 792 more TN in IgG ELISA** | |  |
| **False positives** (patients incorrectly classified as having Schistosomiasis) |  |  |  |  |  |  |  | 9 to 817 | 950 to 950 | 9 to 774 | 900 to 900 | 8 to 688 | 800 to 800 |  |
|  |  |  |  |  |  |  |  | **133 fewer to 941 fewer FP in IgG ELISA** | | **126 fewer to 891 fewer FP in IgG ELISA** | | **112 fewer to 792 fewer FP in IgG ELISA** | |  |

##### Explanations

a. imperfect reference test

### Question: Should IgG SEA-ELISA vs. CCA1 cassette be used to diagnose Schistosomiasis in low endemic regions?

| IgG SEA-ELISA | | CCA1 cassette | |
| --- | --- | --- | --- |
| Sensitivity | 0.97 (95% CI: 0.92 to 0.99) | Sensitivity | -- (95% CI: -- to --) |
| Specificity | 0.53 (95% CI: 0.43 to 0.63) | Specificity | -- (95% CI: -- to --) |

| Prevalences | 5% | 10% | 20% |
| --- | --- | --- | --- |

| Outcome | № of studies (№ of patients) | Study design | Factors that may decrease certainty of evidence | | | | | Effect per 1,000 patients tested | | | | | | Test accuracy CoE |
| --- | --- | --- | --- | --- | --- | --- | --- | --- | --- | --- | --- | --- | --- | --- |
|  |  |  |  |  |  |  |  | pre-test probability of 5% | | pre-test probability of 10% | | pre-test probability of 20% | |  |
|  |  |  | Risk of bias | Indirectness | Inconsistency | Imprecision | Publication bias | IgG SEA-ELISA | CCA1 cassette | IgG SEA-ELISA | CCA1 cassette | IgG SEA-ELISA | CCA1 cassette |  |
| **True positives** (patients with Schistosomiasis) | 1 studies 147 patients | cross-sectional (cohort type accuracy study) | not serious | not serious | not serious | serious ^a^ | none | 49 (46 to 50) | 0 (0 to 0) | 97 (92 to 99) | 0 (0 to 0) | 194 (184 to 198) | 0 (0 to 0) | ⨁⨁⨁◯ MODERATE |
|  |  |  |  |  |  |  |  | **49 more TP in IgG SEA-ELISA** | | **97 more TP in IgG SEA-ELISA** | | **194 more TP in IgG SEA-ELISA** | |  |
| **False negatives** (patients incorrectly classified as not having Schistosomiasis) |  |  |  |  |  |  |  | 1 (0 to 4) | 50 (50 to 50) | 3 (1 to 8) | 100 (100 to 100) | 6 (2 to 16) | 200 (200 to 200) |  |
|  |  |  |  |  |  |  |  | **49 fewer FN in IgG SEA-ELISA** | | **97 fewer FN in IgG SEA-ELISA** | | **194 fewer FN in IgG SEA-ELISA** | |  |
| **True negatives** (patients without Schistosomiasis) | 1 studies 111 patients | cross-sectional (cohort type accuracy study) | not serious | not serious | not serious | serious ^a^ | none | 503 (409 to 598) | 0 (0 to 0) | 477 (387 to 567) | 0 (0 to 0) | 424 (344 to 504) | 0 (0 to 0) | ⨁⨁⨁◯ MODERATE |
|  |  |  |  |  |  |  |  | **503 more TN in IgG SEA-ELISA** | | **477 more TN in IgG SEA-ELISA** | | **424 more TN in IgG SEA-ELISA** | |  |
| **False positives** (patients incorrectly classified as having Schistosomiasis) |  |  |  |  |  |  |  | 447 (352 to 541) | 950 (950 to 950) | 423 (333 to 513) | 900 (900 to 900) | 376 (296 to 456) | 800 (800 to 800) |  |
|  |  |  |  |  |  |  |  | **503 fewer FP in IgG SEA-ELISA** | | **477 fewer FP in IgG SEA-ELISA** | | **424 fewer FP in IgG SEA-ELISA** | |  |

##### Explanations

a. imperfect reference test

### Question: Should IgG SEA-ELISA vs. Urine Microscopy be used to diagnose Schistosomiasis in low endemic regions?

| IgG SEA-ELISA | | Urine Microscopy | |
| --- | --- | --- | --- |
| Sensitivity | 0.78 to 0.95 | Sensitivity | -- to -- |
| Specificity | 0.30 to 0.95 | Specificity | -- to -- |

| Prevalences | 5% | 10% | 20% |
| --- | --- | --- | --- |

| Outcome | № of studies (№ of patients) | Study design | Factors that may decrease certainty of evidence | | | | | Effect per 1,000 patients tested | | | | | | Test accuracy CoE |
| --- | --- | --- | --- | --- | --- | --- | --- | --- | --- | --- | --- | --- | --- | --- |
|  |  |  |  |  |  |  |  | pre-test probability of 5% | | pre-test probability of 10% | | pre-test probability of 20% | |  |
|  |  |  | Risk of bias | Indirectness | Inconsistency | Imprecision | Publication bias | IgG SEA-ELISA | Urine Microscopy | IgG SEA-ELISA | Urine Microscopy | IgG SEA-ELISA | Urine Microscopy |  |
| **True positives** (patients with Schistosomiasis) | 4 studies 197 patients | cross-sectional (cohort type accuracy study) | not serious | not serious | not serious | serious ^a^ | none | 39 to 48 | 0 to 0 | 78 to 95 | 0 to 0 | 156 to 190 | 0 to 0 | ⨁⨁⨁◯ MODERATE |
|  |  |  |  |  |  |  |  | **39 more to 48 more TP in IgG SEA-ELISA** | | **78 more to 95 more TP in IgG SEA-ELISA** | | **156 more to 190 more TP in IgG SEA-ELISA** | |  |
| **False negatives** (patients incorrectly classified as not having Schistosomiasis) |  |  |  |  |  |  |  | 2 to 11 | 50 to 50 | 5 to 22 | 100 to 100 | 10 to 44 | 200 to 200 |  |
|  |  |  |  |  |  |  |  | **39 fewer to 48 fewer FN in IgG SEA-ELISA** | | **78 fewer to 95 fewer FN in IgG SEA-ELISA** | | **156 fewer to 190 fewer FN in IgG SEA-ELISA** | |  |
| **True negatives** (patients without Schistosomiasis) | 4 studies 306 patients | cross-sectional (cohort type accuracy study) | not serious | not serious | not serious | serious ^a^ | none | 285 to 903 | 0 to 0 | 270 to 855 | 0 to 0 | 240 to 760 | 0 to 0 | ⨁⨁⨁◯ MODERATE |
|  |  |  |  |  |  |  |  | **285 more to 903 more TN in IgG SEA-ELISA** | | **270 more to 855 more TN in IgG SEA-ELISA** | | **240 more to 760 more TN in IgG SEA-ELISA** | |  |
| **False positives** (patients incorrectly classified as having Schistosomiasis) |  |  |  |  |  |  |  | 47 to 665 | 950 to 950 | 45 to 630 | 900 to 900 | 40 to 560 | 800 to 800 |  |
|  |  |  |  |  |  |  |  | **285 fewer to 903 fewer FP in IgG SEA-ELISA** | | **270 fewer to 855 fewer FP in IgG SEA-ELISA** | | **240 fewer to 760 fewer FP in IgG SEA-ELISA** | |  |

##### Explanations

a. imperfect reference test

### Question: Should IgM ELISA vs. triplicate KK be used to diagnose Schistosomiasis in low endemic regions?

| IgM ELISA | | triplicate KK | |
| --- | --- | --- | --- |
| Sensitivity | 0.98 (95% CI: 0.89 to 1.00) | Sensitivity | -- (95% CI: -- to --) |
| Specificity | 0.98 (95% CI: 0.92 to 1.00) | Specificity | -- (95% CI: -- to --) |

| Prevalences | 5% | 10% | 20% |
| --- | --- | --- | --- |

| Outcome | № of studies (№ of patients) | Study design | Factors that may decrease certainty of evidence | | | | | Effect per 1,000 patients tested | | | | | | Test accuracy CoE |
| --- | --- | --- | --- | --- | --- | --- | --- | --- | --- | --- | --- | --- | --- | --- |
|  |  |  |  |  |  |  |  | pre-test probability of 5% | | pre-test probability of 10% | | pre-test probability of 20% | |  |
|  |  |  | Risk of bias | Indirectness | Inconsistency | Imprecision | Publication bias | IgM ELISA | triplicate KK | IgM ELISA | triplicate KK | IgM ELISA | triplicate KK |  |
| **True positives** (patients with Schistosomiasis) | 1 studies 50 patients | cross-sectional (cohort type accuracy study) | not serious | not serious | not serious | serious ^a^ | none | 49 (45 to 50) | 0 (0 to 0) | 98 (89 to 100) | 0 (0 to 0) | 196 (178 to 200) | 0 (0 to 0) | ⨁⨁⨁◯ MODERATE |
|  |  |  |  |  |  |  |  | **49 more TP in IgM ELISA** | | **98 more TP in IgM ELISA** | | **196 more TP in IgM ELISA** | |  |
| **False negatives** (patients incorrectly classified as not having Schistosomiasis) |  |  |  |  |  |  |  | 1 (0 to 5) | 50 (50 to 50) | 2 (0 to 11) | 100 (100 to 100) | 4 (0 to 22) | 200 (200 to 200) |  |
|  |  |  |  |  |  |  |  | **49 fewer FN in IgM ELISA** | | **98 fewer FN in IgM ELISA** | | **196 fewer FN in IgM ELISA** | |  |
| **True negatives** (patients without Schistosomiasis) | 1 studies 87 patients | cross-sectional (cohort type accuracy study) | not serious | not serious | not serious | serious ^a^ | none | 931 (874 to 950) | 0 (0 to 0) | 882 (828 to 900) | 0 (0 to 0) | 784 (736 to 800) | 0 (0 to 0) | ⨁⨁⨁◯ MODERATE |
|  |  |  |  |  |  |  |  | **931 more TN in IgM ELISA** | | **882 more TN in IgM ELISA** | | **784 more TN in IgM ELISA** | |  |
| **False positives** (patients incorrectly classified as having Schistosomiasis) |  |  |  |  |  |  |  | 19 (0 to 76) | 950 (950 to 950) | 18 (0 to 72) | 900 (900 to 900) | 16 (0 to 64) | 800 (800 to 800) |  |
|  |  |  |  |  |  |  |  | **931 fewer FP in IgM ELISA** | | **882 fewer FP in IgM ELISA** | | **784 fewer FP in IgM ELISA** | |  |

##### Explanations

a. imperfect reference test

### Question: Should Leukocyturia (R strip) vs. Urine Microscopy be used to diagnose Schistosomiasis in low endemic regions?

| Leukocyturia (R strip) | | Urine Microscopy | |
| --- | --- | --- | --- |
| Sensitivity | 0.32 to 0.73 | Sensitivity | -- to -- |
| Specificity | 0.23 to 0.93 | Specificity | -- to -- |

| Prevalences | 5% | 10% | 20% |
| --- | --- | --- | --- |

| Outcome | № of studies (№ of patients) | Study design | Factors that may decrease certainty of evidence | | | | | Effect per 1,000 patients tested | | | | | | Test accuracy CoE |
| --- | --- | --- | --- | --- | --- | --- | --- | --- | --- | --- | --- | --- | --- | --- |
|  |  |  |  |  |  |  |  | pre-test probability of 5% | | pre-test probability of 10% | | pre-test probability of 20% | |  |
|  |  |  | Risk of bias | Indirectness | Inconsistency | Imprecision | Publication bias | Leukocyturia (R strip) | Urine Microscopy | Leukocyturia (R strip) | Urine Microscopy | Leukocyturia (R strip) | Urine Microscopy |  |
| **True positives** (patients with Schistosomiasis) | 5 studies 687 patients | cross-sectional (cohort type accuracy study) | not serious | not serious | not serious | serious ^a^ | none | 16 to 37 | 0 to 0 | 32 to 73 | 0 to 0 | 64 to 146 | 0 to 0 | ⨁⨁⨁◯ MODERATE |
|  |  |  |  |  |  |  |  | **16 more to 37 more TP in Leukocyturia (R strip)** | | **32 more to 73 more TP in Leukocyturia (R strip)** | | **64 more to 146 more TP in Leukocyturia (R strip)** | |  |
| **False negatives** (patients incorrectly classified as not having Schistosomiasis) |  |  |  |  |  |  |  | 13 to 34 | 50 to 50 | 27 to 68 | 100 to 100 | 54 to 136 | 200 to 200 |  |
|  |  |  |  |  |  |  |  | **16 fewer to 37 fewer FN in Leukocyturia (R strip)** | | **32 fewer to 73 fewer FN in Leukocyturia (R strip)** | | **64 fewer to 146 fewer FN in Leukocyturia (R strip)** | |  |
| **True negatives** (patients without Schistosomiasis) | 5 studies 845 patients | cross-sectional (cohort type accuracy study) | not serious | not serious | not serious | serious ^a^ | none | 219 to 884 | 0 to 0 | 207 to 837 | 0 to 0 | 184 to 744 | 0 to 0 | ⨁⨁⨁◯ MODERATE |
|  |  |  |  |  |  |  |  | **219 more to 884 more TN in Leukocyturia (R strip)** | | **207 more to 837 more TN in Leukocyturia (R strip)** | | **184 more to 744 more TN in Leukocyturia (R strip)** | |  |
| **False positives** (patients incorrectly classified as having Schistosomiasis) |  |  |  |  |  |  |  | 66 to 731 | 950 to 950 | 63 to 693 | 900 to 900 | 56 to 616 | 800 to 800 |  |
|  |  |  |  |  |  |  |  | **219 fewer to 884 fewer FP in Leukocyturia (R strip)** | | **207 fewer to 837 fewer FP in Leukocyturia (R strip)** | | **184 fewer to 744 fewer FP in Leukocyturia (R strip)** | |  |

##### Explanations

a. imperfect reference test

### Question: Should Proteinuria (R strip) vs. Urine Microscopy be used to diagnose Schistosomiasis in low endemic regions?

| Proteinuria (R strip) | | Urine Microscopy | |
| --- | --- | --- | --- |
| Sensitivity | 0.12 to 0.93 | Sensitivity | -- to -- |
| Specificity | 0.11 to 0.99 | Specificity | -- to -- |

| Prevalences | 5% | 10% | 20% |
| --- | --- | --- | --- |

| Outcome | № of studies (№ of patients) | Study design | Factors that may decrease certainty of evidence | | | | | Effect per 1,000 patients tested | | | | | | Test accuracy CoE |
| --- | --- | --- | --- | --- | --- | --- | --- | --- | --- | --- | --- | --- | --- | --- |
|  |  |  |  |  |  |  |  | pre-test probability of 5% | | pre-test probability of 10% | | pre-test probability of 20% | |  |
|  |  |  | Risk of bias | Indirectness | Inconsistency | Imprecision | Publication bias | Proteinuria (R strip) | Urine Microscopy | Proteinuria (R strip) | Urine Microscopy | Proteinuria (R strip) | Urine Microscopy |  |
| **True positives** (patients with Schistosomiasis) | 41 studies 17452 patients | cross-sectional (cohort type accuracy study) | not serious | not serious | not serious | serious ^a^ | none | 6 to 47 | 0 to 0 | 12 to 93 | 0 to 0 | 24 to 186 | 0 to 0 | ⨁⨁⨁◯ MODERATE |
|  |  |  |  |  |  |  |  | **6 more to 47 more TP in Proteinuria (R strip)** | | **12 more to 93 more TP in Proteinuria (R strip)** | | **24 more to 186 more TP in Proteinuria (R strip)** | |  |
| **False negatives** (patients incorrectly classified as not having Schistosomiasis) |  |  |  |  |  |  |  | 3 to 44 | 50 to 50 | 7 to 88 | 100 to 100 | 14 to 176 | 200 to 200 |  |
|  |  |  |  |  |  |  |  | **6 fewer to 47 fewer FN in Proteinuria (R strip)** | | **12 fewer to 93 fewer FN in Proteinuria (R strip)** | | **24 fewer to 186 fewer FN in Proteinuria (R strip)** | |  |
| **True negatives** (patients without Schistosomiasis) | 41 studies 62014 patients | cross-sectional (cohort type accuracy study) | not serious | not serious | not serious | serious ^a^ | none | 105 to 941 | 0 to 0 | 99 to 891 | 0 to 0 | 88 to 792 | 0 to 0 | ⨁⨁⨁◯ MODERATE |
|  |  |  |  |  |  |  |  | **105 more to 941 more TN in Proteinuria (R strip)** | | **99 more to 891 more TN in Proteinuria (R strip)** | | **88 more to 792 more TN in Proteinuria (R strip)** | |  |
| **False positives** (patients incorrectly classified as having Schistosomiasis) |  |  |  |  |  |  |  | 9 to 845 | 950 to 950 | 9 to 801 | 900 to 900 | 8 to 712 | 800 to 800 |  |
|  |  |  |  |  |  |  |  | **105 fewer to 941 fewer FP in Proteinuria (R strip)** | | **99 fewer to 891 fewer FP in Proteinuria (R strip)** | | **88 fewer to 792 fewer FP in Proteinuria (R strip)** | |  |

##### Explanations

a. imperfect reference test

### Question: Should rSP13 ELISA vs. 27 KK be used to diagnose Schistosomiasis in low endemic regions?

| rSP13 ELISA | | 27 KK | |
| --- | --- | --- | --- |
| Sensitivity | 0.96 (95% CI: 0.89 to 0.99) | Sensitivity | -- (95% CI: -- to --) |
| Specificity | 0.70 (95% CI: 0.67 to 0.72) | Specificity | -- (95% CI: -- to --) |

| Prevalences | 5% | 10% | 20% |
| --- | --- | --- | --- |

| Outcome | № of studies (№ of patients) | Study design | Factors that may decrease certainty of evidence | | | | | Effect per 1,000 patients tested | | | | | | Test accuracy CoE |
| --- | --- | --- | --- | --- | --- | --- | --- | --- | --- | --- | --- | --- | --- | --- |
|  |  |  |  |  |  |  |  | pre-test probability of 5% | | pre-test probability of 10% | | pre-test probability of 20% | |  |
|  |  |  | Risk of bias | Indirectness | Inconsistency | Imprecision | Publication bias | rSP13 ELISA | 27 KK | rSP13 ELISA | 27 KK | rSP13 ELISA | 27 KK |  |
| **True positives** (patients with Schistosomiasis) | 1 studies 74 patients | cross-sectional (cohort type accuracy study) | not serious | not serious | not serious | serious ^a^ | none | 48 (45 to 50) | 0 (0 to 0) | 96 (89 to 99) | 0 (0 to 0) | 192 (178 to 198) | 0 (0 to 0) | ⨁⨁⨁◯ MODERATE |
|  |  |  |  |  |  |  |  | **48 more TP in rSP13 ELISA** | | **96 more TP in rSP13 ELISA** | | **192 more TP in rSP13 ELISA** | |  |
| **False negatives** (patients incorrectly classified as not having Schistosomiasis) |  |  |  |  |  |  |  | 2 (0 to 5) | 50 (50 to 50) | 4 (1 to 11) | 100 (100 to 100) | 8 (2 to 22) | 200 (200 to 200) |  |
|  |  |  |  |  |  |  |  | **48 fewer FN in rSP13 ELISA** | | **96 fewer FN in rSP13 ELISA** | | **192 fewer FN in rSP13 ELISA** | |  |
| **True negatives** (patients without Schistosomiasis) | 1 studies 1297 patients | cross-sectional (cohort type accuracy study) | not serious | not serious | not serious | serious ^a^ | none | 665 (637 to 684) | 0 (0 to 0) | 630 (603 to 648) | 0 (0 to 0) | 560 (536 to 576) | 0 (0 to 0) | ⨁⨁⨁◯ MODERATE |
|  |  |  |  |  |  |  |  | **665 more TN in rSP13 ELISA** | | **630 more TN in rSP13 ELISA** | | **560 more TN in rSP13 ELISA** | |  |
| **False positives** (patients incorrectly classified as having Schistosomiasis) |  |  |  |  |  |  |  | 285 (266 to 313) | 950 (950 to 950) | 270 (252 to 297) | 900 (900 to 900) | 240 (224 to 264) | 800 (800 to 800) |  |
|  |  |  |  |  |  |  |  | **665 fewer FP in rSP13 ELISA** | | **630 fewer FP in rSP13 ELISA** | | **560 fewer FP in rSP13 ELISA** | |  |

##### Explanations

a. imperfect reference test

### Question: Should Sm DNA PCR vs. duplicate KK be used to diagnose Schistosomiasis in low endemic regions?

| Sm DNA PCR | | duplicate KK | |
| --- | --- | --- | --- |
| Sensitivity | 1.00 (95% CI: 0.92 to 1.00) | Sensitivity | -- (95% CI: -- to --) |
| Specificity | 0.23 (95% CI: 0.11 to 0.38) | Specificity | -- (95% CI: -- to --) |

| Prevalences | 5% | 10% | 20% |
| --- | --- | --- | --- |

| Outcome | № of studies (№ of patients) | Study design | Factors that may decrease certainty of evidence | | | | | Effect per 1,000 patients tested | | | | | | Test accuracy CoE |
| --- | --- | --- | --- | --- | --- | --- | --- | --- | --- | --- | --- | --- | --- | --- |
|  |  |  |  |  |  |  |  | pre-test probability of 5% | | pre-test probability of 10% | | pre-test probability of 20% | |  |
|  |  |  | Risk of bias | Indirectness | Inconsistency | Imprecision | Publication bias | Sm DNA PCR | duplicate KK | Sm DNA PCR | duplicate KK | Sm DNA PCR | duplicate KK |  |
| **True positives** (patients with Schistosomiasis) | 1 studies 45 patients | cross-sectional (cohort type accuracy study) | not serious | not serious | not serious | serious ^a^ | none | 50 (46 to 50) | 0 (0 to 0) | 100 (92 to 100) | 0 (0 to 0) | 200 (184 to 200) | 0 (0 to 0) | ⨁⨁⨁◯ MODERATE |
|  |  |  |  |  |  |  |  | **50 more TP in Sm DNA PCR** | | **100 more TP in Sm DNA PCR** | | **200 more TP in Sm DNA PCR** | |  |
| **False negatives** (patients incorrectly classified as not having Schistosomiasis) |  |  |  |  |  |  |  | 0 (0 to 4) | 50 (50 to 50) | 0 (0 to 8) | 100 (100 to 100) | 0 (0 to 16) | 200 (200 to 200) |  |
|  |  |  |  |  |  |  |  | **50 fewer FN in Sm DNA PCR** | | **100 fewer FN in Sm DNA PCR** | | **200 fewer FN in Sm DNA PCR** | |  |
| **True negatives** (patients without Schistosomiasis) | 1 studies 44 patients | cross-sectional (cohort type accuracy study) | not serious | not serious | not serious | serious ^a^ | none | 219 (105 to 361) | 0 (0 to 0) | 207 (99 to 342) | 0 (0 to 0) | 184 (88 to 304) | 0 (0 to 0) | ⨁⨁⨁◯ MODERATE |
|  |  |  |  |  |  |  |  | **219 more TN in Sm DNA PCR** | | **207 more TN in Sm DNA PCR** | | **184 more TN in Sm DNA PCR** | |  |
| **False positives** (patients incorrectly classified as having Schistosomiasis) |  |  |  |  |  |  |  | 731 (589 to 845) | 950 (950 to 950) | 693 (558 to 801) | 900 (900 to 900) | 616 (496 to 712) | 800 (800 to 800) |  |
|  |  |  |  |  |  |  |  | **219 fewer FP in Sm DNA PCR** | | **207 fewer FP in Sm DNA PCR** | | **184 fewer FP in Sm DNA PCR** | |  |

##### Explanations

a. imperfect reference test

### Question: Should SWAP ELISA vs. sextuple KK be used to diagnose Schistosomiasis in low endemic regions?

| SWAP ELISA | | sextuple KK | |
| --- | --- | --- | --- |
| Sensitivity | 0.92 (95% CI: 0.87 to 0.95) | Sensitivity | -- (95% CI: -- to --) |
| Specificity | 0.57 (95% CI: 0.51 to 0.63) | Specificity | -- (95% CI: -- to --) |

| Prevalences | 5% | 10% | 20% |
| --- | --- | --- | --- |

| Outcome | № of studies (№ of patients) | Study design | Factors that may decrease certainty of evidence | | | | | Effect per 1,000 patients tested | | | | | | Test accuracy CoE |
| --- | --- | --- | --- | --- | --- | --- | --- | --- | --- | --- | --- | --- | --- | --- |
|  |  |  |  |  |  |  |  | pre-test probability of 5% | | pre-test probability of 10% | | pre-test probability of 20% | |  |
|  |  |  | Risk of bias | Indirectness | Inconsistency | Imprecision | Publication bias | SWAP ELISA | sextuple KK | SWAP ELISA | sextuple KK | SWAP ELISA | sextuple KK |  |
| **True positives** (patients with Schistosomiasis) | 1 studies 187 patients | cross-sectional (cohort type accuracy study) | not serious | not serious | not serious | serious ^a^ | none | 46 (44 to 48) | 0 (0 to 0) | 92 (87 to 95) | 0 (0 to 0) | 184 (174 to 190) | 0 (0 to 0) | ⨁⨁⨁◯ MODERATE |
|  |  |  |  |  |  |  |  | **46 more TP in SWAP ELISA** | | **92 more TP in SWAP ELISA** | | **184 more TP in SWAP ELISA** | |  |
| **False negatives** (patients incorrectly classified as not having Schistosomiasis) |  |  |  |  |  |  |  | 4 (2 to 6) | 50 (50 to 50) | 8 (5 to 13) | 100 (100 to 100) | 16 (10 to 26) | 200 (200 to 200) |  |
|  |  |  |  |  |  |  |  | **46 fewer FN in SWAP ELISA** | | **92 fewer FN in SWAP ELISA** | | **184 fewer FN in SWAP ELISA** | |  |
| **True negatives** (patients without Schistosomiasis) | 1 studies 295 patients | cross-sectional (cohort type accuracy study) | not serious | not serious | not serious | serious ^a^ | none | 542 (485 to 598) | 0 (0 to 0) | 513 (459 to 567) | 0 (0 to 0) | 456 (408 to 504) | 0 (0 to 0) | ⨁⨁⨁◯ MODERATE |
|  |  |  |  |  |  |  |  | **542 more TN in SWAP ELISA** | | **513 more TN in SWAP ELISA** | | **456 more TN in SWAP ELISA** | |  |
| **False positives** (patients incorrectly classified as having Schistosomiasis) |  |  |  |  |  |  |  | 408 (352 to 465) | 950 (950 to 950) | 387 (333 to 441) | 900 (900 to 900) | 344 (296 to 392) | 800 (800 to 800) |  |
|  |  |  |  |  |  |  |  | **542 fewer FP in SWAP ELISA** | | **513 fewer FP in SWAP ELISA** | | **456 fewer FP in SWAP ELISA** | |  |

##### Explanations

a. imperfect reference test

### Question: Should CAA vs. Urine Microscopy be used to diagnose Schistosomiasis in low endemic regions?

| CAA | | Urine Microscopy | |
| --- | --- | --- | --- |
| Sensitivity | 0.16 to 0.97 | Sensitivity | -- to -- |
| Specificity | 0.24 to 1.00 | Specificity | -- to -- |

| Prevalences | 5% | 10% | 20% |
| --- | --- | --- | --- |

| Outcome | № of studies (№ of patients) | Study design | Factors that may decrease certainty of evidence | | | | | Effect per 1,000 patients tested | | | | | | Test accuracy CoE |
| --- | --- | --- | --- | --- | --- | --- | --- | --- | --- | --- | --- | --- | --- | --- |
|  |  |  |  |  |  |  |  | pre-test probability of 5% | | pre-test probability of 10% | | pre-test probability of 20% | |  |
|  |  |  | Risk of bias | Indirectness | Inconsistency | Imprecision | Publication bias | CAA | Urine Microscopy | CAA | Urine Microscopy | CAA | Urine Microscopy |  |
| **True positives** (patients with Schistosomiasis) | 4 studies 564 patients | cross-sectional (cohort type accuracy study) | not serious | not serious | not serious | serious ^a^ | none | 8 to 49 | 0 to 0 | 16 to 97 | 0 to 0 | 32 to 194 | 0 to 0 | ⨁⨁⨁◯ MODERATE |
|  |  |  |  |  |  |  |  | **8 more to 49 more TP in CAA** | | **16 more to 97 more TP in CAA** | | **32 more to 194 more TP in CAA** | |  |
| **False negatives** (patients incorrectly classified as not having Schistosomiasis) |  |  |  |  |  |  |  | 1 to 42 | 50 to 50 | 3 to 84 | 100 to 100 | 6 to 168 | 200 to 200 |  |
|  |  |  |  |  |  |  |  | **8 fewer to 49 fewer FN in CAA** | | **16 fewer to 97 fewer FN in CAA** | | **32 fewer to 194 fewer FN in CAA** | |  |
| **True negatives** (patients without Schistosomiasis) | 4 studies 683 patients | cross-sectional (cohort type accuracy study) | not serious | not serious | not serious | serious ^a^ | none | 228 to 950 | 0 to 0 | 216 to 900 | 0 to 0 | 192 to 800 | 0 to 0 | ⨁⨁⨁◯ MODERATE |
|  |  |  |  |  |  |  |  | **228 more to 950 more TN in CAA** | | **216 more to 900 more TN in CAA** | | **192 more to 800 more TN in CAA** | |  |
| **False positives** (patients incorrectly classified as having Schistosomiasis) |  |  |  |  |  |  |  | 0 to 722 | 950 to 950 | 0 to 684 | 900 to 900 | 0 to 608 | 800 to 800 |  |
|  |  |  |  |  |  |  |  | **228 fewer to 950 fewer FP in CAA** | | **216 fewer to 900 fewer FP in CAA** | | **192 fewer to 800 fewer FP in CAA** | |  |

##### Explanations

a. imperfect reference test

### Question: Should CCA1 cassette vs. Urine Microscopy be used to diagnose Schistosomiasis in low endemic regions?

| CCA1 cassette | | Urine Microscopy | |
| --- | --- | --- | --- |
| Sensitivity | 0.09 to 0.79 | Sensitivity | -- to -- |
| Specificity | 0.44 to 0.98 | Specificity | -- to -- |

| Prevalences | 5% | 10% | 20% |
| --- | --- | --- | --- |

| Outcome | № of studies (№ of patients) | Study design | Factors that may decrease certainty of evidence | | | | | Effect per 1,000 patients tested | | | | | | Test accuracy CoE |
| --- | --- | --- | --- | --- | --- | --- | --- | --- | --- | --- | --- | --- | --- | --- |
|  |  |  |  |  |  |  |  | pre-test probability of 5% | | pre-test probability of 10% | | pre-test probability of 20% | |  |
|  |  |  | Risk of bias | Indirectness | Inconsistency | Imprecision | Publication bias | CCA1 cassette | Urine Microscopy | CCA1 cassette | Urine Microscopy | CCA1 cassette | Urine Microscopy |  |
| **True positives** (patients with Schistosomiasis) | 4 studies 318 patients | cross-sectional (cohort type accuracy study) | not serious | not serious | not serious | serious ^a^ | none | 5 to 40 | 0 to 0 | 9 to 79 | 0 to 0 | 18 to 158 | 0 to 0 | ⨁⨁⨁◯ MODERATE |
|  |  |  |  |  |  |  |  | **5 more to 40 more TP in CCA1 cassette** | | **9 more to 79 more TP in CCA1 cassette** | | **18 more to 158 more TP in CCA1 cassette** | |  |
| **False negatives** (patients incorrectly classified as not having Schistosomiasis) |  |  |  |  |  |  |  | 10 to 45 | 50 to 50 | 21 to 91 | 100 to 100 | 42 to 182 | 200 to 200 |  |
|  |  |  |  |  |  |  |  | **5 fewer to 40 fewer FN in CCA1 cassette** | | **9 fewer to 79 fewer FN in CCA1 cassette** | | **18 fewer to 158 fewer FN in CCA1 cassette** | |  |
| **True negatives** (patients without Schistosomiasis) | 4 studies 673 patients | cross-sectional (cohort type accuracy study) | not serious | not serious | not serious | serious ^a^ | none | 418 to 931 | 0 to 0 | 396 to 882 | 0 to 0 | 352 to 784 | 0 to 0 | ⨁⨁⨁◯ MODERATE |
|  |  |  |  |  |  |  |  | **418 more to 931 more TN in CCA1 cassette** | | **396 more to 882 more TN in CCA1 cassette** | | **352 more to 784 more TN in CCA1 cassette** | |  |
| **False positives** (patients incorrectly classified as having Schistosomiasis) |  |  |  |  |  |  |  | 19 to 532 | 950 to 950 | 18 to 504 | 900 to 900 | 16 to 448 | 800 to 800 |  |
|  |  |  |  |  |  |  |  | **418 fewer to 931 fewer FP in CCA1 cassette** | | **396 fewer to 882 fewer FP in CCA1 cassette** | | **352 fewer to 784 fewer FP in CCA1 cassette** | |  |

##### Explanations

a. imperfect reference test

### Question: Should Colorimetric test vs. Urine Microscopy be used to diagnose Schistosomiasis in low endemic regions?

| Colorimetric test | | Urine Microscopy | |
| --- | --- | --- | --- |
| Sensitivity | 0.52 (95% CI: 0.49 to 0.56) | Sensitivity | -- (95% CI: -- to --) |
| Specificity | 0.75 (95% CI: 0.70 to 0.78) | Specificity | -- (95% CI: -- to --) |

| Prevalences | 5% | 10% | 20% |
| --- | --- | --- | --- |

| Outcome | № of studies (№ of patients) | Study design | Factors that may decrease certainty of evidence | | | | | Effect per 1,000 patients tested | | | | | | Test accuracy CoE |
| --- | --- | --- | --- | --- | --- | --- | --- | --- | --- | --- | --- | --- | --- | --- |
|  |  |  |  |  |  |  |  | pre-test probability of 5% | | pre-test probability of 10% | | pre-test probability of 20% | |  |
|  |  |  | Risk of bias | Indirectness | Inconsistency | Imprecision | Publication bias | Colorimetric test | Urine Microscopy | Colorimetric test | Urine Microscopy | Colorimetric test | Urine Microscopy |  |
| **True positives** (patients with Schistosomiasis) | 1 studies 791 patients | cross-sectional (cohort type accuracy study) | not serious | not serious | not serious | serious ^a^ | none | 26 (25 to 28) | 0 (0 to 0) | 52 (49 to 56) | 0 (0 to 0) | 104 (98 to 112) | 0 (0 to 0) | ⨁⨁⨁◯ MODERATE |
|  |  |  |  |  |  |  |  | **26 more TP in Colorimetric test** | | **52 more TP in Colorimetric test** | | **104 more TP in Colorimetric test** | |  |
| **False negatives** (patients incorrectly classified as not having Schistosomiasis) |  |  |  |  |  |  |  | 24 (22 to 25) | 50 (50 to 50) | 48 (44 to 51) | 100 (100 to 100) | 96 (88 to 102) | 200 (200 to 200) |  |
|  |  |  |  |  |  |  |  | **26 fewer FN in Colorimetric test** | | **52 fewer FN in Colorimetric test** | | **104 fewer FN in Colorimetric test** | |  |
| **True negatives** (patients without Schistosomiasis) | 1 studies 488 patients | cross-sectional (cohort type accuracy study) | not serious | not serious | not serious | serious ^a^ | none | 712 (665 to 741) | 0 (0 to 0) | 675 (630 to 702) | 0 (0 to 0) | 600 (560 to 624) | 0 (0 to 0) | ⨁⨁⨁◯ MODERATE |
|  |  |  |  |  |  |  |  | **712 more TN in Colorimetric test** | | **675 more TN in Colorimetric test** | | **600 more TN in Colorimetric test** | |  |
| **False positives** (patients incorrectly classified as having Schistosomiasis) |  |  |  |  |  |  |  | 238 (209 to 285) | 950 (950 to 950) | 225 (198 to 270) | 900 (900 to 900) | 200 (176 to 240) | 800 (800 to 800) |  |
|  |  |  |  |  |  |  |  | **712 fewer FP in Colorimetric test** | | **675 fewer FP in Colorimetric test** | | **600 fewer FP in Colorimetric test** | |  |

##### Explanations

a. imperfect reference test

### Question: Should COPT vs. duplicate KK be used to diagnose Schistosomiasis in low endemic regions?

| COPT | | duplicate KK | |
| --- | --- | --- | --- |
| Sensitivity | 0.80 (95% CI: 0.28 to 0.99) | Sensitivity | -- (95% CI: -- to --) |
| Specificity | 0.96 (95% CI: 0.94 to 0.97) | Specificity | -- (95% CI: -- to --) |

| Prevalences | 5% | 10% | 20% |
| --- | --- | --- | --- |

| Outcome | № of studies (№ of patients) | Study design | Factors that may decrease certainty of evidence | | | | | Effect per 1,000 patients tested | | | | | | Test accuracy CoE |
| --- | --- | --- | --- | --- | --- | --- | --- | --- | --- | --- | --- | --- | --- | --- |
|  |  |  |  |  |  |  |  | pre-test probability of 5% | | pre-test probability of 10% | | pre-test probability of 20% | |  |
|  |  |  | Risk of bias | Indirectness | Inconsistency | Imprecision | Publication bias | COPT | duplicate KK | COPT | duplicate KK | COPT | duplicate KK |  |
| **True positives** (patients with Schistosomiasis) | 1 studies 5 patients | cross-sectional (cohort type accuracy study) | not serious | not serious | not serious | serious ^a^ | none | 40 (14 to 50) | 0 (0 to 0) | 80 (28 to 99) | 0 (0 to 0) | 160 (56 to 198) | 0 (0 to 0) | ⨁⨁⨁◯ MODERATE |
|  |  |  |  |  |  |  |  | **40 more TP in COPT** | | **80 more TP in COPT** | | **160 more TP in COPT** | |  |
| **False negatives** (patients incorrectly classified as not having Schistosomiasis) |  |  |  |  |  |  |  | 10 (0 to 36) | 50 (50 to 50) | 20 (1 to 72) | 100 (100 to 100) | 40 (2 to 144) | 200 (200 to 200) |  |
|  |  |  |  |  |  |  |  | **40 fewer FN in COPT** | | **80 fewer FN in COPT** | | **160 fewer FN in COPT** | |  |
| **True negatives** (patients without Schistosomiasis) | 1 studies 567 patients | cross-sectional (cohort type accuracy study) | not serious | not serious | not serious | serious ^a^ | none | 912 (893 to 922) | 0 (0 to 0) | 864 (846 to 873) | 0 (0 to 0) | 768 (752 to 776) | 0 (0 to 0) | ⨁⨁⨁◯ MODERATE |
|  |  |  |  |  |  |  |  | **912 more TN in COPT** | | **864 more TN in COPT** | | **768 more TN in COPT** | |  |
| **False positives** (patients incorrectly classified as having Schistosomiasis) |  |  |  |  |  |  |  | 38 (28 to 57) | 950 (950 to 950) | 36 (27 to 54) | 900 (900 to 900) | 32 (24 to 48) | 800 (800 to 800) |  |
|  |  |  |  |  |  |  |  | **912 fewer FP in COPT** | | **864 fewer FP in COPT** | | **768 fewer FP in COPT** | |  |

##### Explanations

a. imperfect reference test

### Question: Should DDIA vs. Urine Microscopy be used to diagnose Schistosomiasis in low endemic regions?

| DDIA | | Urine Microscopy | |
| --- | --- | --- | --- |
| Sensitivity | 0.60 (95% CI: 0.49 to 0.70) | Sensitivity | -- (95% CI: -- to --) |
| Specificity | 0.61 (95% CI: 0.48 to 0.74) | Specificity | -- (95% CI: -- to --) |

| Prevalences | 5% | 10% | 20% |
| --- | --- | --- | --- |

| Outcome | № of studies (№ of patients) | Study design | Factors that may decrease certainty of evidence | | | | | Effect per 1,000 patients tested | | | | | | Test accuracy CoE |
| --- | --- | --- | --- | --- | --- | --- | --- | --- | --- | --- | --- | --- | --- | --- |
|  |  |  |  |  |  |  |  | pre-test probability of 5% | | pre-test probability of 10% | | pre-test probability of 20% | |  |
|  |  |  | Risk of bias | Indirectness | Inconsistency | Imprecision | Publication bias | DDIA | Urine Microscopy | DDIA | Urine Microscopy | DDIA | Urine Microscopy |  |
| **True positives** (patients with Schistosomiasis) | 1 studies 89 patients | cross-sectional (cohort type accuracy study) | not serious | not serious | not serious | serious ^a^ | none | 30 (25 to 35) | 0 (0 to 0) | 60 (49 to 70) | 0 (0 to 0) | 120 (98 to 140) | 0 (0 to 0) | ⨁⨁⨁◯ MODERATE |
|  |  |  |  |  |  |  |  | **30 more TP in DDIA** | | **60 more TP in DDIA** | | **120 more TP in DDIA** | |  |
| **False negatives** (patients incorrectly classified as not having Schistosomiasis) |  |  |  |  |  |  |  | 20 (15 to 25) | 50 (50 to 50) | 40 (30 to 51) | 100 (100 to 100) | 80 (60 to 102) | 200 (200 to 200) |  |
|  |  |  |  |  |  |  |  | **30 fewer FN in DDIA** | | **60 fewer FN in DDIA** | | **120 fewer FN in DDIA** | |  |
| **True negatives** (patients without Schistosomiasis) | 1 studies 57 patients | cross-sectional (cohort type accuracy study) | not serious | not serious | not serious | serious ^a^ | none | 580 (456 to 703) | 0 (0 to 0) | 549 (432 to 666) | 0 (0 to 0) | 488 (384 to 592) | 0 (0 to 0) | ⨁⨁⨁◯ MODERATE |
|  |  |  |  |  |  |  |  | **580 more TN in DDIA** | | **549 more TN in DDIA** | | **488 more TN in DDIA** | |  |
| **False positives** (patients incorrectly classified as having Schistosomiasis) |  |  |  |  |  |  |  | 370 (247 to 494) | 950 (950 to 950) | 351 (234 to 468) | 900 (900 to 900) | 312 (208 to 416) | 800 (800 to 800) |  |
|  |  |  |  |  |  |  |  | **580 fewer FP in DDIA** | | **549 fewer FP in DDIA** | | **488 fewer FP in DDIA** | |  |

##### Explanations

a. imperfect reference test

### Question: Should Helmintex vs. duplicate KK be used to diagnose Schistosomiasis in low endemic regions?

| Helmintex | | duplicate KK | |
| --- | --- | --- | --- |
| Sensitivity | 0.98 (95% CI: 0.90 to 1.00) | Sensitivity | -- (95% CI: -- to --) |
| Specificity | 0.67 (95% CI: 0.62 to 0.72) | Specificity | -- (95% CI: -- to --) |

| Prevalences | 5% | 10% | 20% |
| --- | --- | --- | --- |

| Outcome | № of studies (№ of patients) | Study design | Factors that may decrease certainty of evidence | | | | | Effect per 1,000 patients tested | | | | | | Test accuracy CoE |
| --- | --- | --- | --- | --- | --- | --- | --- | --- | --- | --- | --- | --- | --- | --- |
|  |  |  |  |  |  |  |  | pre-test probability of 5% | | pre-test probability of 10% | | pre-test probability of 20% | |  |
|  |  |  | Risk of bias | Indirectness | Inconsistency | Imprecision | Publication bias | Helmintex | duplicate KK | Helmintex | duplicate KK | Helmintex | duplicate KK |  |
| **True positives** (patients with Schistosomiasis) | 1 studies 55 patients | cross-sectional (cohort type accuracy study) | not serious | not serious | not serious | serious ^a^ | none | 49 (45 to 50) | 0 (0 to 0) | 98 (90 to 100) | 0 (0 to 0) | 196 (180 to 200) | 0 (0 to 0) | ⨁⨁⨁◯ MODERATE |
|  |  |  |  |  |  |  |  | **49 more TP in Helmintex** | | **98 more TP in Helmintex** | | **196 more TP in Helmintex** | |  |
| **False negatives** (patients incorrectly classified as not having Schistosomiasis) |  |  |  |  |  |  |  | 1 (0 to 5) | 50 (50 to 50) | 2 (0 to 10) | 100 (100 to 100) | 4 (0 to 20) | 200 (200 to 200) |  |
|  |  |  |  |  |  |  |  | **49 fewer FN in Helmintex** | | **98 fewer FN in Helmintex** | | **196 fewer FN in Helmintex** | |  |
| **True negatives** (patients without Schistosomiasis) | 1 studies 406 patients | cross-sectional (cohort type accuracy study) | not serious | not serious | not serious | serious ^a^ | none | 637 (589 to 684) | 0 (0 to 0) | 603 (558 to 648) | 0 (0 to 0) | 536 (496 to 576) | 0 (0 to 0) | ⨁⨁⨁◯ MODERATE |
|  |  |  |  |  |  |  |  | **637 more TN in Helmintex** | | **603 more TN in Helmintex** | | **536 more TN in Helmintex** | |  |
| **False positives** (patients incorrectly classified as having Schistosomiasis) |  |  |  |  |  |  |  | 313 (266 to 361) | 950 (950 to 950) | 297 (252 to 342) | 900 (900 to 900) | 264 (224 to 304) | 800 (800 to 800) |  |
|  |  |  |  |  |  |  |  | **637 fewer FP in Helmintex** | | **603 fewer FP in Helmintex** | | **536 fewer FP in Helmintex** | |  |

##### Explanations

a. imperfect reference test

### Question: Should IHA vs. triplicate KK be used to diagnose Schistosomiasis in low endemic regions?

| IHA | | triplicate KK | |
| --- | --- | --- | --- |
| Sensitivity | 0.90 to 0.91 | Sensitivity | -- to -- |
| Specificity | 0.00 to -- | Specificity | -- to -- |

| Prevalences | 5% | 10% | 20% |
| --- | --- | --- | --- |

| Outcome | № of studies (№ of patients) | Study design | Factors that may decrease certainty of evidence | | | | | Effect per 1,000 patients tested | | | | | | Test accuracy CoE |
| --- | --- | --- | --- | --- | --- | --- | --- | --- | --- | --- | --- | --- | --- | --- |
|  |  |  |  |  |  |  |  | pre-test probability of 5% | | pre-test probability of 10% | | pre-test probability of 20% | |  |
|  |  |  | Risk of bias | Indirectness | Inconsistency | Imprecision | Publication bias | IHA | triplicate KK | IHA | triplicate KK | IHA | triplicate KK |  |
| **True positives** (patients with Schistosomiasis) | 2 studies 198 patients | cross-sectional (cohort type accuracy study) | not serious | not serious | not serious | serious ^a^ | none | 45 to 46 | 0 to 0 | 90 to 91 | 0 to 0 | 180 to 182 | 0 to 0 | ⨁⨁⨁◯ MODERATE |
|  |  |  |  |  |  |  |  | **45 more to 46 more TP in IHA** | | **90 more to 91 more TP in IHA** | | **180 more to 182 more TP in IHA** | |  |
| **False negatives** (patients incorrectly classified as not having Schistosomiasis) |  |  |  |  |  |  |  | 4 to 5 | 50 to 50 | 9 to 10 | 100 to 100 | 18 to 20 | 200 to 200 |  |
|  |  |  |  |  |  |  |  | **45 fewer to 46 fewer FN in IHA** | | **90 fewer to 91 fewer FN in IHA** | | **180 fewer to 182 fewer FN in IHA** | |  |
| **True negatives** (patients without Schistosomiasis) | 2 studies 5 patients | cross-sectional (cohort type accuracy study) | not serious | not serious | not serious | serious ^a^ | none | 0 to 0 | 0 to 0 | 0 to 0 | 0 to 0 | 0 to 0 | 0 to 0 | ⨁⨁⨁◯ MODERATE |
|  |  |  |  |  |  |  |  | **0 fewer to 0 fewer TN in IHA** | | **0 fewer to 0 fewer TN in IHA** | | **0 fewer to 0 fewer TN in IHA** | |  |
| **False positives** (patients incorrectly classified as having Schistosomiasis) |  |  |  |  |  |  |  | 950 to 950 | 950 to 950 | 900 to 900 | 900 to 900 | 800 to 800 | 800 to 800 |  |
|  |  |  |  |  |  |  |  | **0 fewer to 0 fewer FP in IHA** | | **0 fewer to 0 fewer FP in IHA** | | **0 fewer to 0 fewer FP in IHA** | |  |

##### Explanations

a. imperfect reference test

### Question: Should IHA vs. Urine Microscopy be used to diagnose Schistosomiasis in low endemic regions?

| IHA | | Urine Microscopy | |
| --- | --- | --- | --- |
| Sensitivity | 0.74 (95% CI: 0.64 to 0.83) | Sensitivity | -- (95% CI: -- to --) |
| Specificity | 0.72 (95% CI: 0.58 to 0.83) | Specificity | -- (95% CI: -- to --) |

| Prevalences | 5% | 10% | 20% |
| --- | --- | --- | --- |

| Outcome | № of studies (№ of patients) | Study design | Factors that may decrease certainty of evidence | | | | | Effect per 1,000 patients tested | | | | | | Test accuracy CoE |
| --- | --- | --- | --- | --- | --- | --- | --- | --- | --- | --- | --- | --- | --- | --- |
|  |  |  |  |  |  |  |  | pre-test probability of 5% | | pre-test probability of 10% | | pre-test probability of 20% | |  |
|  |  |  | Risk of bias | Indirectness | Inconsistency | Imprecision | Publication bias | IHA | Urine Microscopy | IHA | Urine Microscopy | IHA | Urine Microscopy |  |
| **True positives** (patients with Schistosomiasis) | 1 studies 89 patients | cross-sectional (cohort type accuracy study) | not serious | not serious | not serious | serious ^a^ | none | 37 (32 to 42) | 0 (0 to 0) | 74 (64 to 83) | 0 (0 to 0) | 148 (128 to 166) | 0 (0 to 0) | ⨁⨁⨁◯ MODERATE |
|  |  |  |  |  |  |  |  | **37 more TP in IHA** | | **74 more TP in IHA** | | **148 more TP in IHA** | |  |
| **False negatives** (patients incorrectly classified as not having Schistosomiasis) |  |  |  |  |  |  |  | 13 (8 to 18) | 50 (50 to 50) | 26 (17 to 36) | 100 (100 to 100) | 52 (34 to 72) | 200 (200 to 200) |  |
|  |  |  |  |  |  |  |  | **37 fewer FN in IHA** | | **74 fewer FN in IHA** | | **148 fewer FN in IHA** | |  |
| **True negatives** (patients without Schistosomiasis) | 1 studies 57 patients | cross-sectional (cohort type accuracy study) | not serious | not serious | not serious | serious ^a^ | none | 684 (551 to 789) | 0 (0 to 0) | 648 (522 to 747) | 0 (0 to 0) | 576 (464 to 664) | 0 (0 to 0) | ⨁⨁⨁◯ MODERATE |
|  |  |  |  |  |  |  |  | **684 more TN in IHA** | | **648 more TN in IHA** | | **576 more TN in IHA** | |  |
| **False positives** (patients incorrectly classified as having Schistosomiasis) |  |  |  |  |  |  |  | 266 (161 to 399) | 950 (950 to 950) | 252 (153 to 378) | 900 (900 to 900) | 224 (136 to 336) | 800 (800 to 800) |  |
|  |  |  |  |  |  |  |  | **684 fewer FP in IHA** | | **648 fewer FP in IHA** | | **576 fewer FP in IHA** | |  |

##### Explanations

a. imperfect reference test

### Question: Should LAMP be used to diagnose triplicate KK in low endemic regions?

| Sensitivity | 0.95 to 0.97 |
| --- | --- |
| Specificity | -- to 1.00 |

| Prevalences | 5% | 10% | 20% |
| --- | --- | --- | --- |

| Outcome | № of studies (№ of patients) | Study design | Factors that may decrease certainty of evidence | | | | | Effect per 1,000 patients tested | | | Test accuracy CoE |
| --- | --- | --- | --- | --- | --- | --- | --- | --- | --- | --- | --- |
|  |  |  | Risk of bias | Indirectness | Inconsistency | Imprecision | Publication bias | pre-test probability of 5% | pre-test probability of 10% | pre-test probability of 20% |  |
| **True positives** (patients with triplicate KK) | 2 studies 286 patients | cross-sectional (cohort type accuracy study) | not serious | not serious | not serious | serious ^a^ | none | 48 to 49 | 95 to 97 | 190 to 194 | ⨁⨁⨁◯ MODERATE |
| **False negatives** (patients incorrectly classified as not having triplicate KK) |  |  |  |  |  |  |  | 1 to 2 | 3 to 5 | 6 to 10 |  |
| **True negatives** (patients without triplicate KK) | 2 studies 207 patients | cross-sectional (cohort type accuracy study) | not serious | not serious | not serious | serious ^a^ | none | 0 to 950 | 0 to 900 | 0 to 800 | ⨁⨁⨁◯ MODERATE |
| **False positives** (patients incorrectly classified as having triplicate KK) |  |  |  |  |  |  |  | 0 to 950 | 0 to 900 | 0 to 800 |  |

##### Explanations

a. imperfect reference test

### Question: Should LAMP vs. Urine Microscopy be used to diagnose Schistosomiasis in low endemic regions?

| LAMP | | Urine Microscopy | |
| --- | --- | --- | --- |
| Sensitivity | 0.72 to 0.86 | Sensitivity | -- to -- |
| Specificity | 0.39 to 0.88 | Specificity | -- to -- |

| Prevalences | 5% | 10% | 20% |
| --- | --- | --- | --- |

| Outcome | № of studies (№ of patients) | Study design | Factors that may decrease certainty of evidence | | | | | Effect per 1,000 patients tested | | | | | | Test accuracy CoE |
| --- | --- | --- | --- | --- | --- | --- | --- | --- | --- | --- | --- | --- | --- | --- |
|  |  |  |  |  |  |  |  | pre-test probability of 5% | | pre-test probability of 10% | | pre-test probability of 20% | |  |
|  |  |  | Risk of bias | Indirectness | Inconsistency | Imprecision | Publication bias | LAMP | Urine Microscopy | LAMP | Urine Microscopy | LAMP | Urine Microscopy |  |
| **True positives** (patients with Schistosomiasis) | 2 studies 112 patients | cross-sectional (cohort type accuracy study) | not serious | not serious | not serious | serious ^a^ | none | 36 to 43 | 0 to 0 | 72 to 86 | 0 to 0 | 144 to 172 | 0 to 0 | ⨁⨁⨁◯ MODERATE |
|  |  |  |  |  |  |  |  | **36 more to 43 more TP in LAMP** | | **72 more to 86 more TP in LAMP** | | **144 more to 172 more TP in LAMP** | |  |
| **False negatives** (patients incorrectly classified as not having Schistosomiasis) |  |  |  |  |  |  |  | 7 to 14 | 50 to 50 | 14 to 28 | 100 to 100 | 28 to 56 | 200 to 200 |  |
|  |  |  |  |  |  |  |  | **36 fewer to 43 fewer FN in LAMP** | | **72 fewer to 86 fewer FN in LAMP** | | **144 fewer to 172 fewer FN in LAMP** | |  |
| **True negatives** (patients without Schistosomiasis) | 2 studies 154 patients | cross-sectional (cohort type accuracy study) | not serious | not serious | not serious | serious ^a^ | none | 371 to 836 | 0 to 0 | 351 to 792 | 0 to 0 | 312 to 704 | 0 to 0 | ⨁⨁⨁◯ MODERATE |
|  |  |  |  |  |  |  |  | **371 more to 836 more TN in LAMP** | | **351 more to 792 more TN in LAMP** | | **312 more to 704 more TN in LAMP** | |  |
| **False positives** (patients incorrectly classified as having Schistosomiasis) |  |  |  |  |  |  |  | 114 to 579 | 950 to 950 | 108 to 549 | 900 to 900 | 96 to 488 | 800 to 800 |  |
|  |  |  |  |  |  |  |  | **371 fewer to 836 fewer FP in LAMP** | | **351 fewer to 792 fewer FP in LAMP** | | **312 fewer to 704 fewer FP in LAMP** | |  |

##### Explanations

a. imperfect reference test

### Question: Should PCR vs. CCA1 cassette be used to diagnose Schistosomiasis in low endemic regions?

| PCR | | CCA1 cassette | |
| --- | --- | --- | --- |
| Sensitivity | 0.85 (95% CI: 0.78 to 0.90) | Sensitivity | -- (95% CI: -- to --) |
| Specificity | 0.56 (95% CI: 0.46 to 0.65) | Specificity | -- (95% CI: -- to --) |

| Prevalences | 5% | 10% | 20% |
| --- | --- | --- | --- |

| Outcome | № of studies (№ of patients) | Study design | Factors that may decrease certainty of evidence | | | | | Effect per 1,000 patients tested | | | | | | Test accuracy CoE |
| --- | --- | --- | --- | --- | --- | --- | --- | --- | --- | --- | --- | --- | --- | --- |
|  |  |  |  |  |  |  |  | pre-test probability of 5% | | pre-test probability of 10% | | pre-test probability of 20% | |  |
|  |  |  | Risk of bias | Indirectness | Inconsistency | Imprecision | Publication bias | PCR | CCA1 cassette | PCR | CCA1 cassette | PCR | CCA1 cassette |  |
| **True positives** (patients with Schistosomiasis) | 1 studies 147 patients | cross-sectional (cohort type accuracy study) | not serious | not serious | not serious | serious ^a^ | none | 43 (39 to 45) | 0 (0 to 0) | 85 (78 to 90) | 0 (0 to 0) | 170 (156 to 180) | 0 (0 to 0) | ⨁⨁⨁◯ MODERATE |
|  |  |  |  |  |  |  |  | **43 more TP in PCR** | | **85 more TP in PCR** | | **170 more TP in PCR** | |  |
| **False negatives** (patients incorrectly classified as not having Schistosomiasis) |  |  |  |  |  |  |  | 7 (5 to 11) | 50 (50 to 50) | 15 (10 to 22) | 100 (100 to 100) | 30 (20 to 44) | 200 (200 to 200) |  |
|  |  |  |  |  |  |  |  | **43 fewer FN in PCR** | | **85 fewer FN in PCR** | | **170 fewer FN in PCR** | |  |
| **True negatives** (patients without Schistosomiasis) | 1 studies 111 patients | cross-sectional (cohort type accuracy study) | not serious | not serious | not serious | serious ^a^ | none | 532 (437 to 617) | 0 (0 to 0) | 504 (414 to 585) | 0 (0 to 0) | 448 (368 to 520) | 0 (0 to 0) | ⨁⨁⨁◯ MODERATE |
|  |  |  |  |  |  |  |  | **532 more TN in PCR** | | **504 more TN in PCR** | | **448 more TN in PCR** | |  |
| **False positives** (patients incorrectly classified as having Schistosomiasis) |  |  |  |  |  |  |  | 418 (333 to 513) | 950 (950 to 950) | 396 (315 to 486) | 900 (900 to 900) | 352 (280 to 432) | 800 (800 to 800) |  |
|  |  |  |  |  |  |  |  | **532 fewer FP in PCR** | | **504 fewer FP in PCR** | | **448 fewer FP in PCR** | |  |

##### Explanations

a. imperfect reference test

### Question: Should PCR vs. KK be used to diagnose Schistosomiasis in low endemic regions?

| PCR | | KK | |
| --- | --- | --- | --- |
| Sensitivity | 0.97 to 1.00 | Sensitivity | -- to -- |
| Specificity | 0.59 to 0.83 | Specificity | -- to -- |

| Prevalences | 5% | 10% | 20% |
| --- | --- | --- | --- |

| Outcome | № of studies (№ of patients) | Study design | Factors that may decrease certainty of evidence | | | | | Effect per 1,000 patients tested | | | | | | Test accuracy CoE |
| --- | --- | --- | --- | --- | --- | --- | --- | --- | --- | --- | --- | --- | --- | --- |
|  |  |  |  |  |  |  |  | pre-test probability of 5% | | pre-test probability of 10% | | pre-test probability of 20% | |  |
|  |  |  | Risk of bias | Indirectness | Inconsistency | Imprecision | Publication bias | PCR | KK | PCR | KK | PCR | KK |  |
| **True positives** (patients with Schistosomiasis) | 3 studies 319 patients | cross-sectional (cohort type accuracy study) | not serious | not serious | not serious | serious ^a^ | none | 49 to 50 | 0 to 0 | 97 to 100 | 0 to 0 | 194 to 200 | 0 to 0 | ⨁⨁⨁◯ MODERATE |
|  |  |  |  |  |  |  |  | **49 more to 50 more TP in PCR** | | **97 more to 100 more TP in PCR** | | **194 more to 200 more TP in PCR** | |  |
| **False negatives** (patients incorrectly classified as not having Schistosomiasis) |  |  |  |  |  |  |  | 0 to 1 | 50 to 50 | 0 to 3 | 100 to 100 | 0 to 6 | 200 to 200 |  |
|  |  |  |  |  |  |  |  | **49 fewer to 50 fewer FN in PCR** | | **97 fewer to 100 fewer FN in PCR** | | **194 fewer to 200 fewer FN in PCR** | |  |
| **True negatives** (patients without Schistosomiasis) | 3 studies 232 patients | cross-sectional (cohort type accuracy study) | not serious | not serious | not serious | serious ^a^ | none | 561 to 789 | 0 to 0 | 531 to 747 | 0 to 0 | 472 to 664 | 0 to 0 | ⨁⨁⨁◯ MODERATE |
|  |  |  |  |  |  |  |  | **561 more to 789 more TN in PCR** | | **531 more to 747 more TN in PCR** | | **472 more to 664 more TN in PCR** | |  |
| **False positives** (patients incorrectly classified as having Schistosomiasis) |  |  |  |  |  |  |  | 161 to 389 | 950 to 950 | 153 to 369 | 900 to 900 | 136 to 328 | 800 to 800 |  |
|  |  |  |  |  |  |  |  | **561 fewer to 789 fewer FP in PCR** | | **531 fewer to 747 fewer FP in PCR** | | **472 fewer to 664 fewer FP in PCR** | |  |

##### Explanations

a. imperfect reference test

### Question: Should CAA vs. lumped KK be used to diagnose Schistosomiasis in low endemic regions?

| CAA | | lumped KK | |
| --- | --- | --- | --- |
| Sensitivity | 0.10 to 0.96 | Sensitivity | -- to -- |
| Specificity | 0.37 to 0.99 | Specificity | -- to -- |

| Prevalences | 5% | 10% | 20% |
| --- | --- | --- | --- |

| Outcome | № of studies (№ of patients) | Study design | Factors that may decrease certainty of evidence | | | | | Effect per 1,000 patients tested | | | | | | Test accuracy CoE |
| --- | --- | --- | --- | --- | --- | --- | --- | --- | --- | --- | --- | --- | --- | --- |
|  |  |  |  |  |  |  |  | pre-test probability of 5% | | pre-test probability of 10% | | pre-test probability of 20% | |  |
|  |  |  | Risk of bias | Indirectness | Inconsistency | Imprecision | Publication bias | CAA | lumped KK | CAA | lumped KK | CAA | lumped KK |  |
| **True positives** (patients with Schistosomiasis) | 4 studies 524 patients | cross-sectional (cohort type accuracy study) | not serious | not serious | not serious | serious ^a^ | none | 5 to 48 | 0 to 0 | 10 to 96 | 0 to 0 | 20 to 192 | 0 to 0 | ⨁⨁⨁◯ MODERATE |
|  |  |  |  |  |  |  |  | **5 more to 48 more TP in CAA** | | **10 more to 96 more TP in CAA** | | **20 more to 192 more TP in CAA** | |  |
| **False negatives** (patients incorrectly classified as not having Schistosomiasis) |  |  |  |  |  |  |  | 2 to 45 | 50 to 50 | 4 to 90 | 100 to 100 | 8 to 180 | 200 to 200 |  |
|  |  |  |  |  |  |  |  | **5 fewer to 48 fewer FN in CAA** | | **10 fewer to 96 fewer FN in CAA** | | **20 fewer to 192 fewer FN in CAA** | |  |
| **True negatives** (patients without Schistosomiasis) | 4 studies 683 patients | cross-sectional (cohort type accuracy study) | not serious | not serious | not serious | serious ^a^ | none | 352 to 941 | 0 to 0 | 333 to 891 | 0 to 0 | 296 to 792 | 0 to 0 | ⨁⨁⨁◯ MODERATE |
|  |  |  |  |  |  |  |  | **352 more to 941 more TN in CAA** | | **333 more to 891 more TN in CAA** | | **296 more to 792 more TN in CAA** | |  |
| **False positives** (patients incorrectly classified as having Schistosomiasis) |  |  |  |  |  |  |  | 9 to 598 | 950 to 950 | 9 to 567 | 900 to 900 | 8 to 504 | 800 to 800 |  |
|  |  |  |  |  |  |  |  | **352 fewer to 941 fewer FP in CAA** | | **333 fewer to 891 fewer FP in CAA** | | **296 fewer to 792 fewer FP in CAA** | |  |

##### Explanations

a. imperfect reference test

### Question: Should CCA1 cassette vs. 16 KK be used to diagnose Schistosomiasis in low endemic regions?

| CCA1 cassette | | 16 KK | |
| --- | --- | --- | --- |
| Sensitivity | 0.81 (95% CI: 0.63 to 0.93) | Sensitivity | -- (95% CI: -- to --) |
| Specificity | 0.71 (95% CI: 0.64 to 0.77) | Specificity | -- (95% CI: -- to --) |

| Prevalences | 5% | 10% | 20% |
| --- | --- | --- | --- |

| Outcome | № of studies (№ of patients) | Study design | Factors that may decrease certainty of evidence | | | | | Effect per 1,000 patients tested | | | | | | Test accuracy CoE |
| --- | --- | --- | --- | --- | --- | --- | --- | --- | --- | --- | --- | --- | --- | --- |
|  |  |  |  |  |  |  |  | pre-test probability of 5% | | pre-test probability of 10% | | pre-test probability of 20% | |  |
|  |  |  | Risk of bias | Indirectness | Inconsistency | Imprecision | Publication bias | CCA1 cassette | 16 KK | CCA1 cassette | 16 KK | CCA1 cassette | 16 KK |  |
| **True positives** (patients with Schistosomiasis) | 1 studies 31 patients | cross-sectional (cohort type accuracy study) | not serious | not serious | not serious | serious ^a^ | none | 41 (32 to 47) | 0 (0 to 0) | 81 (63 to 93) | 0 (0 to 0) | 162 (126 to 186) | 0 (0 to 0) | ⨁⨁⨁◯ MODERATE |
|  |  |  |  |  |  |  |  | **41 more TP in CCA1 cassette** | | **81 more TP in CCA1 cassette** | | **162 more TP in CCA1 cassette** | |  |
| **False negatives** (patients incorrectly classified as not having Schistosomiasis) |  |  |  |  |  |  |  | 9 (3 to 18) | 50 (50 to 50) | 19 (7 to 37) | 100 (100 to 100) | 38 (14 to 74) | 200 (200 to 200) |  |
|  |  |  |  |  |  |  |  | **41 fewer FN in CCA1 cassette** | | **81 fewer FN in CCA1 cassette** | | **162 fewer FN in CCA1 cassette** | |  |
| **True negatives** (patients without Schistosomiasis) | 1 studies 186 patients | cross-sectional (cohort type accuracy study) | not serious | not serious | not serious | serious ^a^ | none | 675 (608 to 731) | 0 (0 to 0) | 639 (576 to 693) | 0 (0 to 0) | 568 (512 to 616) | 0 (0 to 0) | ⨁⨁⨁◯ MODERATE |
|  |  |  |  |  |  |  |  | **675 more TN in CCA1 cassette** | | **639 more TN in CCA1 cassette** | | **568 more TN in CCA1 cassette** | |  |
| **False positives** (patients incorrectly classified as having Schistosomiasis) |  |  |  |  |  |  |  | 275 (219 to 342) | 950 (950 to 950) | 261 (207 to 324) | 900 (900 to 900) | 232 (184 to 288) | 800 (800 to 800) |  |
|  |  |  |  |  |  |  |  | **675 fewer FP in CCA1 cassette** | | **639 fewer FP in CCA1 cassette** | | **568 fewer FP in CCA1 cassette** | |  |

##### Explanations

a. imperfect reference test

### Question: Should CCA1 cassette vs. Helmintex be used to diagnose Schistosomiasis in low endemic regions?

| CCA1 cassette | | Helmintex | |
| --- | --- | --- | --- |
| Sensitivity | 0.57 (95% CI: 0.42 to 0.70) | Sensitivity | -- (95% CI: -- to --) |
| Specificity | 0.71 (95% CI: 0.64 to 0.78) | Specificity | -- (95% CI: -- to --) |

| Prevalences | 5% | 10% | 20% |
| --- | --- | --- | --- |

| Outcome | № of studies (№ of patients) | Study design | Factors that may decrease certainty of evidence | | | | | Effect per 1,000 patients tested | | | | | | Test accuracy CoE |
| --- | --- | --- | --- | --- | --- | --- | --- | --- | --- | --- | --- | --- | --- | --- |
|  |  |  |  |  |  |  |  | pre-test probability of 5% | | pre-test probability of 10% | | pre-test probability of 20% | |  |
|  |  |  | Risk of bias | Indirectness | Inconsistency | Imprecision | Publication bias | CCA1 cassette | Helmintex | CCA1 cassette | Helmintex | CCA1 cassette | Helmintex |  |
| **True positives** (patients with Schistosomiasis) | 1 studies 53 patients | cross-sectional (cohort type accuracy study) | not serious | not serious | not serious | serious ^a^ | none | 28 (21 to 35) | 0 (0 to 0) | 57 (42 to 70) | 0 (0 to 0) | 114 (84 to 140) | 0 (0 to 0) | ⨁⨁⨁◯ MODERATE |
|  |  |  |  |  |  |  |  | **28 more TP in CCA1 cassette** | | **57 more TP in CCA1 cassette** | | **114 more TP in CCA1 cassette** | |  |
| **False negatives** (patients incorrectly classified as not having Schistosomiasis) |  |  |  |  |  |  |  | 22 (15 to 29) | 50 (50 to 50) | 43 (30 to 58) | 100 (100 to 100) | 86 (60 to 116) | 200 (200 to 200) |  |
|  |  |  |  |  |  |  |  | **28 fewer FN in CCA1 cassette** | | **57 fewer FN in CCA1 cassette** | | **114 fewer FN in CCA1 cassette** | |  |
| **True negatives** (patients without Schistosomiasis) | 1 studies 161 patients | cross-sectional (cohort type accuracy study) | not serious | not serious | not serious | serious ^a^ | none | 675 (608 to 741) | 0 (0 to 0) | 639 (576 to 702) | 0 (0 to 0) | 568 (512 to 624) | 0 (0 to 0) | ⨁⨁⨁◯ MODERATE |
|  |  |  |  |  |  |  |  | **675 more TN in CCA1 cassette** | | **639 more TN in CCA1 cassette** | | **568 more TN in CCA1 cassette** | |  |
| **False positives** (patients incorrectly classified as having Schistosomiasis) |  |  |  |  |  |  |  | 275 (209 to 342) | 950 (950 to 950) | 261 (198 to 324) | 900 (900 to 900) | 232 (176 to 288) | 800 (800 to 800) |  |
|  |  |  |  |  |  |  |  | **675 fewer FP in CCA1 cassette** | | **639 fewer FP in CCA1 cassette** | | **568 fewer FP in CCA1 cassette** | |  |

##### Explanations

a. imperfect reference test

### Question: Should CCA1 cassette vs. lumped KK be used to diagnose Schistosomiasis in low endemic regions?

| CCA1 cassette | | lumped KK | |
| --- | --- | --- | --- |
| Sensitivity | 0.07 to 0.99 | Sensitivity | -- to -- |
| Specificity | 0.03 to 0.93 | Specificity | -- to -- |

| Prevalences | 5% | 10% | 20% |
| --- | --- | --- | --- |

| Outcome | № of studies (№ of patients) | Study design | Factors that may decrease certainty of evidence | | | | | Effect per 1,000 patients tested | | | | | | Test accuracy CoE |
| --- | --- | --- | --- | --- | --- | --- | --- | --- | --- | --- | --- | --- | --- | --- |
|  |  |  |  |  |  |  |  | pre-test probability of 5% | | pre-test probability of 10% | | pre-test probability of 20% | |  |
|  |  |  | Risk of bias | Indirectness | Inconsistency | Imprecision | Publication bias | CCA1 cassette | lumped KK | CCA1 cassette | lumped KK | CCA1 cassette | lumped KK |  |
| **True positives** (patients with Schistosomiasis) | 37 studies 4819 patients | cross-sectional (cohort type accuracy study) | not serious | not serious | not serious | serious ^a^ | none | 4 to 50 | 0 to 0 | 7 to 99 | 0 to 0 | 14 to 198 | 0 to 0 | ⨁⨁⨁◯ MODERATE |
|  |  |  |  |  |  |  |  | **4 more to 50 more TP in CCA1 cassette** | | **7 more to 99 more TP in CCA1 cassette** | | **14 more to 198 more TP in CCA1 cassette** | |  |
| **False negatives** (patients incorrectly classified as not having Schistosomiasis) |  |  |  |  |  |  |  | 0 to 46 | 50 to 50 | 1 to 93 | 100 to 100 | 2 to 186 | 200 to 200 |  |
|  |  |  |  |  |  |  |  | **4 fewer to 50 fewer FN in CCA1 cassette** | | **7 fewer to 99 fewer FN in CCA1 cassette** | | **14 fewer to 198 fewer FN in CCA1 cassette** | |  |
| **True negatives** (patients without Schistosomiasis) | 37 studies 7416 patients | cross-sectional (cohort type accuracy study) | not serious | not serious | not serious | serious ^a^ | none | 28 to 884 | 0 to 0 | 27 to 837 | 0 to 0 | 24 to 744 | 0 to 0 | ⨁⨁⨁◯ MODERATE |
|  |  |  |  |  |  |  |  | **28 more to 884 more TN in CCA1 cassette** | | **27 more to 837 more TN in CCA1 cassette** | | **24 more to 744 more TN in CCA1 cassette** | |  |
| **False positives** (patients incorrectly classified as having Schistosomiasis) |  |  |  |  |  |  |  | 66 to 922 | 950 to 950 | 63 to 873 | 900 to 900 | 56 to 776 | 800 to 800 |  |
|  |  |  |  |  |  |  |  | **28 fewer to 884 fewer FP in CCA1 cassette** | | **27 fewer to 837 fewer FP in CCA1 cassette** | | **24 fewer to 744 fewer FP in CCA1 cassette** | |  |

##### Explanations

a. imperfect reference test

### Question: Should CCA1 cassette vs. RT-PCR be used to diagnose Schistosomiasis in low endemic regions?

| CCA1 cassette | | RT-PCR | |
| --- | --- | --- | --- |
| Sensitivity | 0.65 (95% CI: 0.55 to 0.74) | Sensitivity | -- (95% CI: -- to --) |
| Specificity | 0.73 (95% CI: 0.62 to 0.82) | Specificity | -- (95% CI: -- to --) |

| Prevalences | 5% | 10% | 20% |
| --- | --- | --- | --- |

| Outcome | № of studies (№ of patients) | Study design | Factors that may decrease certainty of evidence | | | | | Effect per 1,000 patients tested | | | | | | Test accuracy CoE |
| --- | --- | --- | --- | --- | --- | --- | --- | --- | --- | --- | --- | --- | --- | --- |
|  |  |  |  |  |  |  |  | pre-test probability of 5% | | pre-test probability of 10% | | pre-test probability of 20% | |  |
|  |  |  | Risk of bias | Indirectness | Inconsistency | Imprecision | Publication bias | CCA1 cassette | RT-PCR | CCA1 cassette | RT-PCR | CCA1 cassette | RT-PCR |  |
| **True positives** (patients with Schistosomiasis) | 1 studies 108 patients | cross-sectional (cohort type accuracy study) | not serious | not serious | not serious | serious ^a^ | none | 33 (28 to 37) | 0 (0 to 0) | 65 (55 to 74) | 0 (0 to 0) | 130 (110 to 148) | 0 (0 to 0) | ⨁⨁⨁◯ MODERATE |
|  |  |  |  |  |  |  |  | **33 more TP in CCA1 cassette** | | **65 more TP in CCA1 cassette** | | **130 more TP in CCA1 cassette** | |  |
| **False negatives** (patients incorrectly classified as not having Schistosomiasis) |  |  |  |  |  |  |  | 17 (13 to 22) | 50 (50 to 50) | 35 (26 to 45) | 100 (100 to 100) | 70 (52 to 90) | 200 (200 to 200) |  |
|  |  |  |  |  |  |  |  | **33 fewer FN in CCA1 cassette** | | **65 fewer FN in CCA1 cassette** | | **130 fewer FN in CCA1 cassette** | |  |
| **True negatives** (patients without Schistosomiasis) | 1 studies 88 patients | cross-sectional (cohort type accuracy study) | not serious | not serious | not serious | serious ^a^ | none | 694 (589 to 779) | 0 (0 to 0) | 657 (558 to 738) | 0 (0 to 0) | 584 (496 to 656) | 0 (0 to 0) | ⨁⨁⨁◯ MODERATE |
|  |  |  |  |  |  |  |  | **694 more TN in CCA1 cassette** | | **657 more TN in CCA1 cassette** | | **584 more TN in CCA1 cassette** | |  |
| **False positives** (patients incorrectly classified as having Schistosomiasis) |  |  |  |  |  |  |  | 256 (171 to 361) | 950 (950 to 950) | 243 (162 to 342) | 900 (900 to 900) | 216 (144 to 304) | 800 (800 to 800) |  |
|  |  |  |  |  |  |  |  | **694 fewer FP in CCA1 cassette** | | **657 fewer FP in CCA1 cassette** | | **584 fewer FP in CCA1 cassette** | |  |

##### Explanations

a. imperfect reference test

### Question: Should CCA1 cassette vs. single KK be used to diagnose Schistosomiasis in low endemic regions?

| CCA1 cassette | | single KK | |
| --- | --- | --- | --- |
| Sensitivity | 0.83 (95% CI: 0.52 to 0.98) | Sensitivity | -- (95% CI: -- to --) |
| Specificity | 0.66 (95% CI: 0.59 to 0.73) | Specificity | -- (95% CI: -- to --) |

| Prevalences | 5% | 10% | 20% |
| --- | --- | --- | --- |

| Outcome | № of studies (№ of patients) | Study design | Factors that may decrease certainty of evidence | | | | | Effect per 1,000 patients tested | | | | | | Test accuracy CoE |
| --- | --- | --- | --- | --- | --- | --- | --- | --- | --- | --- | --- | --- | --- | --- |
|  |  |  |  |  |  |  |  | pre-test probability of 5% | | pre-test probability of 10% | | pre-test probability of 20% | |  |
|  |  |  | Risk of bias | Indirectness | Inconsistency | Imprecision | Publication bias | CCA1 cassette | single KK | CCA1 cassette | single KK | CCA1 cassette | single KK |  |
| **True positives** (patients with Schistosomiasis) | 1 studies 12 patients | cross-sectional (cohort type accuracy study) | not serious | not serious | not serious | serious ^a^ | none | 42 (26 to 49) | 0 (0 to 0) | 83 (52 to 98) | 0 (0 to 0) | 166 (104 to 196) | 0 (0 to 0) | ⨁⨁⨁◯ MODERATE |
|  |  |  |  |  |  |  |  | **42 more TP in CCA1 cassette** | | **83 more TP in CCA1 cassette** | | **166 more TP in CCA1 cassette** | |  |
| **False negatives** (patients incorrectly classified as not having Schistosomiasis) |  |  |  |  |  |  |  | 8 (1 to 24) | 50 (50 to 50) | 17 (2 to 48) | 100 (100 to 100) | 34 (4 to 96) | 200 (200 to 200) |  |
|  |  |  |  |  |  |  |  | **42 fewer FN in CCA1 cassette** | | **83 fewer FN in CCA1 cassette** | | **166 fewer FN in CCA1 cassette** | |  |
| **True negatives** (patients without Schistosomiasis) | 1 studies 205 patients | cross-sectional (cohort type accuracy study) | not serious | not serious | not serious | serious ^a^ | none | 627 (561 to 694) | 0 (0 to 0) | 594 (531 to 657) | 0 (0 to 0) | 528 (472 to 584) | 0 (0 to 0) | ⨁⨁⨁◯ MODERATE |
|  |  |  |  |  |  |  |  | **627 more TN in CCA1 cassette** | | **594 more TN in CCA1 cassette** | | **528 more TN in CCA1 cassette** | |  |
| **False positives** (patients incorrectly classified as having Schistosomiasis) |  |  |  |  |  |  |  | 323 (256 to 389) | 950 (950 to 950) | 306 (243 to 369) | 900 (900 to 900) | 272 (216 to 328) | 800 (800 to 800) |  |
|  |  |  |  |  |  |  |  | **627 fewer FP in CCA1 cassette** | | **594 fewer FP in CCA1 cassette** | | **528 fewer FP in CCA1 cassette** | |  |

##### Explanations

a. imperfect reference test

### Question: Should CCA2 cassette vs. lumped KK be used to diagnose Schistosomiasis in low endemic regions?

| CCA2 cassette | | lumped KK | |
| --- | --- | --- | --- |
| Sensitivity | 0.45 to 0.63 | Sensitivity | -- to -- |
| Specificity | 0.96 to 0.96 | Specificity | -- to -- |

| Prevalences | 5% | 10% | 20% |
| --- | --- | --- | --- |

| Outcome | № of studies (№ of patients) | Study design | Factors that may decrease certainty of evidence | | | | | Effect per 1,000 patients tested | | | | | | Test accuracy CoE |
| --- | --- | --- | --- | --- | --- | --- | --- | --- | --- | --- | --- | --- | --- | --- |
|  |  |  |  |  |  |  |  | pre-test probability of 5% | | pre-test probability of 10% | | pre-test probability of 20% | |  |
|  |  |  | Risk of bias | Indirectness | Inconsistency | Imprecision | Publication bias | CCA2 cassette | lumped KK | CCA2 cassette | lumped KK | CCA2 cassette | lumped KK |  |
| **True positives** (patients with Schistosomiasis) | 2 studies 19 patients | cross-sectional (cohort type accuracy study) | not serious | not serious | not serious | serious ^a^ | none | 23 to 32 | 0 to 0 | 45 to 63 | 0 to 0 | 90 to 126 | 0 to 0 | ⨁⨁⨁◯ MODERATE |
|  |  |  |  |  |  |  |  | **23 more to 32 more TP in CCA2 cassette** | | **45 more to 63 more TP in CCA2 cassette** | | **90 more to 126 more TP in CCA2 cassette** | |  |
| **False negatives** (patients incorrectly classified as not having Schistosomiasis) |  |  |  |  |  |  |  | 18 to 27 | 50 to 50 | 37 to 55 | 100 to 100 | 74 to 110 | 200 to 200 |  |
|  |  |  |  |  |  |  |  | **23 fewer to 32 fewer FN in CCA2 cassette** | | **45 fewer to 63 fewer FN in CCA2 cassette** | | **90 fewer to 126 fewer FN in CCA2 cassette** | |  |
| **True negatives** (patients without Schistosomiasis) | 2 studies 181 patients | cross-sectional (cohort type accuracy study) | not serious | not serious | not serious | serious ^a^ | none | 912 to 912 | 0 to 0 | 864 to 864 | 0 to 0 | 768 to 768 | 0 to 0 | ⨁⨁⨁◯ MODERATE |
|  |  |  |  |  |  |  |  | **912 more to 912 more TN in CCA2 cassette** | | **864 more to 864 more TN in CCA2 cassette** | | **768 more to 768 more TN in CCA2 cassette** | |  |
| **False positives** (patients incorrectly classified as having Schistosomiasis) |  |  |  |  |  |  |  | 38 to 38 | 950 to 950 | 36 to 36 | 900 to 900 | 32 to 32 | 800 to 800 |  |
|  |  |  |  |  |  |  |  | **912 fewer to 912 fewer FP in CCA2 cassette** | | **864 fewer to 864 fewer FP in CCA2 cassette** | | **768 fewer to 768 fewer FP in CCA2 cassette** | |  |

##### Explanations

a. imperfect reference test

### Question: Should FLOTAC (all) vs. KK be used to diagnose Schistosomiasis in low endemic regions?

| FLOTAC (all) | | KK | |
| --- | --- | --- | --- |
| Sensitivity | 0.71 to 0.99 | Sensitivity | -- to -- |
| Specificity | 0.65 to 0.78 | Specificity | -- to -- |

| Prevalences | 5% | 10% | 20% |
| --- | --- | --- | --- |

| Outcome | № of studies (№ of patients) | Study design | Factors that may decrease certainty of evidence | | | | | Effect per 1,000 patients tested | | | | | | Test accuracy CoE |
| --- | --- | --- | --- | --- | --- | --- | --- | --- | --- | --- | --- | --- | --- | --- |
|  |  |  |  |  |  |  |  | pre-test probability of 5% | | pre-test probability of 10% | | pre-test probability of 20% | |  |
|  |  |  | Risk of bias | Indirectness | Inconsistency | Imprecision | Publication bias | FLOTAC (all) | KK | FLOTAC (all) | KK | FLOTAC (all) | KK |  |
| **True positives** (patients with Schistosomiasis) | 3 studies 216 patients | cross-sectional (cohort type accuracy study) | not serious | not serious | not serious | serious ^a^ | none | 36 to 50 | 0 to 0 | 71 to 99 | 0 to 0 | 142 to 198 | 0 to 0 | ⨁⨁⨁◯ MODERATE |
|  |  |  |  |  |  |  |  | **36 more to 50 more TP in FLOTAC (all)** | | **71 more to 99 more TP in FLOTAC (all)** | | **142 more to 198 more TP in FLOTAC (all)** | |  |
| **False negatives** (patients incorrectly classified as not having Schistosomiasis) |  |  |  |  |  |  |  | 0 to 14 | 50 to 50 | 1 to 29 | 100 to 100 | 2 to 58 | 200 to 200 |  |
|  |  |  |  |  |  |  |  | **36 fewer to 50 fewer FN in FLOTAC (all)** | | **71 fewer to 99 fewer FN in FLOTAC (all)** | | **142 fewer to 198 fewer FN in FLOTAC (all)** | |  |
| **True negatives** (patients without Schistosomiasis) | 3 studies 120 patients | cross-sectional (cohort type accuracy study) | not serious | not serious | not serious | serious ^a^ | none | 617 to 741 | 0 to 0 | 585 to 702 | 0 to 0 | 520 to 624 | 0 to 0 | ⨁⨁⨁◯ MODERATE |
|  |  |  |  |  |  |  |  | **617 more to 741 more TN in FLOTAC (all)** | | **585 more to 702 more TN in FLOTAC (all)** | | **520 more to 624 more TN in FLOTAC (all)** | |  |
| **False positives** (patients incorrectly classified as having Schistosomiasis) |  |  |  |  |  |  |  | 209 to 333 | 950 to 950 | 198 to 315 | 900 to 900 | 176 to 280 | 800 to 800 |  |
|  |  |  |  |  |  |  |  | **617 fewer to 741 fewer FP in FLOTAC (all)** | | **585 fewer to 702 fewer FP in FLOTAC (all)** | | **520 fewer to 624 fewer FP in FLOTAC (all)** | |  |

##### Explanations

a. imperfect reference test

### Question: Should Helmintex vs. RT-PCR be used to diagnose Schistosomiasis in low endemic regions?

| Helmintex | | RT-PCR | |
| --- | --- | --- | --- |
| Sensitivity | 0.68 (95% CI: 0.58 to 0.77) | Sensitivity | -- (95% CI: -- to --) |
| Specificity | 0.92 (95% CI: 0.83 to 0.97) | Specificity | -- (95% CI: -- to --) |

| Prevalences | 5% | 10% | 20% |
| --- | --- | --- | --- |

| Outcome | № of studies (№ of patients) | Study design | Factors that may decrease certainty of evidence | | | | | Effect per 1,000 patients tested | | | | | | Test accuracy CoE |
| --- | --- | --- | --- | --- | --- | --- | --- | --- | --- | --- | --- | --- | --- | --- |
|  |  |  |  |  |  |  |  | pre-test probability of 5% | | pre-test probability of 10% | | pre-test probability of 20% | |  |
|  |  |  | Risk of bias | Indirectness | Inconsistency | Imprecision | Publication bias | Helmintex | RT-PCR | Helmintex | RT-PCR | Helmintex | RT-PCR |  |
| **True positives** (patients with Schistosomiasis) | 1 studies 104 patients | cross-sectional (cohort type accuracy study) | not serious | not serious | not serious | serious ^a^ | none | 34 (29 to 39) | 0 (0 to 0) | 68 (58 to 77) | 0 (0 to 0) | 136 (116 to 154) | 0 (0 to 0) | ⨁⨁⨁◯ MODERATE |
|  |  |  |  |  |  |  |  | **34 more TP in Helmintex** | | **68 more TP in Helmintex** | | **136 more TP in Helmintex** | |  |
| **False negatives** (patients incorrectly classified as not having Schistosomiasis) |  |  |  |  |  |  |  | 16 (11 to 21) | 50 (50 to 50) | 32 (23 to 42) | 100 (100 to 100) | 64 (46 to 84) | 200 (200 to 200) |  |
|  |  |  |  |  |  |  |  | **34 fewer FN in Helmintex** | | **68 fewer FN in Helmintex** | | **136 fewer FN in Helmintex** | |  |
| **True negatives** (patients without Schistosomiasis) | 1 studies 72 patients | cross-sectional (cohort type accuracy study) | not serious | not serious | not serious | serious ^a^ | none | 874 (789 to 922) | 0 (0 to 0) | 828 (747 to 873) | 0 (0 to 0) | 736 (664 to 776) | 0 (0 to 0) | ⨁⨁⨁◯ MODERATE |
|  |  |  |  |  |  |  |  | **874 more TN in Helmintex** | | **828 more TN in Helmintex** | | **736 more TN in Helmintex** | |  |
| **False positives** (patients incorrectly classified as having Schistosomiasis) |  |  |  |  |  |  |  | 76 (28 to 161) | 950 (950 to 950) | 72 (27 to 153) | 900 (900 to 900) | 64 (24 to 136) | 800 (800 to 800) |  |
|  |  |  |  |  |  |  |  | **874 fewer FP in Helmintex** | | **828 fewer FP in Helmintex** | | **736 fewer FP in Helmintex** | |  |

##### Explanations

a. imperfect reference test

### Question: Should RT-PCR be used to diagnose Schistosomiasis in low endemic regions?

| Sensitivity | 1.00 (95% CI: 0.91 to 1.00) |
| --- | --- |
| Specificity | 0.56 (95% CI: 0.48 to 0.63) |

| Prevalences | 5% | 10% | 20% |
| --- | --- | --- | --- |

| Outcome | № of studies (№ of patients) | Study design | Factors that may decrease certainty of evidence | | | | | Effect per 1,000 patients tested | | | Test accuracy CoE |
| --- | --- | --- | --- | --- | --- | --- | --- | --- | --- | --- | --- |
|  |  |  | Risk of bias | Indirectness | Inconsistency | Imprecision | Publication bias | pre-test probability of 5% | pre-test probability of 10% | pre-test probability of 20% |  |
| **True positives** (patients with Schistosomiasis) | 1 studies 41 patients | cross-sectional (cohort type accuracy study) | not serious | not serious | not serious | serious ^a^ | none | 50 (46 to 50) | 100 (91 to 100) | 200 (182 to 200) | ⨁⨁⨁◯ MODERATE |
| **False negatives** (patients incorrectly classified as not having Schistosomiasis) |  |  |  |  |  |  |  | 0 (0 to 4) | 0 (0 to 9) | 0 (0 to 18) |  |
| **True negatives** (patients without Schistosomiasis) | 1 studies 165 patients | cross-sectional (cohort type accuracy study) | not serious | not serious | not serious | serious ^a^ | none | 532 (456 to 598) | 504 (432 to 567) | 448 (384 to 504) | ⨁⨁⨁◯ MODERATE |
| **False positives** (patients incorrectly classified as having Schistosomiasis) |  |  |  |  |  |  |  | 418 (352 to 494) | 396 (333 to 468) | 352 (296 to 416) |  |

##### Explanations

a. imperfect reference test

### Question: Should RT-PCR vs. sextuple KK be used to diagnose Schistosomiasis in low endemic regions?

| RT-PCR | | sextuple KK | |
| --- | --- | --- | --- |
| Sensitivity | 0.95 (95% CI: 0.87 to 0.99) | Sensitivity | -- (95% CI: -- to --) |
| Specificity | 0.69 (95% CI: 0.60 to 0.76) | Specificity | -- (95% CI: -- to --) |

| Prevalences | 5% | 10% | 20% |
| --- | --- | --- | --- |

| Outcome | № of studies (№ of patients) | Study design | Factors that may decrease certainty of evidence | | | | | Effect per 1,000 patients tested | | | | | | Test accuracy CoE |
| --- | --- | --- | --- | --- | --- | --- | --- | --- | --- | --- | --- | --- | --- | --- |
|  |  |  |  |  |  |  |  | pre-test probability of 5% | | pre-test probability of 10% | | pre-test probability of 20% | |  |
|  |  |  | Risk of bias | Indirectness | Inconsistency | Imprecision | Publication bias | RT-PCR | sextuple KK | RT-PCR | sextuple KK | RT-PCR | sextuple KK |  |
| **True positives** (patients with Schistosomiasis) | 1 studies 64 patients | cross-sectional (cohort type accuracy study) | not serious | not serious | not serious | serious ^a^ | none | 48 (44 to 50) | 0 (0 to 0) | 95 (87 to 99) | 0 (0 to 0) | 190 (174 to 198) | 0 (0 to 0) | ⨁⨁⨁◯ MODERATE |
|  |  |  |  |  |  |  |  | **48 more TP in RT-PCR** | | **95 more TP in RT-PCR** | | **190 more TP in RT-PCR** | |  |
| **False negatives** (patients incorrectly classified as not having Schistosomiasis) |  |  |  |  |  |  |  | 2 (0 to 6) | 50 (50 to 50) | 5 (1 to 13) | 100 (100 to 100) | 10 (2 to 26) | 200 (200 to 200) |  |
|  |  |  |  |  |  |  |  | **48 fewer FN in RT-PCR** | | **95 fewer FN in RT-PCR** | | **190 fewer FN in RT-PCR** | |  |
| **True negatives** (patients without Schistosomiasis) | 1 studies 134 patients | cross-sectional (cohort type accuracy study) | not serious | not serious | not serious | serious ^a^ | none | 656 (570 to 722) | 0 (0 to 0) | 621 (540 to 684) | 0 (0 to 0) | 552 (480 to 608) | 0 (0 to 0) | ⨁⨁⨁◯ MODERATE |
|  |  |  |  |  |  |  |  | **656 more TN in RT-PCR** | | **621 more TN in RT-PCR** | | **552 more TN in RT-PCR** | |  |
| **False positives** (patients incorrectly classified as having Schistosomiasis) |  |  |  |  |  |  |  | 294 (228 to 380) | 950 (950 to 950) | 279 (216 to 360) | 900 (900 to 900) | 248 (192 to 320) | 800 (800 to 800) |  |
|  |  |  |  |  |  |  |  | **656 fewer FP in RT-PCR** | | **621 fewer FP in RT-PCR** | | **552 fewer FP in RT-PCR** | |  |

##### Explanations

a. imperfect reference test
