## Supplementary material for "Diagnostic tests for Schistosomiasis for low prevalence settings: a systematic review and Meta-Analysis": Heterogeneity & publication bias

### CCA1 cassette vs duplicate KK

#### Heterogeneity

```
## ================================================
##
## ======== Heterogeneity Analysis for DOR ========
##
## ================================================
##
## Random-Effects Model (k = 17; tau^2 estimator: REML)
##
## tau^2 (estimated amount of total heterogeneity): 1.4713 (SE = 0.6222)
## tau (square root of estimated tau^2 value):      1.2130
## I^2 (total heterogeneity / total variability):   88.66%
## H^2 (total variability / sampling variability):  8.82
##
## Test for Heterogeneity:
## Q(df = 16) = 112.6012, p-val < .0001
##
## Model Results:
##
## estimate      se    zval    pval   ci.lb   ci.ub
##   2.1019  0.3231  6.5055  <.0001  1.4687  2.7352  ***
##
## ---
## Signif. codes:  0 '***' 0.001 '**' 0.01 '*' 0.05 '.' 0.1 ' ' 1
##
## ========================================================================
##
## ======== Heterogeneity Analysis for Sensitivity and Specificity ========
##
## ========================================================================
## Heterogeneity test for sensitivity
## $statistic
## X-squared
##  292.4661
##
## $p.value
## [1] 9.243177e-53
##
## [1] "I^2:  94.53 %"
##
##
## Heterogeneity test for specificity
## $statistic
## X-squared
##    530.94
##
## $p.value
## [1] 9.662395e-103
##
## [1] "I^2:  96.99 %"
```

#### Publication Bias

```

## ==========================================================
##
## ========= Funnel Plot, Egger's regression test &  ========
##
## ========= Begg's rank test for lnDOR vs SE(lnDOR) ========
##
## ==========================================================
```

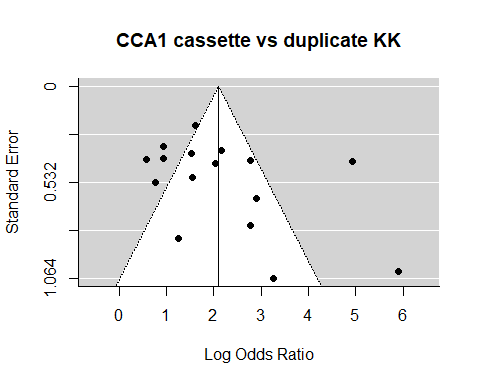

```
##
## Regression Test for Funnel Plot Asymmetry
##
## model:     mixed-effects meta-regression model
## predictor: standard error
##
## test for funnel plot asymmetry: z = 1.9655, p = 0.0494
##
##
## Rank Correlation Test for Funnel Plot Asymmetry
##
## Kendall's tau = 0.3037, p = 0.0907


## ==========================================================
##
## ========= Deek's Funnel Plot &                    ========
##
## ========= Asymmetry test for lnDOR vs 1/sqrt(ESS) ========
##
## ==========================================================
```

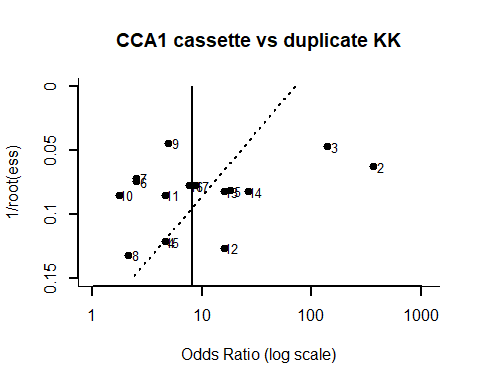

```
##
## Regression Test for Funnel Plot Asymmetry
##
## model:     weighted regression with multiplicative dispersion
## predictor: standard error
##
## test for funnel plot asymmetry: t = -1.4358, df = 15, p = 0.1716
```

### CCA1 cassette vs quadruple KK

#### Heterogeneity

```

## ================================================
##
## ======== Heterogeneity Analysis for DOR ========
##
## ================================================
##
## Random-Effects Model (k = 11; tau^2 estimator: REML)
##
## tau^2 (estimated amount of total heterogeneity): 0.6312 (SE = 0.4067)
## tau (square root of estimated tau^2 value):      0.7945
## I^2 (total heterogeneity / total variability):   81.56%
## H^2 (total variability / sampling variability):  5.42
##
## Test for Heterogeneity:
## Q(df = 10) = 54.7345, p-val < .0001
##
## Model Results:
##
## estimate      se    zval    pval   ci.lb   ci.ub
##   2.2162  0.2925  7.5768  <.0001  1.6429  2.7894  ***
##
## ---
## Signif. codes:  0 '***' 0.001 '**' 0.01 '*' 0.05 '.' 0.1 ' ' 1
##
## ========================================================================
##
## ======== Heterogeneity Analysis for Sensitivity and Specificity ========
##
## ========================================================================
## Heterogeneity test for sensitivity
## $statistic
## X-squared
##  59.29295
##
## $p.value
## [1] 4.930608e-09
##
## [1] "I^2:  83.13 %"
##
##
## Heterogeneity test for specificity
## $statistic
## X-squared
##  250.3836
##
## $p.value
## [1] 4.507931e-48
##
## [1] "I^2:  96.01 %"
```

#### Publication Bias

```

## ==========================================================
##
## ========= Funnel Plot, Egger's regression test &  ========
##
## ========= Begg's rank test for lnDOR vs SE(lnDOR) ========
##
## ==========================================================
```

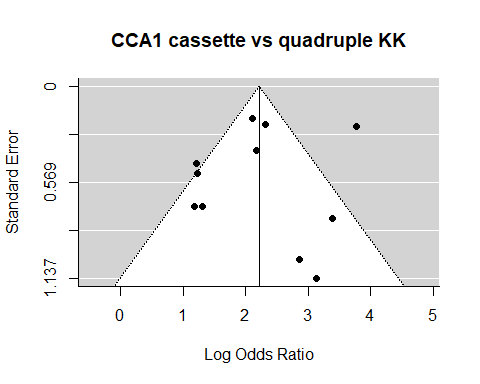

```
##
## Regression Test for Funnel Plot Asymmetry
##
## model:     mixed-effects meta-regression model
## predictor: standard error
##
## test for funnel plot asymmetry: z = -0.0991, p = 0.9211
##
##
## Rank Correlation Test for Funnel Plot Asymmetry
##
## Kendall's tau = 0.2364, p = 0.3587


##
## ==========================================================
##
## ========= Deek's Funnel Plot &                    ========
##
## ========= Asymmetry test for lnDOR vs 1/sqrt(ESS) ========
##
## ==========================================================
```

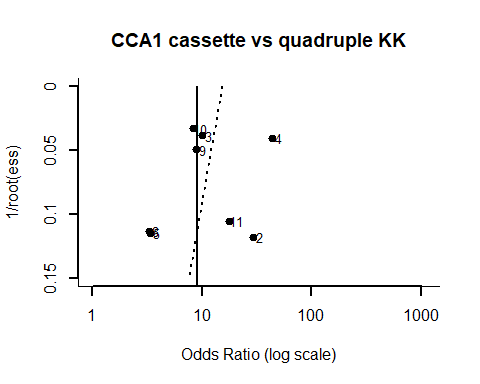

```
##
## Regression Test for Funnel Plot Asymmetry
##
## model:     weighted regression with multiplicative dispersion
## predictor: standard error
##
## test for funnel plot asymmetry: t = -0.6192, df = 9, p = 0.5511
```

### CCA1 cassette vs sextuple KK

#### Heterogeneity

```

##
## ================================================
##
## ======== Heterogeneity Analysis for DOR ========
##
## ================================================
##
## Random-Effects Model (k = 7; tau^2 estimator: REML)
##
## tau^2 (estimated amount of total heterogeneity): 0.2403 (SE = 0.2548)
## tau (square root of estimated tau^2 value):      0.4902
## I^2 (total heterogeneity / total variability):   59.48%
## H^2 (total variability / sampling variability):  2.47
##
## Test for Heterogeneity:
## Q(df = 6) = 13.9651, p-val = 0.0300
##
## Model Results:
##
## estimate      se    zval    pval   ci.lb   ci.ub
##   2.3571  0.2570  9.1708  <.0001  1.8533  2.8608  ***
##
## ---
## Signif. codes:  0 '***' 0.001 '**' 0.01 '*' 0.05 '.' 0.1 ' ' 1
##
## ========================================================================
##
## ======== Heterogeneity Analysis for Sensitivity and Specificity ========
##
## ========================================================================
## Heterogeneity test for sensitivity
## $statistic
## X-squared
##  34.56929
##
## $p.value
## [1] 5.221737e-06
##
## [1] "I^2:  82.64 %"
##
##
## Heterogeneity test for specificity
## $statistic
## X-squared
##   35.3468
##
## $p.value
## [1] 3.691477e-06
##
## [1] "I^2:  83.03 %"
```

#### Publication Bias

```

## ==========================================================
##
## ========= Funnel Plot, Egger's regression test &  ========
##
## ========= Begg's rank test for lnDOR vs SE(lnDOR) ========
##
## ==========================================================
```

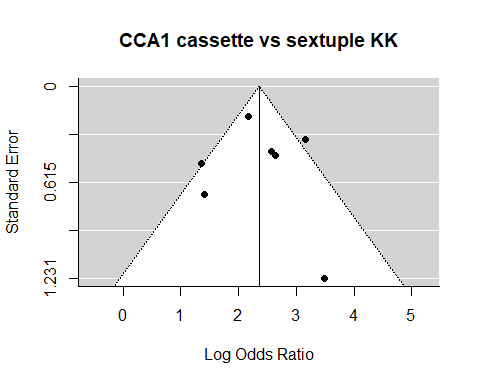

```
##
## Regression Test for Funnel Plot Asymmetry
##
## model:     mixed-effects meta-regression model
## predictor: standard error
##
## test for funnel plot asymmetry: z = 0.0462, p = 0.9632
##
##
## Rank Correlation Test for Funnel Plot Asymmetry
##
## Kendall's tau = -0.0476, p = 1.0000
##


## ==========================================================
##
## ========= Deek's Funnel Plot &                    ========
##
## ========= Asymmetry test for lnDOR vs 1/sqrt(ESS) ========
##
## ==========================================================
```

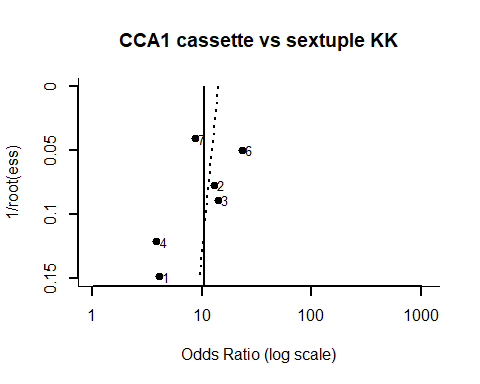

```
##
## Regression Test for Funnel Plot Asymmetry
##
## model:     weighted regression with multiplicative dispersion
## predictor: standard error
##
## test for funnel plot asymmetry: t = -0.3792, df = 5, p = 0.7201
```

### SmCTF-RDT vs quadruple KK

#### Heterogeneity

```

##
## ================================================
##
## ======== Heterogeneity Analysis for DOR ========
##
## ================================================
##
## Random-Effects Model (k = 4; tau^2 estimator: REML)
##
## tau^2 (estimated amount of total heterogeneity): 1.5058 (SE = 1.8545)
## tau (square root of estimated tau^2 value):      1.2271
## I^2 (total heterogeneity / total variability):   70.49%
## H^2 (total variability / sampling variability):  3.39
##
## Test for Heterogeneity:
## Q(df = 3) = 7.9454, p-val = 0.0472
##
## Model Results:
##
## estimate      se    zval    pval    ci.lb   ci.ub
##   0.7511  0.7546  0.9954  0.3195  -0.7278  2.2301
##
## ---
## Signif. codes:  0 '***' 0.001 '**' 0.01 '*' 0.05 '.' 0.1 ' ' 1
##
## ========================================================================
##
## ======== Heterogeneity Analysis for Sensitivity and Specificity ========
##
## ========================================================================
## Heterogeneity test for sensitivity
## $statistic
## X-squared
##  22.32815
##
## $p.value
## [1] 5.573872e-05
##
## [1] "I^2:  86.56 %"
##
##
## Heterogeneity test for specificity
## $statistic
## X-squared
##  4.082923
##
## $p.value
## [1] 0.2526481
##
## [1] "I^2:  26.52 %"
```

#### Publication Bias

```

## ==========================================================
##
## ========= Funnel Plot, Egger's regression test &  ========
##
## ========= Begg's rank test for lnDOR vs SE(lnDOR) ========
##
## ==========================================================
```

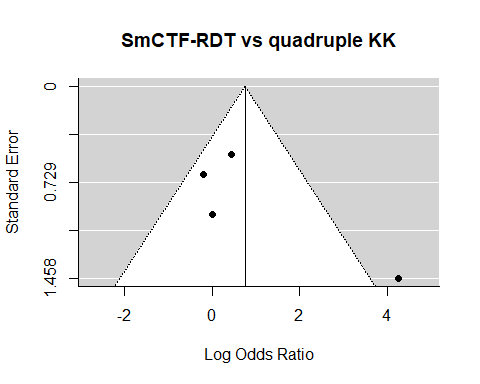

```
##
## Regression Test for Funnel Plot Asymmetry
##
## model:     mixed-effects meta-regression model
## predictor: standard error
##
## test for funnel plot asymmetry: z = 1.7807, p = 0.0750
##
##
## Rank Correlation Test for Funnel Plot Asymmetry
##
## Kendall's tau = 0.3333, p = 0.7500
##


## ==========================================================
##
## ========= Deek's Funnel Plot &                    ========
##
## ========= Asymmetry test for lnDOR vs 1/sqrt(ESS) ========
##
## ==========================================================
```

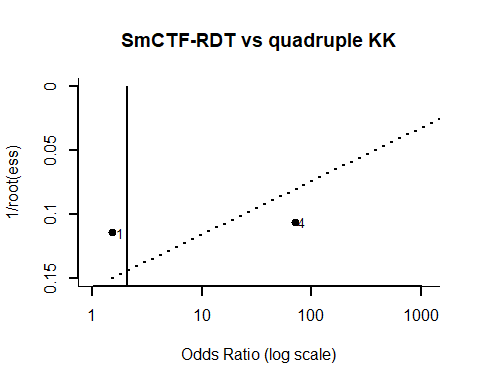

```
##
## Regression Test for Funnel Plot Asymmetry
##
## model:     weighted regression with multiplicative dispersion
## predictor: standard error
##
## test for funnel plot asymmetry: t = -1.3981, df = 2, p = 0.2970
```

### IgG ELISA vs triplicate KK

#### Heterogeneity

```

##
## ================================================
##
## ======== Heterogeneity Analysis for DOR ========
##
## ================================================
##
## Random-Effects Model (k = 4; tau^2 estimator: REML)
##
## tau^2 (estimated amount of total heterogeneity): 8.8562 (SE = 7.8737)
## tau (square root of estimated tau^2 value):      2.9759
## I^2 (total heterogeneity / total variability):   94.73%
## H^2 (total variability / sampling variability):  18.99
##
## Test for Heterogeneity:
## Q(df = 3) = 31.2223, p-val < .0001
##
## Model Results:
##
## estimate      se    zval    pval   ci.lb   ci.ub
##   3.8182  1.5527  2.4591  0.0139  0.7750  6.8614  *
##
## ---
## Signif. codes:  0 '***' 0.001 '**' 0.01 '*' 0.05 '.' 0.1 ' ' 1
##
## ========================================================================
##
## ======== Heterogeneity Analysis for Sensitivity and Specificity ========
##
## ========================================================================
## Heterogeneity test for sensitivity
## $statistic
## X-squared
##  17.29169
##
## $p.value
## [1] 0.0006155247
##
## [1] "I^2:  82.65 %"
##
##
## Heterogeneity test for specificity
## $statistic
## X-squared
##  149.8377
##
## $p.value
## [1] 2.856102e-32
##
## [1] "I^2:  98 %"
```

#### Publication Bias

```

## ==========================================================
##
## ========= Funnel Plot, Egger's regression test &  ========
##
## ========= Begg's rank test for lnDOR vs SE(lnDOR) ========
##
## ==========================================================
```

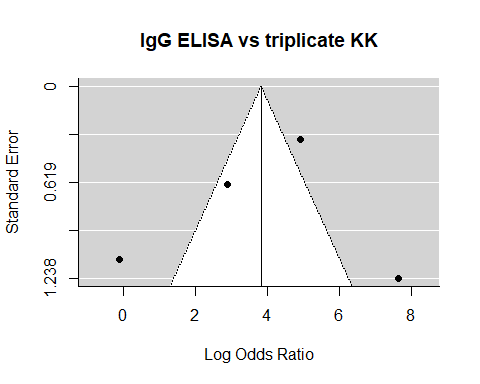

```
##
## Regression Test for Funnel Plot Asymmetry
##
## model:     mixed-effects meta-regression model
## predictor: standard error
##
## test for funnel plot asymmetry: z = 0.0109, p = 0.9913
##
##
## Rank Correlation Test for Funnel Plot Asymmetry
##
## Kendall's tau = -0.3333, p = 0.7500
##


## ==========================================================
##
## ========= Deek's Funnel Plot &                    ========
##
## ========= Asymmetry test for lnDOR vs 1/sqrt(ESS) ========
##
## ==========================================================
```

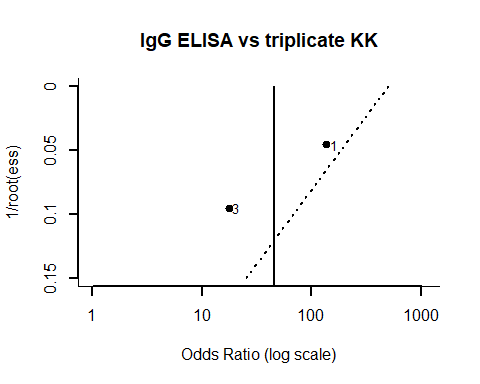

```
##
## Regression Test for Funnel Plot Asymmetry
##
## model:     weighted regression with multiplicative dispersion
## predictor: standard error
##
## test for funnel plot asymmetry: t = -0.5999, df = 2, p = 0.6095
```

### IgG SEA-ELISA vs Urine Microscopy

#### Heterogeneity

```

##
## ================================================
##
## ======== Heterogeneity Analysis for DOR ========
##
## ================================================
##
## Random-Effects Model (k = 4; tau^2 estimator: REML)
##
## tau^2 (estimated amount of total heterogeneity): 0.5902 (SE = 0.8330)
## tau (square root of estimated tau^2 value):      0.7682
## I^2 (total heterogeneity / total variability):   59.20%
## H^2 (total variability / sampling variability):  2.45
##
## Test for Heterogeneity:
## Q(df = 3) = 7.2751, p-val = 0.0636
##
## Model Results:
##
## estimate      se    zval    pval   ci.lb   ci.ub
##   3.2420  0.5058  6.4101  <.0001  2.2507  4.2333  ***
##
## ---
## Signif. codes:  0 '***' 0.001 '**' 0.01 '*' 0.05 '.' 0.1 ' ' 1
##
## ========================================================================
##
## ======== Heterogeneity Analysis for Sensitivity and Specificity ========
##
## ========================================================================
## Heterogeneity test for sensitivity
## $statistic
## X-squared
##  8.832465
##
## $p.value
## [1] 0.03160332
##
## [1] "I^2:  66.03 %"
##
##
## Heterogeneity test for specificity
## $statistic
## X-squared
##  93.89254
##
## $p.value
## [1] 3.19376e-20
##
## [1] "I^2:  96.8 %"
```

#### Publication Bias

```

## ==========================================================
##
## ========= Funnel Plot, Egger's regression test &  ========
##
## ========= Begg's rank test for lnDOR vs SE(lnDOR) ========
##
## ==========================================================
```

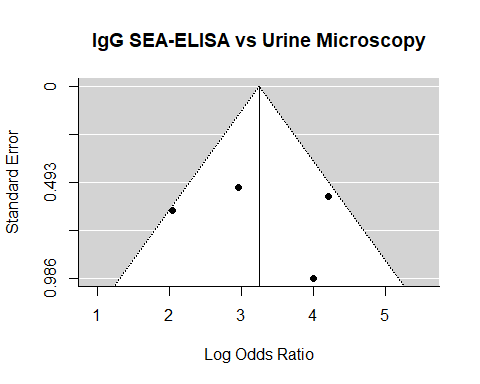

```
##
## Regression Test for Funnel Plot Asymmetry
##
## model:     mixed-effects meta-regression model
## predictor: standard error
##
## test for funnel plot asymmetry: z = 0.3883, p = 0.6978
##
##
## Rank Correlation Test for Funnel Plot Asymmetry
##
## Kendall's tau = 0.0000, p = 1.0000


##
## ==========================================================
##
## ========= Deek's Funnel Plot &                    ========
##
## ========= Asymmetry test for lnDOR vs 1/sqrt(ESS) ========
##
## ==========================================================
```

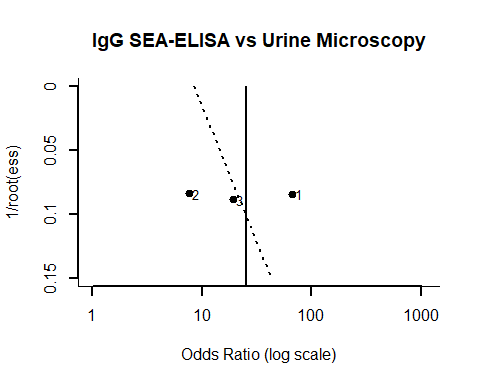

```
##
## Regression Test for Funnel Plot Asymmetry
##
## model:     weighted regression with multiplicative dispersion
## predictor: standard error
##
## test for funnel plot asymmetry: t = 0.4041, df = 2, p = 0.7252
```

### Haematuria (R strip) vs Urine Microscopy

#### Heterogeneity

```

##
## ================================================
##
## ======== Heterogeneity Analysis for DOR ========
##
## ================================================
##
## Random-Effects Model (k = 72; tau^2 estimator: REML)
##
## tau^2 (estimated amount of total heterogeneity): 1.6720 (SE = 0.3067)
## tau (square root of estimated tau^2 value):      1.2930
## I^2 (total heterogeneity / total variability):   98.19%
## H^2 (total variability / sampling variability):  55.24
##
## Test for Heterogeneity:
## Q(df = 71) = 3370.5327, p-val < .0001
##
## Model Results:
##
## estimate      se     zval    pval   ci.lb   ci.ub
##   2.9575  0.1600  18.4830  <.0001  2.6439  3.2712  ***
##
## ---
## Signif. codes:  0 '***' 0.001 '**' 0.01 '*' 0.05 '.' 0.1 ' ' 1
##
## ========================================================================
##
## ======== Heterogeneity Analysis for Sensitivity and Specificity ========
##
## ========================================================================
## Heterogeneity test for sensitivity
## $statistic
## X-squared
##  3057.142
##
## $p.value
## [1] 0
##
## [1] "I^2:  97.68 %"
##
##
## Heterogeneity test for specificity
## $statistic
## X-squared
##  11096.52
##
## $p.value
## [1] 0
##
## [1] "I^2:  99.36 %"
```

#### Publication Bias

```

## ==========================================================
##
## ========= Funnel Plot, Egger's regression test &  ========
##
## ========= Begg's rank test for lnDOR vs SE(lnDOR) ========
##
## ==========================================================
```

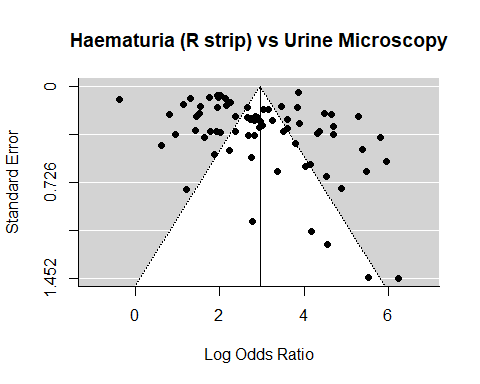

```
##
## Regression Test for Funnel Plot Asymmetry
##
## model:     mixed-effects meta-regression model
## predictor: standard error
##
## test for funnel plot asymmetry: z = 3.8162, p = 0.0001
##
##
## Rank Correlation Test for Funnel Plot Asymmetry
##
## Kendall's tau = 0.2019, p = 0.0119
##


## ==========================================================
##
## ========= Deek's Funnel Plot &                    ========
##
## ========= Asymmetry test for lnDOR vs 1/sqrt(ESS) ========
##
## ==========================================================
```

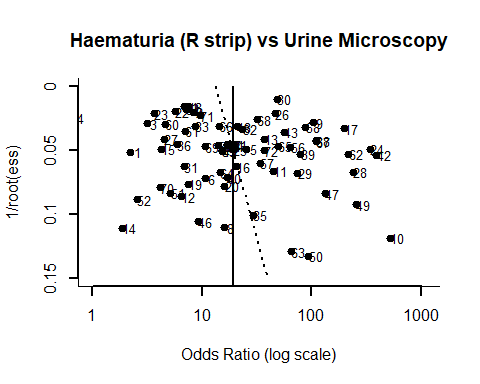

```
##
## Regression Test for Funnel Plot Asymmetry
##
## model:     weighted regression with multiplicative dispersion
## predictor: standard error
##
## test for funnel plot asymmetry: t = 0.9168, df = 70, p = 0.3624
```

### Proteinuria (R strip) vs Urine Microscopy

#### Heterogeneity

```

##
## ================================================
##
## ======== Heterogeneity Analysis for DOR ========
##
## ================================================
##
## Random-Effects Model (k = 41; tau^2 estimator: REML)
##
## tau^2 (estimated amount of total heterogeneity): 1.8180 (SE = 0.4324)
## tau (square root of estimated tau^2 value):      1.3483
## I^2 (total heterogeneity / total variability):   98.46%
## H^2 (total variability / sampling variability):  64.90
##
## Test for Heterogeneity:
## Q(df = 40) = 2150.2704, p-val < .0001
##
## Model Results:
##
## estimate      se    zval    pval   ci.lb   ci.ub
##   1.9526  0.2176  8.9733  <.0001  1.5261  2.3791  ***
##
## ---
## Signif. codes:  0 '***' 0.001 '**' 0.01 '*' 0.05 '.' 0.1 ' ' 1
##
## ========================================================================
##
## ======== Heterogeneity Analysis for Sensitivity and Specificity ========
##
## ========================================================================
## Heterogeneity test for sensitivity
## $statistic
## X-squared
##  3547.146
##
## $p.value
## [1] 0
##
## [1] "I^2:  98.87 %"
##
##
## Heterogeneity test for specificity
## $statistic
## X-squared
##  10593.17
##
## $p.value
## [1] 0
##
## [1] "I^2:  99.62 %"
```

#### Publication Bias

```

## ==========================================================
##
## ========= Funnel Plot, Egger's regression test &  ========
##
## ========= Begg's rank test for lnDOR vs SE(lnDOR) ========
##
## ==========================================================
```

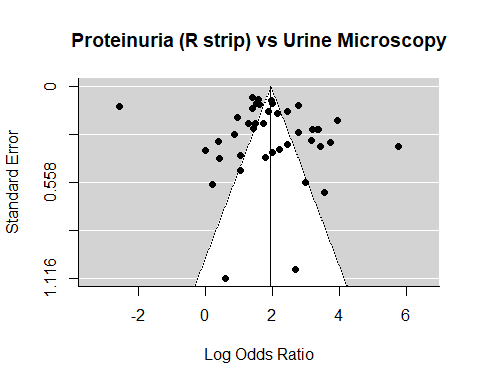

```
##
## Regression Test for Funnel Plot Asymmetry
##
## model:     mixed-effects meta-regression model
## predictor: standard error
##
## test for funnel plot asymmetry: z = 0.3599, p = 0.7189
##
##
## Rank Correlation Test for Funnel Plot Asymmetry
##
## Kendall's tau = -0.0195, p = 0.8671


##
## ==========================================================
##
## ========= Deek's Funnel Plot &                    ========
##
## ========= Asymmetry test for lnDOR vs 1/sqrt(ESS) ========
##
## ==========================================================
```

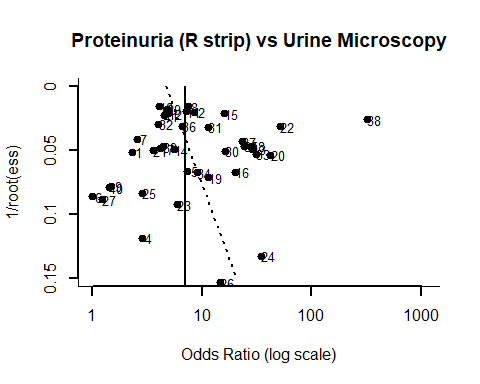

```
##
## Regression Test for Funnel Plot Asymmetry
##
## model:     weighted regression with multiplicative dispersion
## predictor: standard error
##
## test for funnel plot asymmetry: t = 0.7014, df = 39, p = 0.4872
```

### Leukocyturia vs Urine Microscopy

#### Heterogeneity

```

##
## ================================================
##
## ======== Heterogeneity Analysis for DOR ========
##
## ================================================
##
## Random-Effects Model (k = 5; tau^2 estimator: REML)
##
## tau^2 (estimated amount of total heterogeneity): 0.7410 (SE = 0.6375)
## tau (square root of estimated tau^2 value):      0.8608
## I^2 (total heterogeneity / total variability):   86.66%
## H^2 (total variability / sampling variability):  7.50
##
## Test for Heterogeneity:
## Q(df = 4) = 30.1487, p-val < .0001
##
## Model Results:
##
## estimate      se    zval    pval    ci.lb   ci.ub
##   0.7346  0.4257  1.7254  0.0845  -0.0999  1.5690  .
##
## ---
## Signif. codes:  0 '***' 0.001 '**' 0.01 '*' 0.05 '.' 0.1 ' ' 1
##
## ========================================================================
##
## ======== Heterogeneity Analysis for Sensitivity and Specificity ========
##
## ========================================================================
## Heterogeneity test for sensitivity
## $statistic
## X-squared
##   34.6945
##
## $p.value
## [1] 5.36731e-07
##
## [1] "I^2:  88.47 %"
##
##
## Heterogeneity test for specificity
## $statistic
## X-squared
##  276.7514
##
## $p.value
## [1] 1.117839e-58
##
## [1] "I^2:  98.55 %"
```

#### Publication Bias

```

## ==========================================================
##
## ========= Funnel Plot, Egger's regression test &  ========
##
## ========= Begg's rank test for lnDOR vs SE(lnDOR) ========
##
## ==========================================================
```

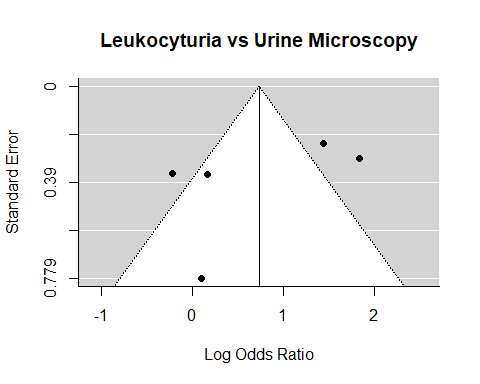

```
##
## Regression Test for Funnel Plot Asymmetry
##
## model:     mixed-effects meta-regression model
## predictor: standard error
##
## test for funnel plot asymmetry: z = -1.0346, p = 0.3009
##
##
## Rank Correlation Test for Funnel Plot Asymmetry
##
## Kendall's tau = -0.2000, p = 0.8167


##
## ==========================================================
##
## ========= Deek's Funnel Plot &                    ========
##
## ========= Asymmetry test for lnDOR vs 1/sqrt(ESS) ========
##
## ==========================================================
```

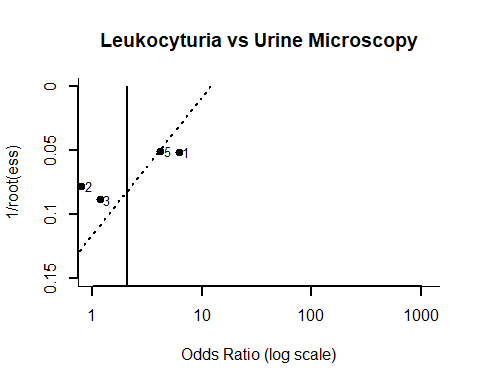

```
##
## Regression Test for Funnel Plot Asymmetry
##
## model:     weighted regression with multiplicative dispersion
## predictor: standard error
##
## test for funnel plot asymmetry: t = -1.6286, df = 3, p = 0.2019
```

### CCA1 vs Urine Microscopy

#### Heterogeneity

```

##
## ================================================
##
## ======== Heterogeneity Analysis for DOR ========
##
## ================================================
##
## Random-Effects Model (k = 4; tau^2 estimator: REML)
##
## tau^2 (estimated amount of total heterogeneity): 0.3628 (SE = 0.4281)
## tau (square root of estimated tau^2 value):      0.6023
## I^2 (total heterogeneity / total variability):   74.79%
## H^2 (total variability / sampling variability):  3.97
##
## Test for Heterogeneity:
## Q(df = 3) = 12.1292, p-val = 0.0070
##
## Model Results:
##
## estimate      se    zval    pval   ci.lb   ci.ub
##   1.2988  0.3644  3.5645  0.0004  0.5847  2.0130  ***
##
## ---
## Signif. codes:  0 '***' 0.001 '**' 0.01 '*' 0.05 '.' 0.1 ' ' 1
##
## ========================================================================
##
## ======== Heterogeneity Analysis for Sensitivity and Specificity ========
##
## ========================================================================
## Heterogeneity test for sensitivity
## $statistic
## X-squared
##  77.00153
##
## $p.value
## [1] 1.349131e-16
##
## [1] "I^2:  96.1 %"
##
##
## Heterogeneity test for specificity
## $statistic
## X-squared
##  84.74317
##
## $p.value
## [1] 2.946343e-18
##
## [1] "I^2:  96.46 %"
```

#### Publication Bias

```

## ==========================================================
##
## ========= Funnel Plot, Egger's regression test &  ========
##
## ========= Begg's rank test for lnDOR vs SE(lnDOR) ========
##
## ==========================================================
```

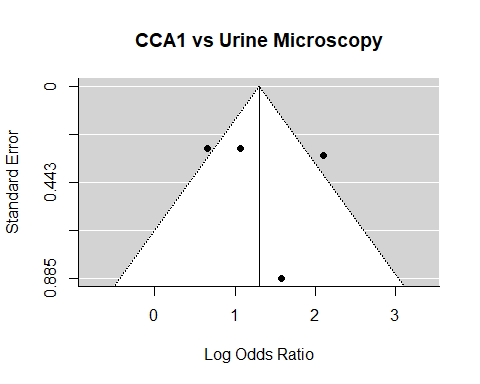

```
##
## Regression Test for Funnel Plot Asymmetry
##
## model:     mixed-effects meta-regression model
## predictor: standard error
##
## test for funnel plot asymmetry: z = 0.3730, p = 0.7092
##
##
## Rank Correlation Test for Funnel Plot Asymmetry
##
## Kendall's tau = 0.6667, p = 0.3333


##
## ==========================================================
##
## ========= Deek's Funnel Plot &                    ========
##
## ========= Asymmetry test for lnDOR vs 1/sqrt(ESS) ========
##
## ==========================================================
```

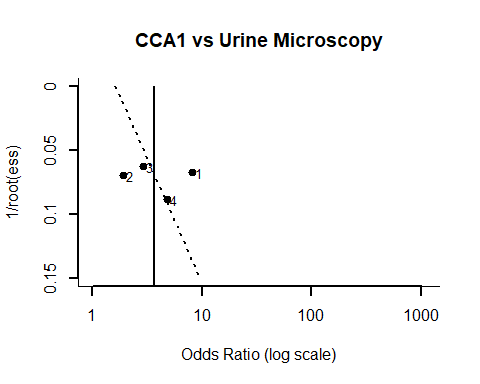

```
##
## Regression Test for Funnel Plot Asymmetry
##
## model:     weighted regression with multiplicative dispersion
## predictor: standard error
##
## test for funnel plot asymmetry: t = 0.2602, df = 2, p = 0.8191
```

### CAA vs Urine Microscopy

#### Heterogeneity

```

##
## ================================================
##
## ======== Heterogeneity Analysis for DOR ========
##
## ================================================
##
## Random-Effects Model (k = 4; tau^2 estimator: REML)
##
## tau^2 (estimated amount of total heterogeneity): 4.1391 (SE = 3.8268)
## tau (square root of estimated tau^2 value):      2.0345
## I^2 (total heterogeneity / total variability):   95.04%
## H^2 (total variability / sampling variability):  20.17
##
## Test for Heterogeneity:
## Q(df = 3) = 15.4591, p-val = 0.0015
##
## Model Results:
##
## estimate      se    zval    pval   ci.lb   ci.ub
##   2.6069  1.0830  2.4071  0.0161  0.4843  4.7296  *
##
## ---
## Signif. codes:  0 '***' 0.001 '**' 0.01 '*' 0.05 '.' 0.1 ' ' 1
##
## ========================================================================
##
## ======== Heterogeneity Analysis for Sensitivity and Specificity ========
##
## ========================================================================
## Heterogeneity test for sensitivity
## $statistic
## X-squared
##  146.1362
##
## $p.value
## [1] 1.795497e-31
##
## [1] "I^2:  97.95 %"
##
##
## Heterogeneity test for specificity
## $statistic
## X-squared
##  250.4547
##
## $p.value
## [1] 5.217599e-54
##
## [1] "I^2:  98.8 %"
```

#### Publication Bias

```

## ==========================================================
##
## ========= Funnel Plot, Egger's regression test &  ========
##
## ========= Begg's rank test for lnDOR vs SE(lnDOR) ========
##
## ==========================================================
```

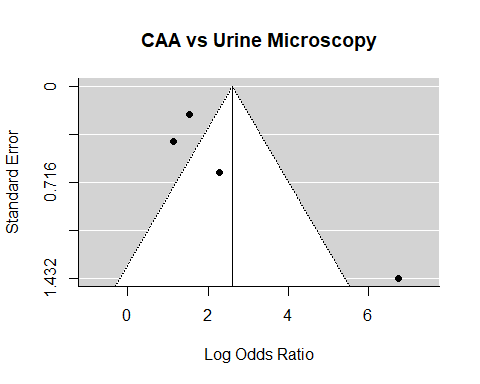

```
##
## Regression Test for Funnel Plot Asymmetry
##
## model:     mixed-effects meta-regression model
## predictor: standard error
##
## test for funnel plot asymmetry: z = 3.0047, p = 0.0027
##
##
## Rank Correlation Test for Funnel Plot Asymmetry
##
## Kendall's tau = 0.6667, p = 0.3333


##
## ==========================================================
##
## ========= Deek's Funnel Plot &                    ========
##
## ========= Asymmetry test for lnDOR vs 1/sqrt(ESS) ========
##
## ==========================================================
```

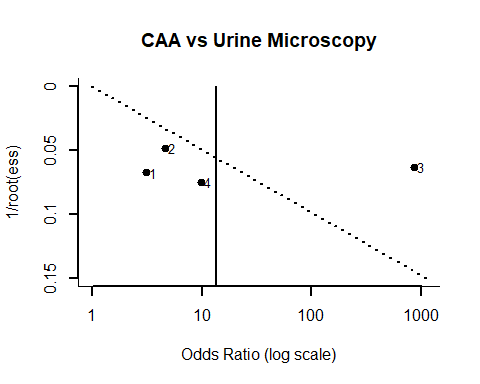

```
##
## Regression Test for Funnel Plot Asymmetry
##
## model:     weighted regression with multiplicative dispersion
## predictor: standard error
##
## test for funnel plot asymmetry: t = 0.3160, df = 2, p = 0.7819
```

### CCA1 vs KK

#### Heterogeneity

```

##
## ================================================
##
## ======== Heterogeneity Analysis for DOR ========
##
## ================================================
##
## Random-Effects Model (k = 37; tau^2 estimator: REML)
##
## tau^2 (estimated amount of total heterogeneity): 1.2506 (SE = 0.3646)
## tau (square root of estimated tau^2 value):      1.1183
## I^2 (total heterogeneity / total variability):   88.77%
## H^2 (total variability / sampling variability):  8.91
##
## Test for Heterogeneity:
## Q(df = 36) = 267.5978, p-val < .0001
##
## Model Results:
##
## estimate      se    zval    pval   ci.lb   ci.ub
##   1.9922  0.2060  9.6715  <.0001  1.5885  2.3960  ***
##
## ---
## Signif. codes:  0 '***' 0.001 '**' 0.01 '*' 0.05 '.' 0.1 ' ' 1
##
## ========================================================================
##
## ======== Heterogeneity Analysis for Sensitivity and Specificity ========
##
## ========================================================================
## Heterogeneity test for sensitivity
## $statistic
## X-squared
##  911.7971
##
## $p.value
## [1] 4.69832e-168
##
## [1] "I^2:  96.05 %"
##
##
## Heterogeneity test for specificity
## $statistic
## X-squared
##  849.5284
##
## $p.value
## [1] 4.703195e-155
##
## [1] "I^2:  95.76 %"
```

#### Publication Bias

```

## ==========================================================
##
## ========= Funnel Plot, Egger's regression test &  ========
##
## ========= Begg's rank test for lnDOR vs SE(lnDOR) ========
##
## ==========================================================
```

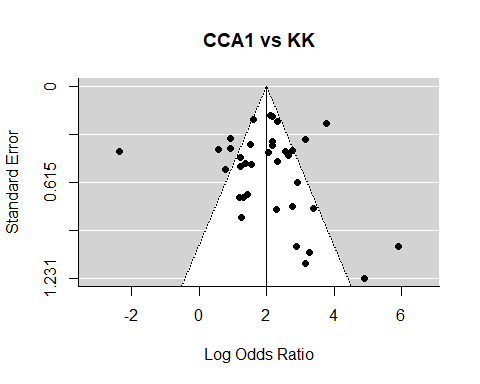

```
##
## Regression Test for Funnel Plot Asymmetry
##
## model:     mixed-effects meta-regression model
## predictor: standard error
##
## test for funnel plot asymmetry: z = 2.0089, p = 0.0446
##
##
## Rank Correlation Test for Funnel Plot Asymmetry
##
## Kendall's tau = 0.1188, p = 0.3014


##
## ==========================================================
##
## ========= Deek's Funnel Plot &                    ========
##
## ========= Asymmetry test for lnDOR vs 1/sqrt(ESS) ========
##
## ==========================================================
```

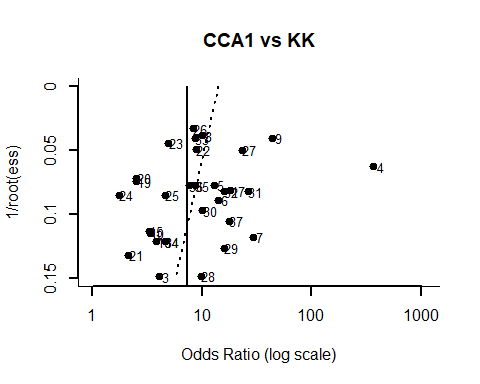

```
##
## Regression Test for Funnel Plot Asymmetry
##
## model:     weighted regression with multiplicative dispersion
## predictor: standard error
##
## test for funnel plot asymmetry: t = -0.9044, df = 35, p = 0.3719
```

### CAA vs KK

#### Heterogeneity

```

##
## ================================================
##
## ======== Heterogeneity Analysis for DOR ========
##
## ================================================
##
## Random-Effects Model (k = 4; tau^2 estimator: REML)
##
## tau^2 (estimated amount of total heterogeneity): 0.0000 (SE = 0.1841)
## tau (square root of estimated tau^2 value):      0.0005
## I^2 (total heterogeneity / total variability):   0.00%
## H^2 (total variability / sampling variability):  1.00
##
## Test for Heterogeneity:
## Q(df = 3) = 4.8116, p-val = 0.1861
##
## Model Results:
##
## estimate      se    zval    pval   ci.lb   ci.ub
##   1.8368  0.2444  7.5144  <.0001  1.3577  2.3158  ***
##
## ---
## Signif. codes:  0 '***' 0.001 '**' 0.01 '*' 0.05 '.' 0.1 ' ' 1
##
## ========================================================================
##
## ======== Heterogeneity Analysis for Sensitivity and Specificity ========
##
## ========================================================================
## Heterogeneity test for sensitivity
## $statistic
## X-squared
##  210.6622
##
## $p.value
## [1] 2.094405e-45
##
## [1] "I^2:  98.58 %"
##
##
## Heterogeneity test for specificity
## $statistic
## X-squared
##  104.8104
##
## $p.value
## [1] 1.435266e-22
##
## [1] "I^2:  97.14 %"
```

#### Publication Bias

```

## ==========================================================
##
## ========= Funnel Plot, Egger's regression test &  ========
##
## ========= Begg's rank test for lnDOR vs SE(lnDOR) ========
##
## ==========================================================
```

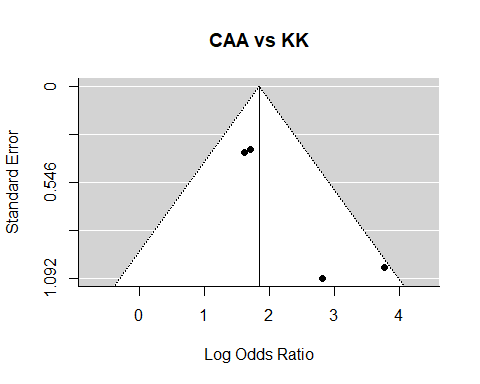

```
##
## Regression Test for Funnel Plot Asymmetry
##
## model:     mixed-effects meta-regression model
## predictor: standard error
##
## test for funnel plot asymmetry: z = 2.0517, p = 0.0402
##
##
## Rank Correlation Test for Funnel Plot Asymmetry
##
## Kendall's tau = 0.3333, p = 0.7500


##
## ==========================================================
##
## ========= Deek's Funnel Plot &                    ========
##
## ========= Asymmetry test for lnDOR vs 1/sqrt(ESS) ========
##
## ==========================================================
```

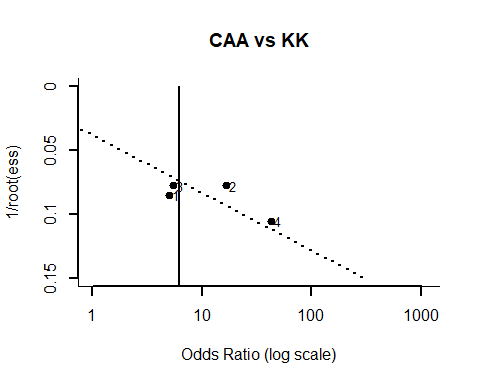

```
##
## Regression Test for Funnel Plot Asymmetry
##
## model:     weighted regression with multiplicative dispersion
## predictor: standard error
##
## test for funnel plot asymmetry: t = 1.1490, df = 2, p = 0.3694
```
